# Evaluating the roles of weather and bird dynamics in accurately forecasting West Nile virus infection in mosquitoes and humans

**DOI:** 10.64898/2026.08.27.26361564

**Authors:** Kayode Oshinubi, Jeffrey Covington, Nicole Busser, John Townsend, James Will, Irene Ruberto, Melissa Kretschmer, Ye Chen, Eck Doerry, Crystal Hepp, Joseph Mihaljevic

## Abstract

Mosquito-borne diseases pose a growing public health challenge as climate change reshapes vector population dynamics. West Nile virus (WNV), transmitted between birds and Culex mosquitoes, disproportionately affects Maricopa County, Arizona, one of the nation’s highest-burden counties, yet whether models that include weather and avian dynamics improve forecast accuracy remains unclear. Using a 15-year weekly time series of mosquito abundance, mosquito infection prevalence, and human cases, we developed four mechanistic model configurations of varying complexity, from mosquito-human dynamics alone to full models incorporating avian dynamics and weather forcing. We fitted each model to the weekly-observed data, generated probabilistic 1- and 2-week-ahead forecast horizons, and evaluated forecasts against a historical baseline. All configurations fit the data equally regardless of weather or avian dynamics. However, models incorporating both birds and weather created more accurate forecasts of mosquito abundance and mosquito infection prevalence, and all configurations outperformed the baseline for forecasting human cases. Forecast accuracy was highest in summer and fall, and ensemble aggregation sometimes outperformed every individual model, stabilizing predictions across the 15-year record. These findings indicate that avian and weather dynamics are most critical for predicting mosquito-specific data, positioning this framework as a scalable tool for public health planning for WNV surveillance under climate change.

## Introduction

West Nile virus (WNV) is the leading cause of domestically acquired mosquito-borne disease in the continental United States, with more than 33,000 cases of human neuroinvasive disease and more than 3000 deaths reported since its introduction in 1999 [1, 2]. Transmitted through an enzootic cycle involving *Culex* mosquitoes and migratory birds as amplifying reservoir hosts, WNV spread rapidly across North America following its emergence in New York City in 1999 and has since become endemic across major parts of the country [3, 4].

Predicting the spillover of vector-borne, zoonotic pathogens from animal populations to humans is inherently difficult due to the complex ecological interactions among wildlife hosts, vectors, and climate [5, 6]. A standard approach for linking climate to the spread of disease requires developing epidemiological models that are explicit about how climate affects specific transmission processes, followed by rigorous testing against longitudinal epidemiological data [7–10]. Statistical models have long explored associations between climate variables and WNV outbreaks [11], and a review identified more than 70 such models [12]. These analyses collectively demonstrate that temperature, precipitation, and drought are significantly associated with interannual variation in WNV incidence [13–21]. Temperature modulates key traits of the mosquito and virus in this system, including the extrinsic incubation period of the virus in mosquitoes, the biting rate of vectors, and the developmental rates of larval mosquitoes, while precipitation and hydrological conditions regulate larval habitat availability and adult mosquito abundance [15, 22–25]. Despite this wealth of statistical evidence, however, few studies have rigorously assessed the mechanistic links between climate and WNV transmission, and fewer still have leveraged these mechanistic insights for operational, probabilistic forecasting [9, 26, 27].

Recent mechanistic modeling studies of WNV show promising results in improving our understanding by linking temperature fluctuations to infection risk via temperature-dependent transmission rates [28–30]. However, most prior modeling frameworks either assume fixed mosquito and bird population sizes, omit the effects of precipitation on mosquito population dynamics, or focus on annual rather than sub-seasonal forecasting horizons [11, 31]. As a consequence, we do not yet have an adequate mechanistic understanding of how mosquito population sizes fluctuate in response to climate on sub-seasonal timescales, whether these fluctuations drive enzootic transmission dynamics, and whether the seasonal timing and magnitude of susceptible bird recruitment modulates these effects [3, 21].

Efforts to evaluate and compare WNV forecasting approaches have accelerated in recent years. The CDC WNV Forecasting Challenges of 2020 and 2022 organized multi-team probabilistic forecasts of the total number of West Nile virus neuroinvasive disease (WNND) cases expected per county for the full calendar year, generated from historical county-level annual case counts supplied by ArboNET together with whatever additional covariates (climate, demographic, mosquito surveillance, or land use) individual teams chose to incorporate. Submissions were updated periodically over the course of the season but always targeted the same single annual total [32, 33]. Another study aggregated case and climate data to the regional level and produced annual rather than sub-seasonal WNND forecasts [34]. This approach used a Bayesian regression framework driven by monthly climate covariates, most often drought indices and temperature, drawn from the preceding October through July.

Analyses of these forecast challenges demonstrated that forecast performance for WNV remains limited, as simple historically derived baseline forecasts have repeatedly outperformed more complex models, including those incorporating climate covariates, with general under-prediction across most models and only modest improvement over historical baselines [32–34]. Within these challenges, models that incorporated weather or demographic covariates tended to score somewhat better than those using mosquito surveillance or land-use data. Even the best climate-informed models, though, rarely outperformed a simple historical baseline forecast. This is part of why more mechanistic approaches, ones that explicitly represent mosquitobird-human transmission dynamics, are needed.

Other recent modeling studies have advanced sub-seasonal WNV forecasting but still highlight gaps in our understanding. Wimberly et al. [35] demonstrated that integrating mosquito surveillance data with meteorological variables substantially improved weekly WNV forecasts in South Dakota relative to models based on either data stream alone. Separately, DeFelice et al. [26] showed that a coupled model-inference framework combining bird–mosquito–human transmission dynamics with data assimilation could accurately predict seasonal human WNV case totals up to nine weeks in advance in Long Island, New York. Incorporating temperature forcing into that same framework further improved forecast accuracy across 12 geographically diverse US counties [27]. Yet these cutting-edge forecasting studies still don’t account for key metrics of both mosquito and human disease risk, including mosquito abundance, mosquito infection prevalence, and human cases, within a single framework. None have explicitly coupled avian reservoir recruitment dynamics with weather-driven mosquito population growth, even though the timing of bird recruitment relative to mosquito emergence is likely what determines whether a given year’s weather conditions translate into an outbreak. Also, none have formally partitioned the independent contributions of weather forcing and bird biology to forecast performance, a distinction that matters for public health agencies deciding where to invest limited surveillance resources, whether in weather monitoring, bird surveillance, deployment of more traps, or all.

Here, we address these gaps using a 15-year (2006–2019, 2021) retrospective forecasting study in Maricopa County, Arizona. Arizona has had annual WNV incidence since 2003 and has contributed approximately 4.8% of all neuroinvasive disease cases nationwide. In some years Arizona accounted for over 20% of national cases, and within Arizona, Maricopa County annually ranks among the top ten counties for WNV disease burden with a cumulative human case count of 3099 between 1999 to 2024 [1, 36]. Also, in 2025 alone, preliminary data shows that Maricopa County has 57 total human cases, [2] while in 2021 there was a large statewide outbreak with 1710 human cases and 127 deaths. The county’s semi-arid climate characterized by hot, dry summers punctuated by a monsoon season (July-September) creates distinctive and interannually variable conditions for *Culex quinquefasciatus* mosquito population dynamics, making it an ideal setting for evaluating weather-driven forecasting models. Understanding, and ultimately forecasting, the factors that drive this disproportionate burden is therefore a public health priority of national significance.

We developed and compared four mechanistic Ordinary Differential Equation (ODE) model configurations of WNV transmission that differ systematically in whether they incorporate weather forcing (temperature and 30-day accumulated precipitation) [37], and whether they explicitly represent an avian reservoir (Figure 1). This design allows us to directly answer the central question of this study: *do weather data and avian reservoir dynamics improve short-term probabilistic forecasts of WNV, and for which surveillance targets?* We fit our models using Bayesian data assimilation, specifically the Ensemble Kalman Filter (EnKF) [38, 39], which is a powerful technique previously applied to influenza and RSV forecasting [39, 40]. Forecast performance was evaluated against a historical baseline using the Weighted Interval Score (WIS) [41, 42] across 1- and 2-week-ahead horizons, seasons, and years. We further evaluated five ensemble combination strategies [43–45] to assess whether aggregating across model configurations provides additional forecast improvement. To our knowledge, this study represents the most comprehensive retrospective probabilistic WNV forecasting evaluation conducted in a single high-burden county and the first to formally disentangle the contributions of weather forcing and avian reservoir dynamics to short-term intra-seasonal forecast accuracy using a systematic multi-model comparison framework and proper probabilistic scoring rules evaluated simultaneously across three epidemiological targets.

**Figure 1.**
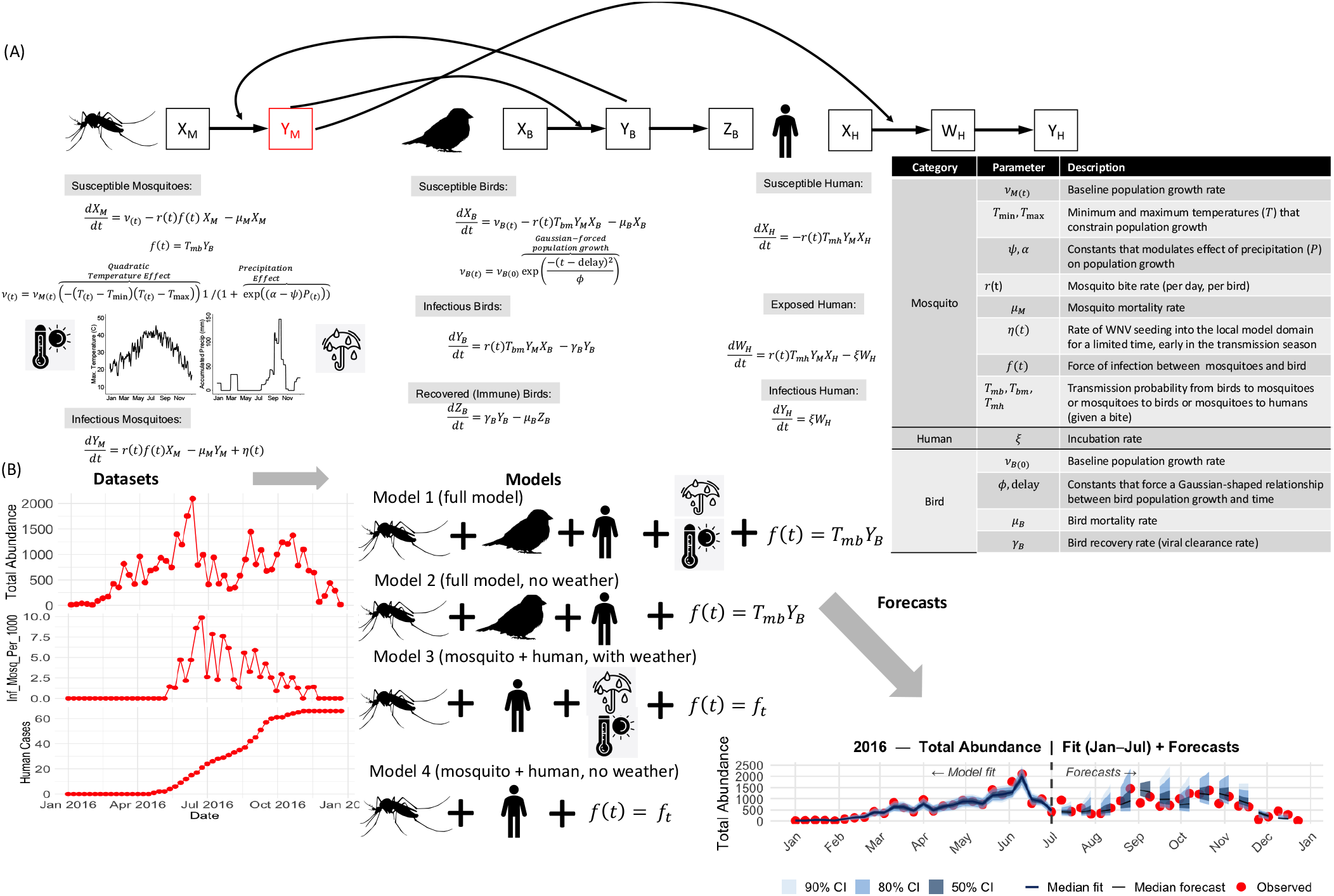
Schematic overview of the four WNV model configurations and the model-filter forecasting framework. Panel (A) shows the mechanistic ODE structure for the full model, including compartmental dynamics for mosquitoes, avian reservoir hosts, and humans, together with the governing differential equations and parameter definitions. Panel (B) illustrates the end-to-end pipeline: weekly surveillance data on mosquito abundance, mosquito infection prevalence, and human WNV cases are used to fit four model configurations of varying complexity via the Ensemble Kalman Filter; probabilistic 1- and 2-week-ahead forecasts are generated using the Ornstein-Uhlenbeck parameter propagation framework.

## Results

### Model fit to surveillance data

When we implement a fully retrospective model-fitting experiment for all four model configurations, it successfully reproduced the seasonal dynamics of total mosquito abundance, IM1000 (kindly note that throughout the text, mosquito infection prevalence (IM1000) is an estimate of prevalence from testing mosquito pools per 1000.), and cumulative human WNV cases across the 15-year dataset (Figure 2, Figures S1–S12, S62-S64 in the Supplementary Material). Specifically, across years, the WIS scores for the model fits of all three observation targets are indistinguishable (Figure 2, top row).

**Figure 2.**
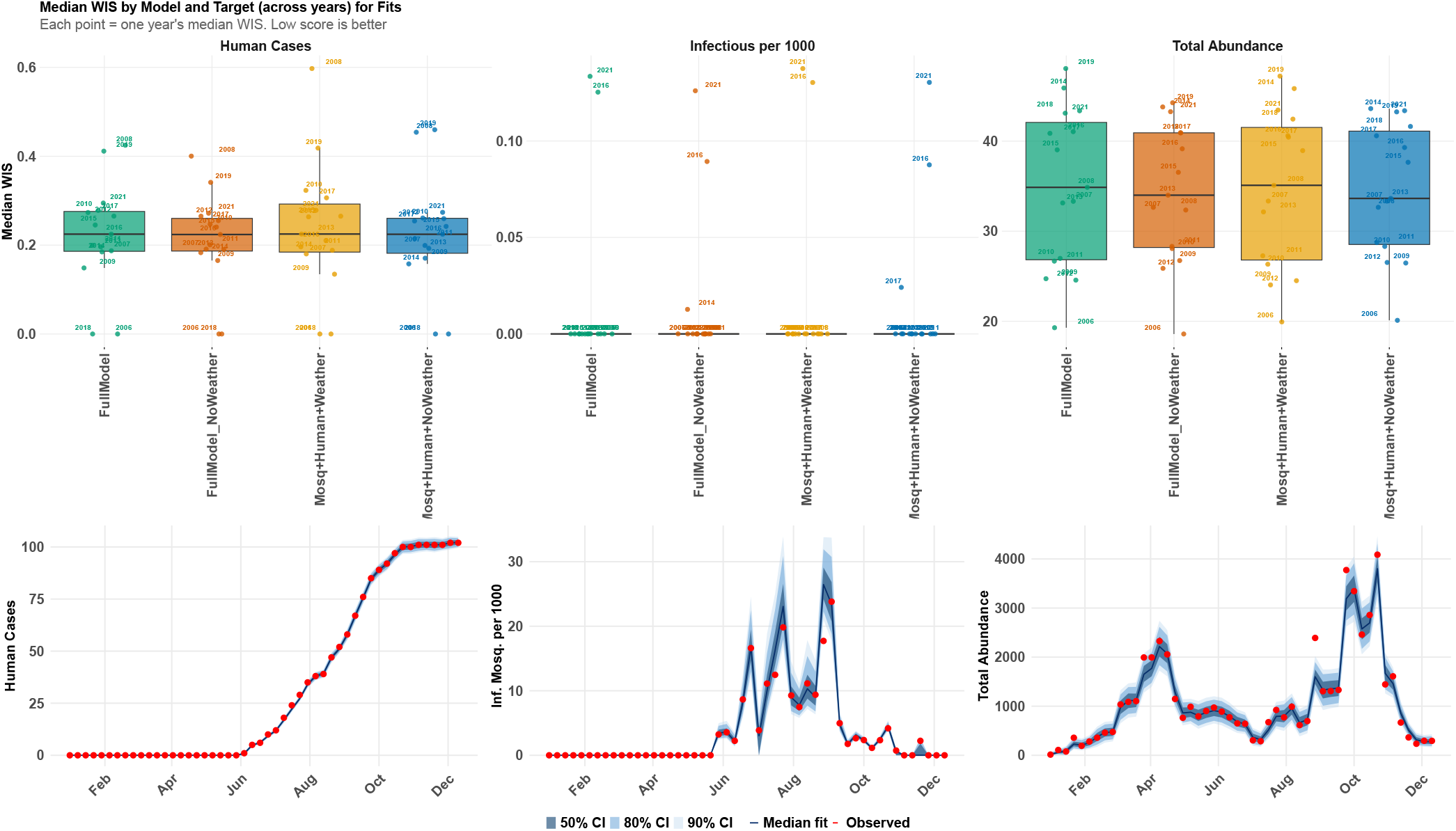
*(Top)* Median Weighted Interval Score (WIS) for in-sample model fits aggregated across all assimilation weeks per year, shown separately for human WNV cases (left), mosquito infection prevalence (IM1000) (center), and total mosquito abundance (right). Each point represents the median WIS for a single year; lower values indicate better fit accuracy. All four model configurations produced comparable fit quality across targets, suggesting that the inclusion of weather forcing or bird dynamics does not substantially alter within-sample fitting performance. *(Bottom)* Model fit for 2014 across three surveillance targets. Each pair shows the EnKF posterior fit (shaded ribbons: 50%, 80%, and 90% credible intervals; solid line: posterior median; red points: observed data). 2014 was selected because it represents a moderate-burden year. Results shown are for the Full Model configuration.

Across models that include weather, the Full Model and the mosquito with human model (M + H), and all 15 study years, the fitted temperature response consistently peaked in a narrow range. The posterior median temperature at which mosquito population growth is maximized was 31.9°C (range 30.9–32.6°C across years and models), with posterior median estimates of *T*_min_ = 19.0°C and *T*_max_ = 44.6°C (see Figure S53 in the Supplementary Material). This consistency across years and independent model configurations indicates that the fitted thermal response is a stable feature of the transmission system, rather than a result specific to a single year’s weather or one model’s structure.

All model fits explained the non-linear, dynamical patterns of mosquito abundance within and among years, even as years varied drastically in mosquito population dynamics (Figure 2). Therefore, our models explain patterns in mosquito abundance that go well beyond what is usually captured in WNV forecasting challenges, which have more commonly focused on mosquito infection rate or human case counts rather than raw abundance [26, 27] (see Figures S1–S12, S62-S64 in the Supplementary Material). Moreover, our model captures detailed patterns of mosquito infection prevalence across the study period, even as climate varied dramatically from year to year. Models incorporating weather forcing (Full Model and M + H) produced slightly tighter posterior intervals during spring and early summer for both mosquito abundance and IM1000, consistent with the mechanistic role of temperature and precipitation in driving early-season mosquito emergence.

In contrast, all four model configurations were able to fit human case data equally well, regardless of weather forcing (median WIS of model fits: Full Model (no W), 0.214; M + H (no W), 0.216; Full Model, 0.219; M + H, 0.222). This suggests that intervening processes between mosquito infection and reported human cases (spillover probability, human exposure behavior, and surveillance reporting) buffer human case data from the same direct meteorological sensitivity observed for abundance and infection prevalence (see Figure 2 and Figures S9-S12, S62 in the supplementary file).

Models without weather forcing (Full Model (no W) and M + H (no W)) still achieved good fits for total abundance by allowing the time-varying population growth rate *ν*_*M*_(*t*) to absorb seasonal trends (see Figure 2 and Figures S1-S4, S63 in the supplementary file). Fit quality was substantially worse in spring than during the rest of the year for all four configurations (median fit WIS: 38.2–39.0 in March–May, versus 31.6–32.3 for the remainder of the year), and within spring specifically, the two models without weather configurations showed modestly higher (worse) fit WIS than models with weather (Full Model (no W): 39.0; M + H (no W): 38.9; versus Full Model: 38.2; M + H: 38.5).

Fits to mosquito infection prevalence (IM1000) were more variable across years than fits to the other two targets (see Figure 2 and Figures S5-S8, S63 in the supplementary file). 2006 stood out as the most difficult year to fit across all four model configurations (median fit WIS during weeks with non-zero observed prevalence, hereafter the active transmission season: 1.44, roughly double the next-highest year, 2012, at 0.73). This is consistent with 2006 being the first year of the Maricopa County trapping program, when the network consisted of only 285 traps, compared to 519 the following year and more than 500 in every subsequent year through 2017 (see Methods and Figure S71 in the supplementary file). The reduced spatial coverage in this pilot year, rather than a genuine failure of the underlying mechanistic structure, is the more likely explanation for the elevated fit error. In years with more established surveillance infrastructure, the four model configurations achieved comparable active-season (see Method for description) fit quality to one another (e.g., in 2021, median fit WIS ranged narrowly from 0.345 to 0.359 across all four configurations), indicating that once trap coverage was sufficient, all four model structures fit IM1000 with similar precision within a given year, and that the year-to-year variability described above is driven primarily by surveillance coverage and interannual differences in transmission intensity rather than by which mechanistic components a given configuration includes.

### Probabilistic forecast accuracy

While retrospective model fits did not vary substantially among model configurations, our analysis reveals that forecast accuracy depends systematically on both avian transmission dynamics and weather forcing (Figure 3, Figures S13-S48 in the Supplementary file). Weather forcing is essential for forecasting total mosquito abundance. Models that include weather substantially outperform models without weather (median relative WIS (log scale): Full Model, − 0.099; M + H, − 0.117; versus Full Model (no W), 0.113; M + H (no W), 0.114), with M + H achieving a marginally lower median relative WIS (log scale) than Full Model. This is consistent with the mechanistic role of temperature and precipitation in driving mosquito population dynamics [37]. Models that explicitly represent weather-driven recruitment capture seasonal transitions in mosquito abundance more accurately than those relying solely on the time-varying growth rate. Performance at the 1-week-ahead horizon was uniformly better than at the 2-week-ahead horizon, reflecting the greater difficulty of forecasting mosquito abundance two weeks in advance [46]. For mosquito infection prevalence (IM1000), avian dynamics strongly impact forecast accuracy (median relative WIS (log scale): Full Model, 0.150; Full Model (no W), 0.483), with weather forcing providing a further, moderate improvement on top of avian dynamics. For human WNV cases, by contrast, all four configurations achieve comparable, meaningful performance above baseline regardless of birds or weather (median relative WIS (log scale) ranging from 1.09 to 0.954 among M + H, Full Model (no W), and M + H (no W); Full Model, − 1.02), indicating that the shared mosquito-to-human transmission pathway alone captures the dynamics needed for useful short-term prediction.

**Figure 3.**
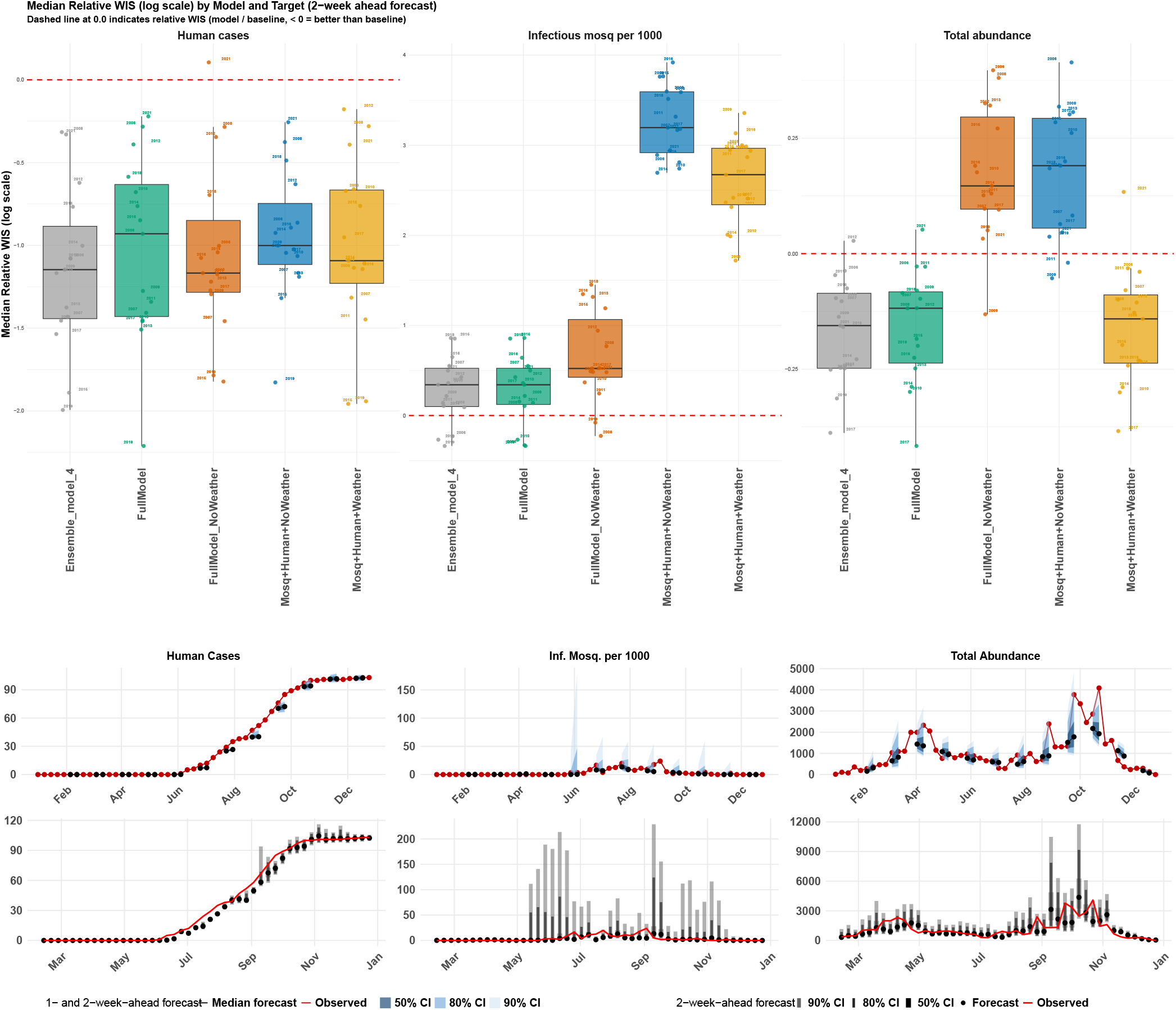
*(Top)* Median relative WIS on the log scale for 2-week-ahead probabilistic forecasts, where values below the red dashed line (0.0) indicate that the model outperforms the historical baseline. Each point represents the median relative WIS for a single year. *(Middle)* 1- and 2-week-ahead probabilistic forecasts for 2014 generated at every fourth assimilation step (shaded ribbons: 50%, 80%, and 90% prediction intervals; black line: median forecast; red line: observed data). Rows correspond to the three forecast targets: total mosquito abundance (last), infectious mosquitoes per 1,000 (middle), and cumulative human WNV cases (first). Results shown are for the Full Model configuration. *(Bottom)* Probabilistic 2-week-ahead forecasts for 2014 for the full model configuration. Each target shows the sequence of 2-week-ahead forecast distributions generated at each of the 46 assimilation steps within a single calendar year (2014). Shaded ribbons indicate the 50% (darkest), 80% (medium), and 90% (lightest) prediction intervals; the solid line shows the forecast median; the red line shows the observed case values. Forecasts were generated by drawing 1,000 samples from the EnKF posterior at each assimilation step and propagating the time-varying parameters two steps forward via the Ornstein-Uhlenbeck process, with each successive step conditioned on the preceding OU-propagated parameter value.

In addition, all four individual model configurations consistently outperformed the baseline across most years and both forecast horizons (1- and 2-week-ahead forecasts). Three of the four configurations, Full Model, M + H, and M + H (no W), achieved a relative WIS (log scale) below zero (i.e., better than baseline) in 100% of the 30 year-horizon combinations evaluated; Full Model (no W) did so in 93.3% of combinations. All four configurations therefore cluster within a narrow 93.3–100% range, indicating that adding avian or weather dynamics does not meaningfully change how often a model beats the baseline for this target. This is consistent with the shared mosquito-human transmission pathway, rather than the additional mechanistic components, driving forecast performance here. Ensemble methods, particularly Ensemble Model 4, achieved the most consistently negative relative WIS (log scale) scores for this target, outperforming the baseline in every year evaluated.

For mosquito infection prevalence (IM1000), performance was more heterogeneous across years and models. No individual model consistently beat the baseline across all 15 years; however, Full Model and Full Model (no W), the two configurations that include bird dynamics, clearly outperformed M + H and M + H (no W), with median relative WIS (log scale) roughly an order of magnitude lower (0.150 and 0.483, respectively, versus 2.24 and 2.83), confirming that avian recruitment dynamics are the dominant driver of forecast performance for this target. Weather forcing provided a further, more moderate improvement on top of this avian structure, with Full Model (0.150) outperforming Full Model (no W) (0.483). M + H and M + H (no W) underperformed the historical baseline for IM1000 in every one of the 15 study years (median relative WIS (log scale) ranging from 1.72–2.76 and 2.41–3.45, respectively), with no significant association between a year’s monsoon precipitation and the degree of underperformance (Spearman’s *ρ* = − 0.36, *p* = 0.19 for M + H ; *ρ* = − 0.18, *p* = 0.53 for M + H (no W); see Supplementary Text S1 for details on how monsoon precipitation and this correlation were computed). This matches the finding above that avian dynamics, not weather, are the dominant driver of IM1000 performance accuracy (see Figures S57-S60 in the Supplementary File), at least in our model configurations. Without a bird compartment, these two configurations cannot capture the dynamics that drive mosquito infection prevalence, no matter that year’s weather conditions. Among ensemble methods, Ensemble Model 4 achieved median relative WIS (log scale) identical to Full Model at both forecast horizons (0.016 at 1 week and 0.35 at 2 weeks), the most competitive performance of any ensemble method for this target, consistent with this weighting procedure concentrating ensemble weight on Full Model, the most skilled individual configuration for this surveillance target [47, 48].

Years of high WNV prevalence in mosquitoes appear to be driven by high recruitment of susceptible birds, and this effect can be amplified by an overlap of high mosquito abundance. For example, in 2010 there was high WNV prevalence in mosquitoes but not particularly high mosquito abundance, suggesting that the outbreak was possibly driven by high bird reproduction and/or immigration (i.e., high recruitment of susceptible birds). In contrast, 2019 was characterized by large mosquito populations overlapping the peak timing of susceptible bird recruitment, potentially causing the elevated WNV prevalence in mosquitoes observed that year. The unprecedented 2021 outbreak, which produced the largest WNV case total ever recorded in Maricopa County [49], similarly reflects a confluence of high mosquito abundance and elevated transmission pressure that our model captures retrospectively, albeit with wider uncertainty intervals reflecting the extreme nature of that season (Figure 2).

### Forecast accuracy varies substantially by year

Consistent with findings in Cramer et al. [42] for COVID-19, the relative performance of all individual models showed high variability across years (Supplementary Figures S13-48, S57–S60 in the supplementary file). Even the best-performing individual configuration, the Full Model, ranked in the top half of all nine models (the four individual configurations plus five ensemble methods) in only 62.2% of year-target-horizon combinations evaluated, with the remaining three individual configurations performing less consistently (46.7%, 41.1%, and 11.1% of combinations for Full Model (no W), M + H, and M + H (no W), respectively). This indicates that mechanistic models alone substantially benefited from ensembling, especially in some years. High-burden years such as 2021 were associated with markedly larger absolute WIS values for human cases (median absolute WIS = 2.02), more than double that of any other year in the study period (2008: 1.00), though relative WIS scores did not deteriorate correspondingly, suggesting that when transmission is elevated, mechanistic models benefit from more observational signal to condition their forecasts. This pattern was not uniform across all years. In 2008, a year with moderate disease burden, we observed the second-highest absolute WIS for human cases in the study period (median absolute WIS = 1.00), indicating that factors beyond transmission intensity alone, potentially including reporting irregularities or atypical case timing, can also drive forecast difficulty in a given year. Still, in the biggest outbreak years that matter most for public health response, our models maintain or improve their forecast performance advantage over the baseline.

### Seasonal patterns in forecast performance

Forecast performance for all three surveillance targets improved steadily over the course of the calendar year, from winter through fall, though the point at which models began to outperform the baseline differed by target. For human cases, median relative WIS (log scale) declined from 0.66 in February to − 2.31 in December for the Full Model, becoming better than the historical baseline around May. For total abundance, the two models with weather configurations (Full Model and M + H) scored below baseline in nearly every month from February onward (e.g., Full Model: − 0.01 in February, − 1.05 in December), while the two models without weather configurations remained worse than baseline until November. Ensemble Model 4 closely tracked the Full Model across the season for both human cases (e.g., − 2.26 in December, versus − 2.31 for Full Model) and IM1000 ( −0.48 in December, versus − 0.83 for Full Model), confirming that this ensemble method reproduces the seasonal advantage of the best individual configuration for these two targets. The size and timing of these seasonal trends differed by target, as detailed below (Figure 4 for all years, Figure 5 for 2014, and Supplementary Figures S58, S60, S65–S70 for all nine model configurations).

**Figure 4.**
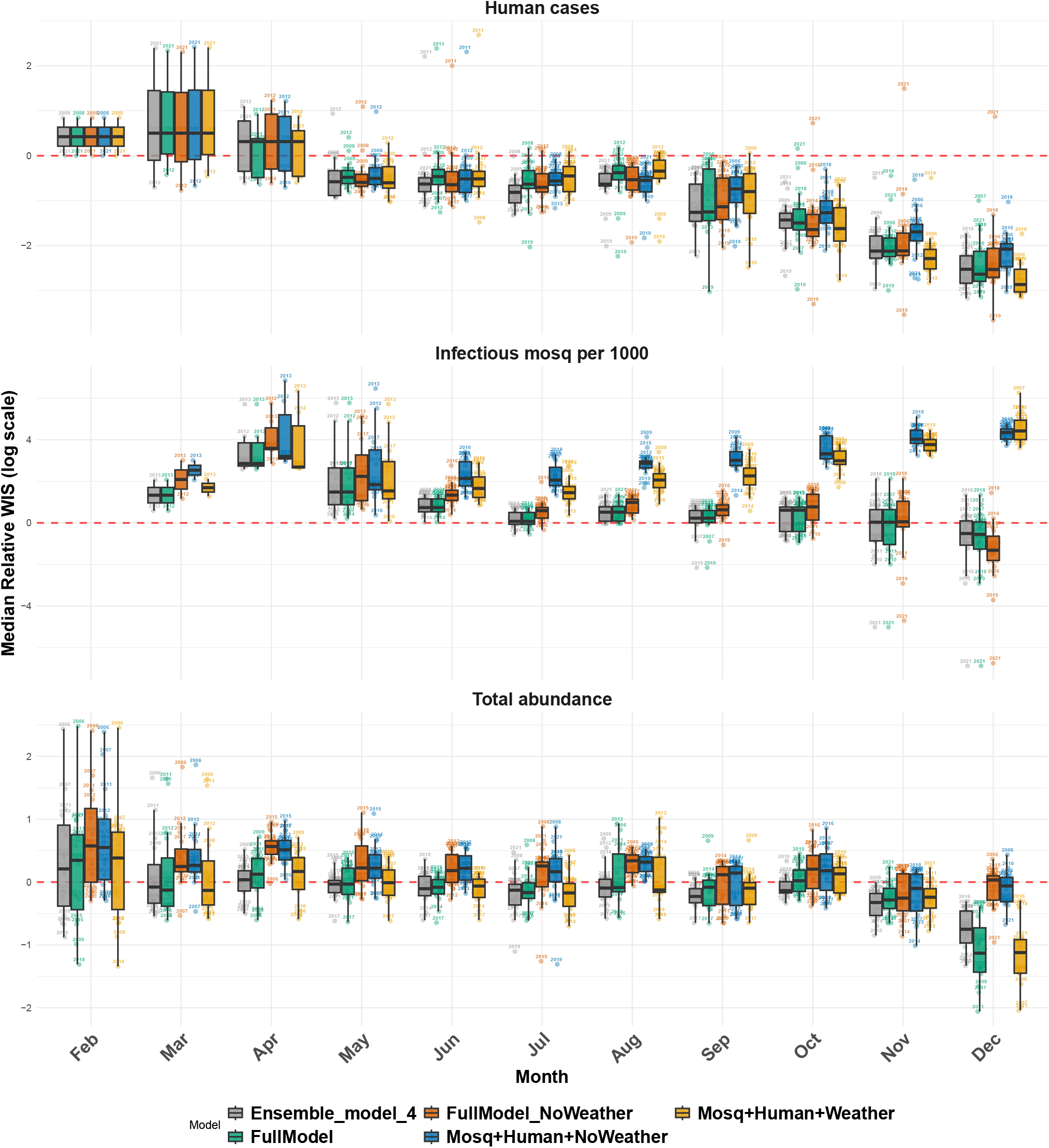
Seasonal patterns in 2-week-ahead forecast performance across model configurations and surveillance targets, aggregated over the 15-year study period (2006–2019, 2021). Each boxplot summarizes the distribution of per-year median relative weighted interval score (WIS) on the log scale for a given model, month, and target, where each box aggregates across all available years for that calendar month. The red dashed line at 0.0 marks the boundary between better-than-baseline (below) and worse-than-baseline (above) performance. Results are shown for human WNV cases (top), infectious mosquitoes per 1,000 (middle), and total mosquito abundance (bottom). January is excluded, as forecasts were not generated for that month.

**Figure 5.**
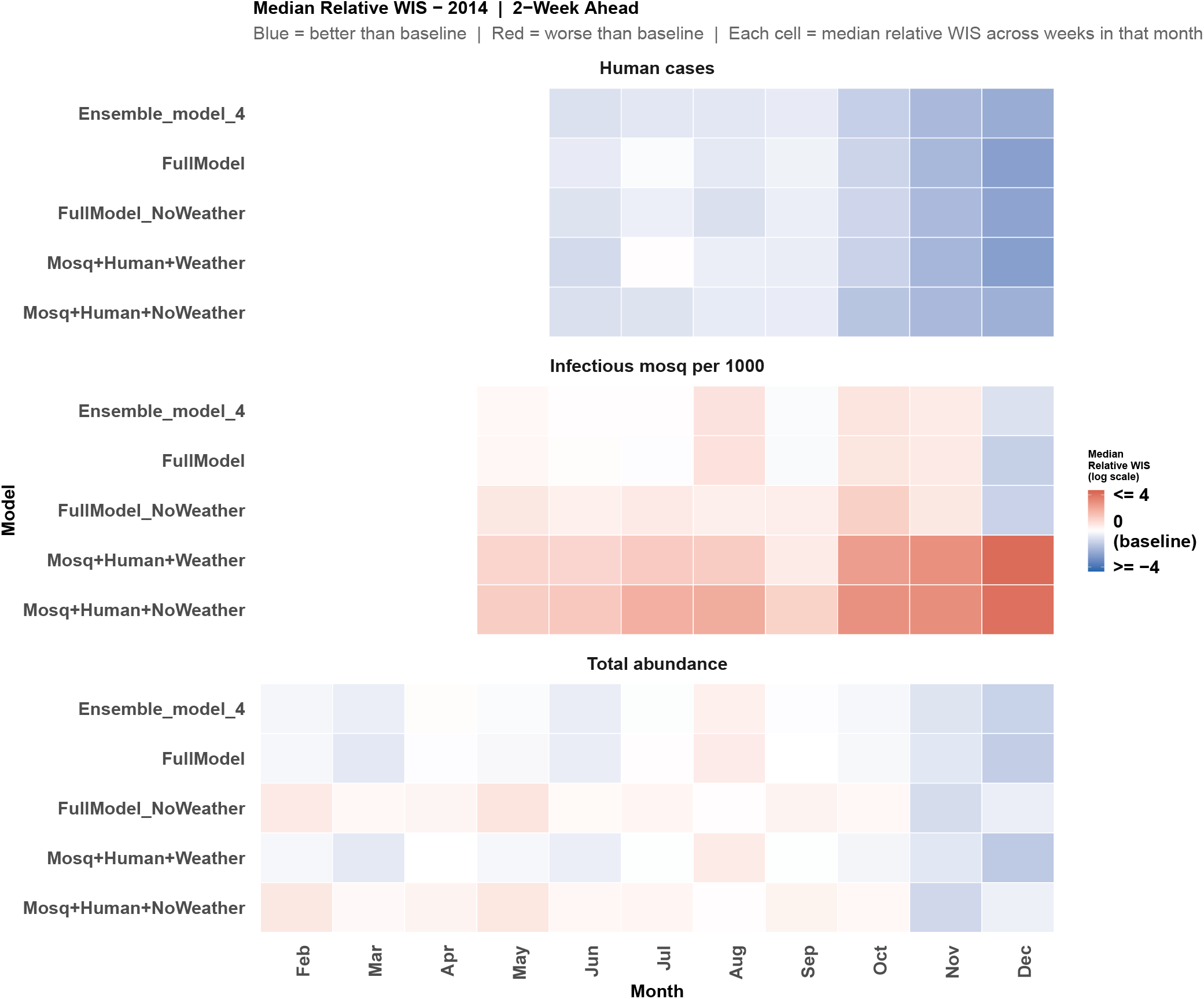
Median relative weighted interval score (WIS) on the log scale for 2-week-ahead probabilistic forecasts by model configuration, calendar month, and surveillance target for 2014. Each cell represents the median relative WIS aggregated across all forecast weeks falling within that calendar month, where relative WIS is defined as the log ratio of the model WIS to the historical baseline WIS. Values below zero (blue) indicate that the model outperformed the historical baseline; values above zero (red) indicate worse-than-baseline performance. Results are shown for five model configurations: the Ensemble Model 4, the Full Model, Full Model (no W), M + H, and M + H (no W), across three surveillance targets: human WNV cases (left), mosquito infection prevalence (IM1000) (center), and total mosquito abundance (right).

For human cases, all four configurations substantially outperformed the baseline during summer (July–September) and fall (October–December), when rising case counts give mechanistic models more of the current season’s trajectory to draw on. Median relative WIS (log scale) for Full Model fell from 0.63 in July to 2.31 in December, a pattern closely mirrored by Ensemble Model 4 ( − 0.77 in July, − 2.26 in December). Performance in winter (February–March; no forecasts were generated in January) and early spring (April) was at or slightly worse than baseline (median relative WIS (log scale) 0.42–0.66 across configurations), as expected given the near-zero case counts typical of those months.

For mosquito infection prevalence (IM1000), all four individual configurations scored worse than baseline for most of the year, reflecting the inherent difficulty of forecasting this target. Among them, the Full Model had the smallest gap above baseline in summer and fall (median relative WIS (log scale): − 0.02 in July, 0.24 averaged across fall), and the Full Model and the Full Model (no W) were the only two configurations to score better than baseline at any point, doing so in November and December. Ensemble Model 4 showed the same pattern, scoring below baseline in November ( − 0.22) and December ( − 0.48). The two configurations without bird dynamics (M + H and M + H (no W)) never scored below baseline in any month and instead grew steadily worse across the season, from a median relative WIS (log scale) of about 1.4–1.6 in March to 4.3–4.4 in December. This late-season divide between the models with bird configurations (including Ensemble Model 4, which weights them most heavily) and the non-bird configurations is consistent with mechanistic knowledge of bird population dynamics providing a signal that models without bird configurations lack.

For total abundance, all four individual configurations clustered relatively close to baseline from February through April (median relative WIS (log scale) between − 0.19 and 0.40), before the two models with weather configurations (Full Model and M + H) diverged from models without weather. From May through December, Full Model and M + H scored below baseline in every month (e.g., Full Model: − 0.02 in October, − 1.05 in December), while Full Model (no W) and M + H (no W) remained at or above baseline until November. Ensemble Model 4 followed the models with a weather pattern throughout, scoring below baseline from March onward and reaching − 0.68 in December. This reflects the importance of temperature and precipitation forcing during spring mosquito emergence, when year-to-year variation in weather is the main reason abundance trajectories differ from one year to the next.

### Forecast accuracy degrades with the horizon

Averaging across all years and targets, 2-week-ahead forecasts showed higher relative WIS values than 1-week-ahead forecasts for all models. This degradation was most pronounced for total mosquito abundance, where the 2-week-ahead median WIS was approximately 20-40% higher than at the 1-week-ahead horizon. This pattern is consistent with general findings on forecast accuracy degradation with increasing lead time in epidemic forecasting [42, 50] and reflects the fundamental limit imposed by the growing uncertainty of the forecasted parameter trajectories over a two-week window [46]. For human cases the horizon effect was relatively modest, consistent with the smooth cumulative nature of that target, which makes consecutive weeks strongly correlated.

### Ensemble combination improves forecast stability

Ensemble methods improved forecast accuracy and year-to-year stability relative to individual mechanistic models, but the degree of improvement and the ranking of specific ensemble approaches differed substantially across the three surveillance targets [32, 42, 50] (Figures 3–5, Supplementary Figures S57–S60, S65-S71 in the supplementary file). No single ensemble method ranked first across all three targets, and the performance spread among ensemble approaches was far larger than the spread among individual mechanistic models for some targets, underscoring that the choice of ensemble method is consequential and target-dependent. Ensemble Model 4 was the most broadly reliable ensemble method across all three targets, ranking first for infectious mosquito prevalence (IM1000; median relative WIS (log scale): 0.148, tied with Full Model at 0.150) and total abundance ( −0.137), and second for human cases ( −1.15, within 0.05 log-units of the best-performing ensemble method, Ensemble Model 3). Its year-to-year variability was lower than, or comparable to, that of the best-performing individual model for each target (SD: 0.433 vs. 0.481 for human cases; 0.310 vs. 0.312 for IM1000; 0.096 vs. 0.113 for total abundance), confirming that its optimization against the WIS objective translates into consistent gains across both targets and years [43, 47, 48]. More details in the supplementary text S2 in the supplementary file.

## Discussion

Mathematical models play a powerful role in revealing the mechanistic drivers of the spread of infectious disease in wildlife and human hosts [51, 52]. Our results clarify the relative importance of weather forcing and avian reservoir dynamics for WNV forecasting. Both matter, but not equally, and not for the same surveillance target. This study represents the first comprehensive evaluation of key biological mechanisms in the WNV system that drive short-term forecast accuracy. Our integration of models and data suggests that patterns of WNV prevalence in mosquitoes are driven by weather’s effects on mosquito demography and by the timing and magnitude of susceptible bird recruitment.

Our strategic comparison of four model structures allowed us to isolate the independent contributions of avian reservoir dynamics and weather forcing to forecast accuracy. First, our models allow non-linear effects of temperature and precipitation to regulate dynamical patterns of mosquito population abundances, enabling a realistic explanation of temporal fluctuations in the number of infectious mosquitoes in the environment, which mediates infection risk. Second, previous mechanistic forecasting efforts [26, 27] did not allow for realistic, dynamically changing bird population densities; our model fills this gap, and our factorial model comparison allows us to isolate its contribution.

### Weather forcing is essential for mosquito-specific targets

Weather forcing was essential for forecasting total mosquito abundance, and provided a moderate improvement for mosquito infection prevalence (IM1000). The temperature constraint (Eq. 15) and the logistic precipitation term together captured threshold-like seasonal responses, contributing to the stability of fits and forecasts across the 15-year record [37]. The importance of weather was most pronounced in spring, when temperature-dependent emergence is the dominant driver of mosquito population growth. This is consistent with the broader literature linking temperature and precipitation to *Cx. quinquefasciatus* population dynamics across the US [14, 19, 20, 23, 34], and with our own prior mechanistic analysis of this system [37]. Our fitted temperature response peaks near 31.9°C, consistent across years and model configurations, which corresponds reasonably well with the thermal performance of the development rate and egg hatching proportion of *Cx. quinquefasciatus* [53]. While a recent study [53] estimated the thermal optimum of WNV transmission at 25.2°C using a trait-based *R*_0_ framework, our mechanistic approach operates at the population level and integrates temperature effects on the net mosquito growth rate rather than individual traits, providing a complementary and operationally tractable estimate of climate-abundance relationships.

### Susceptible bird recruitment mediates WNV outbreak intensity

Model configurations that included avian reservoir dynamics outperformed those without them for infectious mosquito prevalence (IM1000), and the addition of bird dynamics to the weather-inclusive configuration further improved fits to total mosquito abundance, consistent with the known role of avian amplification in driving secondary peaks in mosquito infection prevalence [3, 4]. We interpret the year-specific patterns described above (2010, 2019, 2021) cautiously, because the bird recruitment function in our model is not directly parameterized from independent bird surveillance data but is instead estimated from the observed transmission dynamics themselves, we cannot definitively attribute improved forecast performance to birds specifically, as opposed to other unmeasured biological processes captured by the additional flexibility of the bird compartment. What the model does show is that including an avian compartment is necessary to reproduce the temporal dynamics of IM1000 across the study period in a way that weather forcing alone cannot achieve and that the magnitude and timing of estimated bird recruitment co-vary meaningfully with observed interannual variation in WNV prevalence. These patterns are biologically plausible given regional variability in avian host competence and the known outsized influence of particular bird species on WNV amplification [4, 54, 55], but they remain model-inferred rather than directly observed.

Three targeted extensions are needed to move from these model-inferred conclusions toward validated mechanistic inference. First, independent longitudinal data on bird population densities and WNV seroprevalence in Maricopa County are needed to directly parameterize and validate the bird recruitment function, rather than estimating it from transmission data alone. Second, species-specific demography and migration dynamics should be incorporated, since the current model treats the avian reservoir as a single homogeneous compartment and cannot distinguish the contributions of highly competent host species (e.g., house sparrows, house finches) from those of less competent ones. Third, the environmental drivers of bird recruitment itself, including the effects of temperature and precipitation on breeding success and immigration timing, should be characterized to allow mechanistic prediction of the timing and intensity of bird-driven outbreak amplification, rather than estimating recruitment as a latent state inferred from the data.

### The role of drought, hydrology, and climate legacy effects

Several aspects of the hydrology-WNV relationship merit further investigation. Hydrology, drought, and historical precipitation patterns vary drastically across regions and can have nuanced effects on WNV outbreaks. Drought as long as six months prior has been hypothesized to alter contact rates between birds and mosquitoes through aggregation around available water sources [15, 21], but drought has also been associated with fewer mosquitoes, perhaps through reduced breeding habitat and concentration of larval predators [13]. Our model includes the effects of 30-day accumulated precipitation to account for the larval development period of mosquitoes but does not currently capture legacy effects of drought or other climatic events on mosquito or bird population dynamics.

In the desert Southwest context of Maricopa County, we hypothesize that drought can lead to enhanced irrigation and other anthropogenic water sources that create localized mosquito breeding hotspots, a mechanism not currently represented in our county-level model [37]. While our forecast error analysis did not reveal systematic directional bias in the weather-inclusive model configurations during the driest years of the study period, this mechanism may nonetheless contribute to residual variability in abundance that is not captured by county-average precipitation alone. Incorporating spatially explicit anthropogenic water use as a modulator of mosquito population growth rates therefore represents a promising direction for improving model realism, particularly for forecasting localized abundance hotspots during drought years in irrigated urban landscapes.

### WNV endemicity and the closed-system assumption

Our model assumes recurrent WNV outbreaks arise from virus over-wintering in resident birds or mosquitoes, without importation of new viral variants; in other words, a closed enzootic system. This assumption is well-supported for Maricopa County, where phylogenomic analyses have shown that WNV in Arizona is predominantly endemic and that Arizona serves as a source of WNV for several areas of the US Southwest [36]. In contrast, in the northeastern US there is a higher rate of viral reintroduction with few endemic strains [56], likely due to lower overwintering rates of mosquitoes in hard-freeze climates. Extending our framework to such regions would, therefore, require adding stochastic viral seeding events [26] to capture the more sporadic, importation-driven dynamics of WNV in those areas.

### Human WNV case forecasting is robust to model complexity

Mechanistic forecasting consistently outperformed the historical baseline for human WNV cases regardless of model complexity. Even the most parsimonious model configuration (M + H (no W)) provided useful short-term predictions. This finding may have practical implications for resource-constrained public health agencies requiring only human case early warning. A model that requires neither bird surveillance data nor real-time weather inputs may be sufficient. This result also contextualizes the negative findings from the CDC WNV Forecasting Challenges [32, 33], where models failed to consistently outperform historical baselines at annual time scales. Our retrospective framework demonstrates that performance is achievable at sub-seasonal (weekly) horizons, suggesting that the annual aggregation used in those challenges may obscure meaningful within-season predictive power.

### Ensemble modeling and forecast stability

Beyond the overall stabilization benefit of ensembling described above, the mechanism behind these results is instructive. For IM1000, where individual model configurations disagreed most sharply (see Results), ensemble methods that weight models approximately equally (Ensemble Models 1, 2, and 5) were dragged toward the majority of poorly performing configurations, including Ensemble Model 5, which showed no competitive advantage here despite its theoretical strength in combining distributional uncertainty [44, 45]. The Ensemble Model 4 avoided this problem by concentrating weight on the model configurations that include the bird, which actually explain IM1000 dynamics [47, 48].

This suggests that when individual models disagree substantially and that disagreement has a clear mechanistic basis, as it does here with bird dynamics separating strong from weak configurations, a performance-weighted or performance-optimized combination is preferable to simple aggregation. This mirrors findings from the COVID-19 Forecast Hub, where ensemble forecasts were the only models to outperform the baseline across all locations and weeks [42], and from the CDC WNV Forecasting Challenges, where no single model consistently outperformed historical baselines [32, 33]. Moreover, previous literature [32, 42, 50] supports that not all ensemble models can outperform the baseline, which we also observed in our analysis (see Figures S57–S60 in the Supplementary Material). The fact that all our model configurations forecast human cases accurately and better than baseline (regardless of whether birds or weather are included) suggests that for this target, the mosquito-human transmission pathway captures sufficient seasonal dynamics to provide useful short-term predictions.

The reversal in Ensemble Model 3’s ranking between human cases and total abundance (see Results) illustrates a related risk. A method well-suited to one target can perform poorly, or worse than baseline, on another. Ensemble Model 4 avoided this failure mode by ranking consistently well across all three targets. This trade-off is consistent with Oidtman et al. [57]’s finding in the context of the 2015–2016 Zika epidemic in Colombia, where individual models occasionally outperformed ensembles early in outbreaks but ensembles outperformed individual models on average. Our results extend this finding by showing that the same target-dependence applies across ensemble methods themselves, not just between individual models and ensembles, and that operational multi-target forecasting systems should prioritize ensemble methods that perform robustly across all targets over those that excel on a subset.

### Implications for vector control and public health policy

Beyond demonstrating forecast performance, our findings carry direct implications for operational vector-borne disease management in Maricopa County and similarly arid, urbanized settings where WNV is endemic. Vector control interventions such as larviciding, adulticiding, and source reduction typically require on the order of days to weeks to implement and to take effect, a timescale well matched to the 1- and 2-week forecast horizons evaluated here. More importantly, our results are consistent with outbreak intensity being shaped by the coincidence of peak avian recruitment timing with high mosquito abundance, rather than by either factor alone, a mechanistically interpretable signal that purely statistical or machine-learning forecasts cannot provide. Years in which these two processes appear to have converged, such as 2019 and 2021, produced the most severe transmission seasons in our retrospective analysis in terms of both peak mosquito abundance and cumulative human case counts. A forecast system capable of flagging such convergence in near-real time could give vector control districts an early, mechanistically grounded basis for intensifying surveillance and control efforts ahead of the seasonal case peak, rather than responding only after trap-based infection rates or human cases have already risen. The reduced year-to-year variability achieved by ensemble methods, particularly Ensemble Model 4 (see Results), is similarly relevant from an operational standpoint, since public health agencies require forecasts that are reliable from year to year, not merely accurate on average, to justify routine integration into decision-making.

Translating these retrospective results into a deployable early-warning tool will require confronting the broader adoptability challenges recently articulated for climate-sensitive disease forecasting. Phung et al. [58] propose a “3-U” framework, useful, usable, and used, for advancing digital prediction tools from research demonstrations into routine public health practice. Our results speak most directly to the “useful” criterion, showing that a mechanistic ensemble Kalman filter approach can achieve sufficient accuracy and lead time to outperform a historical baseline across three surveillance targets. Progressing toward “usable” and “used” status, however, will require engagement with local vector control and public health agencies to determine whether forecast outputs align with existing operational thresholds, along with prospective validation of forecast-informed interventions to demonstrate real-world efficacy and cost-effectiveness. The 2021 WNV outbreak in this same county, the largest ever recorded in a single U.S. jurisdiction [49], underscores the public health stakes of closing this gap between forecast performance and operational deployment of the model.

### Limitations and future directions

Our ensemble weights were estimated using the same within-year data used for evaluation, constituting in-sample optimization [43]. A fully prospective evaluation, estimating weights from prior years and applying them to the current year, would provide a more operationally realistic assessment and will be a priority for future work. Our framework was developed for a single county; extension to other high-burden regions with different ecological and climatic profiles is needed to establish the broader applicability of our findings. We did not model interannual variation in human or bird immunity, which may contribute to the elevated transmission observed in years such as 2019 and 2021 and could be incorporated through susceptible-recovered-susceptible compartment extensions. Future implementations should also consider finer spatial scales to capture the microclimate and irrigation heterogeneity within Maricopa County that can produce localized mosquito abundance hotspots even during county-wide drought years [37]. As discussed in the preceding paragraph, direct parameterization of the bird recruitment function from independent longitudinal surveillance data remains a critical next step for validating the avian-driven mechanisms inferred here. Operational deployment will also require integration with real-time numerical weather forecasts, which are reliable only up to approximately 10 days ahead [46]; as seasonal climate forecast products improve, particularly for the Southwest monsoon, extending the forecast horizon beyond two weeks will become increasingly feasible. Extension to longer horizons, for example, using seasonal climate outlooks or probabilistic ensemble weather forecasts as forcing inputs, represents an important direction for future work and will be explored as part of a prospective real-time deployment of this framework in Maricopa County. Finally, future implementations could explore particle flow filter approaches [59], which may offer improved performance in years such as 2021, where the posterior distribution departs substantially from Gaussianity due to extreme transmission dynamics. Our analysis emphasizes that the combination of susceptible bird recruitment and weather-driven mosquito abundance mediates WNV transmission risk in Maricopa County and that a characteristic mismatch between the timing of favorable mosquito climate (cooler, wetter monsoon conditions) and peak susceptible bird recruitment underlies the interannual variability in outbreak intensity. Dynamical models that explicitly represent how climate regulates both components of this interaction will improve our understanding of WNV across diverse ecoregions [52, 60].

## Methods

### Methodology overview

We developed four mechanistic compartmental models of WNV transmission dynamics that differ systematically in their complexity. All four models share a core mosquito-to-human transmission structure based on ordinary differential equations, but they vary in whether they explicitly represent the dynamics and transmission from avian reservoir host (migratory birds), and whether mosquito population growth is modulated by observed daily weather conditions (temperature and precipitation). By systematically varying the inclusion of weather forcing and bird dynamics across four configurations, we are able to evaluate the contribution of each component to forecast performance. Our motive is therefore to test which mechanistic dynamics most contribute to model fit and model forecast accuracy.

We fit each model with an Ensemble Kalman Filter (EnKF), sequentially assimilating weekly observations of mosquito abundance, mosquito infection prevalence (IM1000), and cumulative human WNV cases to generate rolling 1- and 2-week-ahead forecasts across each 15-year study year (2006–2019, 2021).

We evaluated the forecast performance of the four model configurations against a historical baseline model (i.e., a type of null model), using the raw and log-scaled Weighted Interval Score (WIS and log(WIS)). We calculated forecast performance across 1- and 2-week-ahead horizons, monthly, and all 15 years, enabling a detailed decomposition of where and when each model configuration adds predictive value. Finally, we also created five ensemble models using different combination methods to assess whether ensembling forecasts across the four model configurations provides further improvements in forecast performance. Below, we describe each component of the framework in detail.

### Data

#### Mosquito abundance data

Weekly mosquito surveillance data were collected and aggregated by the Maricopa County Environmental Services Vector Control Division [36], using a network of carbon dioxide-baited light traps deployed throughout the county, with coverage concentrated in residential and accessible areas. The trap network grew substantially over the study period, from 285 traps in 2006, the program’s pilot year, to 828 traps by 2021 (Supplementary Figure S71), and the time-varying baseline mosquito population growth rate is intended to partly absorb this trend, since more traps are expected to capture more mosquitoes independent of any change in the underlying population. Given the especially limited spatial coverage in 2006, model fit and forecast quality for that year should be interpreted with this constraint in mind.

Each trap was operated on a weekly schedule with a 12-hour collection window, yielding surveillance records for up to 50–53 weeks per calendar year depending on the year. Field collections were sorted taxonomically by species and sex, and our analysis was restricted to adult female *Cx. quinquefasciatus*, which constitutes the primary WNV amplification vector in the region and accounts for approximately 80% of WNV-positive mosquito pools detected in Maricopa County. The county-level abundance index used in model fitting was defined as the aggregate weekly count of female *Cx. quinquefasciatus* captured across all operational routine traps in a given week. Further methodological details regarding data collection and quality control procedures are described in [36, 37, 61].

Across the 15 years of the study period (2006–2019 and 2021), we generated 1- and 2-week-ahead forecasts for three epidemiological targets (total mosquito abundance, mosquito infection prevalence (IM1000), and cumulative human WNV cases), using four individual model configurations and five ensemble methods, yielding a total of 15 ×

46 × 2 × 3 × 9 = 37,260 individual forecast-observation pairs. The evaluation period captured substantial interannual variability in WNV transmission intensity, including high-burden years such as 2019 and 2021 and relatively low-burden years such as 2009 and 2018 (Figure 2). This time period also includes high variation in mosquito population dynamics: four of the 15 study years (2014, 2018, 2019, and 2021) fell within the top quartile of peak seasonal total abundance. Three of these four years (2014, 2018, and 2021) coincided with above-median monsoon-season (June–September) precipitation, consistent with rainfall-driven expansion of mosquito breeding habitat. 2019 is a notable exception, with near-record peak abundance despite monsoon precipitation among the lowest of the study period, suggesting that factors other than seasonal rainfall, such as the overlap with avian recruitment timing discussed below, drove that year’s outbreak.

#### Mosquito infection prevalence (IM1000)

As a measure of WNV prevalence in the mosquito population, the proportion of infectious mosquitoes was estimated as:

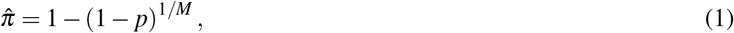

where *p* is the proportion of pools that tested positive for WNV, and *M* is the average number of mosquitoes tested per pool [51]. The WNV prevalence in mosquitoes (IM1000) was then estimated as 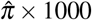, and this derived quantity served as the second observational target in our model-filter system.

#### Human WNV case data

Weekly human WNV case data for Maricopa County were obtained from Maricopa County Department of Public Health. The dataset contained aggregated numbers of cases by week. The resulting weekly incidence series was then transformed to cumulative cases, which increased across weeks in a given year. Cumulative case counts were used because we needed a data type that linked directly to a state variable in the model, rather than being “new events.” This way, the model state variable also has all the hospitalizations that have occurred. The study period spanned 2006–2019 and 2021; year 2020 was excluded due to a low case count of 3 attributed to mosquitoes not testing positive in high numbers. It is unclear the mechanism that led to this low incidence year.

#### Temperature and precipitation data

County-level daily maximum temperature (degrees Celsius) and precipitation (millimeters) were derived from the PRISM gridded climate dataset [62], which provides spatially continuous climate surfaces at 4 km × 4 km resolution across the contiguous United States. For each day in the study period, all raster grid cells intersecting the Maricopa County administrative boundary (2010 US Census Bureau delineation) were averaged to produce a single county-level daily estimate. We used average daily maximum temperature (°*C*) as well as the 30-day rolling accumulation of average daily precipitation (mm) divided by a constant of 50 [37]. This linear scaling of the precipitation data was applied for numerical reasons: observed 30-day accumulated precipitation in Maricopa County spans a wide range (0–230 mm across the full year, with non-monsoon months typically between 0–50 mm and monsoon peaks reaching 100–230 mm), and dividing by 50 places the precipitation covariate on a 0–5 scale, which is more convenient for the exponential effects we assume in the model [37].

### Dynamical models of WNV transmission

We define this family of models as “mechanistic” in the sense that they explicitly represent biological processes (mosquito recruitment, infection, host-to-host transmission, and human incubation) while using flexible statistical functions to characterize the effects of weather on demographic rates, following our earlier work [37]. This differs from fully stage-structured models that simulate each mosquito life stage explicitly with temperature-dependent developmental rates. Our approach instead prioritizes fitting temporal patterns in field-collected surveillance data while maintaining the simplest key mechanistic structure necessary to represent WNV transmission across mosquito, bird, and human hosts.

#### Model 1 — mosquito + bird + human + weather (Full Model)

The full model tracks susceptible (*X*_*M*_) and infectious (*Y*_*M*_) mosquitoes, susceptible (*X*_*B*_), infectious (*Y*_*B*_), and recovered/immune (*Z*_*B*_) birds, and susceptible (*X*_*H*_), exposed (*W*_*H*_), and infectious & cumulative hospitalized (*Y*_*H*_) humans:

#### Mosquitoes

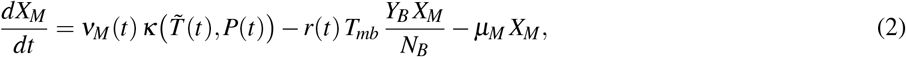

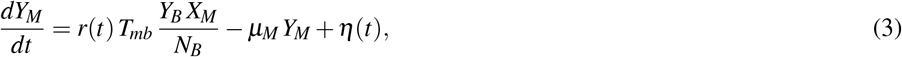

where *N*_*B*_ = *X*_*B*_ + *Y*_*B*_ + *Z*_*B*_ is the total bird population, and *η*(*t*) is a small seeding rate representing WNV reintroduction via migratory birds following the kind of description in [27] (see Supplementary text S1 for full specification). *T*_*mb*_ is the transmission probability from infectious mosquitoes to susceptible birds per bite. *µ*_*M*_ = 0.05 day^−1^ is the mosquito mortality rate, which we fix so that the time-varying birth rate can drive temporal fluctuations in overall mosquito population abundance. See the time-varying parameters and weather constraints section below for the definition of *r*(*t*), *ν*_*M*_(*t*), and the weather-driven mosquito growth function, 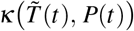

#### Birds

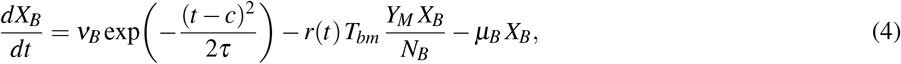

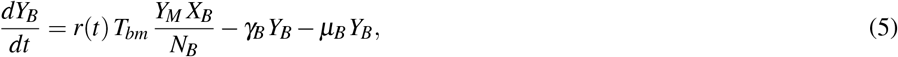

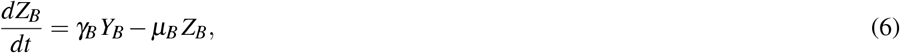

where the Gaussian term forces a seasonal immigration pulse of susceptible birds centered on a day *c* with spread *τ, T*_*bm*_ is the transmission probability from infectious bird to susceptible mosquito per bite, *µ*_*B*_ = 1*/*120 day^−1^ is the bird mortality rate; and *γ*_*B*_ is the bird recovery (viral clearance) rate. We multiplied the population growth rate *ν*_*B*_ by 10 for numerical reasons. A representative figure for the simulation of the susceptible bird population can be found in Supplementary Figure S51.

#### Humans

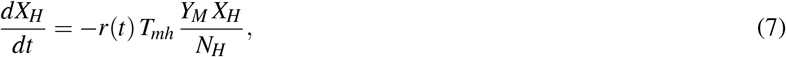

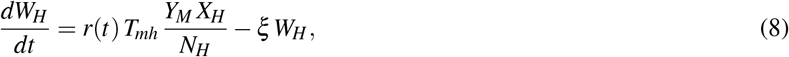

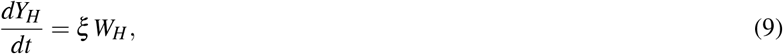

where *N*_*H*_ = *X*_*H*_ + *W*_*H*_ + *Y*_*H*_ is the total human population (fixed at the county-level census estimate for each year), *T*_*mh*_ is the transmission probability from infectious mosquitoes to susceptible humans per bite, and *ξ* is the human incubation rate.

#### Model 2 — full model, no weather (Full Model (no W))

Model 2 retains the full three-host structure but replaces the weather-driven growth term with a time-varying baseline:

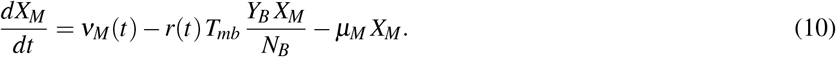

All other compartments follow the same equations as Model 1 (Eqs. 3–9). This configuration isolates the contribution of avian reservoir dynamics from weather forcing.

#### Model 3 — mosquito + human, with weather (M + H)

Model 3 removes the avian reservoir and treats the force of infection as *f* (*t*), a time-varying parameter that substitutes for the bird-to-mosquito transmission term *T*_*mb*_ *Y*_*B*_*/N*_*B*_ estimated by the EnKF. However, the model retains the weather-forced dynamics of the mosquito population growth:

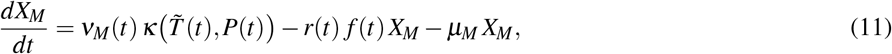

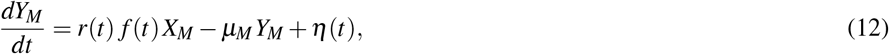

with human dynamics following Eqs. 7–9.

#### Model 4 — mosquito + human, no weather (M + H (no W))

Model 4 is the most parsimonious configuration, removing both the avian reservoir and weather forcing:

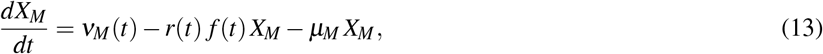

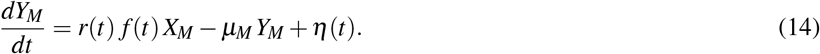

Human dynamics follow Eqs. 7–9. This model serves as the mechanistic baseline against which the added value of weather and bird information is assessed.

#### Time-varying parameters and weather constraints

Three parameters were treated as time-varying across all model configurations: *ν*_*M*_(*t*), the baseline mosquito population growth rate, *f* (*t*), force of infection between mosquitoes and birds in the absence of bird population, and *r*(*t*), the mosquito bite rate (per day, per bird or human). In models incorporating weather, the mosquito growth rate is modulated by both temperature and precipitation. The temperature effect was truncated to zero, so that effects outside of *T*_min_ and *T*_max_ were fixed at zero. The temperature effect is defined as follows:

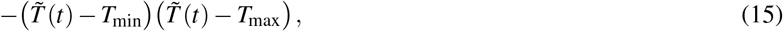

while the precipitation effect is defined as follows:

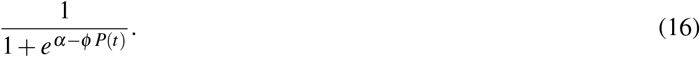

Note that *T*_min_ and *T*_max_ are themselves estimated parameters. Precipitation *P*(*t*) (30-day accumulated, scaled by 50) was used directly as observed. Temperature 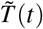 is the daily maximum temperature.

The weather-driven mosquito growth function takes the form [37]:

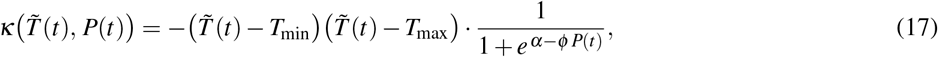

where the quadratic temperature term represents a thermal performance curve of population growth rate controlled by *T*_min_ and *T*_max_, and the logistic precipitation term controlled by constants *α* and *φ* captures a saturating response of growth rate to accumulated rainfall. As precipitation increases, the growth rate increases up to a saturation level; the parameters *α* and *φ* jointly determine at what level of rainfall saturation occurs and how steeply the response rises [37].

### Model initialization and ensemble construction

For each year and model configuration, we initialized an ensemble of *N* = 8,000. State variables and static parameters were drawn from uniform prior distributions spanning biologically plausible ranges (Supplemental table S3). Each year was fitted and forecast independently (i.e., the model had no information retained from prior years), and the human population *N*_*H*_ was fixed at the county-level census estimate for the respective year.

### Observation model

The three surveillance targets assimilated by the EnKF at each weekly update step are derived from the model state variables through the following observation equations:

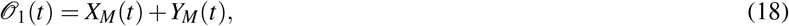

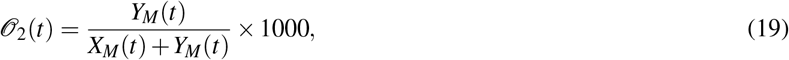

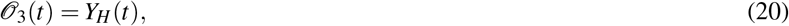

where *O*_1_(*t*) is the predicted total mosquito abundance (corresponding to the observed weekly county-wide trap count), *O*_2_(*t*) is the predicted number of WNV prevalence in mosquitoes, estimated as infections per 1000 (IM1000) (corresponding to the observed IM1000 derived from pool testing data; see Eq. 1), and *O*_3_(*t*) is the predicted cumulative human WNV case count (corresponding to the observed cumulative reported cases). These three quantities constitute the observation vector **y**(*t*) = [*O*_1_(*t*), *O*_2_(*t*), *O*_3_(*t*)]^⊤^ used in the EnKF update step.

The EnKF update is derived under a Gaussian likelihood, so our observation model assumes each of the three targets, total mosquito abundance, WNV prevalence in mosquitoes, and cumulative human cases, is normally distributed around its predicted value, with error variance given by the diagonal entries of *R*_temp_. Every observation is still assumed to be normally distributed around its predicted value. What changes from one observation to the next is the size of that variance. *R*_temp_ scales with the magnitude of each observation and target, so the model tolerates larger absolute errors when observed counts are high (e.g., peak-season mosquito abundance) than when they are low (e.g., early-season case counts). This Gaussian assumption is standard in EnKF applications to epidemiological time series [39, 40] and is convenient for real-time sequential assimilation, but it is still an approximation of the true, discrete count data, which can be overdispersed relative to a Gaussian. We also do not explicitly model underreporting of human cases or overdispersion in mosquito trap counts; both are absorbed, imperfectly, into the scaled variance of *R*_temp_ rather than modeled directly. Replacing this Gaussian observation error with an explicit count-based likelihood, such as a negative binomial model as used in dengue forecasting [51], might be a more principled way to handle these issues and is a natural next step for this framework.

### Data assimilation: Ensemble Kalman Filter (EnKF)

We fitted each dynamical model with the Ensemble Kalman Filter (EnKF) [38, 63, 64] to assimilate weekly surveillance observations and simultaneously estimate model state variables and parameters, following the model-filter framework applied previously to influenza [39, 40] and RSV [40]. The EnKF operates iteratively: the ensemble is integrated forward one week (the *forecast step*), generating a prior distribution over state variables and parameters, and when an observation is available, the filter performs an *update step* using the Kalman gain (see supplementary text S1 for more context).

A key methodological contribution of our framework is the use of an Ornstein-Uhlenbeck (log OU) stochastic process [65] to propagate three time-varying model parameters, the baseline mosquito growth rate *ν*_*M*_(*t*), the force of infection from birds *f* (*t*), and the mosquito bite rate *r*(*t*), forward between assimilation steps. This allows the filter to track gradual seasonal changes in transmission intensity while providing a principled probabilistic basis for generating multi-week-ahead forecasts.

The OU process is a mean-reverting stochastic process defined by the stochastic differential equation. We used the form in [66]:

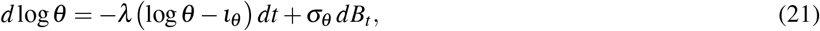

where *θ* ∈ {*ν*_*M*_(*t*), *f* (*t*), *r*(*t*)}, *ι*_*θ*_ is the long-run mean on the log scale, *λ* is the mean-reversion rate, *σ*_*θ*_ is the diffusion coefficient, and *B*_*t*_ is a standard Wiener process. The discrete-time solution used in our implementation is:

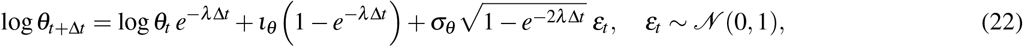

with Δ*t* = 1 days.

During the fitting phase, OU propagation operates at a daily time step (Δ*t* = 1 day) within each weekly assimilation window. At each observation week, seven successive daily OU steps are taken for *ν*_*M*_(*t*), *f* (*t*), and *r*(*t*), producing a sequence of seven daily parameter values. These daily trajectories are passed directly to the ODE model. We provide additional context on how we applied the OU process to our time-varying parameters in Supplementary Text S1 in the supplementary file.

### Probabilistic forecasting

Forecasts were generated at each of the 46 weekly assimilation steps. Following a five-week spin-up period at the start of each year, the EnKF was refitted weekly to all observations available up to that point, and each refit produced 1- and 2-week-ahead forecasts; the window of observations used for fitting therefore grew by one week at each step, so that every forecast was conditioned on the complete observation history up to that point and never had access to future data. At each step, we drew Ω = 1,000 trajectories from the posterior ensemble (out of the full *N* = 8,000 members) and propagated each one forward using the same daily OU process used during fitting: 7 daily steps to reach the 1-week-ahead horizon, then 7 more to reach the 2-week-ahead horizon, each day conditioned on the value from the day before rather than restarting from the weekly posterior. The resulting daily parameter values were passed to the ODE model, which integrated the system one day at a time, producing a 1,000-member forecast ensemble for each target. Forecast distributions were summarized as 23 quantile levels ({0.01, 0.025, 0.05, 0.10,…, 0.95, 0.975, 0.99 }), retaining only ensemble members with physically plausible values (total abundance *>* 0, infectious fraction *<* 100%).

For the weather-forced model configurations (Full Model and M + H), we used the true, observed weather for the forecast period rather than a weather forecast. This is an idealized best case. A real deployment would have to rely on weather forecasts instead, and these are uncertain and become less reliable more than about 10 days out [46]. Future work should test how that added uncertainty carries through to the disease forecasts.

We capped our forecasts at two weeks for two reasons. First, this stays within the roughly 10-day window where weather forecasts are still reliable. Second, mosquito recruitment, infection, and human exposure in our model all play out over days to two weeks, so this is the timescale where a mechanistic model has the most to offer over a purely statistical one.

### Baseline model

All model configurations were evaluated against the symmetrized random-walk baseline model used across the CDC’s FluSight and COVID-19 Forecast Hubs [42, 67, 68]. Historical baseline forecasts derived from empirical case distributions have consistently served as a competitive benchmark in WNV forecast evaluations as well [32, 34]. The baseline is defined as:

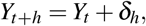

where the median forecast at horizon *h* equals the most recently observed value *Y*_*t*_, and forecast quantiles are derived empirically from the symmetrized distribution of historical weekly differences *d*_*s*_ = *y*_*s*_ − *y*_*s*−1_, truncated to prevent negative case or abundance counts. We implemented this using the `epipredict` R package [69], applying the same 23 quantile levels used for all mechanistic model forecasts, independently to each of the three surveillance targets. For human cases, weekly counts were transformed prior to baseline forecasting to match the cumulative reporting convention used elsewhere; details are in Supplementary Text S1 in the Supplementary Material.

### Evaluation metrics: Weighted Interval Score and relative WIS

Forecast performance was quantified using the Weighted Interval Score (WIS), a proper scoring metric for quantile forecasts that generalizes the mean absolute error [41]. WIS has been adopted as the primary metric in CDC-sponsored forecasting hubs [42]. Given a set of *K* central prediction intervals and observed value *y*, WIS is defined as:

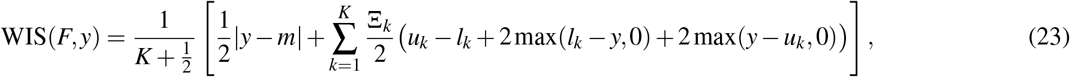

where *m* is the forecast median and (*l*_*k*_, *u*_*k*_) are the lower and upper bounds of the *k*-th central prediction interval at coverage level 1 − Ξ_*k*_ [41, 42]. WIS was computed using the `epipredict` R package [69, 70] using the 23 quantile levels as defined in the probabilistic forecasting section.

To enable comparison across targets, scales, and years, we report the *relative WIS* on the log scale, following the log-transformed relative WIS convention adopted by the CDC’s FluSight influenza forecasting hub [71]:

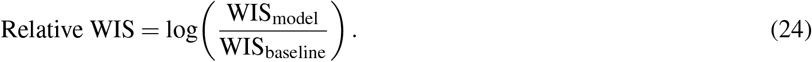

Negative values indicate that a model outperforms the baseline. Relative WIS scores were computed separately for each year, calendar week, season (winter: January-March; spring: April-June; summer: July-September; fall: October-December), and forecast horizon (1- and 2-week ahead). We explained how WIS and log Relative WIS scores are calculated visually in Supplementary Figure S50.

Mosquito infection prevalence (IM1000) is frequently at or near zero outside the transmission season. We restrict WIS summaries to weeks with non-zero observed prevalence when comparing fit across years. We refer to this subset of weeks as the *active transmission season*. Including the many structurally near-zero weeks outside this window causes the median WIS to collapse toward zero regardless of model performance during the transmission season itself, obscuring rather than clarifying year-to-year comparisons. All other WIS summaries reported in this manuscript (e.g., annual and monthly relative WIS (log scale) for forecast evaluation) use all available weeks, including the near-zero season, since those weeks are informative for evaluating forecast calibration even when the target itself is near zero.

### Ensemble combination methods

We evaluated five ensemble combination strategies built from the four individual model configurations: quantile averaging, quantile median, regression-based weighting, WIS-optimized weighting, and a distributional linear pool. The first four follow the definitions of Sherratt et al. [43], adapted here to a within-year retrospective framework: all 46 forecast iterations from a given year were used both to estimate ensemble weights and to evaluate forecast performance, rather than a train/test split. This within-year approach is consistent with the broader rationale for ensembling in the infectious disease forecasting literature, where combined forecasts often outperform individual models on average even when individual models temporarily outperform the ensemble [42, 50, 57]. Full mathematical specifications for all five methods are provided in Supplementary Text S1. Two ensembles combined forecasts without regard to individual model performance: quantile averaging (Ensemble Model 1) and the quantile median (Ensemble Model 2), which differ mainly in their sensitivity to outlier forecasts, with the median offering somewhat greater robustness when one model diverges sharply from the others. Two ensembles instead estimated combination weights from the data: a regression-based ensemble (Ensemble Model 3), which learns unconstrained weights via ordinary least squares, and a WIS-optimized ensemble (Ensemble Model 4), which estimates non-negative, sum-to-one weights by directly minimizing WIS [72]. The fifth approach, the linear pool (Ensemble Model 5), combines the full predictive distributions of the four models rather than their pointwise quantiles, averaging their cumulative distribution functions before inverting to recover quantile forecasts [44, 45, 47].

These five methods differed substantially in how consistently they performed across the three surveillance targets (Supplementary Figures S57–S60, S65-70). The regression ensemble, for example, achieved the best median relative WIS (log scale) of any configuration for human cases but the worst for total abundance, illustrating that unconstrained, target-specific weighting can trade reliability on one target for performance on another. We focus the main text on the WIS-optimized ensemble (Ensemble Model 4) because it was the only method that ranked first or near-first across all three targets simultaneously, with lower year-to-year variability than the best individual model for each. It is this cross-target consistency, rather than peak performance on any single target, that we consider most relevant to an operational system tracking mosquito abundance, infection prevalence, and human cases together. Full pairwise comparisons among all five ensemble methods are provided in Supplementary Text S1 and Supplementary Figures S57–S60, S65-70.

## Supplementary information

The supplementary file provides additional methodological detail, interpretation of results, tables, and figures accompanying the main text.

## Declarations

### Ethics approval and consent to participate

There is no ethical issue related to this research.

### Data availability

The original data that support the findings of this study are available in the GitHub repository https://github.com/NAU-CCL/WNV-Forecasting/tree/main/datasets.

### Code availability

The code used in this study for analyses and to create the tables and figures is publicly available in the GitHub repository https://github.com/NAU-CCL/WNV-Forecasting.

### Author contribution

Conceptualization—K.O. and J.M.; methodology—K.O., J.C., and J.M.; software—K.O.; validation—K.O., J.C., Y.C., E.D., N.B., C.M.H., J.T., J.W., I.R., M.K., and J.M.; formal analysis—K.O.; investigation—K.O., J.C., Y.C., E.D., N.B., C.M.H., J.T., J.W., I.R., M.K., and J.M.; resources—J.M.; data curation—K.O. and J.M.; writing and original draft preparation—K.O. and J.M.; writing—review and editing—K.O., J.C., Y.C., E.D., N.B., C.M.H., J.T., J.W., I.R., M.K., and J.M.; visualization—K.O. and J.M.; supervision—J.M.; project administration—J.M. All authors have read and agreed to the published version of the manuscript.

