## Supplemental File 1 for "Evaluating the roles of weather and bird dynamics in accurately forecasting West Nile virus infection in mosquitoes and humans"

Oshinubi et al.

August 27, 2026

This supplementary material provides additional methodological detail, interpretation of results, tables, and figures accompanying the main text. Section 1 gives full specifications for model components summarized briefly in the main methods. Section 2 gives additional details on the interpretation of results. Section 3 provides parameter, state variable, and calibration tables. Section 4 provides supplementary figures.

**3 Supplementary Tables** 8

**4 Supplementary Figures** 11

#### List of Supplementary Tables

- Table S1: Summary of the four WNV model configurations
- Table S2: State variables of the full model
- Table S3: Model parameters, prior ranges and estimation status
- Table S4: Year-specific OU diffusion parameter $\sigma_{\nu_M}$

### List of Supplementary Figures

#### Model fits (Figures S1–S12)

- Figure S1: Full Model: EnKF posterior fit, all 15 years, total mosquito abundance
- Figure S2: Full Model (no weather): EnKF posterior fit, all 15 years, total mosquito abundance
- Figure S3: Mosquito + Human (with weather): EnKF posterior fit, all 15 years, total mosquito abundance
- Figure S4: Mosquito + Human (no weather): EnKF posterior fit, all 15 years, total mosquito abundance
- Figure S5: Full Model: EnKF posterior fit, all 15 years, infectious mosquitoes per 1000 (IM1000)
- Figure S6: Full Model (no weather): EnKF posterior fit, all 15 years, infectious mosquitoes per 1000 (IM1000)
- Figure S7: Mosquito + Human (with weather): EnKF posterior fit, all 15 years, infectious mosquitoes per 1000 (IM1000)
- Figure S8: Mosquito + Human (no weather): EnKF posterior fit, all 15 years, infectious mosquitoes per 1000 (IM1000)
- Figure S9: Full Model: EnKF posterior fit, all 15 years, cumulative human cases
- Figure S10: Full Model (no weather): EnKF posterior fit, all 15 years, cumulative human cases
- Figure S11: Mosquito + Human (no weather): EnKF posterior fit, all 15 years, cumulative human cases
- Figure S12: Mosquito + Human (with weather): EnKF posterior fit, all 15 years, cumulative human cases

#### Full Model forecasts (Figures S13–S21)

- Figure S13: Full Model: 1-week-ahead forecasts, all 15 years, total mosquito abundance
- Figure S14: Full Model: 2-week-ahead forecasts, all 15 years, total mosquito abundance
- Figure S15: Full Model: 2-week forecast fans, all 15 years, total mosquito abundance
- Figure S16: Full Model: 1-week-ahead forecasts, all 15 years, infectious mosquitoes per 1000 (IM1000)
- Figure S17: Full Model: 2-week-ahead forecasts, all 15 years, infectious mosquitoes per 1000 (IM1000)
- Figure S18: Full Model: 2-week forecast fans, all 15 years, infectious mosquitoes per 1000 (IM1000)
- Figure S19: Full Model: 2-week forecast fans, all 15 years, cumulative human cases
- Figure S20: Full Model: 1-week-ahead forecasts, all 15 years, cumulative human cases
- Figure S21: Full Model: 2-week-ahead forecasts, all 15 years, cumulative human cases

#### Full Model without weather: forecasts (Figures S22–S30)

- Figure S22: Full Model (no weather): 1-week-ahead forecasts, all 15 years, total mosquito abundance
- Figure S23: Full Model (no weather): 2-week-ahead forecasts, all 15 years, total mosquito abundance
- Figure S24: Full Model (no weather): 2-week forecast fans, all 15 years, total mosquito abundance
- Figure S25: Full Model (no weather): 1-week-ahead forecasts, all 15 years, infectious mosquitoes per 1000 (IM1000)
- Figure S26: Full Model (no weather): 2-week-ahead forecasts, all 15 years, infectious mosquitoes per 1000 (IM1000)
- Figure S27: Full Model (no weather): 2-week forecast fans, all 15 years, infectious mosquitoes per 1000 (IM1000)

- Figure S28: Full Model (no weather): 2-week forecast fans, all 15 years, cumulative human cases
- Figure S29: Full Model (no weather): 1-week-ahead forecasts, all 15 years, cumulative human cases
- Figure S30: Full Model (no weather): 2-week-ahead forecasts, all 15 years, cumulative human cases

**Mosquito + Human with weather: forecasts (Figures S31–S39)**

- Figure S31: Mosquito + Human (with weather): 1-week-ahead forecasts, all 15 years, total mosquito abundance
- Figure S32: Mosquito + Human (with weather): 2-week-ahead forecasts, all 15 years, total mosquito abundance
- Figure S33: Mosquito + Human (with weather): 2-week forecast fans, all 15 years, total mosquito abundance
- Figure S34: Mosquito + Human (with weather): 1-week-ahead forecasts, all 15 years, infectious mosquitoes per 1000 (IM1000)
- Figure S35: Mosquito + Human (with weather): 2-week-ahead forecasts, all 15 years, infectious mosquitoes per 1000 (IM1000)
- Figure S36: Mosquito + Human (with weather): 2-week forecast fans, all 15 years, infectious mosquitoes per 1000 (IM1000)
- Figure S37: Mosquito + Human (with weather): 2-week forecast fans, all 15 years, cumulative human cases
- Figure S38: Mosquito + Human (with weather): 1-week-ahead forecasts, all 15 years, cumulative human cases
- Figure S39: Mosquito + Human (with weather): 2-week-ahead forecasts, all 15 years, cumulative human cases

**Mosquito + Human without weather: forecasts (Figures S40–S48)**

- Figure S40: Mosquito + Human (no weather): 1-week-ahead forecasts, all 15 years, total mosquito abundance
- Figure S41: Mosquito + Human (no weather): 2-week-ahead forecasts, all 15 years, total mosquito abundance
- Figure S42: Mosquito + Human (no weather): 2-week forecast fans, all 15 years, total mosquito abundance
- Figure S43: Mosquito + Human (no weather): 1-week-ahead forecasts, all 15 years, infectious mosquitoes per 1000 (IM1000)
- Figure S44: Mosquito + Human (no weather): 2-week-ahead forecasts, all 15 years, infectious mosquitoes per 1000 (IM1000)
- Figure S45: Mosquito + Human (no weather): 2-week forecast fans, all 15 years, infectious mosquitoes per 1000 (IM1000)
- Figure S46: Mosquito + Human (no weather): 2-week forecast fans, all 15 years, cumulative human cases
- Figure S47: Mosquito + Human (no weather): 1-week-ahead forecasts, all 15 years, cumulative human cases
- Figure S48: Mosquito + Human (no weather): 2-week-ahead forecasts, all 15 years, cumulative human cases

**Model components and diagnostics (Figures S49–S54)**

- Figure S49: Temperature and precipitation response functions (2016 data)
- Figure S50: Schematic of the WIS and relative WIS calculation
- Figure S51: Simulated susceptible bird population, Full Model, 2014
- Figure S52: Adaptive OU diffusion parameter $\sigma$ over time, 2014
- Figure S53: Posterior histograms of the static parameters, Full Model, 2014
- Figure S54: Daily ensemble trajectories of the time-varying parameters $\nu_M(t)$ and $r(t)$, 2014

##### Baseline and relative-WIS summaries (Figures S55–S61)

- Figure S55: Baseline model: median WIS by year, 1- and 2-week horizons
- Figure S56: Baseline model: WIS by calendar month
- Figure S57: Median log relative WIS by model and target, 1-week horizon (all nine configurations)
- Figure S58: Median log relative WIS by calendar month, 1-week horizon (all nine configurations)
- Figure S59: Median log relative WIS by model and target, 2-week horizon (all nine configurations)
- Figure S60: Median log relative WIS by calendar month, 2-week horizon (all nine configurations)
- Figure S61: Baseline model: heatmap of median WIS by month and year

##### Fit-WIS and relative-WIS heatmaps, trap network (Figures S62–S71)

- Figure S62: Heatmap of median in-sample fit WIS by model, month and year: human cases
- Figure S63: Heatmap of median in-sample fit WIS by model, month and year: total abundance
- Figure S64: Heatmap of median in-sample fit WIS by model, month and year: IM1000
- Figure S65: Heatmap of median log relative WIS by model, month and year: human cases, 1-week horizon
- Figure S66: Heatmap of median log relative WIS by model, month and year: human cases, 2-week horizon
- Figure S67: Heatmap of median log relative WIS by model, month and year: IM1000, 1-week horizon
- Figure S68: Heatmap of median log relative WIS by model, month and year: IM1000, 2-week horizon
- Figure S69: Heatmap of median log relative WIS by model, month and year: total abundance, 1-week horizon
- Figure S70: Heatmap of median log relative WIS by model, month and year: total abundance, 2-week horizon
- Figure S71: Number of operational mosquito traps in Maricopa County, 2006–2024

##### Baseline model forecasts (Figures S72–S80)

- Figure S72: Baseline model: 1-week-ahead forecasts, all 15 years, total mosquito abundance
- Figure S73: Baseline model: 2-week-ahead forecasts, all 15 years, total mosquito abundance
- Figure S74: Baseline model: 2-week forecast fans, all 15 years, total mosquito abundance
- Figure S75: Baseline model: 1-week-ahead forecasts, all 15 years, infectious mosquitoes per 1000 (IM1000)
- Figure S76: Baseline model: 2-week-ahead forecasts, all 15 years, infectious mosquitoes per 1000 (IM1000)
- Figure S77: Baseline model: 2-week forecast fans, all 15 years, infectious mosquitoes per 1000 (IM1000)
- Figure S78: Baseline model: 2-week forecast fans, all 15 years, cumulative human cases
- Figure S79: Baseline model: 1-week-ahead forecasts, all 15 years, cumulative human cases
- Figure S80: Baseline model: 2-week-ahead forecasts, all 15 years, cumulative human cases

### 1 Supplementary Methods

#### 1.1 Prior distributions for time-varying parameters

Consistent with the prior ranges given in Table S3, the three time-varying parameters, $\nu_M(t)$, $f(t)$, and $r(t)$, were initialized by sampling uniformly on the log scale rather than the natural scale:

$$\log \nu_M(0) \sim \mathscr{U}(\log 0.0001, \log 7), \qquad \log f(0) \sim \mathscr{U}(\log 0.0001, \log 0.001), \qquad \log r(0) \sim \mathscr{U}(\log 0.0001, \log 0.001).$$

#### 1.2 Perturbed-observation Ensemble Kalman Filter update

Perturbed observations were used in the EnKF update step to avoid filter degeneracy [1]. The base observation noise covariance matrix $R$ was specified as a diagonal matrix:

$$R = \text{diag}(5.0,\ 0.001,\ 0.05), \tag{1}$$

where the three diagonal entries correspond to the three simultaneously assimilated targets in order: total mosquito abundance ($X_M + Y_M$), mosquito infection prevalence (IM1000), and cumulative human WNV cases ($Y_H$). The relatively large base noise for total abundance (5.0) reflects the high week-to-week variability in trap counts and imperfect observation of the true countywide mosquito population. The small value for IM1000 (0.001) reflects the fact that this quantity is a derived prevalence estimate and is thus smoother and less subject to sampling noise. The intermediate value for human cases (0.05) reflects moderate reporting uncertainty in weekly case tallies.

At each assimilation step, $R$ was scaled dynamically in proportion to the magnitude of the current observation to produce the step-specific noise matrix $R_{\text{temp}}$:

$$R_{\text{temp}} = \text{diag}(y_1 \cdot R_{11},\ \max(y_2 \cdot R_{22},\ R_{22}),\ \max(y_3 \cdot R_{33},\ R_{33})), \tag{2}$$

where $y_1$, $y_2$, $y_3$ are the current observed values of total abundance, IM1000, and cumulative human cases, respectively, and $R_{ii}$ are the corresponding diagonal entries of $R$. This formulation is analogous to the observation error variance scaling used by Shaman et al. [2] for influenza forecasting. Observation noise grows proportionally with the magnitude of the observation for total abundance, while for IM1000 and human cases a floor equal to the base noise value is enforced via the $\max(\cdot)$ operator. The floor prevents observation noise from collapsing to zero during weeks of near-zero prevalence or case counts, which would otherwise cause the filter to overweight those near-zero observations and destabilize the ensemble.

For each ensemble member $i$, an observation noise vector $\varepsilon_i \sim \mathscr{N}(\mathbf{0}, R_{\text{temp}})$ was drawn independently from the multivariate normal distribution with covariance $R_{\text{temp}}$, and the ensemble update was computed as:

$$\mathbf{x}_i^a = \mathbf{x}_i^f + K\left(\mathbf{y} - \mathbf{H}\mathbf{x}_i^f + \varepsilon_i\right), \tag{3}$$

where $\mathbf{x}_i^f$ is the prior state vector of ensemble member $i$, $\mathbf{y}$ is the observation vector, $\mathbf{H}$ is the observation operator mapping state variables to observed quantities, and $K$ is the Kalman gain:

$$K = P_{xy}^f \left(P_{yy}^f + R_{\text{temp}}\right)^{-1}, \tag{4}$$

where $P_{xy}^f$ is the cross-covariance between the ensemble state and predicted observations, and $P_{yy}^f$ is the variance of predicted observations across ensemble members. The pseudoinverse was used in place of the standard matrix inverse to ensure numerical stability in cases where $P_{yy}^f + R_{\text{temp}}$ is near-singular [1]. The addition of the independently drawn noise vector $\varepsilon_i$ for each ensemble member preserves ensemble spread and prevents collapse of the posterior distribution to a single point estimate.

This stochastic EnKF implementation differs from deterministic variants such as the Ensemble Adjustment Kalman Filter (EAKF) used by Reis and Shaman [3], and was chosen because it naturally accommodates the non-linear mapping between model state variables and the three heterogeneous observation types assimilated simultaneously each week. More computationally intensive filtering approaches, such as particle flow filters [4], offer theoretical advantages for highly non-linear systems but are substantially more demanding to implement and tune at the scale of a 15-year, 8,000-member ensemble. We leave exploration of such alternatives as a direction for future work, particularly for extreme transmission years such as 2021, where the posterior distribution may depart substantially from Gaussianity.

After each update, all state variables and parameters were constrained to remain non-negative; abundances below one were set to zero.

##### 1.3 Ornstein-Uhlenbeck process: daily propagation and adaptive diffusion

The diffusion parameter $\sigma_r$ for the bite rate $r(t)$ was set adaptively at each weekly assimilation step: a low value ($\sigma_r = 0.002$, long-run mean $\iota_r = \log(0.001)$) was used during weeks prior to a year-specific onset index $i_{\text{year}}$, at which point meaningful mosquito activity is not yet expected, switching to a higher value ($\sigma_r = 1.50$, $\iota_r = \log(0.05)$) once sufficient mosquito activity is detected. The same adaptive value was applied uniformly across all seven daily OU steps within each observation week. The same switching schedule was applied to $f(t)$ in Models 3 and 4. This adaptive schedule allowed the filter to remain numerically stable during periods of near-zero transmission while responding rapidly to emerging outbreaks. A representative figure of the diffusion parameter is shown in Supplementary Figure S52.

In Models 3 and 4, which lack an explicit avian reservoir, $f(t)$ is estimated jointly with $v_M(t)$ and $r(t)$ by the EnKF as a data-driven substitute for the bird-to-mosquito transmission term. Biologically, $f(t)$ captures the combined effect of infectious bird density and mosquito-bird contact rate that would otherwise be generated mechanistically by the bird compartment in Models 1 and 2. We applied the same OU propagation framework to $f(t)$ as to $r(t)$, using identical OU parameters ($\lambda$, $\iota_r$, $\sigma_r$) and the same adaptive switching schedule based on $i_{\text{year}}$:

$$\log f(t+\Delta t) = \log f(t)\, e^{-\lambda \Delta t} + \iota_r \left(1 - e^{-\lambda \Delta t}\right) + \sigma_r \sqrt{1 - e^{-2\lambda \Delta t}}\, \varepsilon_t, \quad \varepsilon_t \sim \mathcal{N}(0,1). \tag{5}$$

This formulation allows Models 3 and 4 to capture the seasonal dynamics of avian-to-mosquito transmission pressure in a data-driven manner, even in the absence of explicit bird compartments, while maintaining a consistent probabilistic framework across all four model configurations. In total, Models 1 and 2 estimate two time-varying parameters ($v_M(t)$ and $r(t)$), while Models 3 and 4 estimate three ($v_M(t)$, $r(t)$, and $f(t)$).

The diffusion parameter governing $v_M(t)$, denoted $\sigma_{v_M}$, was treated differently across model configurations. For models without an explicit avian reservoir (Models 3 and 4), $\sigma_{v_M}$ was fixed at 3.10 across all years, reflecting the greater process noise needed for the filter to track mosquito population dynamics in the absence of bird-mediated amplification signals. For models that include bird dynamics (Models 1 and 2), $\sigma_{v_M}$ was allowed to vary by year, taking values between 1.10 and 2.00 depending on the observed epidemiological characteristics of each year (Table S4). Years with historically higher transmission intensity or more variable mosquito dynamics (e.g., 2007, 2009, 2011, 2012, 2018, 2019) were assigned higher diffusion values ($\sigma_{v_M} = 1.50$), while years with more stable dynamics were assigned lower values ($\sigma_{v_M} = 1.10$), with 2021 assigned an intermediate value of 2.00. This year-specific calibration reflects the fact that, in models with bird compartments, the additional state variables provide informative signals that reduce the need for large process noise in the mosquito growth rate, while year-to-year variation in monsoon intensity and bird migratory timing still necessitates modest adjustment of the diffusion parameter across the study period.

##### 1.4 Baseline model: transformation to cumulative human case forecasts

For human WNV cases, an additional processing step was required to align the baseline forecast with the cumulative case series used as the observational target in our analysis. Because the baseline model generates forecasts of weekly incident cases at each assimilation step, the baseline forecast distribution at each forecast horizon was first constructed on the incident (weekly) scale and then transformed to the cumulative scale by summing forecasted incident cases from the start of the year through the forecast target week.

Specifically, at each assimilation step $t^*$, the baseline forecast quantiles for the target week $t^* + h$ (where $h \in \{1,2\}$) were computed as the empirical quantiles of the cumulative case distribution obtained by adding the historical weekly incidence draws to the running cumulative total observed up to week $t^*$. This cumulation was applied independently at each quantile level from 0.01 to 0.99:

$$\hat{C}_{t^*+h}^{(\tau)} = C_{t^*} + \sum_{k=1}^{h} \hat{I}_{t^*+k}^{(\tau)}, \quad \tau \in \{0.01, 0.025, \ldots, 0.975, 0.99\}, \tag{6}$$

where $C_{t^*}$ is the observed cumulative case count up to the current assimilation week, $\hat{I}_{t^*+k}^{(\tau)}$ is the $\tau$-th quantile of the baseline forecast for incident cases at week $t^*+k$, and $\hat{C}_{t^*+h}^{(\tau)}$ is the resulting cumulative forecast quantile at the target week. WIS was then computed by evaluating these cumulative forecast quantiles against the observed cumulative case total at week $t^*+h$, ensuring that the baseline and mechanistic model forecasts are assessed on a common scale and that all WIS scores are directly comparable across model configurations. For total mosquito abundance and IM1000, no such transformation was necessary, as both targets are expressed on a non-cumulative weekly scale.

#### 1.5 Ensemble model construction

**Ensemble model 1.** **Quantile averaging** [5]: $\hat{q}_{i,\tau}^{\text{avg}} = \frac{1}{4}\sum_{k=1}^{4} \hat{q}_{i,\tau}^{(k)}$.

**Ensemble model 2.** **Quantile median** [5]: $\hat{q}_{i,\tau}^{\text{med}} = \text{median}_k\, \hat{q}_{i,\tau}^{(k)}$.

**Ensemble model 3.** **Regression ensemble** [5]: ordinary least-squares regression of observed values on the four model median forecasts, estimated using all 46 forecast iterations from the same year, with the resulting intercept and coefficients applied at each quantile level.

**Ensemble model 4.** **WIS-optimized ensemble** [5]: a constrained linear combination whose weights minimize total WIS over all available observations from the same year, subject to non-negativity and unit-sum constraints. Weights were optimized using the `nloptr` package [6] with the COBYLA algorithm.

**Ensemble model 5.** **Linear pool** [7–9]: if $F_k(x)$ denotes the CDF implied by model $k$'s quantile forecast, the pooled CDF is $F^{\text{LP}}(x) = \frac{1}{4}\sum_{k=1}^{4} F_k(x)$. CDF inversion to recover quantile forecasts was performed by linear interpolation on a grid of 500 points spanning the range of all model forecasts.

Unlike Sherratt et al. [5], who estimated ensemble weights on a training set and evaluated on a held-out test set, our retrospective framework estimated and evaluated weights using the same within-year data. This constitutes a form of in-sample optimization and may modestly overstate the performance of the regression and WIS-optimized ensembles relative to a truly prospective evaluation, a limitation we return to in the Discussion in the main text.

#### 1.6 Weather constraint and mosquito infection importation term

##### Temperature truncation

$$\tilde{T}(t) = \begin{cases} T(t) & \text{if } T_{\min} \le T(t) \le T_{\max}, \\ T_{\min} & \text{otherwise.} \end{cases} \tag{7}$$

Precipitation $P(t)$ is 30-day accumulated rainfall scaled by 50 [10]. The combined weather growth function is: $\kappa(\tilde{T}(t), P(t)) = -(\tilde{T}(t) - T_{\min})(\tilde{T}(t) - T_{\max}) \cdot [1 + e^{\alpha - \phi P(t)}]^{-1}$, as defined in the main text. Supplementary Figure S49 illustrates this function using 2016 temperature and precipitation data.

##### Mosquito infection importation term

$$\eta(t) = \begin{cases} 0.01 & \text{if } i_{\text{year}} \le t \le 180, \\ 0 & \text{otherwise,} \end{cases} \tag{8}$$

where $i_{\text{year}}$ is a year-specific onset day (ranging from day 25 to day 100) capturing interannual variation in WNV reintroduction via migratory birds, following the kind of description in [11].

### 2 Supplementary Results

#### 2.1 Association between monsoon precipitation and IM1000 forecast performance

To test whether year-to-year variation in monsoon precipitation explains the degree to which the non-avian model configurations (M + H and M + H (no W)) underperformed the historical baseline for IM1000, we computed monsoon-season cumulative precipitation for each study year and correlated it with each model's annual IM1000 forecast performance.

Monsoon-season precipitation was defined as the sum of daily precipitation from June through September for a given year, using the same daily precipitation series described in Methods. For each model $\times$ year combination, we computed the median relative WIS across all IM1000 forecast iterations and horizons within that year (the same annual summary statistic used to construct Figure 3 in the main text). We then tested the association between annual monsoon precipitation and annual median relative WIS across the 15 study years using Spearman's rank correlation, separately for each of the two non-avian configurations. Spearman's correlation was used rather than Pearson's because it does not assume a linear relationship between precipitation and forecast performance, which is a more conservative choice given the modest sample size ($n = 15$ years) and no strong prior expectation of linearity.

#### 2.2 Ensemble performance by surveillance target

Ensemble Model 3 achieved the lowest median relative WIS of any model configuration for human cases ($-1.20$, rank 1 of 9), outperforming Ensemble Model 4 by 0.05 log-units. However, this advantage came at a substantial cost in year-to-year reliability: Ensemble Model 3 had the highest year-to-year variability of any of the nine configurations for human cases (SD $= 0.713$, compared to 0.433 for Ensemble Model 4), meaning that its superior average performance masked considerable instability across individual years. More critically, Ensemble Model 3 ranked last of all nine configurations for total abundance (median relative WIS: 0.429), the only ensemble method to fail to outperform the historical baseline for that target.

For IM1000, a three-tier performance structure was apparent among the nine configurations. Ensemble Model 4 and Full Model formed a clearly superior tier (median relative WIS 0.148 and 0.150), performing roughly three times better than the second-tier configuration (Full Mode (no W), 0.483) and an order of magnitude better than the remaining ensemble methods (Ensemble Models 1, 2, and 5: 1.77–2.16; Ensemble Model 3: 0.774). This tier structure maps directly onto the bird-dynamics findings described earlier. The configurations that perform best for IM1000 are precisely those that give the greatest weight to the bird-inclusive individual models, which themselves dominate IM1000 forecasting performance. For Ensemble Model 4, this weighting emerges implicitly from WIS optimization, concentrating weight on Full Model; for Ensemble Model 3, partial weighting toward bird-inclusive configurations still yields substantially better IM1000 performance than simple averaging (rank 4 vs. ranks 5–7 for Ensemble Models 2, 5, and 1).

For total abundance, Ensemble Models 1, 2, 4, and 5 all beat the historical baseline (median relative WIS: $-0.0388$ to $-0.137$) with comparable and lower year-to-year variability than the best individual model (SD: 0.093–0.097 vs. 0.113). The exception was again Ensemble Model 3, which was the worst-performing configuration of all nine, consistent with its instability across targets.

We therefore highlight Ensemble Model 4 as the recommended ensemble approach for a multi-target operational system monitoring mosquito abundance, infection prevalence, and human cases simultaneously. It achieves rank 1 on two of the three targets and near-rank 1 with substantially lower year-to-year variability on the third, without the target-specific catastrophic failures observed in methods that are optimized for a narrower objective. Ensemble Models 1 and 2, while slightly worse on average, offer a viable lower-overhead alternative for the two targets where individual model disagreement is modest (human cases and total abundance) but are not recommended for IM1000, where the performance gap relative to Ensemble Model 4 exceeds an order of magnitude.

### 3 Supplementary Tables

**Table S1:** Summary of the four WNV model configurations evaluated in this study.

| | **Model 1** Full Model | **Model 2** Full, No Weather | **Model 3** Mosq+Human+Weather | **Model 4** Mosq+Human |
|---|---|---|---|---|
| Mosquito dynamics | ✓ | ✓ | ✓ | ✓ |
| Bird reservoir | ✓ | ✓ | × | × |
| Weather forcing | ✓ | × | ✓ | × |
| Human dynamics | ✓ | ✓ | ✓ | ✓ |
| State variables | 8 | 8 | 5 | 5 |
| Static parameters | 12 | 8 | 6 | 2 |
| Time-varying parameters | 2 | 2 | 3 | 3 |

**Table S2:** State variables for the full model (Model 1). Subsets used in Models 2–4 are as described in Table S1.

| **Variable** | **Description** | **Host** |
|---|---|---|
| $X_M$ | Susceptible mosquitoes | Mosquito |
| $Y_M$ | Infectious mosquitoes | Mosquito |
| $X_B$ | Susceptible birds | Bird |
| $Y_B$ | Infectious birds | Bird |
| $Z_B$ | Recovered (immune) birds | Bird |
| $X_H$ | Susceptible humans | Human |
| $W_H$ | Exposed humans | Human |
| $Y_H$ | Infectious humans (cumulative) | Human |

**Table S3:** Model parameters, prior ranges for ensemble initialisation, and estimation status. TV = time-varying (EnKF + OU process); Static = estimated by EnKF, fixed within a week.

| **Param.** | **Description** | **Prior range** | **Units** | **Type** | **Models** |
|---|---|---|---|---|---|
| $\nu_M(t)$[a] | Baseline mosquito growth rate | $\mathscr{U}(0.0001, 7)$ | day$^{-1}$ | TV | 1–4 |
| $r(t)$ | Mosquito bite rate | $\mathscr{U}(0.0001, 0.001)$ | day$^{-1}$ | TV | 1–4 |
| $f(t)$ | Force of infection substituting for bird-to-mosquito transmission | $\mathscr{U}(0.0001, 0.001)$ | unitless | TV | 3–4 |
| $T_{\min}$ | Min. temp. for growth | $\mathscr{U}(17, 20)$ | °C | Static | 1, 3 |
| $T_{\max}$ | Max. temp. for growth | $\mathscr{U}(42, 48)$ | °C | Static | 1, 3 |
| $\alpha$ | Precip. effect shape | $\mathscr{U}(0.7, 1.8)$ | unitless | Static | 1, 3 |
| $\phi$ | Precip. effect scale | $\mathscr{U}(0.95, 1.8)$ | unitless | Static | 1, 3 |
| $T_{mb}$ | Mosq.→bird transmission prob. | $\mathscr{U}(0.14, 0.55)$ | unitless | Static | 1–2 |
| $T_{bm}$ | Bird→mosq. transmission prob. | $\mathscr{U}(0.14, 0.55)$ | unitless | Static | 1–2 |
| $T_{mh}$ | Mosq.→human transmission prob. | $\mathscr{U}(0.014, 0.055)$ | unitless | Static | 1–4 |
| $\nu_B$[b] | Bird baseline growth rate | $\mathscr{U}(0.1, 0.9)$ | day$^{-1}$ | Static | 1–2 |
| $c$ | Bird peak arrival day | $\mathscr{U}(180, 200)$ | day | Static | 1–2 |
| $\tau$ | Bird arrival pulse width | $\mathscr{U}(300, 420)$ | day$^2$ | Static | 1–2 |
| $\gamma_B$ | Bird recovery rate | $\mathscr{U}(0.1, 0.2)$ | day$^{-1}$ | Static | 1–2 |
| $\xi$ | Human incubation rate | $\mathscr{U}(0.05, 0.14)$ | day$^{-1}$ | Static | 1–4 |

[a] The full model prior for $\nu_M(t)$ is $\mathscr{U}(0.0001, 2)$. [b]Sampled $\nu_B$ is multiplied by 10 before entering Eq. 4 in the main text, for numerical reasons.

**Table S4:** Year-specific diffusion parameter $\sigma_{\nu_M}$ for the Ornstein-Uhlenbeck process governing the baseline mosquito growth rate $\nu_M(t)$ in models with weather (Models 1 and 3). Models without weather (Models 2 and 4) used a fixed value of $\sigma_{\nu_M} = 3.10$ across all years.

| **Year** | $\sigma_{\nu_M}$ |
|---|---|
| 2006 | 1.10 |
| 2007 | 1.50 |
| 2008 | 1.10 |
| 2009 | 1.50 |
| 2010 | 1.10 |
| 2011 | 1.50 |
| 2012 | 1.50 |
| 2013 | 1.10 |
| 2014 | 1.10 |
| 2015 | 1.10 |
| 2016 | 1.10 |
| 2017 | 1.10 |
| 2018 | 1.50 |
| 2019 | 1.50 |
| 2021 | 2.00 |

### 4 Supplementary Figures

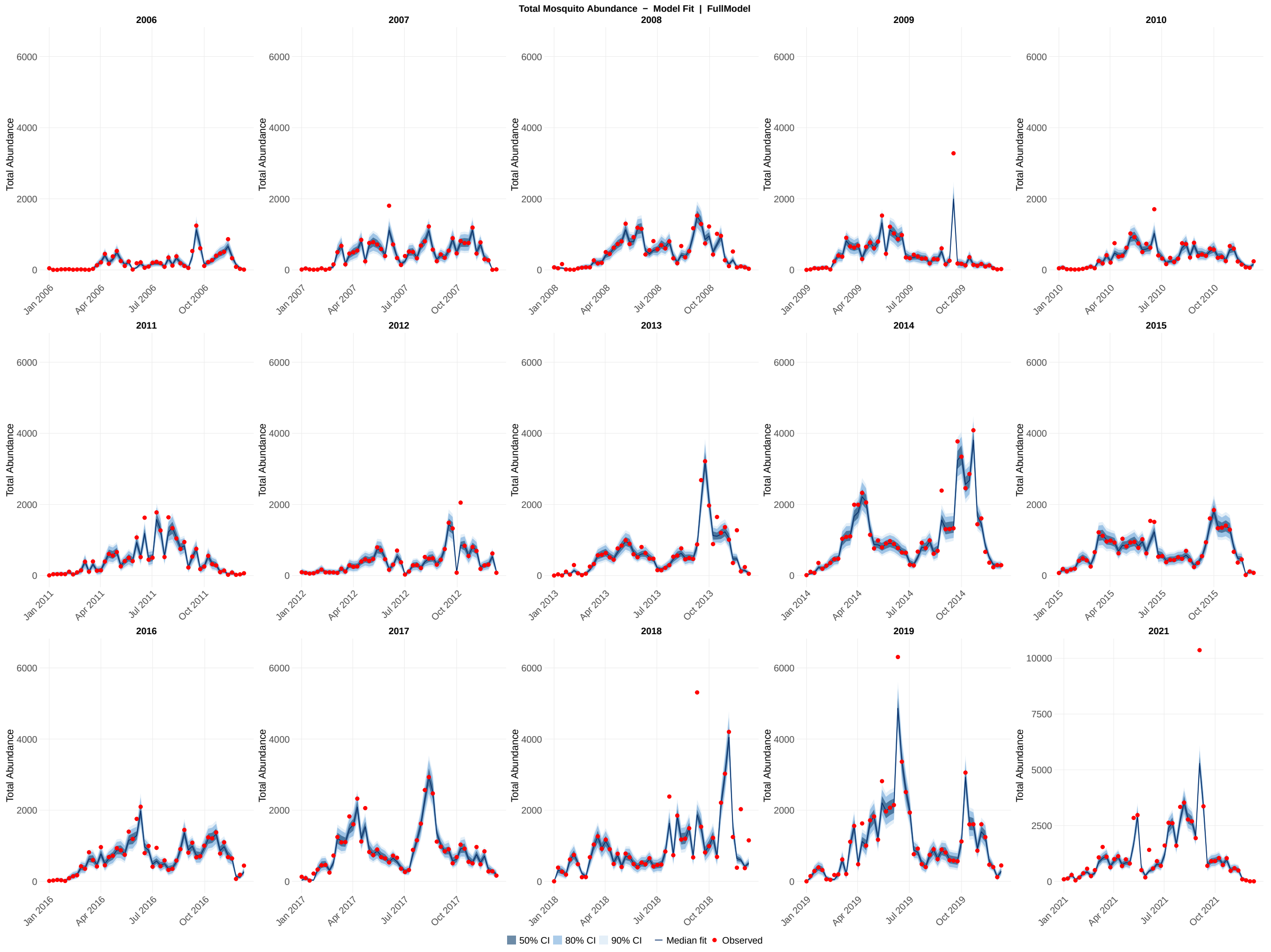

**Figure S1:** EnKF posterior model fit for total abundance across all 15 study years (2006–2019, 2021) for the full model configuration. Each panel shows the EnKF posterior distribution for a single calendar year. Shaded ribbons indicate the 50% (darkest), 80% (medium), and 90% (lightest) credible intervals of the posterior ensemble; the solid line shows the posterior median; red points show the observed weekly human case values. The y-axis scale for 2021 is unconstrained to accommodate the atypically high values observed that year; all other years share a common axis scale (0–6000 for total abundance; adjust as appropriate for other targets). Fits were generated over each year using the Ensemble Kalman Filter with Ornstein-Uhlenbeck parameter propagation.

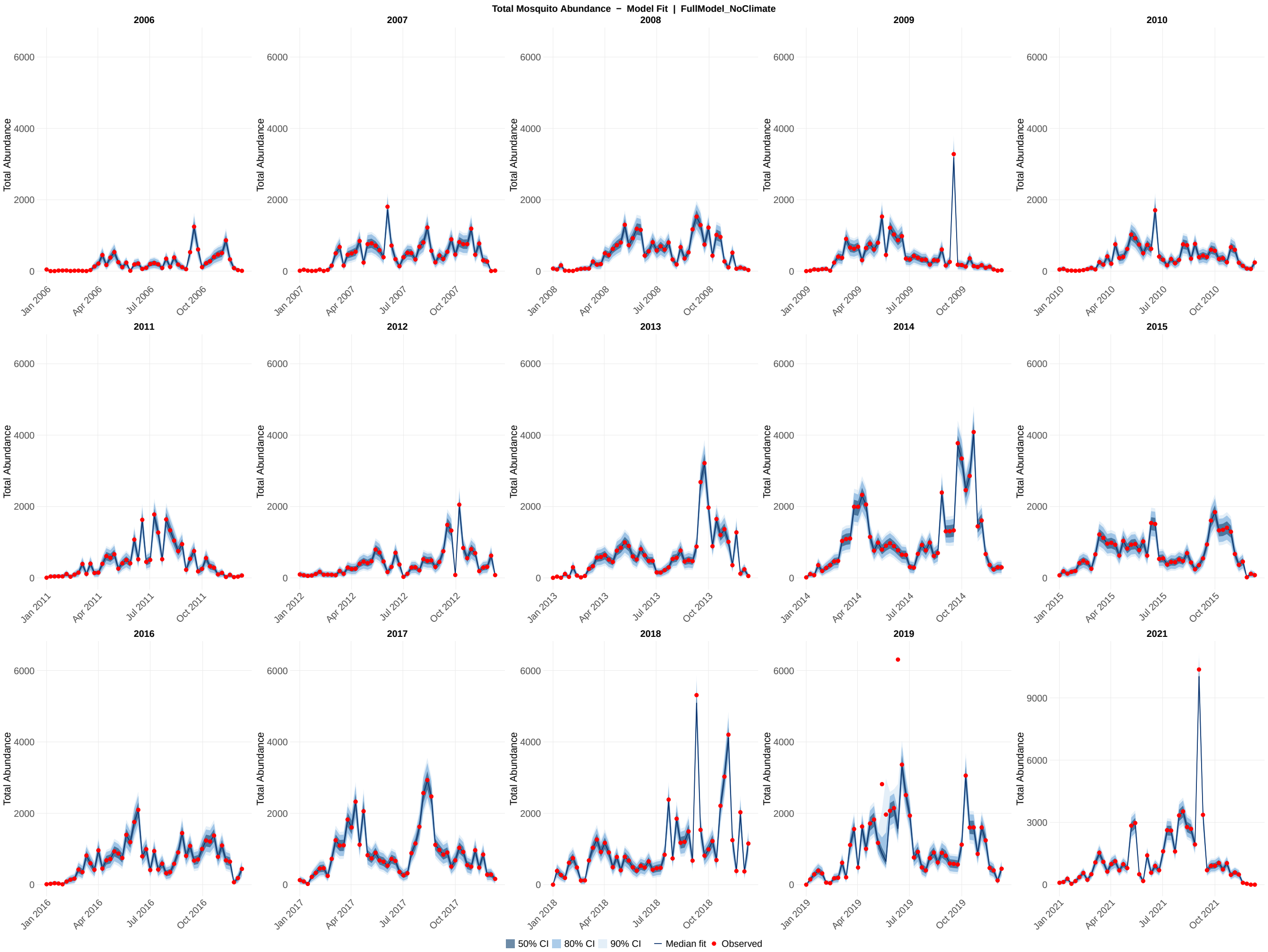

**Figure S2:** EnKF posterior model fit for total abundance across all 15 study years (2006–2019, 2021) for the full model without weather configuration. Each panel shows the EnKF posterior distribution for a single calendar year. Shaded ribbons indicate the 50% (darkest), 80% (medium), and 90% (lightest) credible intervals of the posterior ensemble; the solid line shows the posterior median; red points show the observed weekly human case values. The y-axis scale for 2021 is unconstrained to accommodate the atypically high values observed that year; all other years share a common axis scale (0–6000 for total abundance; adjust as appropriate for other targets). Fits were generated over each year using the Ensemble Kalman Filter with Ornstein-Uhlenbeck parameter propagation.

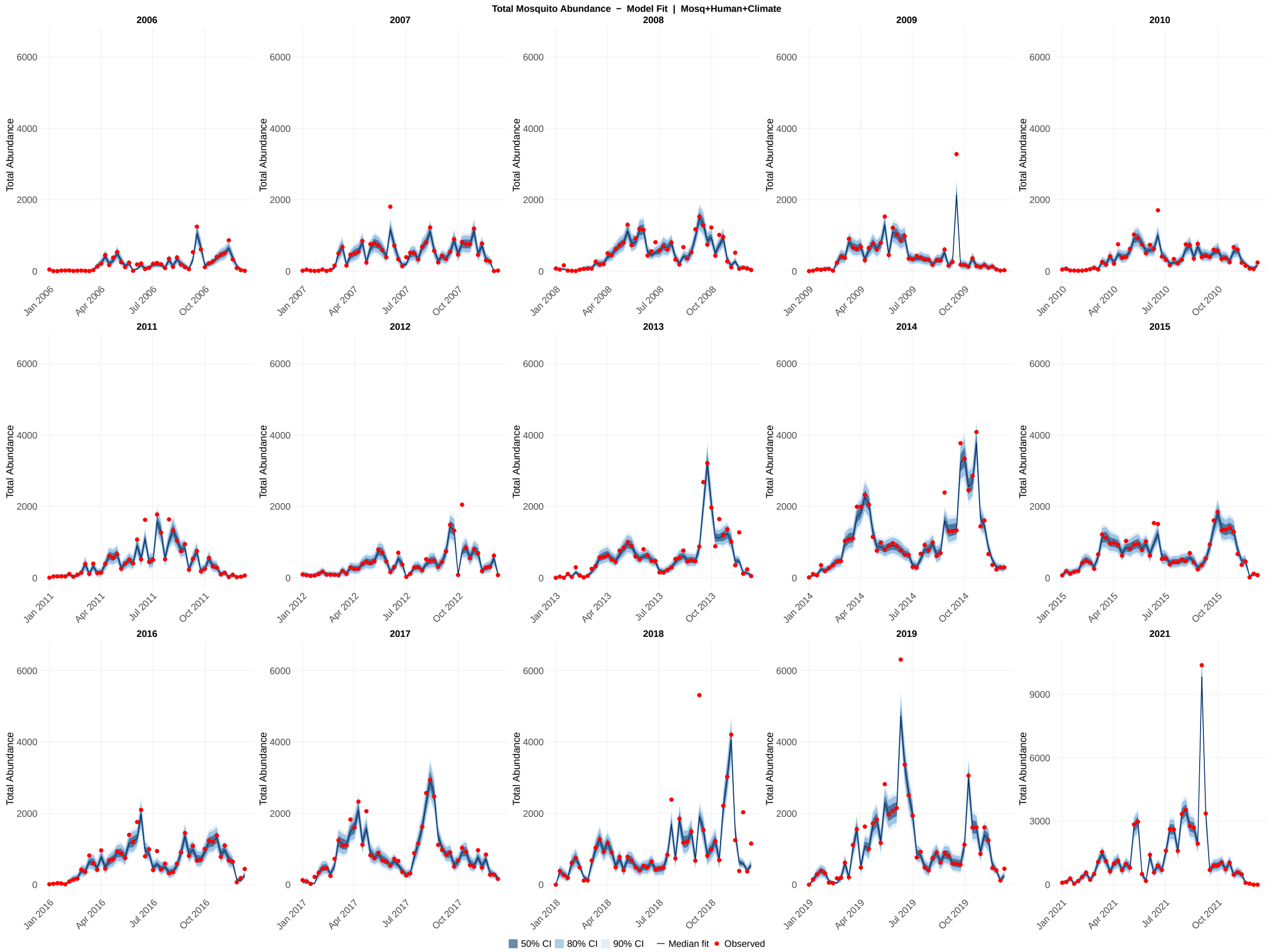

**Figure S3:** EnKF posterior model fit for total abundance across all 15 study years (2006–2019, 2021) for the mosquito + human with weather model configuration. Each panel shows the EnKF posterior distribution for a single calendar year. Shaded ribbons indicate the 50% (darkest), 80% (medium), and 90% (lightest) credible intervals of the posterior ensemble; the solid line shows the posterior median; red points show the observed weekly human case values. The y-axis scale for 2021 is unconstrained to accommodate the atypically high values observed that year; all other years share a common axis scale (0–6000 for total abundance; adjust as appropriate for other targets). Fits were generated over each year using the Ensemble Kalman Filter with Ornstein-Uhlenbeck parameter propagation.

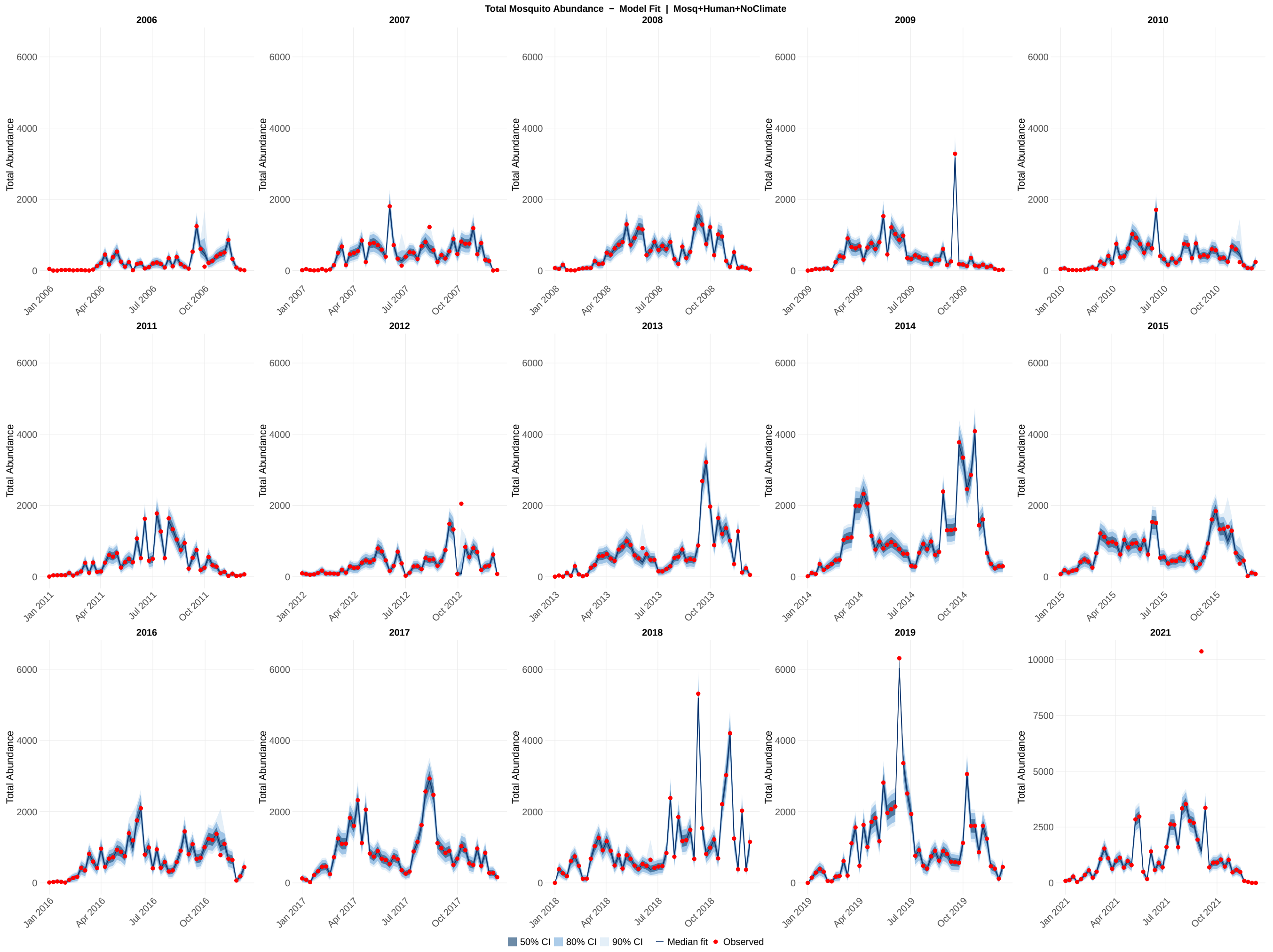

**Figure S4:** EnKF posterior model fit for total abundance across all 15 study years (2006–2019, 2021) for the mosquito + human without weather model configuration. Each panel shows the EnKF posterior distribution for a single calendar year. Shaded ribbons indicate the 50% (darkest), 80% (medium), and 90% (lightest) credible intervals of the posterior ensemble; the solid line shows the posterior median; red points show the observed weekly human case values. The y-axis scale for 2021 is unconstrained to accommodate the atypically high values observed that year; all other years share a common axis scale (0–6000 for total abundance; adjust as appropriate for other targets). Fits were generated over each year using the Ensemble Kalman Filter with Ornstein-Uhlenbeck parameter propagation.

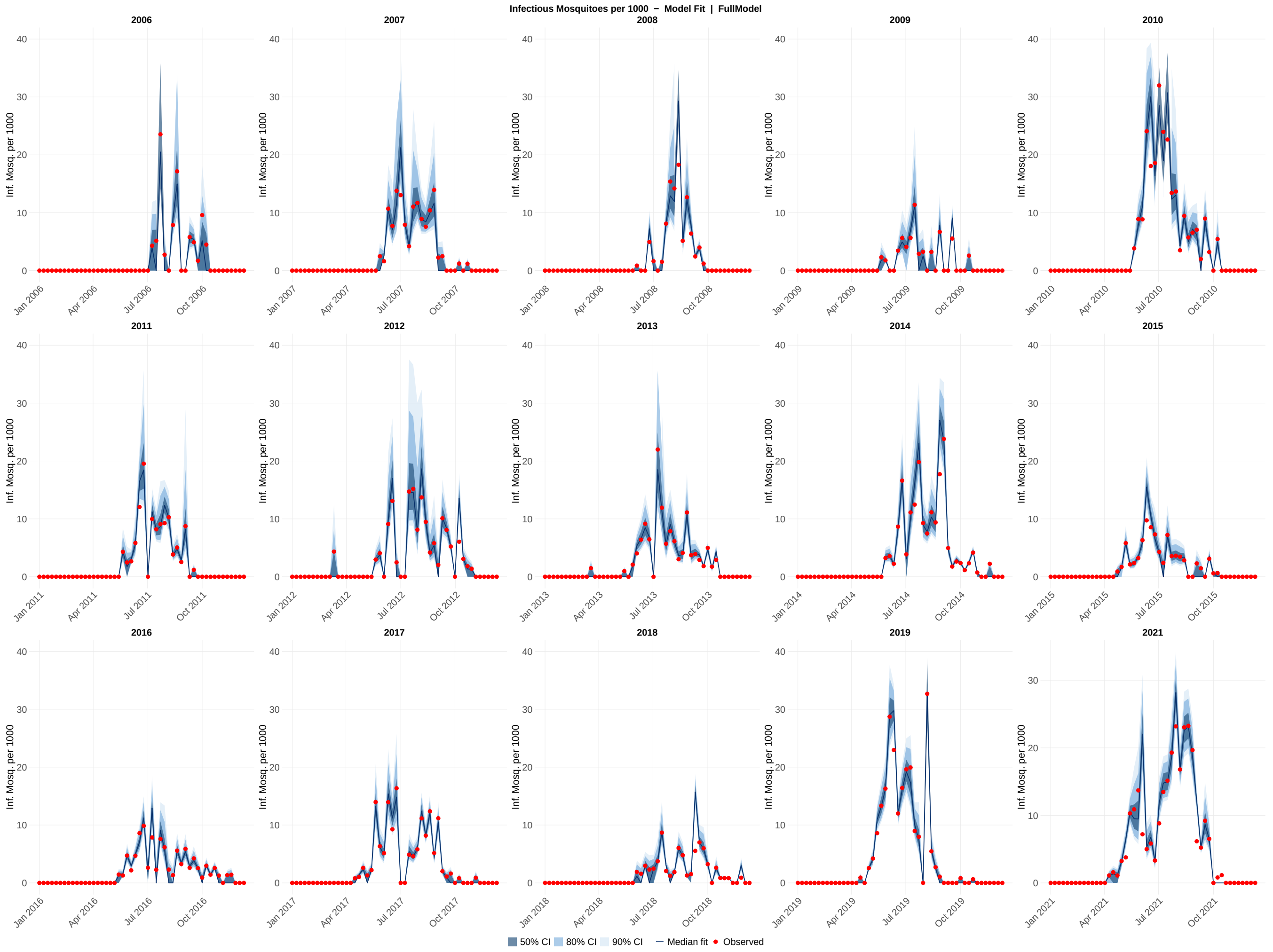

**Figure S5:** EnKF posterior model fit for infectious mosquito per 1000 across all 15 study years (2006–2019, 2021) for the full model configuration. Each panel shows the EnKF posterior distribution for a single calendar year. Shaded ribbons indicate the 50% (darkest), 80% (medium), and 90% (lightest) credible intervals of the posterior ensemble; the solid line shows the posterior median; red points show the observed weekly infectious mosquito per 1000 values. The y-axis scale for 2021 is unconstrained to accommodate the atypically high values observed that year; all other years share a common axis scale (0–40 for infectious mosquito per 1000; adjust as appropriate for other targets). Fits were generated over each year using the Ensemble Kalman Filter with Ornstein-Uhlenbeck parameter propagation.

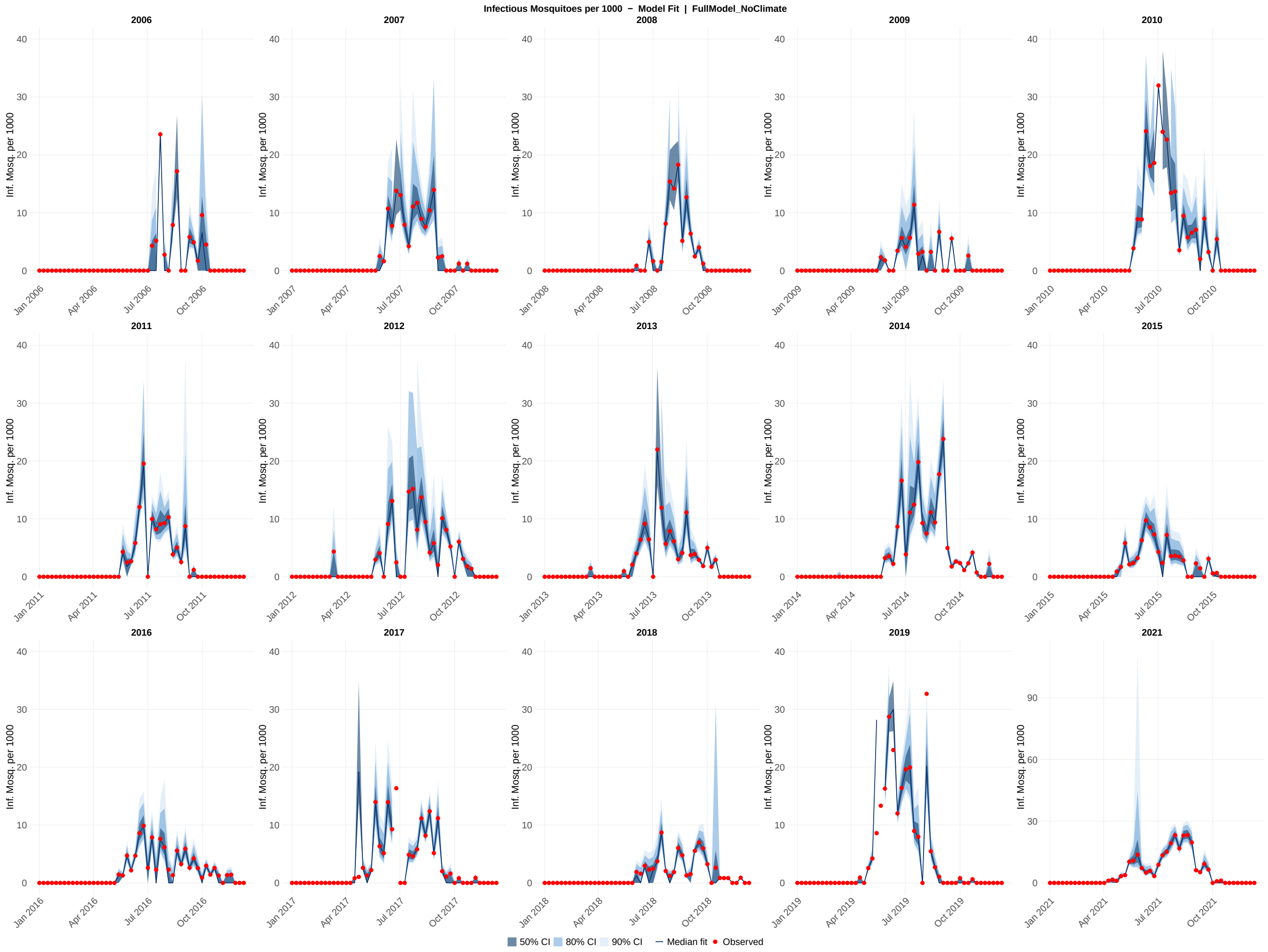

**Figure S6:** EnKF posterior model fit for infectious mosquito per 1000 across all 15 study years (2006–2019, 2021) for the full model without weather configuration. Each panel shows the EnKF posterior distribution for a single calendar year. Shaded ribbons indicate the 50% (darkest), 80% (medium), and 90% (lightest) credible intervals of the posterior ensemble; the solid line shows the posterior median; red points show the observed weekly infectious mosquito per 1000 values. The y-axis scale for 2021 is unconstrained to accommodate the atypically high values observed that year; all other years share a common axis scale (0–40 for infectious mosquito per 1000; adjust as appropriate for other targets). Fits were generated over each year using the Ensemble Kalman Filter with Ornstein-Uhlenbeck parameter propagation.

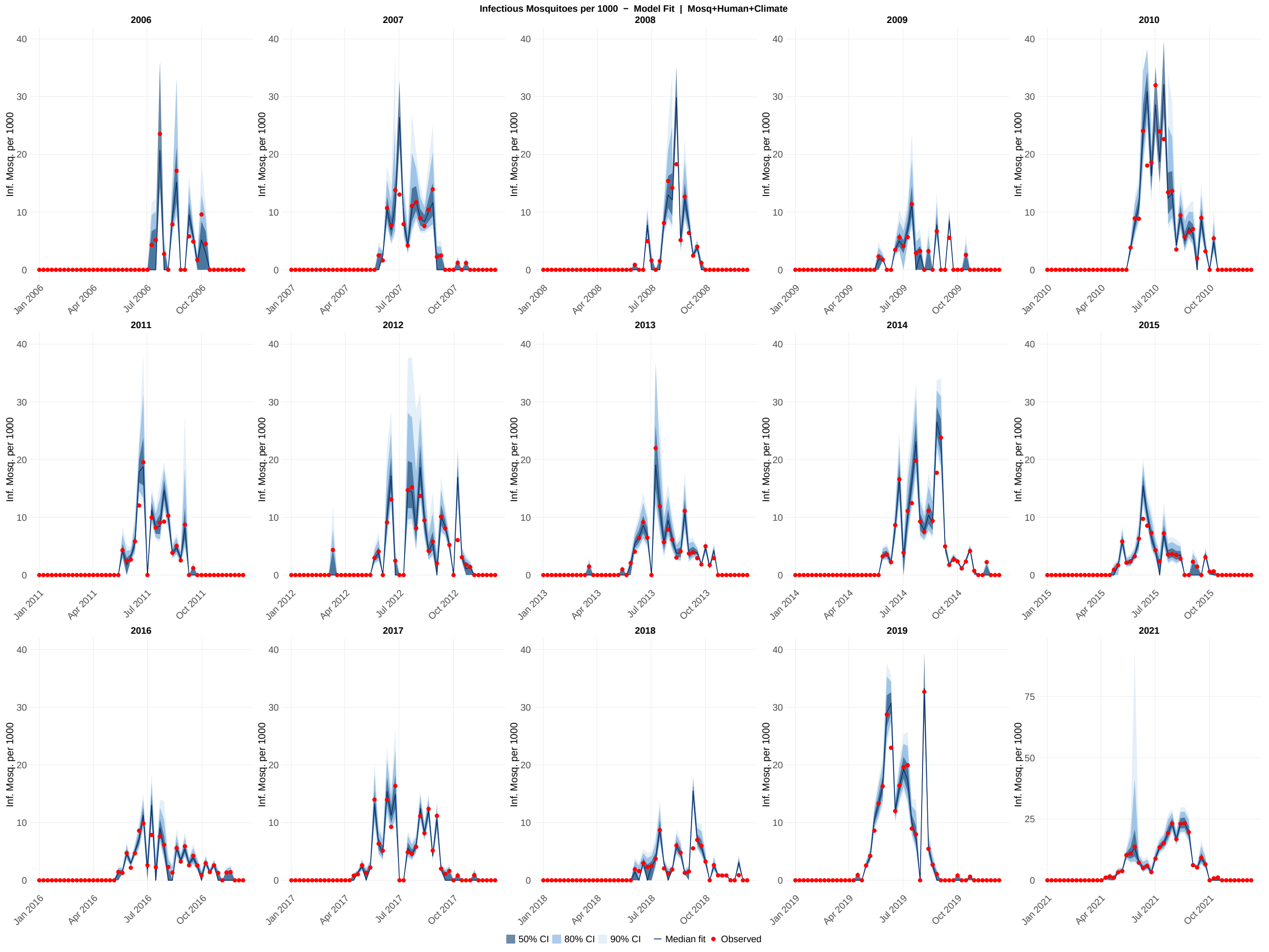

**Figure S7:** EnKF posterior model fit for infectious mosquito per 1000 across all 15 study years (2006–2019, 2021) for the mosquito + human with weather model configuration. Each panel shows the EnKF posterior distribution for a single calendar year. Shaded ribbons indicate the 50% (darkest), 80% (medium), and 90% (lightest) credible intervals of the posterior ensemble; the solid line shows the posterior median; red points show the observed weekly infectious mosquito per 1000 values. The y-axis scale for 2021 is unconstrained to accommodate the atypically high values observed that year; all other years share a common axis scale (0–40 for infectious mosquito per 1000; adjust as appropriate for other targets). Fits were generated over each year using the Ensemble Kalman Filter with Ornstein-Uhlenbeck parameter propagation.

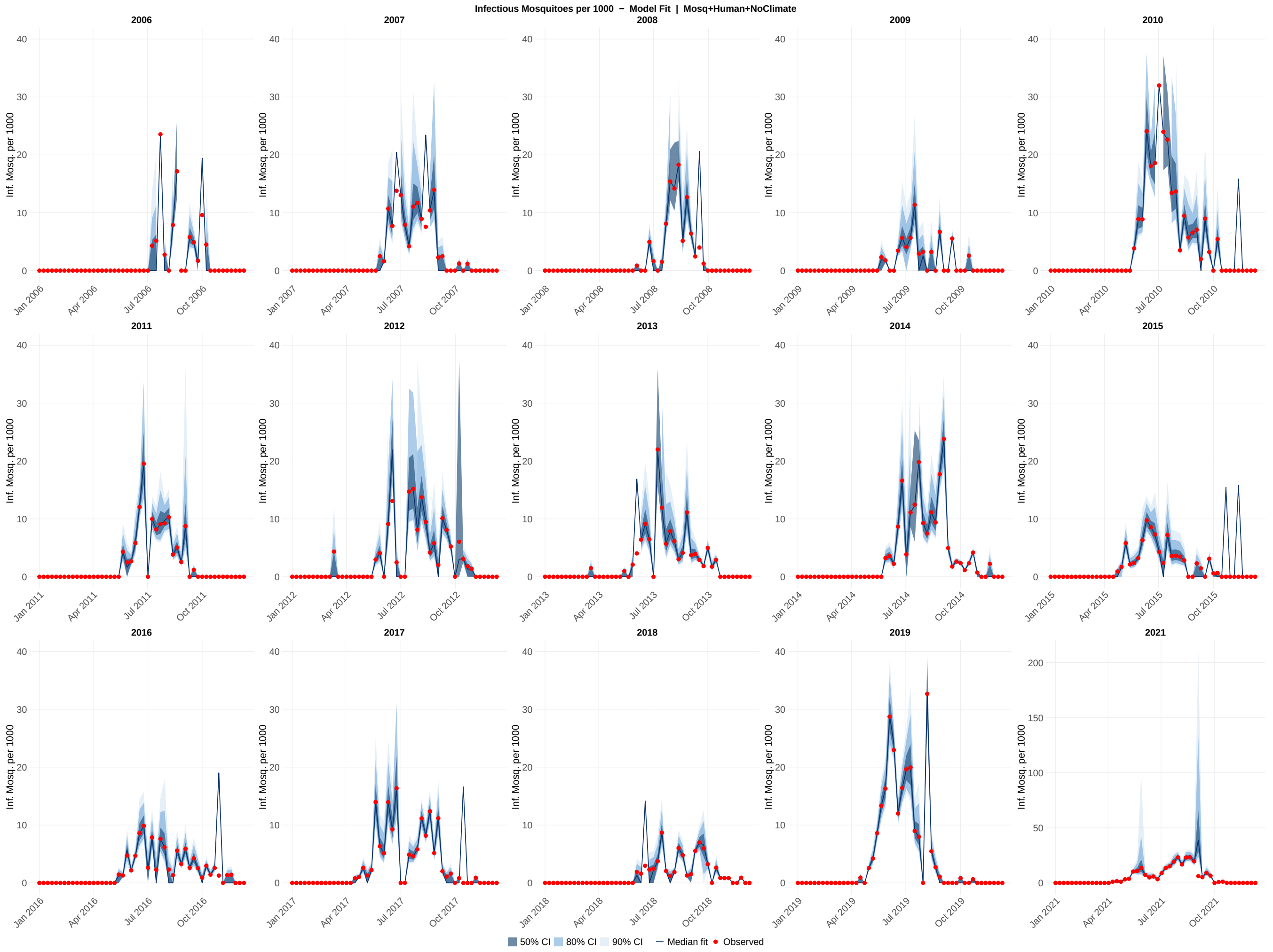

**Figure S8:** EnKF posterior model fit for infectious mosquito per 1000 across all 15 study years (2006–2019, 2021) for the mosquito + human without weather model configuration. Each panel shows the EnKF posterior distribution for a single calendar year. Shaded ribbons indicate the 50% (darkest), 80% (medium), and 90% (lightest) credible intervals of the posterior ensemble; the solid line shows the posterior median; red points show the observed weekly infectious mosquito per 1000 values. The y-axis scale for 2021 is unconstrained to accommodate the atypically high values observed that year; all other years share a common axis scale (0–40 for infectious mosquito per 1000; adjust as appropriate for other targets). Fits were generated over each year using the Ensemble Kalman Filter with Ornstein-Uhlenbeck parameter propagation.

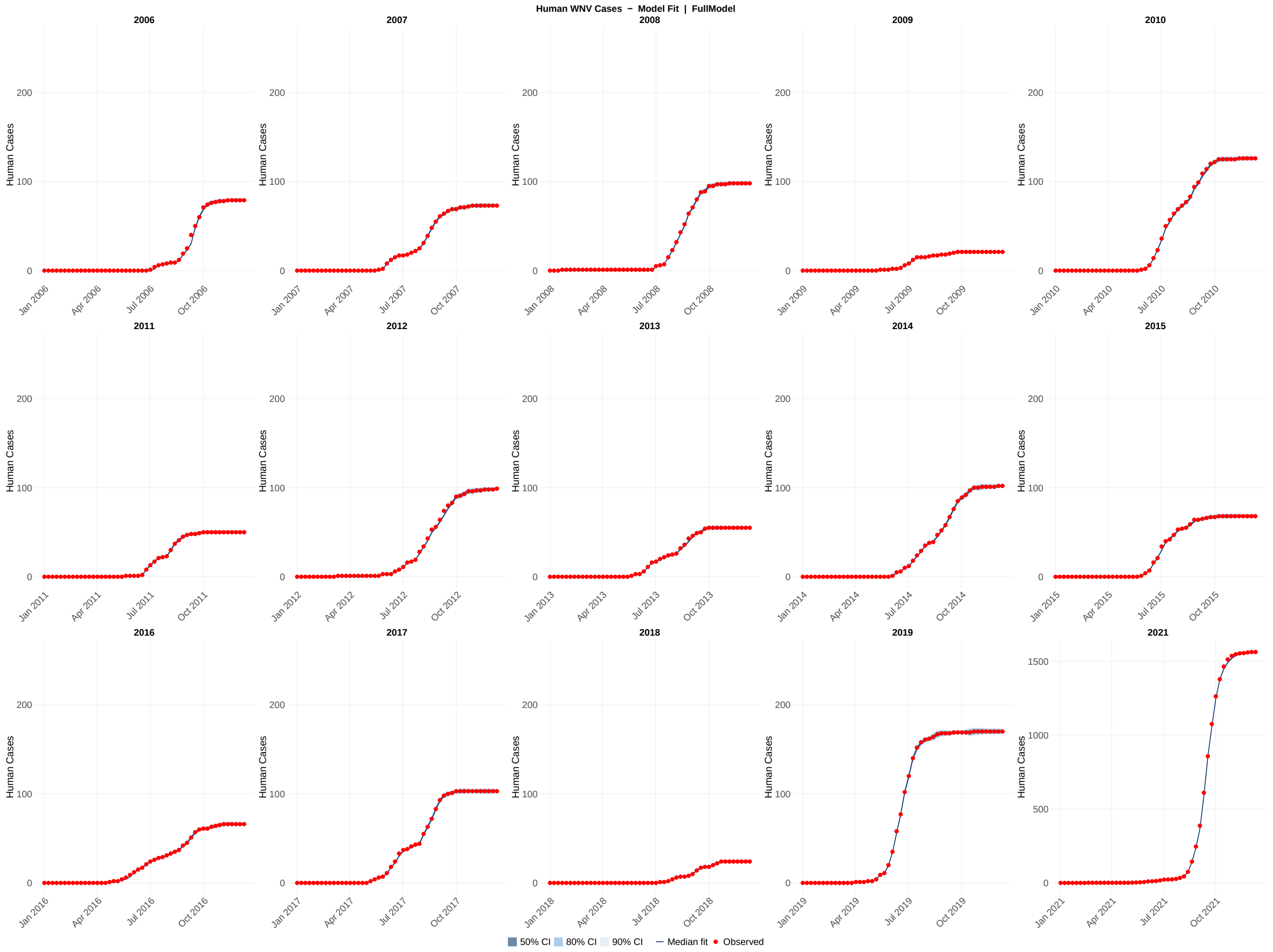

**Figure S9:** EnKF posterior model fit for human cases across all 15 study years (2006–2019, 2021) for the full model configuration. Each panel shows the EnKF posterior distribution for a single calendar year. Shaded ribbons indicate the 50% (darkest), 80% (medium), and 90% (lightest) credible intervals of the posterior ensemble; the solid line shows the posterior median; red points show the observed weekly human cases values. The y-axis scale for 2021 is unconstrained to accommodate the atypically high values observed that year; all other years share a common axis scale (0–200 for human cases; adjust as appropriate for other targets). Fits were generated over each year using the Ensemble Kalman Filter with Ornstein–Uhlenbeck parameter propagation.

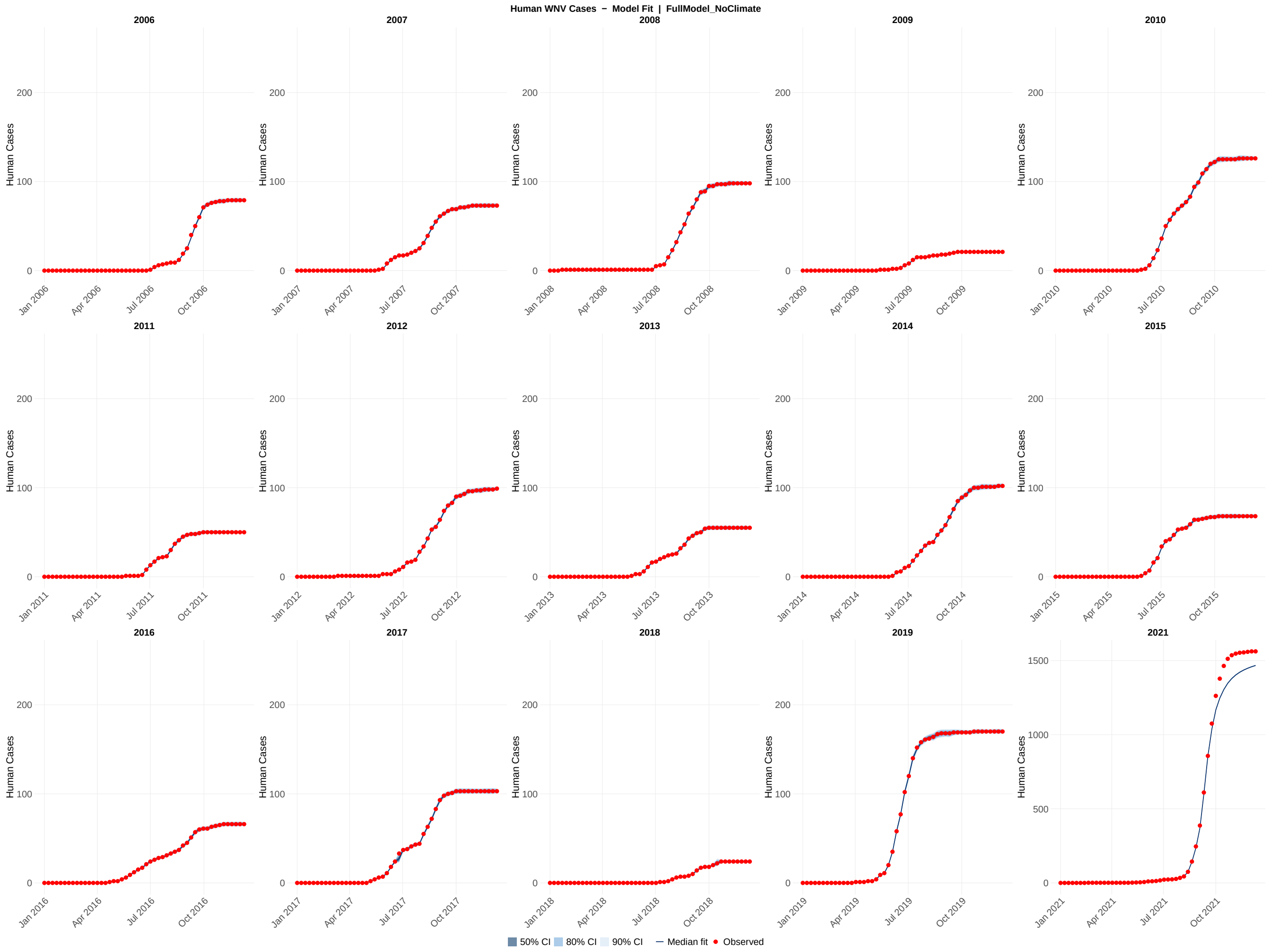

**Figure S10:** EnKF posterior model fit for human cases across all 15 study years (2006–2019, 2021) for the full model without weather configuration. Each panel shows the EnKF posterior distribution for a single calendar year. Shaded ribbons indicate the 50% (darkest), 80% (medium), and 90% (lightest) credible intervals of the posterior ensemble; the solid line shows the posterior median; red points show the observed weekly human cases values. The y-axis scale for 2021 is unconstrained to accommodate the atypically high values observed that year; all other years share a common axis scale (0–200 for human cases; adjust as appropriate for other targets). Fits were generated over each year using the Ensemble Kalman Filter with Ornstein–Uhlenbeck parameter propagation.

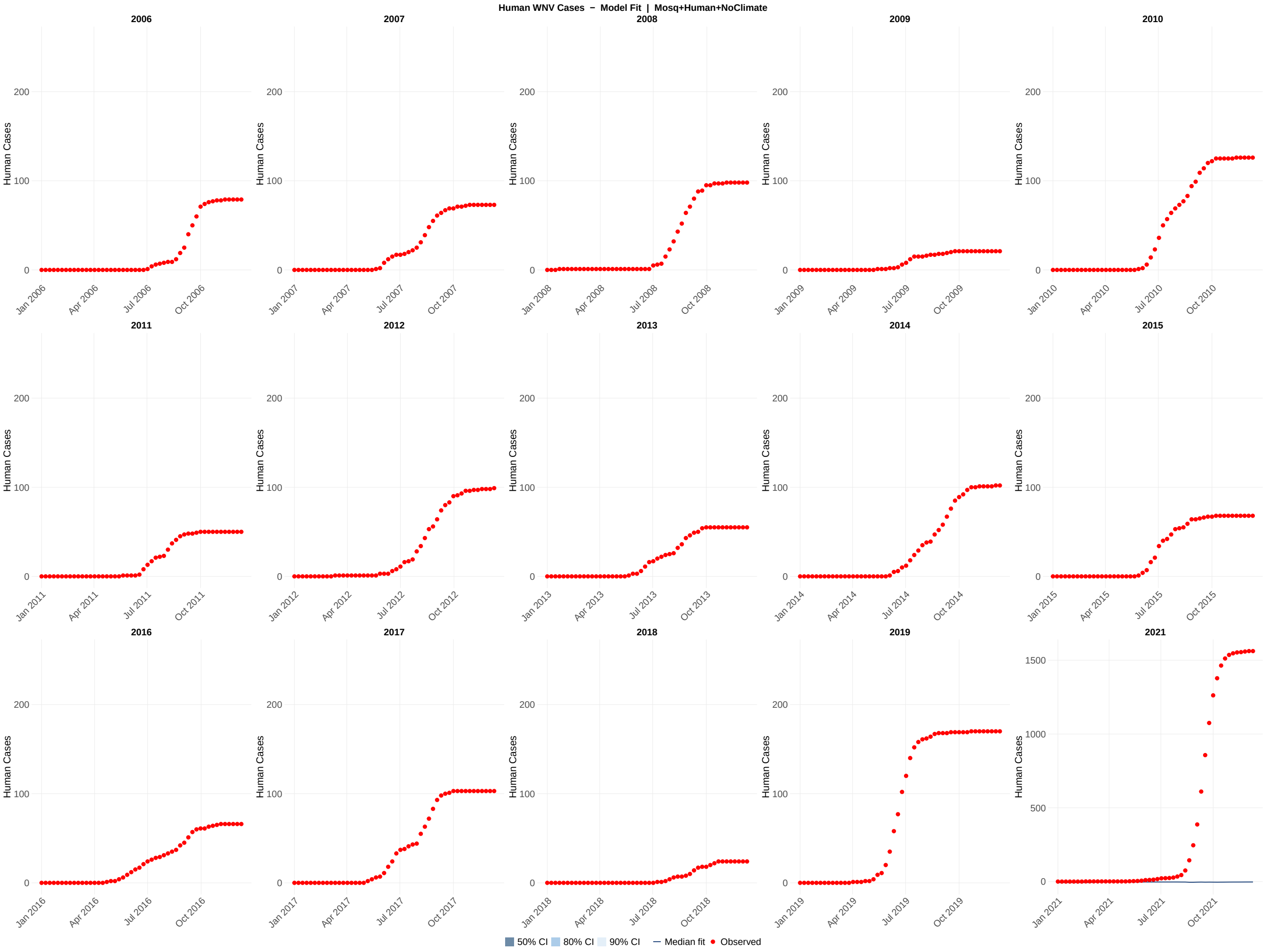

**Figure S11:** EnKF posterior model fit for human cases across all 15 study years (2006–2019, 2021) for the mosquito + human without weather model configuration. Each panel shows the EnKF posterior distribution for a single calendar year. Shaded ribbons indicate the 50% (darkest), 80% (medium), and 90% (lightest) credible intervals of the posterior ensemble; the solid line shows the posterior median; red points show the observed weekly human cases values. The y-axis scale for 2021 is unconstrained to accommodate the atypically high values observed that year; all other years share a common axis scale (0–200 for human cases; adjust as appropriate for other targets). Fits were generated over each year using the Ensemble Kalman Filter with Ornstein–Uhlenbeck parameter propagation.

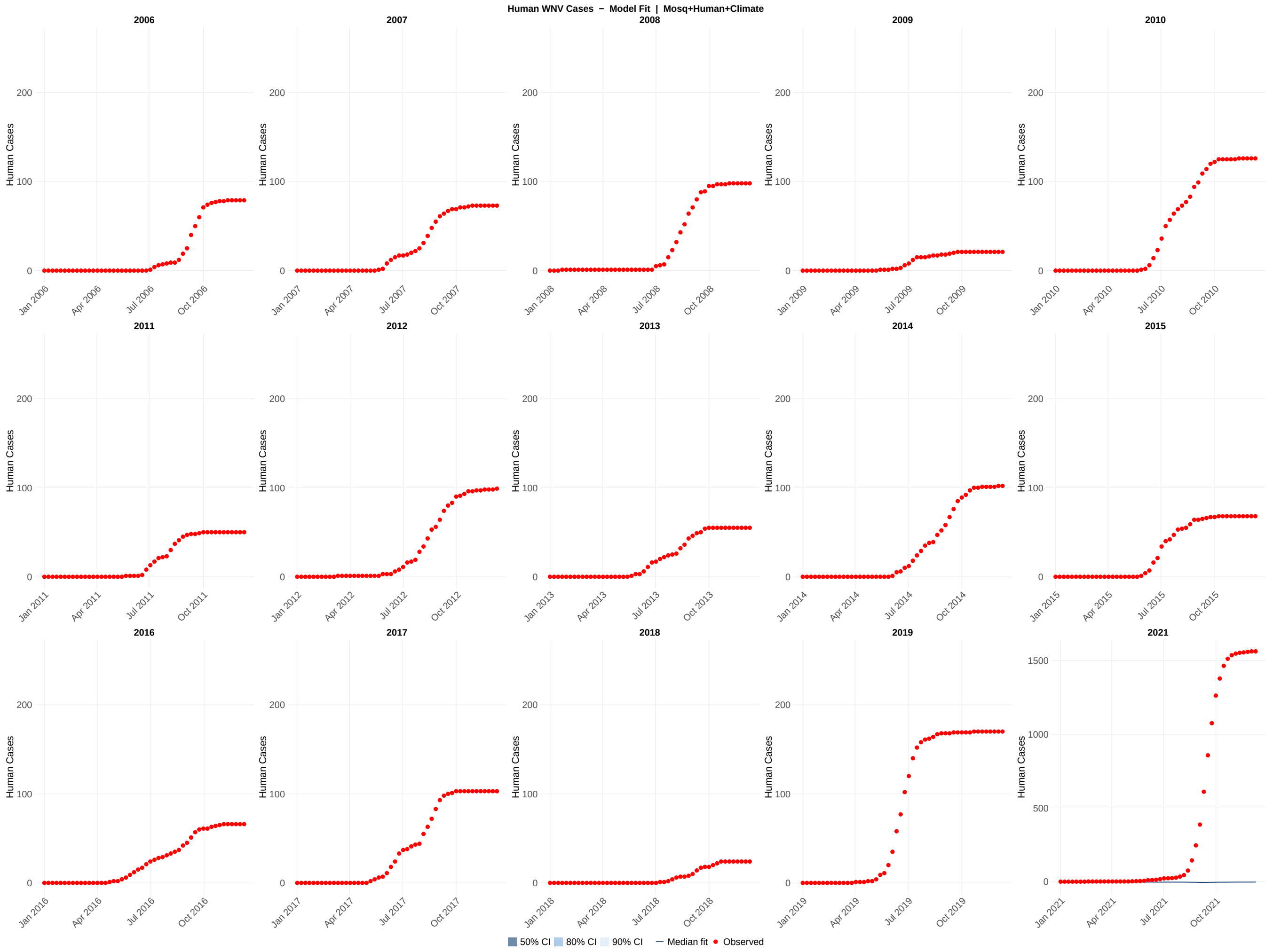

**Figure S12:** EnKF posterior model fit for human cases across all 15 study years (2006–2019, 2021) for the mosquito + human with weather model configuration. Each panel shows the EnKF posterior distribution for a single calendar year. Shaded ribbons indicate the 50% (darkest), 80% (medium), and 90% (lightest) credible intervals of the posterior ensemble; the solid line shows the posterior median; red points show the observed weekly human cases values. The y-axis scale for 2021 is unconstrained to accommodate the atypically high values observed that year; all other years share a common axis scale (0–200 for human cases; adjust as appropriate for other targets). Fits were generated over each year using the Ensemble Kalman Filter with Ornstein–Uhlenbeck parameter propagation.

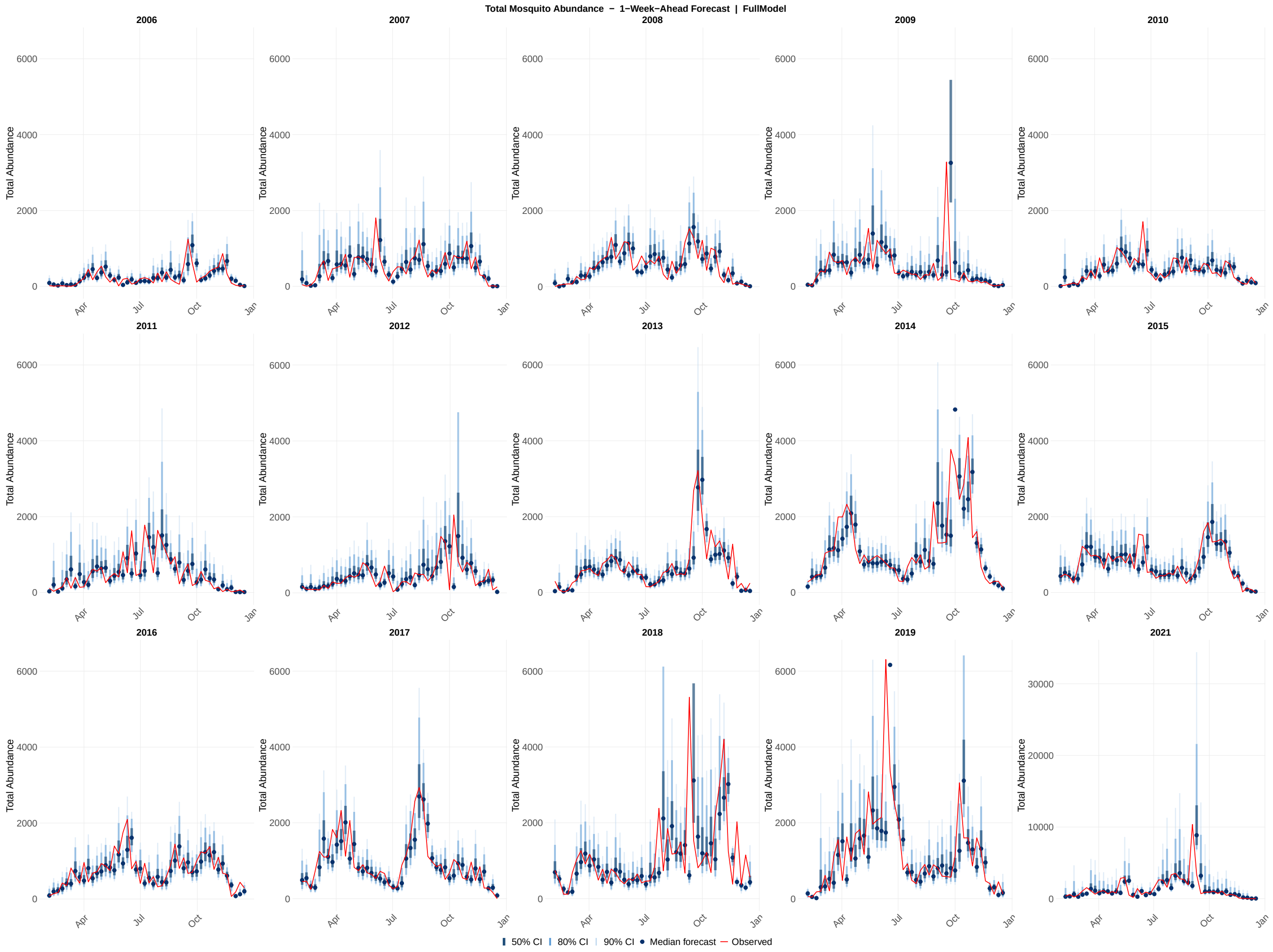

**Figure S13:** Probabilistic 1-week-ahead forecasts of total abundance across all 15 study years (2006–2019, 2021) for the full model configuration. Each panel shows the sequence of 1-week-ahead forecast distributions generated at each of the 46 assimilation steps within a single calendar year. Shaded ribbons indicate the 50% (darkest), 80% (medium), and 90% (lightest) prediction intervals; the solid line shows the forecast median; the red line shows the observed total abundance values. The y-axis scale for 2021 is unconstrained; all other years share a common axis scale. Forecasts were generated by drawing 1,000 samples from the EnKF posterior at each assimilation step, propagating the time-varying parameters one week forward via the Ornstein-Uhlenbeck process, and integrating the ODE model through the forecast horizon.

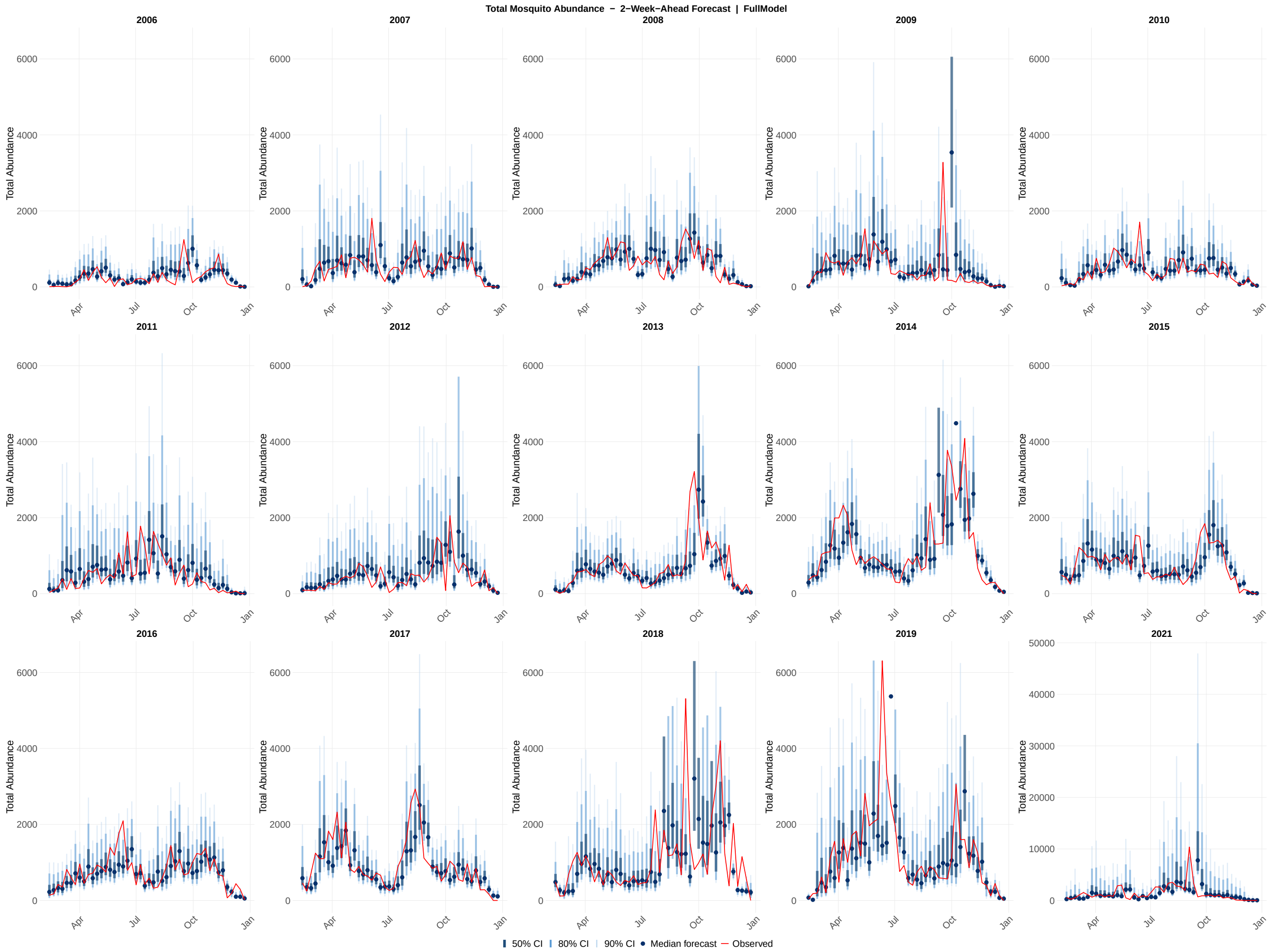

**Figure S14:** Probabilistic 2-week-ahead forecasts of total abundance across all 15 study years (2006–2019, 2021) for the full model configuration. Each panel shows the sequence of 2-week-ahead forecast distributions generated at each of the 46 assimilation steps within a single calendar year. Shaded ribbons indicate the 50% (darkest), 80% (medium), and 90% (lightest) prediction intervals; the solid line shows the forecast median; the red line shows the observed total abundance values. The y-axis scale for 2021 is unconstrained; all other years share a common axis scale. Forecasts were generated by drawing 1,000 samples from the EnKF posterior at each assimilation step and propagating the time-varying parameters two steps forward via the Ornstein-Uhlenbeck process, with each successive step conditioned on the preceding OU-propagated parameter value.

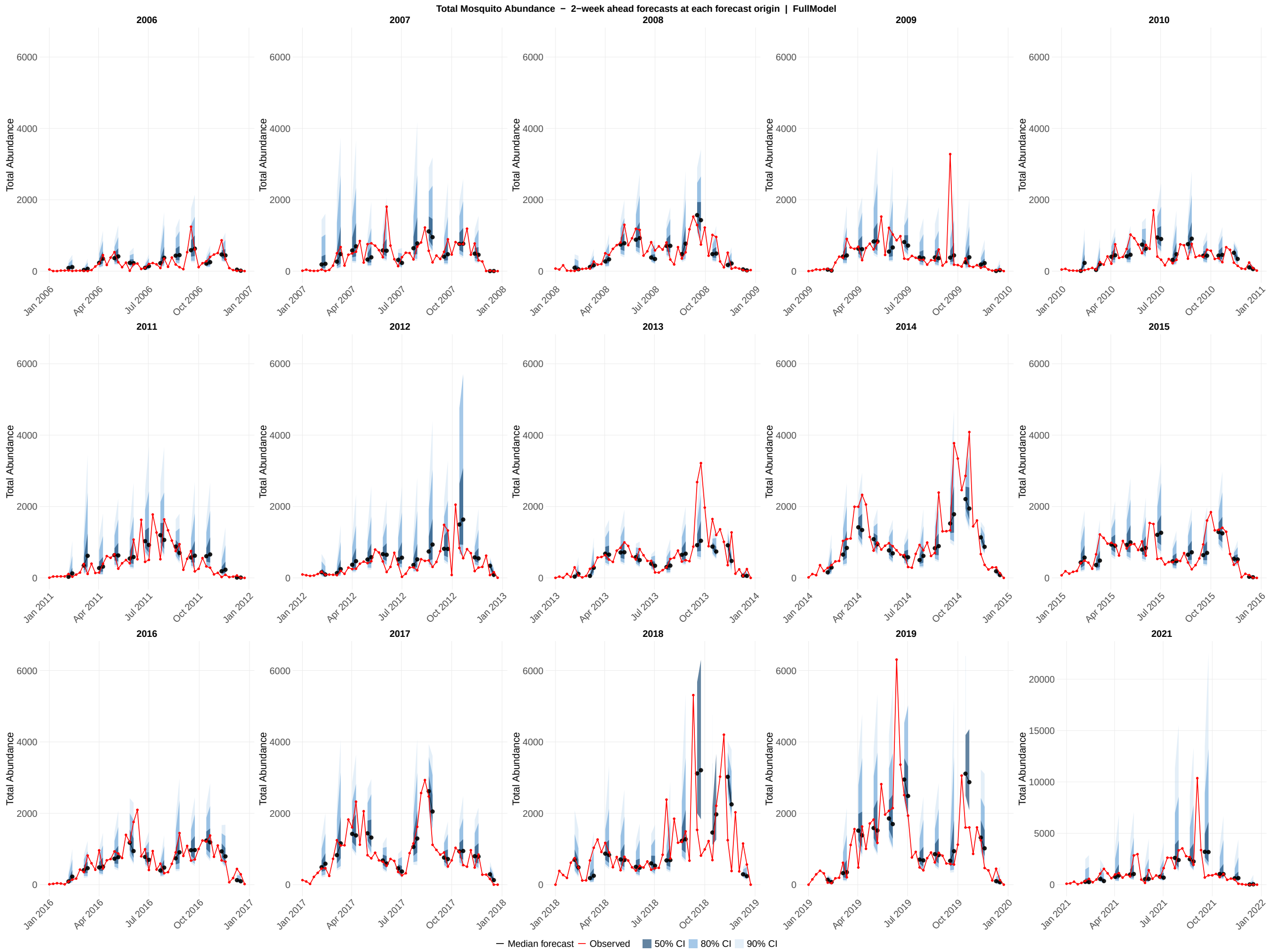

**Figure S15:** Probabilistic forecasts of total abundance across all 15 study years (2006–2019, 2021) for the full model configuration. Each panel displays an overlapping 2-week-ahead forecast generated at every fourth assimilation step within a single calendar year, providing a visual summary of how forecast uncertainty evolves across the transmission season. Each forecast horizon originates at a forecast origin date and extends through the 1-week-ahead and 2-week-ahead target dates; shaded ribbons show the 50% (darkest), 80% (medium), and 90% (lightest) prediction intervals at those two horizons; the colored line connecting the two horizon points shows the median forecast trajectory; the red line shows the observed total abundance values. The y-axis scale for 2021 is unconstrained; all other years share a common axis scale.

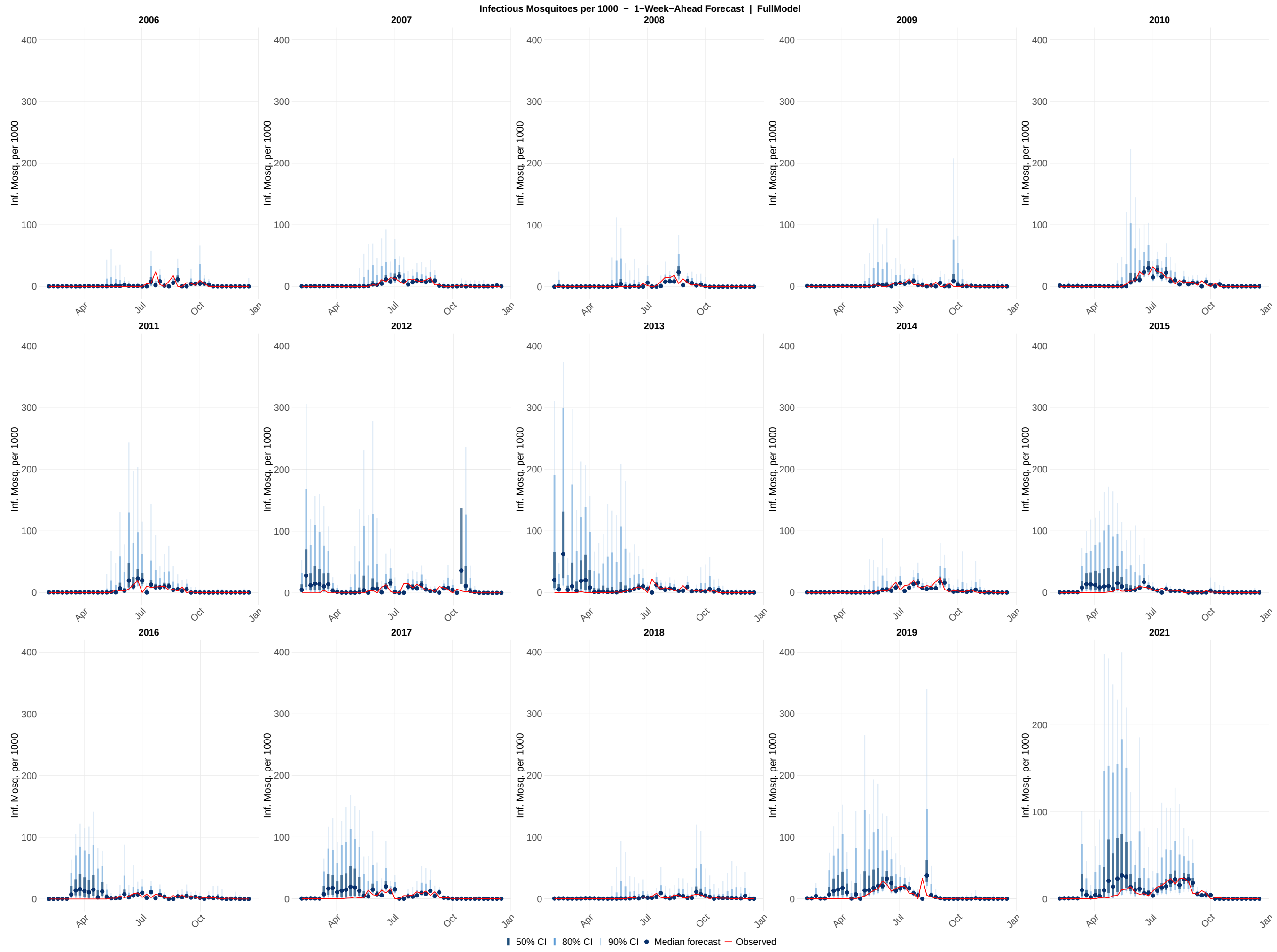

**Figure S16:** Probabilistic 1-week-ahead forecasts of infectious mosquitoes per 1000 across all 15 study years (2006–2019, 2021) for the full model configuration. Each panel shows the sequence of 1-week-ahead forecast distributions generated at each of the 46 assimilation steps within a single calendar year. Shaded ribbons indicate the 50% (darkest), 80% (medium), and 90% (lightest) prediction intervals; the solid line shows the forecast median; the red line shows the observed infectious mosquito per 1000 values. The y-axis scale for 2021 is unconstrained; all other years share a common axis scale. Forecasts were generated by drawing 1,000 samples from the EnKF posterior at each assimilation step, propagating the time-varying parameters one week forward via the Ornstein-Uhlenbeck process, and integrating the ODE model through the forecast horizon.

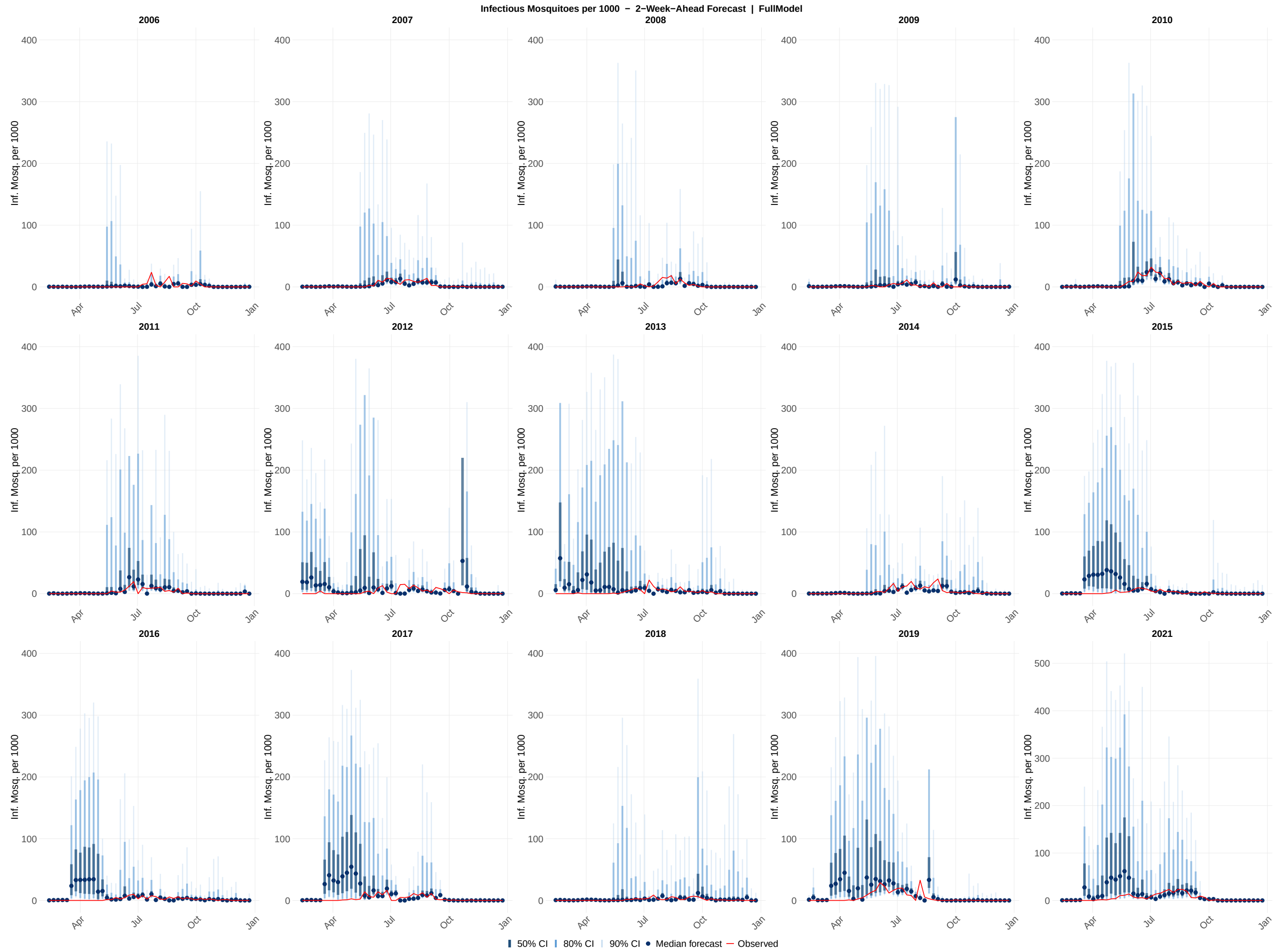

**Figure S17:** Probabilistic 2-week-ahead forecasts of infectious mosquito per 1000 across all 15 study years (2006–2019, 2021) for the full model configuration. Each panel shows the sequence of 2-week-ahead forecast distributions generated at each of the 46 assimilation steps within a single calendar year. Shaded ribbons indicate the 50% (darkest), 80% (medium), and 90% (lightest) prediction intervals; the solid line shows the forecast median; and the red line shows the observed infectious mosquito per 1000 values. The y-axis scale for 2021 is unconstrained; all other years share a common axis scale. Forecasts were generated by drawing 1,000 samples from the EnKF posterior at each assimilation step and propagating the time-varying parameters two steps forward via the Ornstein-Uhlenbeck process, with each successive step conditioned on the preceding OU-propagated parameter value.

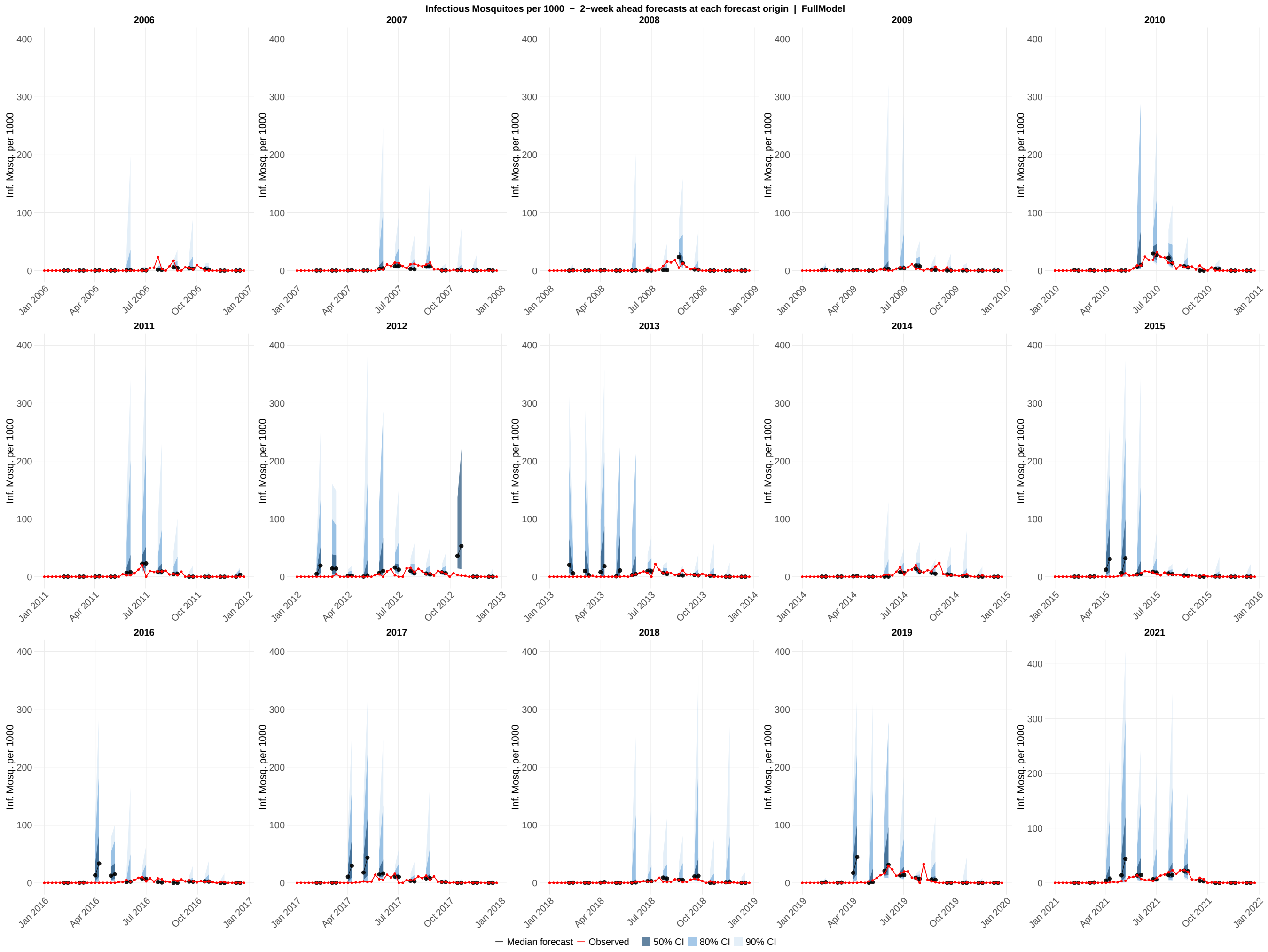

**Figure S18:** Probabilistic forecasts of infectious mosquitoes per 1000 all 15 study years (2006–2019, 2021) for the full model configuration. Each panel displays overlapping 2-week-ahead forecast generated at every fourth assimilation step within a single calendar year, providing a visual summary of how forecast uncertainty evolves across the transmission season. Each forecast horizon originates at a forecast origin date and extends through the 1-week-ahead and 2-week-ahead target dates; shaded ribbons show the 50% (darkest), 80% (medium), and 90% (lightest) prediction intervals at those two horizons; the colored line connecting the two horizon points shows the median forecast trajectory; the red line shows the observed infectious mosquito per 1000 values. The y-axis scale for 2021 is unconstrained; all other years share a common axis scale.

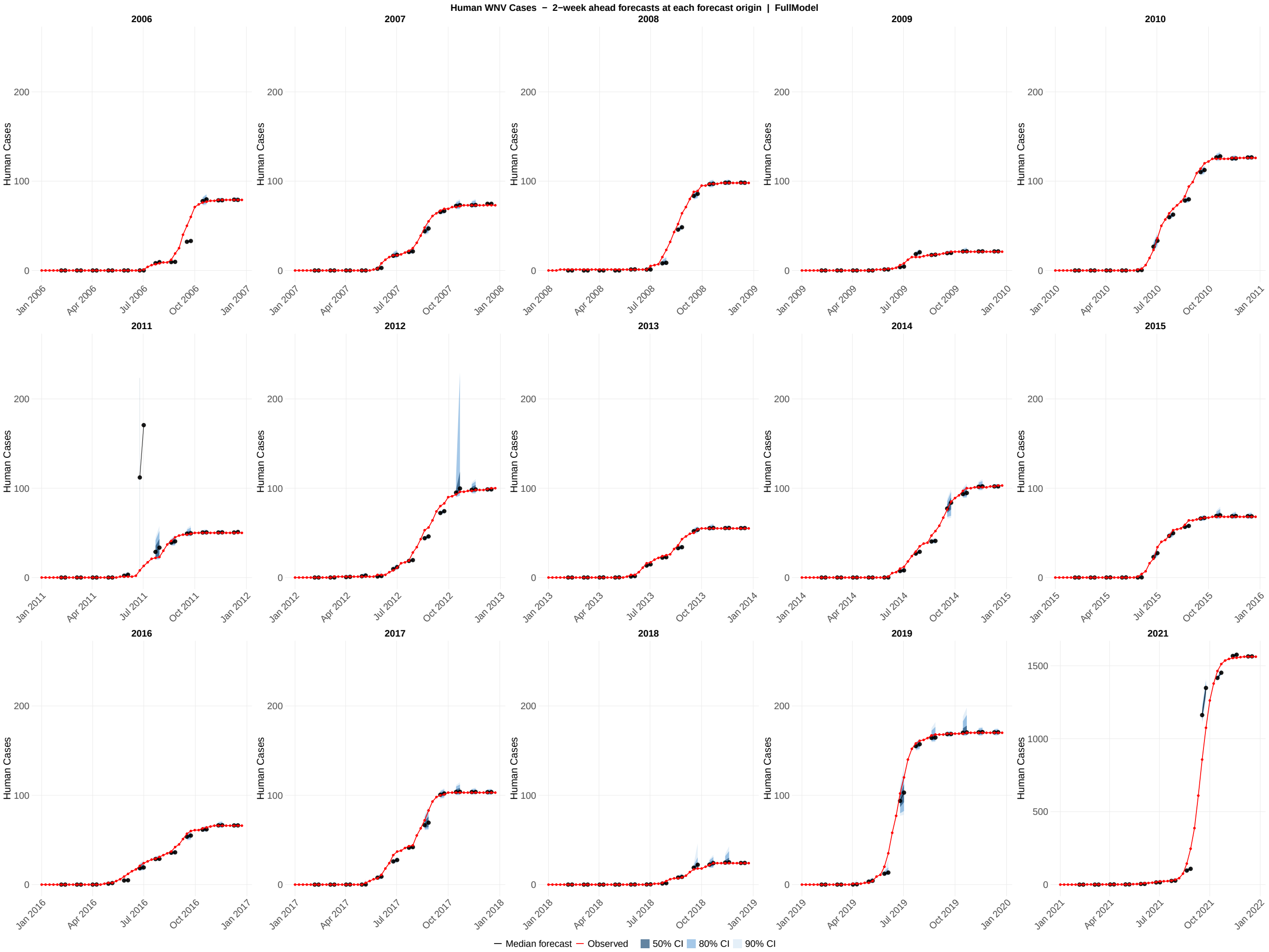

**Figure S19:** Probabilistic forecasts of human cases across all 15 study years (2006–2019, 2021) for the full model configuration. Each panel displays overlapping 2-week-ahead forecast generated at every fourth assimilation step within a single calendar year, providing a visual summary of how forecast uncertainty evolves across the transmission season. Each forecast horizon originates at a forecast origin date and extends through the 1-week-ahead and 2-week-ahead target dates; shaded ribbons show the 50% (darkest), 80% (medium), and 90% (lightest) prediction intervals at those two horizons; the colored line connecting the two horizon points shows the median forecast trajectory; the red line shows the observed human cases values. The y-axis scale for 2021 is unconstrained; all other years share a common axis scale.

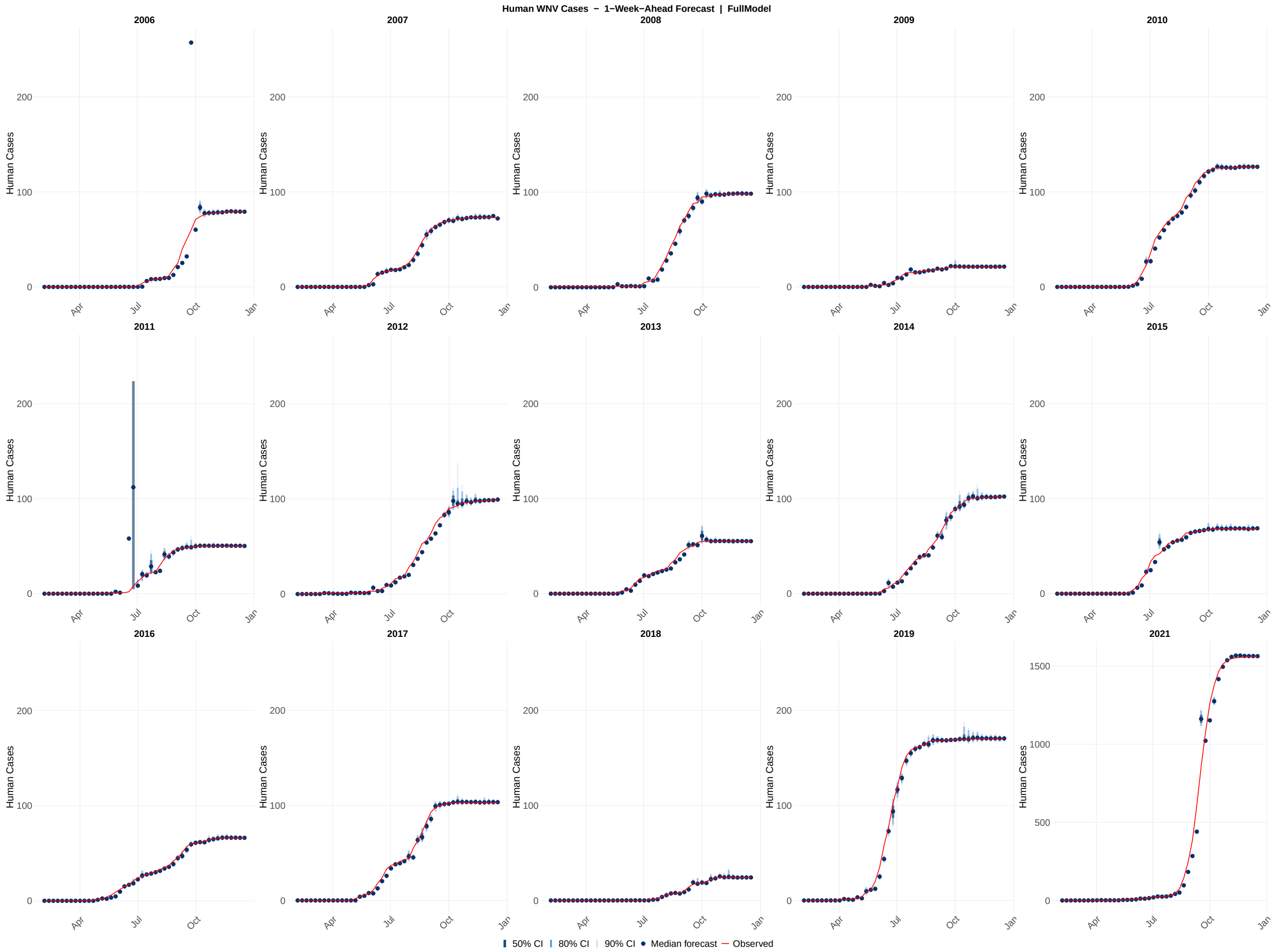

**Figure S20:** Probabilistic 1-week-ahead forecasts of human cases across all 15 study years (2006–2019, 2021) for the full model configuration. Each panel shows the sequence of 1-week-ahead forecast distributions generated at each of the 46 assimilation steps within a single calendar year. Shaded ribbons indicate the 50% (darkest), 80% (medium), and 90% (lightest) prediction intervals; the solid line shows the forecast median; the red line shows the observed human cases values. The y-axis scale for 2021 is unconstrained; all other years share a common axis scale. Forecasts were generated by drawing 1,000 samples from the EnKF posterior at each assimilation step, propagating the time-varying parameters one week forward via the Ornstein–Uhlenbeck process, and integrating the ODE model through the forecast horizon.

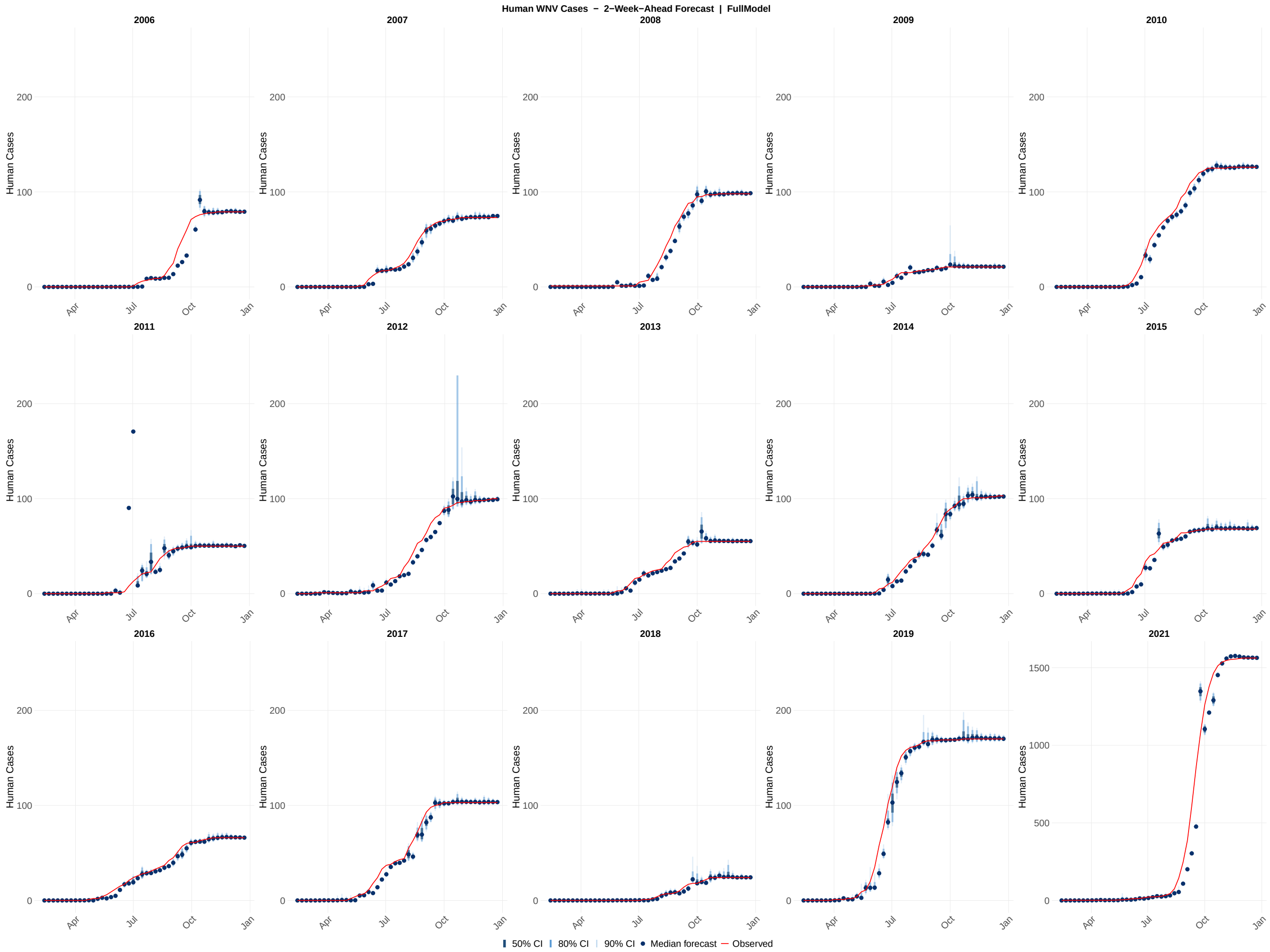

**Figure S21:** Probabilistic 2-week-ahead forecasts of human cases across all 15 study years (2006–2019, 2021) for the full model configuration. Each panel shows the sequence of 2-week-ahead forecast distributions generated at each of the 46 assimilation steps within a single calendar year. Shaded ribbons indicate the 50% (darkest), 80% (medium), and 90% (lightest) prediction intervals; the solid line shows the forecast median; the red line shows the observed human cases values. The y-axis scale for 2021 is unconstrained; all other years share a common axis scale. Forecasts were generated by drawing 1,000 samples from the EnKF posterior at each assimilation step and propagating the time-varying parameters two steps forward via the Ornstein–Uhlenbeck process, with each successive step conditioned on the preceding OU-propagated parameter value.

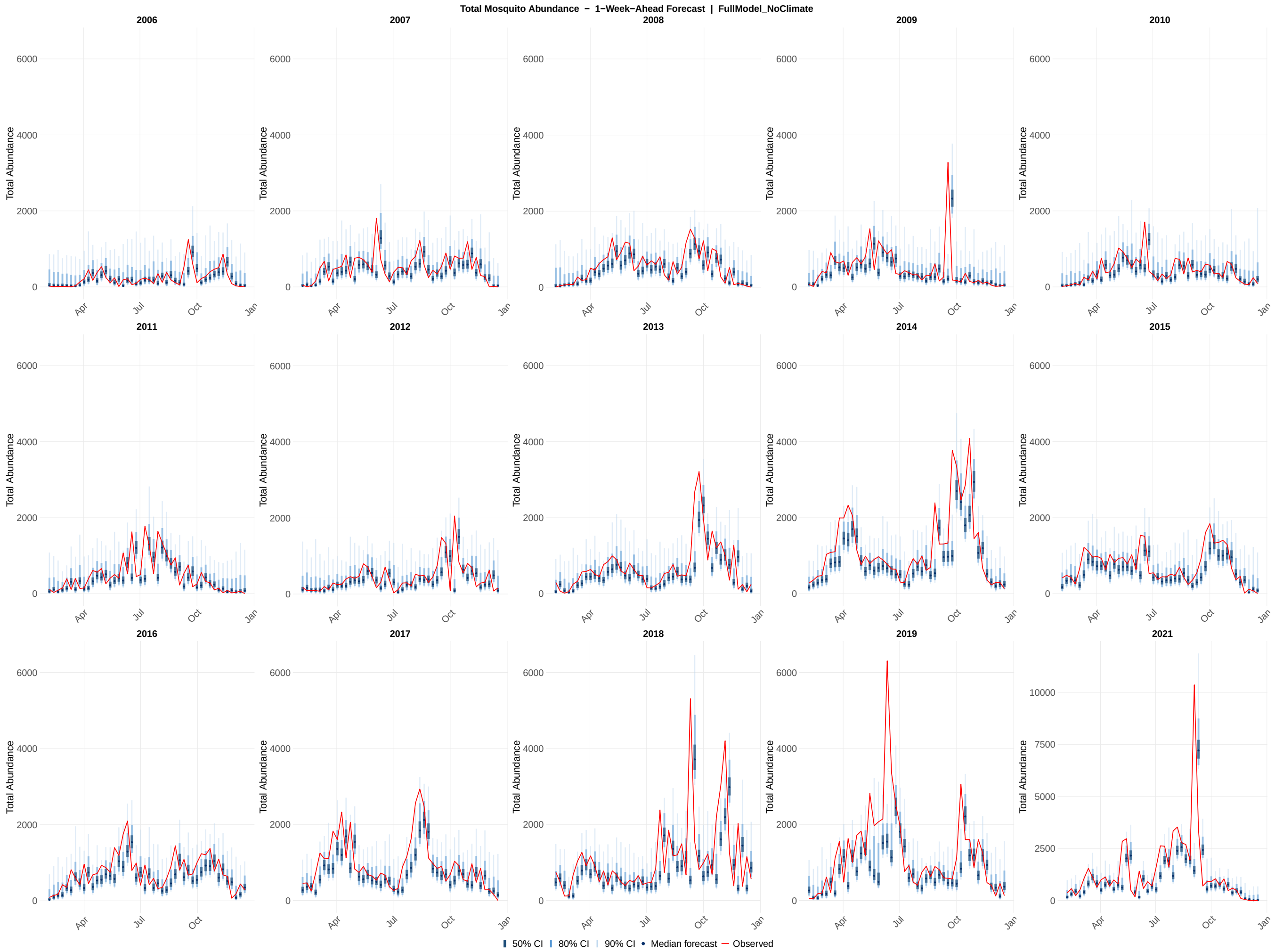

**Figure S22:** Probabilistic 1-week-ahead forecasts of total abundance across all 15 study years (2006–2019, 2021) for the full model without weather configuration. Each panel shows the sequence of 1-week-ahead forecast distributions generated at each of the 46 assimilation steps within a single calendar year. Shaded ribbons indicate the 50% (darkest), 80% (medium), and 90% (lightest) prediction intervals; the solid line shows the forecast median; the red line shows the observed total abundance values. The y-axis scale for 2021 is unconstrained; all other years share a common axis scale. Forecasts were generated by drawing 1,000 samples from the EnKF posterior at each assimilation step, propagating the time-varying parameters one week forward via the Ornstein-Uhlenbeck process, and integrating the ODE model through the forecast horizon.

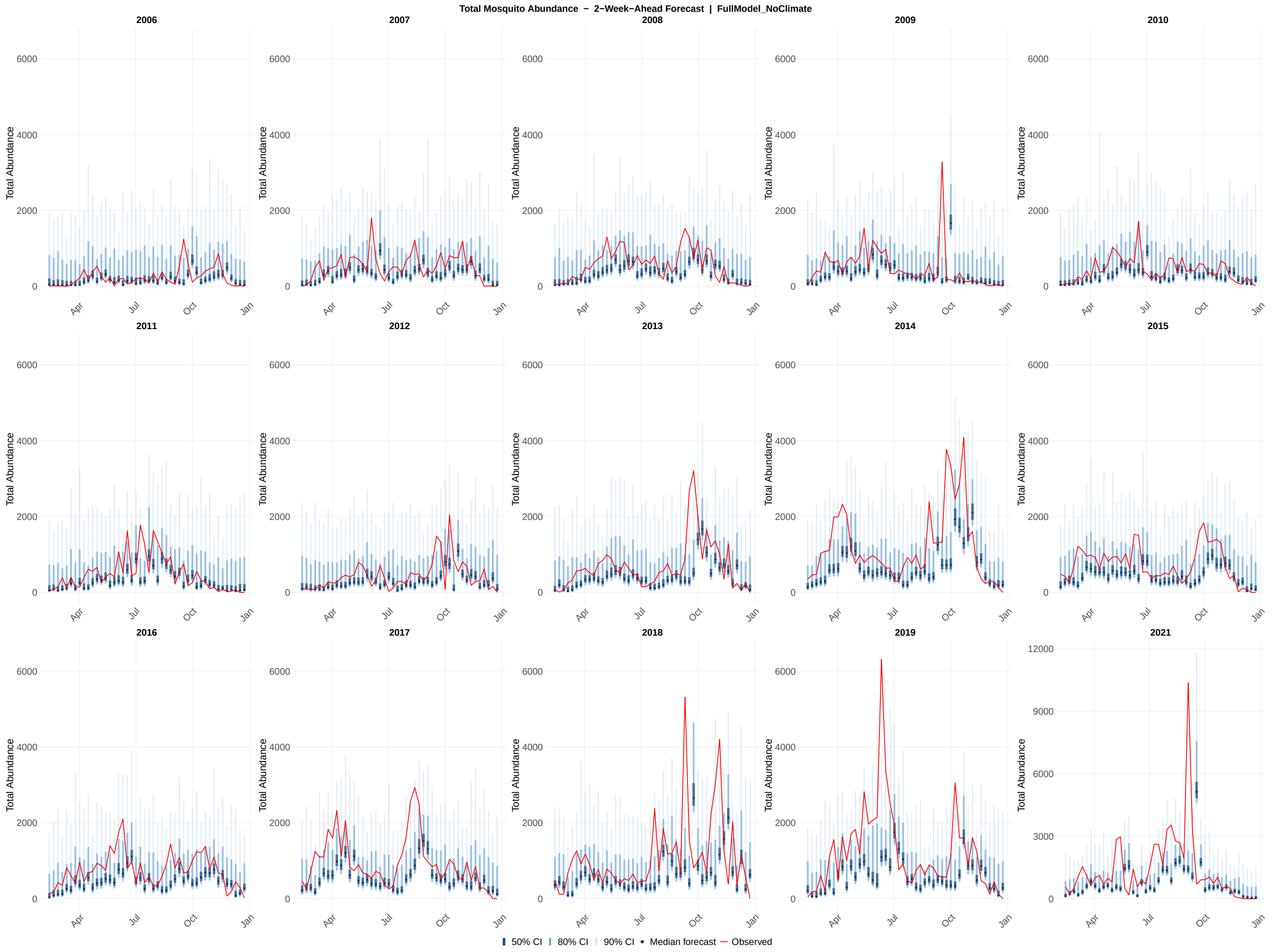

**Figure S23:** Probabilistic 2-week-ahead forecasts of total abundance across all 15 study years (2006–2019, 2021) for the full model without weather configuration. Each panel shows the sequence of 2-week-ahead forecast distributions generated at each of the 46 assimilation steps within a single calendar year. Shaded ribbons indicate the 50% (darkest), 80% (medium), and 90% (lightest) prediction intervals; the solid line shows the forecast median; the red line shows the observed total abundance values. The y-axis scale for 2021 is unconstrained; all other years share a common axis scale. Forecasts were generated by drawing 1,000 samples from the EnKF posterior at each assimilation step and propagating the time-varying parameters two steps forward via the Ornstein-Uhlenbeck process, with each successive step conditioned on the preceding OU-propagated parameter value.

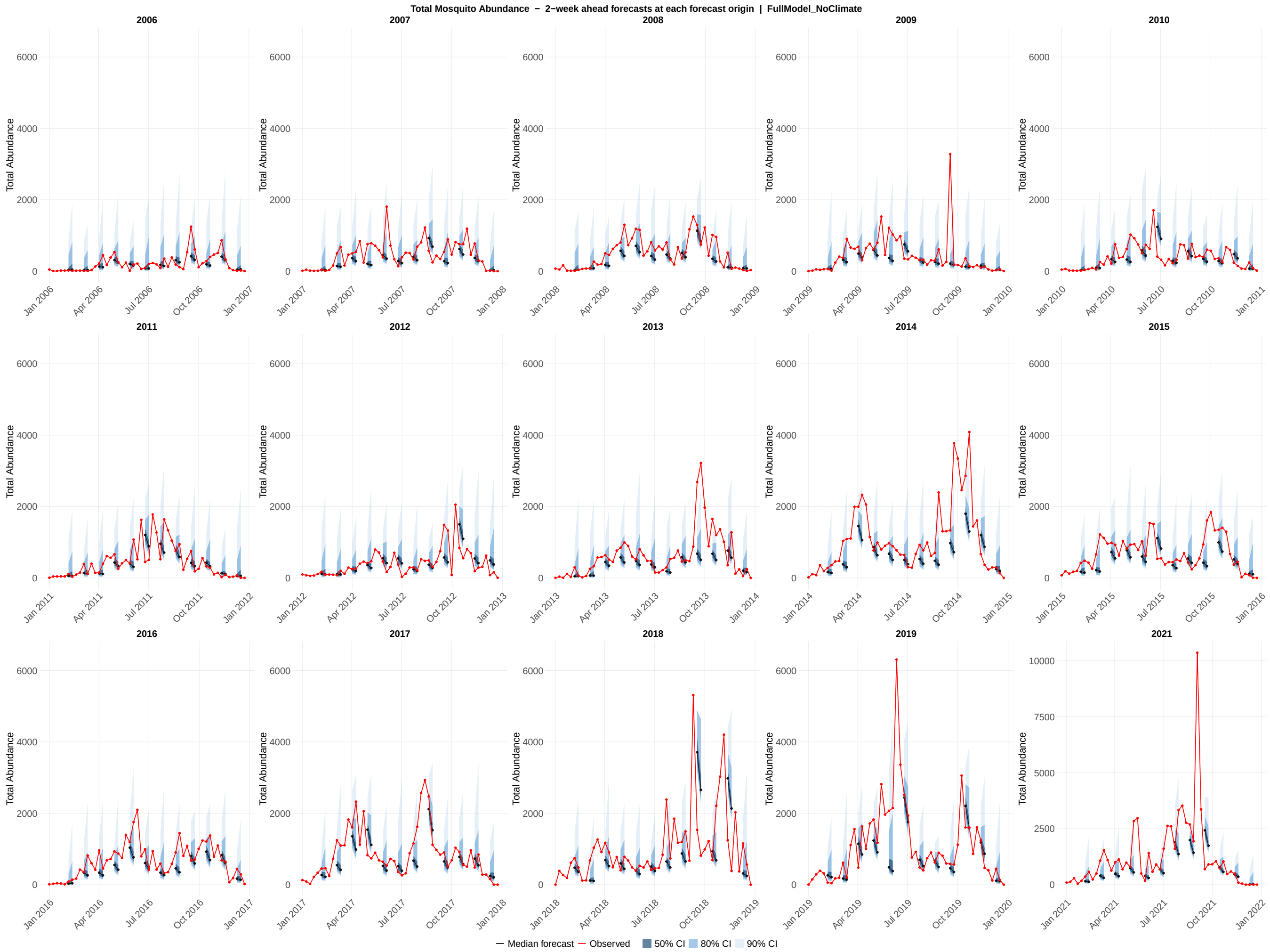

**Figure S24:** Probabilistic forecasts of total abundance across all 15 study years (2006–2019, 2021) for the full model without weather configuration. Each panel displays an overlapping 2-week-ahead forecast generated at every fourth assimilation step within a single calendar year, providing a visual summary of how forecast uncertainty evolves across the transmission season. Each forecast horizon originates at a forecast origin date and extends through the 1-week-ahead and 2-week-ahead target dates; shaded ribbons show the 50% (darkest), 80% (medium), and 90% (lightest) prediction intervals at those two horizons; the colored line connecting the two horizon points shows the median forecast trajectory; the red line shows the observed total abundance values. The y-axis scale for 2021 is unconstrained; all other years share a common axis scale.

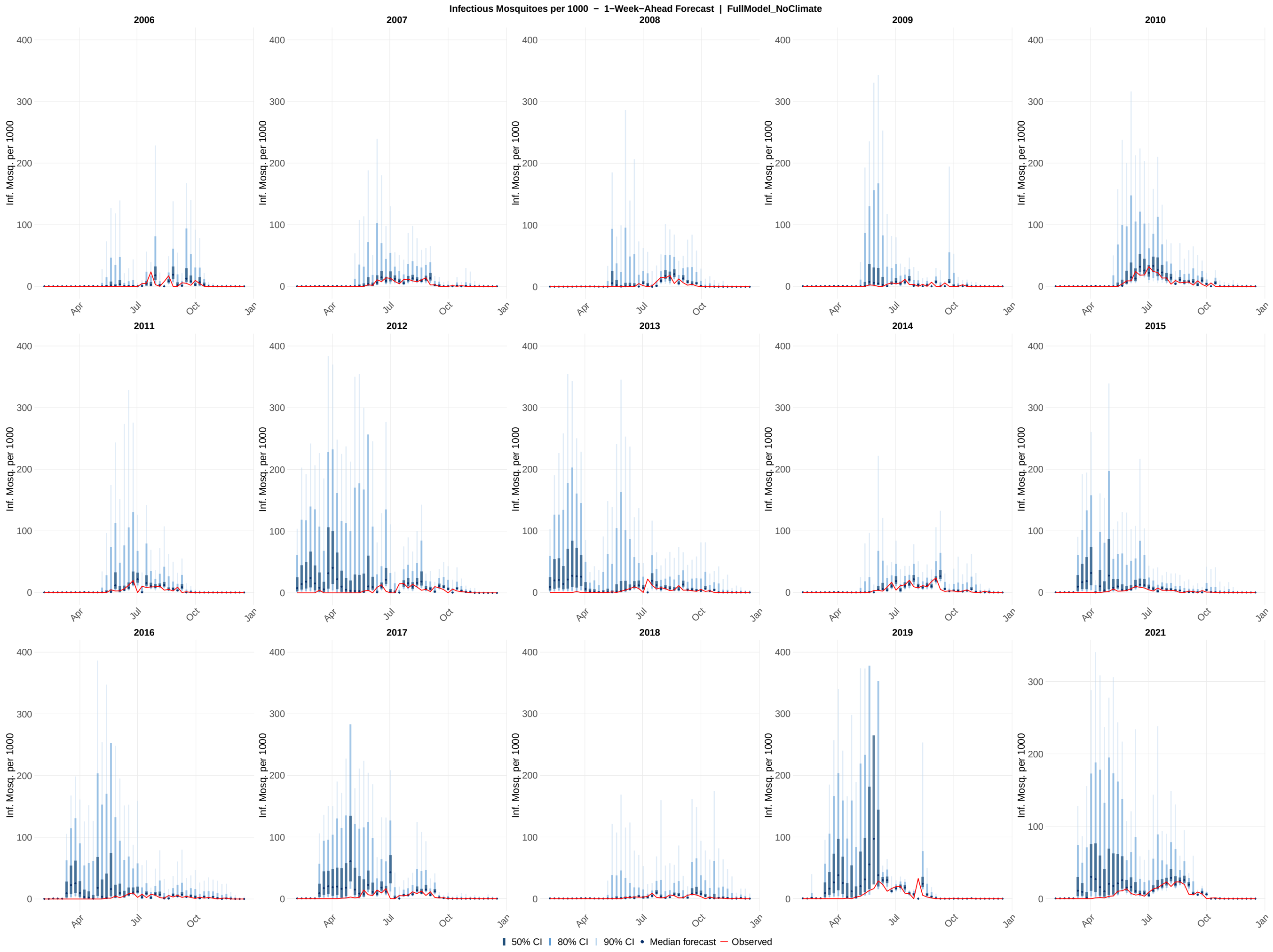

**Figure S25:** Probabilistic 1-week-ahead forecasts of infectious mosquitoes per 1000 across all 15 study years (2006–2019, 2021) for the full model without weather configuration. Each panel shows the sequence of 1-week-ahead forecast distributions generated at each of the 46 assimilation steps within a single calendar year. Shaded ribbons indicate the 50% (darkest), 80% (medium), and 90% (lightest) prediction intervals; the solid line shows the forecast median; the red line shows the observed infectious mosquito per 1000 values. The y-axis scale for 2021 is unconstrained; all other years share a common axis scale. Forecasts were generated by drawing 1,000 samples from the EnKF posterior at each assimilation step, propagating the time-varying parameters one week forward via the Ornstein-Uhlenbeck process, and integrating the ODE model through the forecast horizon.

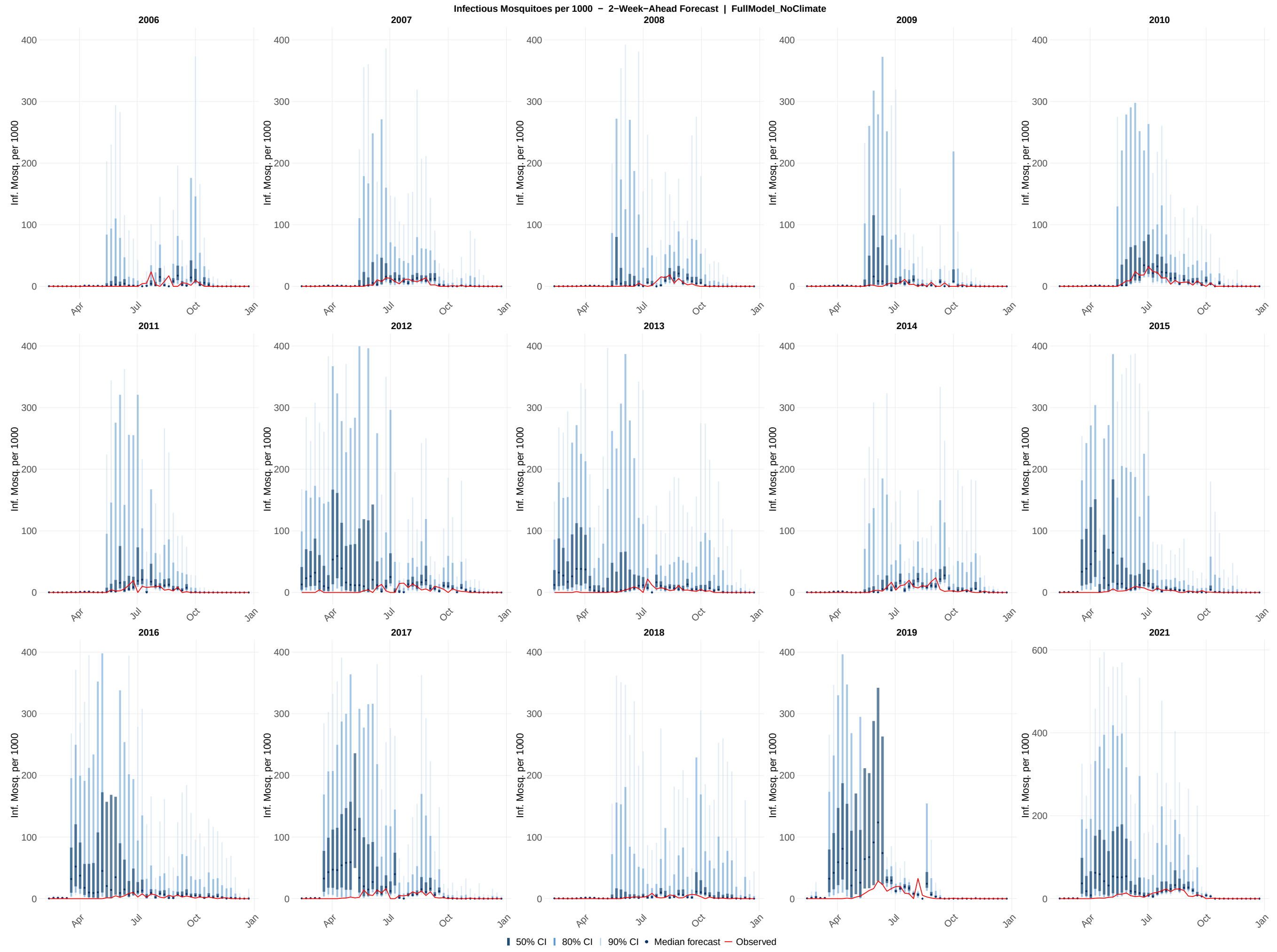

**Figure S26:** Probabilistic 2-week-ahead forecasts of infectious mosquito per 1000 across all 15 study years (2006–2019, 2021) for the full model without weather configuration. Each panel shows the sequence of 2-week-ahead forecast distributions generated at each of the 46 assimilation steps within a single calendar year. Shaded ribbons indicate the 50% (darkest), 80% (medium), and 90% (lightest) prediction intervals; the solid line shows the forecast median; and the red line shows the observed infectious mosquito per 1000 values. The y-axis scale for 2021 is unconstrained; all other years share a common axis scale. Forecasts were generated by drawing 1,000 samples from the EnKF posterior at each assimilation step and propagating the time-varying parameters two steps forward via the Ornstein-Uhlenbeck process, with each successive step conditioned on the preceding OU-propagated parameter value.

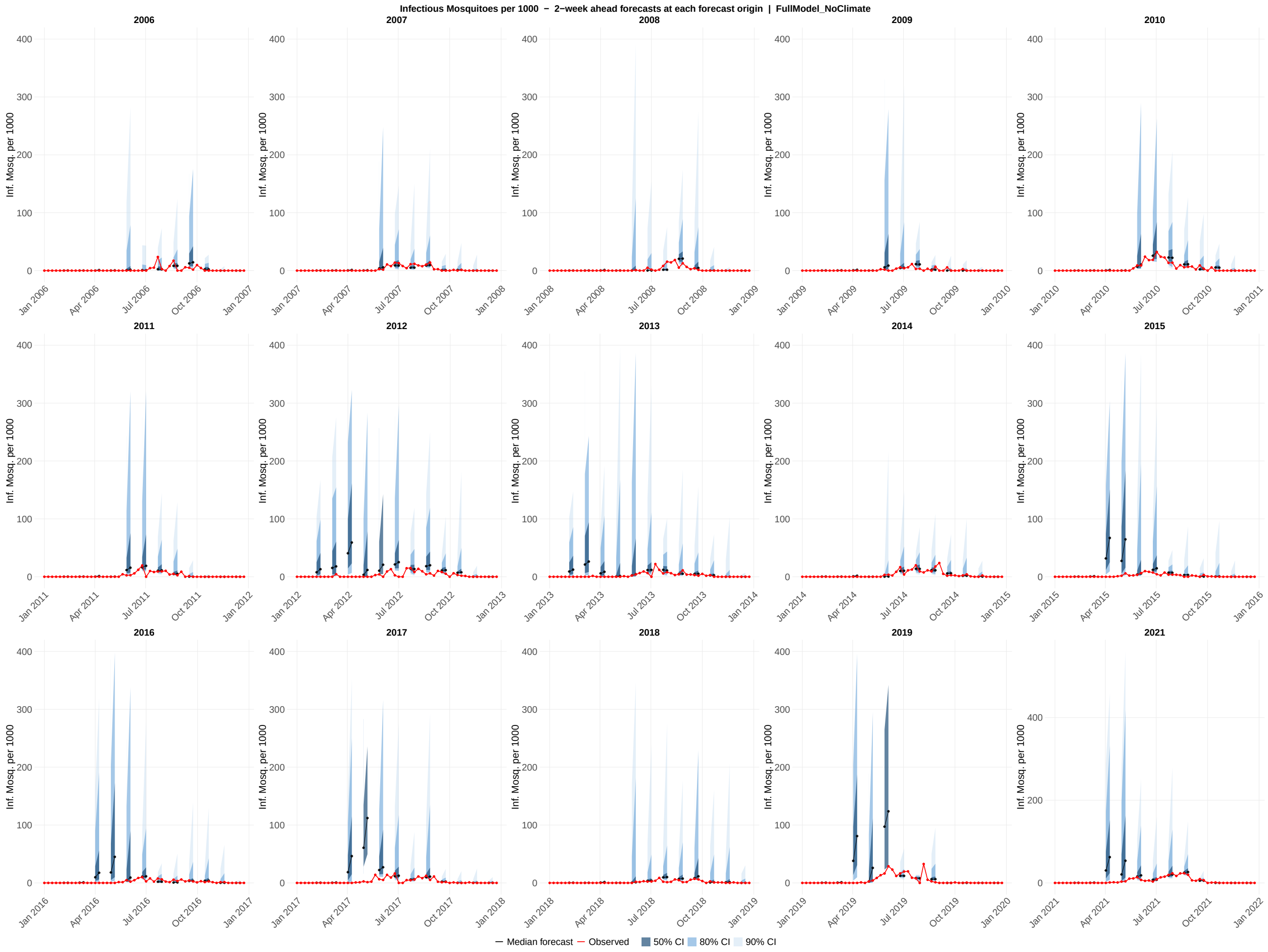

**Figure S27:** Probabilistic forecasts of infectious mosquitoes per 1000 all 15 study years (2006–2019, 2021) for the full model without weather configuration. Each panel displays overlapping 2-week-ahead forecast generated at every fourth assimilation step within a single calendar year, providing a visual summary of how forecast uncertainty evolves across the transmission season. Each forecast horizon originates at a forecast origin date and extends through the 1-week-ahead and 2-week-ahead target dates; shaded ribbons show the 50% (darkest), 80% (medium), and 90% (lightest) prediction intervals at those two horizons; the colored line connecting the two horizon points shows the median forecast trajectory; the red line shows the observed infectious mosquito per 1000 values. The y-axis scale for 2021 is unconstrained; all other years share a common axis scale.

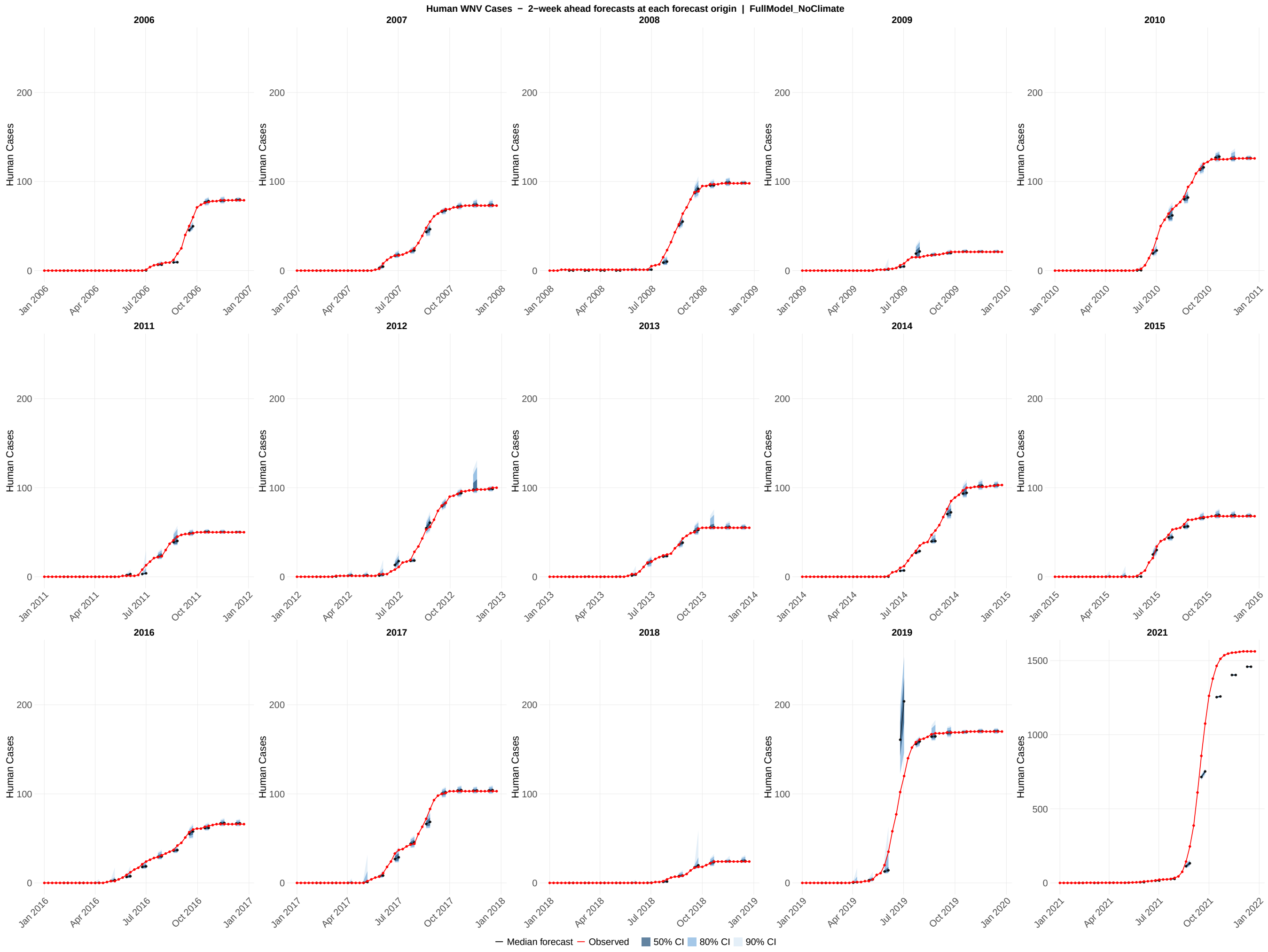

**Figure S28:** Probabilistic forecasts of human cases across all 15 study years (2006–2019, 2021) for the full model without weather configuration. Each panel displays overlapping 2-week-ahead forecast generated at every fourth assimilation step within a single calendar year, providing a visual summary of how forecast uncertainty evolves across the transmission season. Each forecast horizon originates at a forecast origin date and extends through the 1-week-ahead and 2-week-ahead target dates; shaded ribbons show the 50% (darkest), 80% (medium), and 90% (lightest) prediction intervals at those two horizons; the colored line connecting the two horizon points shows the median forecast trajectory; the red line shows the observed human cases values. The y-axis scale for 2021 is unconstrained; all other years share a common axis scale.

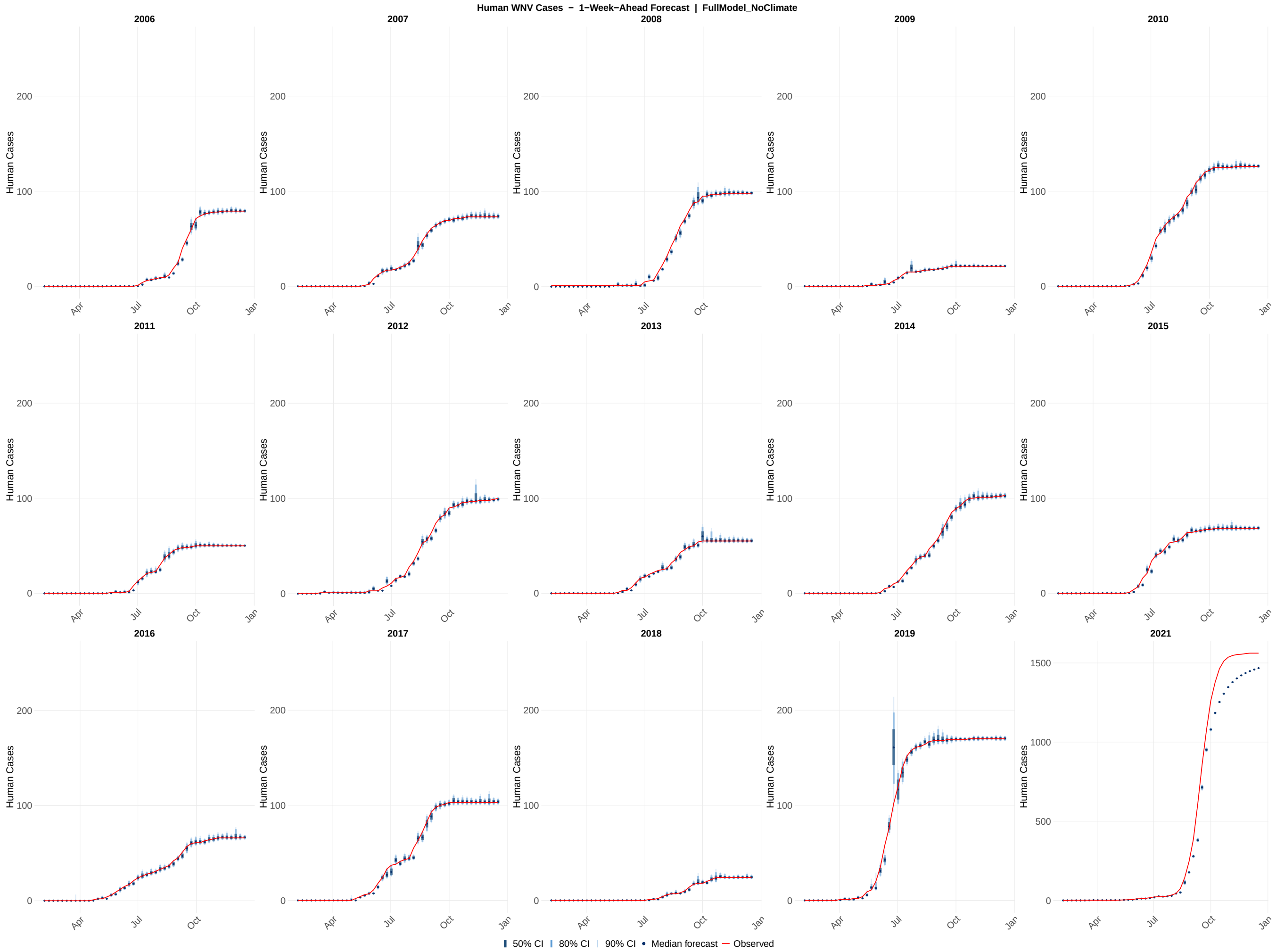

**Figure S29:** Probabilistic 1-week-ahead forecasts of human cases across all 15 study years (2006–2019, 2021) for the full model without weather configuration. Each panel shows the sequence of 1-week-ahead forecast distributions generated at each of the 46 assimilation steps within a single calendar year. Shaded ribbons indicate the 50% (darkest), 80% (medium), and 90% (lightest) prediction intervals; the solid line shows the forecast median; the red line shows the observed human cases values. The y-axis scale for 2021 is unconstrained; all other years share a common axis scale. Forecasts were generated by drawing 1,000 samples from the EnKF posterior at each assimilation step, propagating the time-varying parameters one week forward via the Ornstein–Uhlenbeck process, and integrating the ODE model through the forecast horizon.

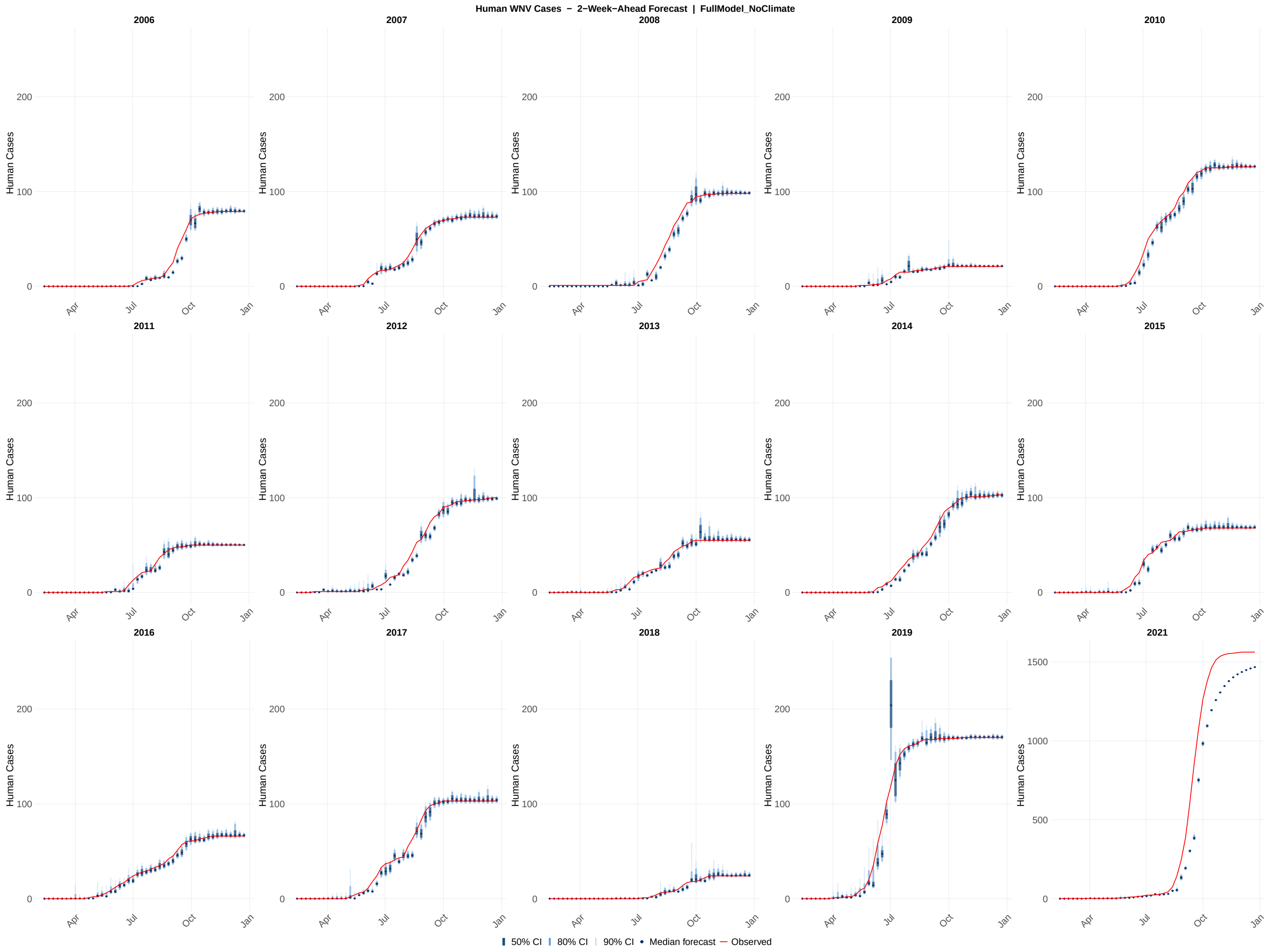

**Figure S30:** Probabilistic 2-week-ahead forecasts of human cases across all 15 study years (2006–2019, 2021) for the full model without weather configuration. Each panel shows the sequence of 2-week-ahead forecast distributions generated at each of the 46 assimilation steps within a single calendar year. Shaded ribbons indicate the 50% (darkest), 80% (medium), and 90% (lightest) prediction intervals; the solid line shows the forecast median; the red line shows the observed human cases values. The y-axis scale for 2021 is unconstrained; all other years share a common axis scale. Forecasts were generated by drawing 1,000 samples from the EnKF posterior at each assimilation step and propagating the time-varying parameters two steps forward via the Ornstein–Uhlenbeck process, with each successive step conditioned on the preceding OU-propagated parameter value.

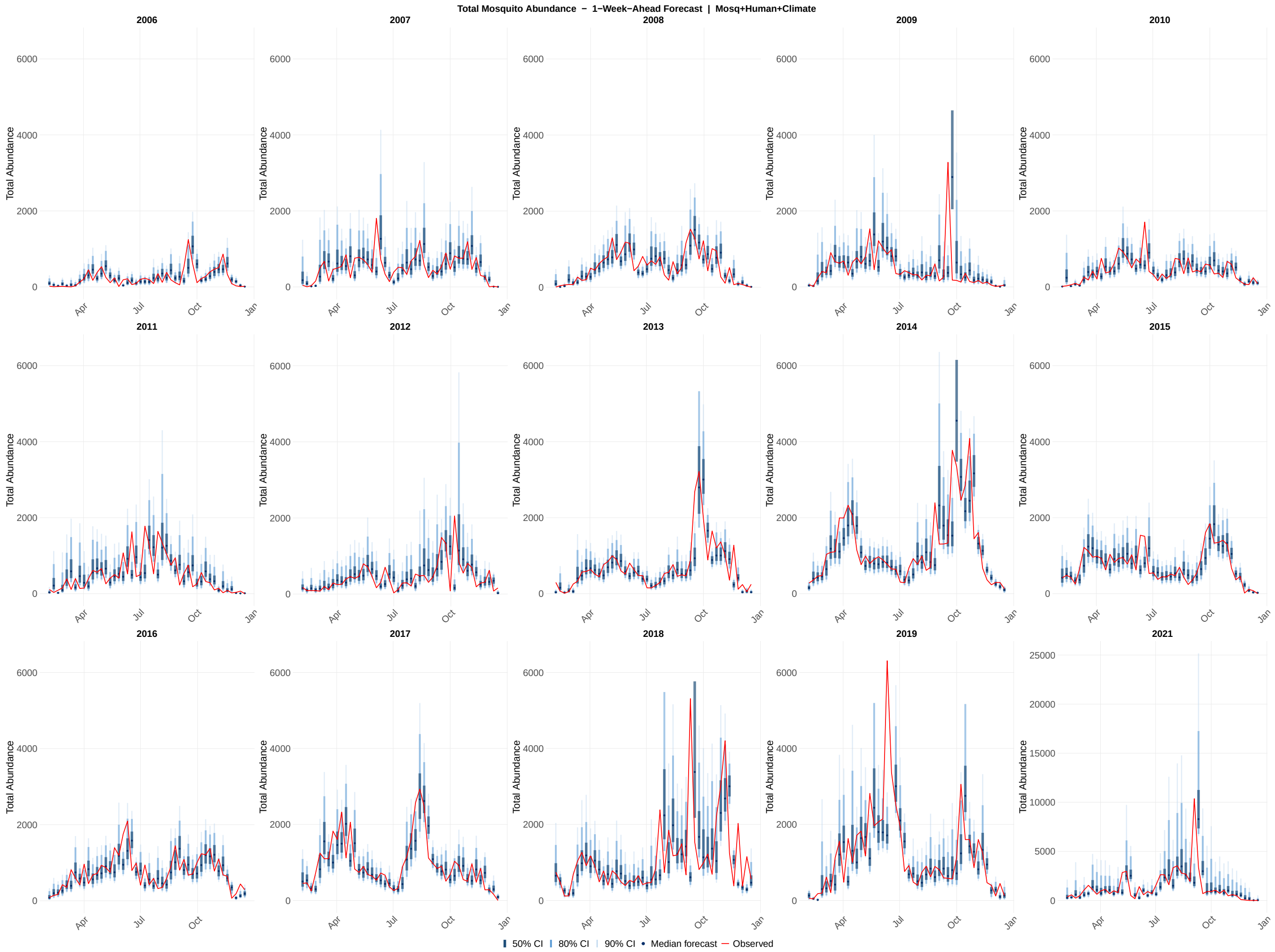

**Figure S31:** Probabilistic 1-week-ahead forecasts of total abundance across all 15 study years (2006–2019, 2021) for the mosquito + human with weather model configuration. Each panel shows the sequence of 1-week-ahead forecast distributions generated at each of the 46 assimilation steps within a single calendar year. Shaded ribbons indicate the 50% (darkest), 80% (medium), and 90% (lightest) prediction intervals; the solid line shows the forecast median; the red line shows the observed total abundance values. The y-axis scale for 2021 is unconstrained; all other years share a common axis scale. Forecasts were generated by drawing 1,000 samples from the EnKF posterior at each assimilation step, propagating the time-varying parameters one week forward via the Ornstein-Uhlenbeck process, and integrating the ODE model through the forecast horizon.

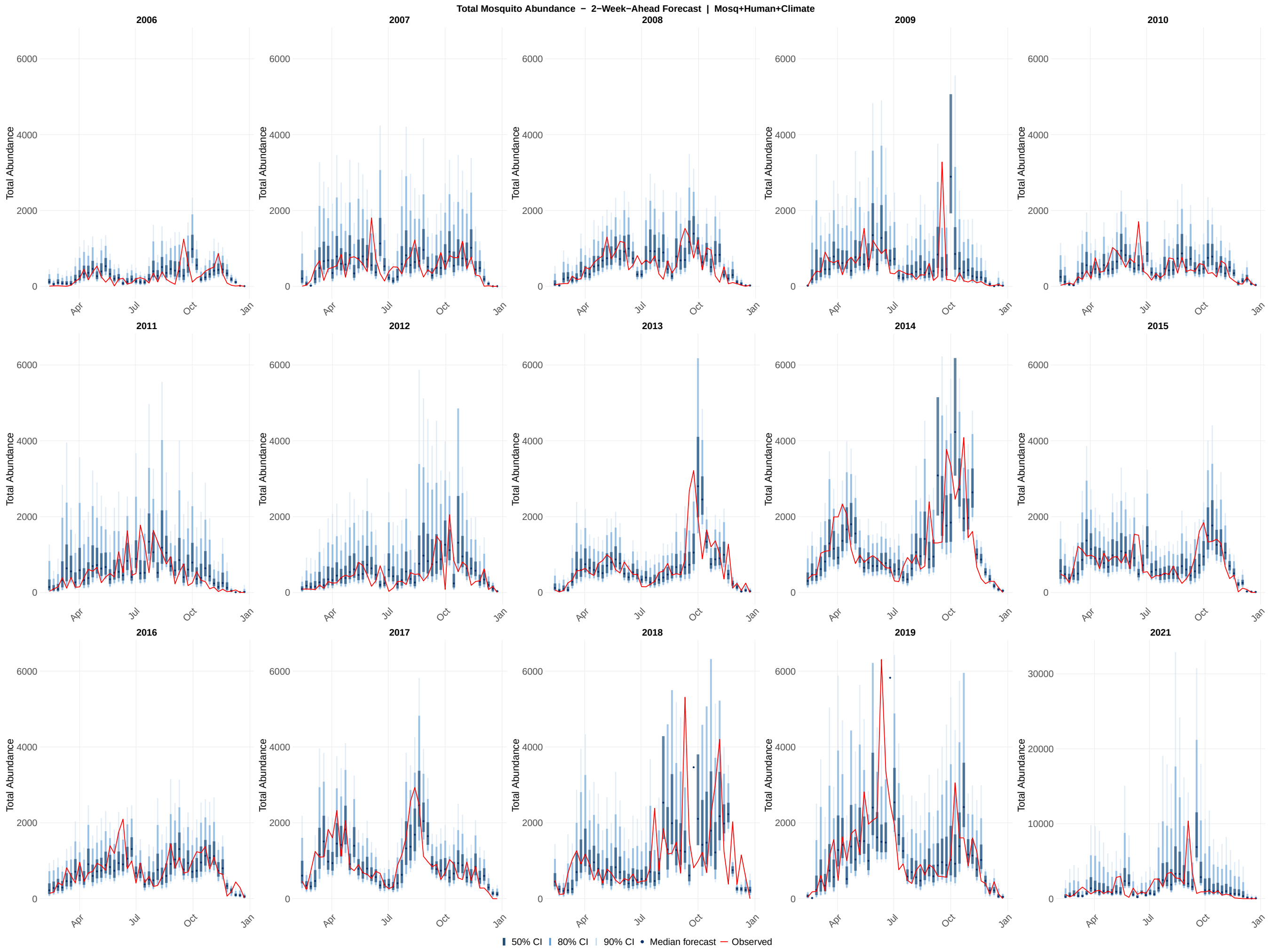

**Figure S32:** Probabilistic 2-week-ahead forecasts of total abundance across all 15 study years (2006–2019, 2021) for the mosquito + human with weather model configuration. Each panel shows the sequence of 2-week-ahead forecast distributions generated at each of the 46 assimilation steps within a single calendar year. Shaded ribbons indicate the 50% (darkest), 80% (medium), and 90% (lightest) prediction intervals; the solid line shows the forecast median; the red line shows the observed total abundance values. The y-axis scale for 2021 is unconstrained; all other years share a common axis scale. Forecasts were generated by drawing 1,000 samples from the EnKF posterior at each assimilation step and propagating the time-varying parameters two steps forward via the Ornstein-Uhlenbeck process, with each successive step conditioned on the preceding OU-propagated parameter value.

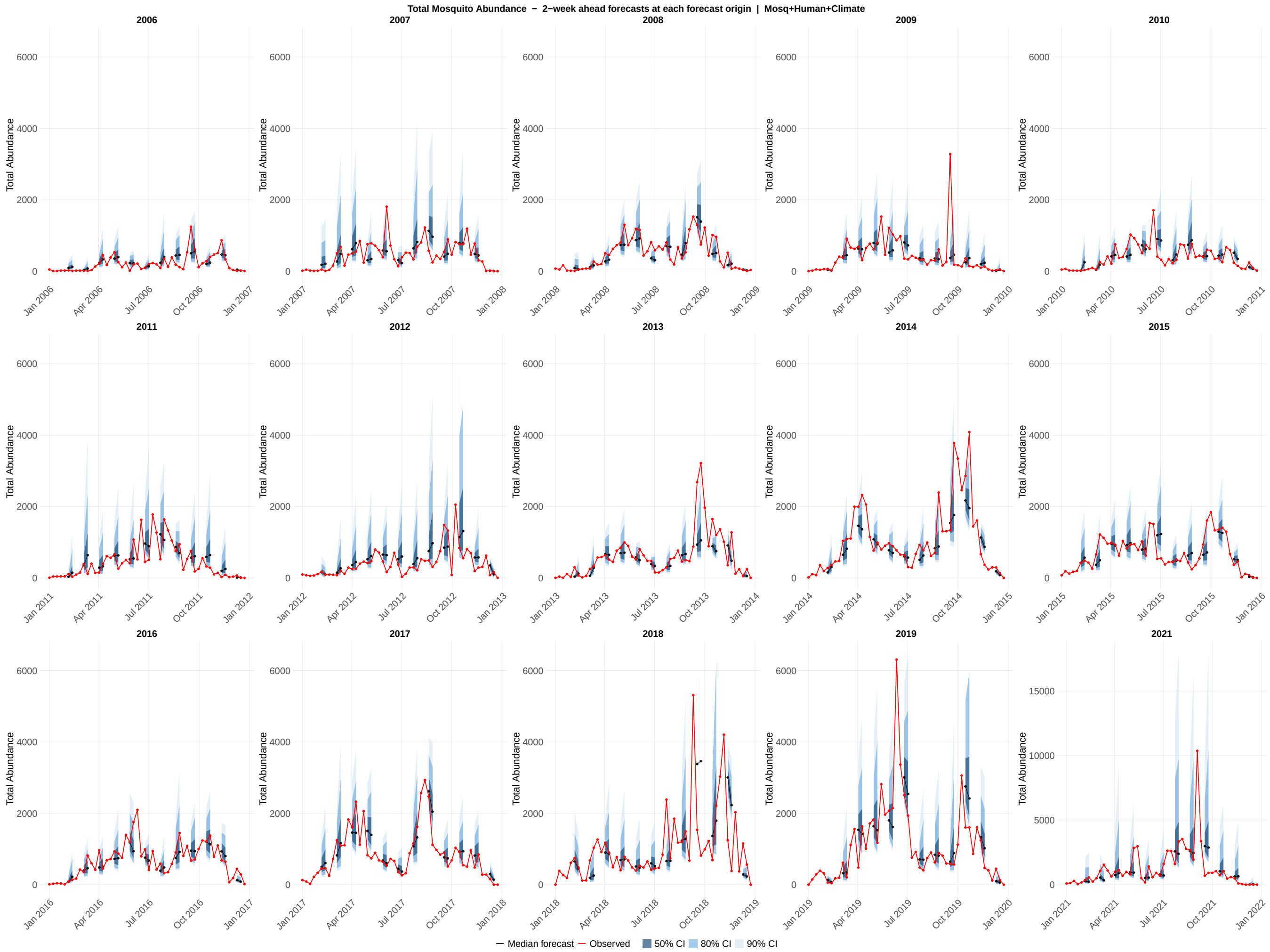

**Figure S33:** Probabilistic forecasts of total abundance across all 15 study years (2006–2019, 2021) for the mosquito + human with weather model configuration. Each panel displays an overlapping 2-week-ahead forecast generated at every fourth assimilation step within a single calendar year, providing a visual summary of how forecast uncertainty evolves across the transmission season. Each forecast horizon originates at a forecast origin date and extends through the 1-week-ahead and 2-week-ahead target dates; shaded ribbons show the 50% (darkest), 80% (medium), and 90% (lightest) prediction intervals at those two horizons; the colored line connecting the two horizon points shows the median forecast trajectory; the red line shows the observed total abundance values. The y-axis scale for 2021 is unconstrained; all other years share a common axis scale.

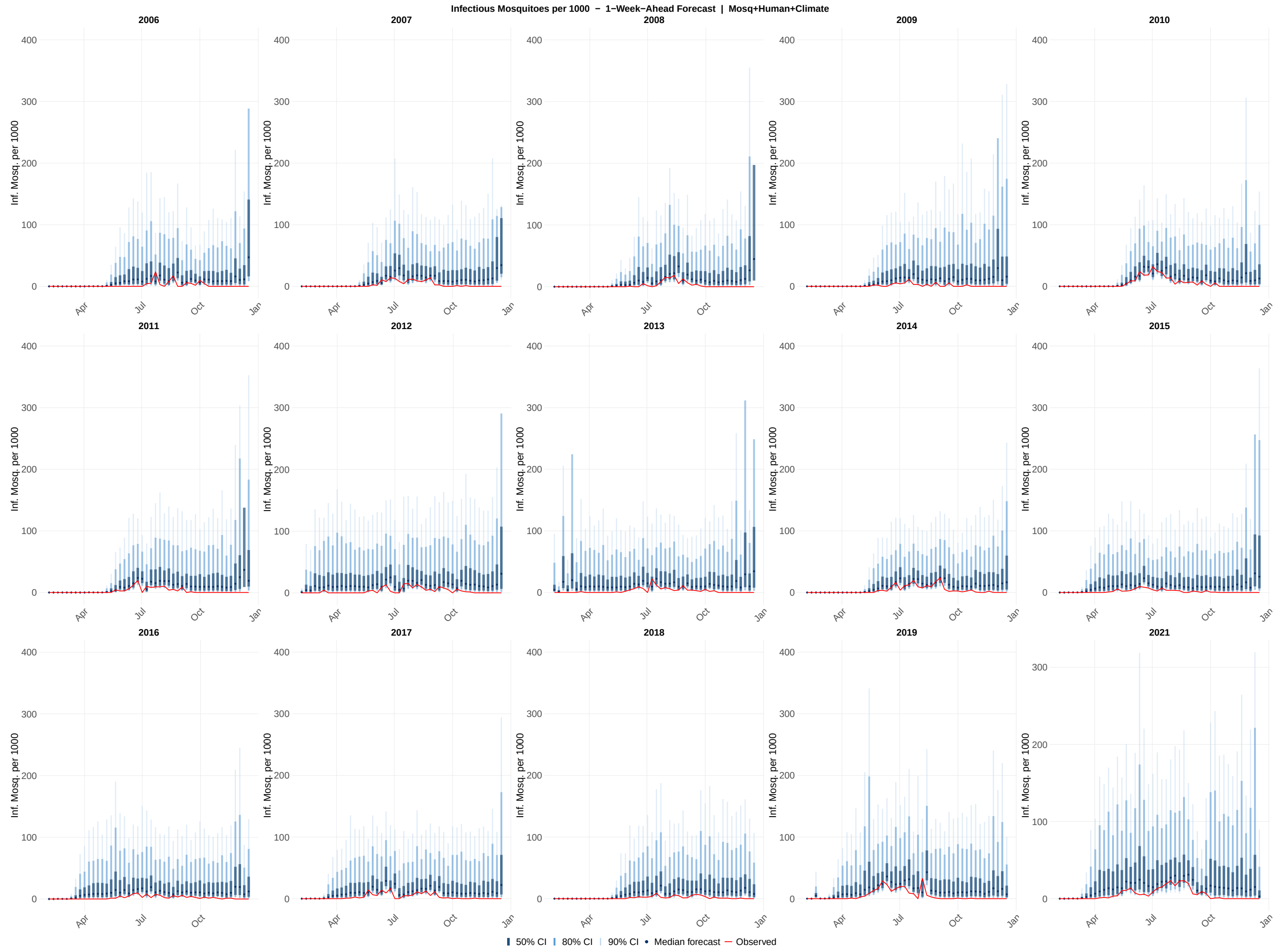

**Figure S34:** Probabilistic 1-week-ahead forecasts of infectious mosquitoes per 1000 across all 15 study years (2006–2019, 2021) for the mosquito + human with weather model configuration. Each panel shows the sequence of 1-week-ahead forecast distributions generated at each of the 46 assimilation steps within a single calendar year. Shaded ribbons indicate the 50% (darkest), 80% (medium), and 90% (lightest) prediction intervals; the solid line shows the forecast median; the red line shows the observed infectious mosquito per 1000 values. The y-axis scale for 2021 is unconstrained; all other years share a common axis scale. Forecasts were generated by drawing 1,000 samples from the EnKF posterior at each assimilation step, propagating the time-varying parameters one week forward via the Ornstein-Uhlenbeck process, and integrating the ODE model through the forecast horizon.

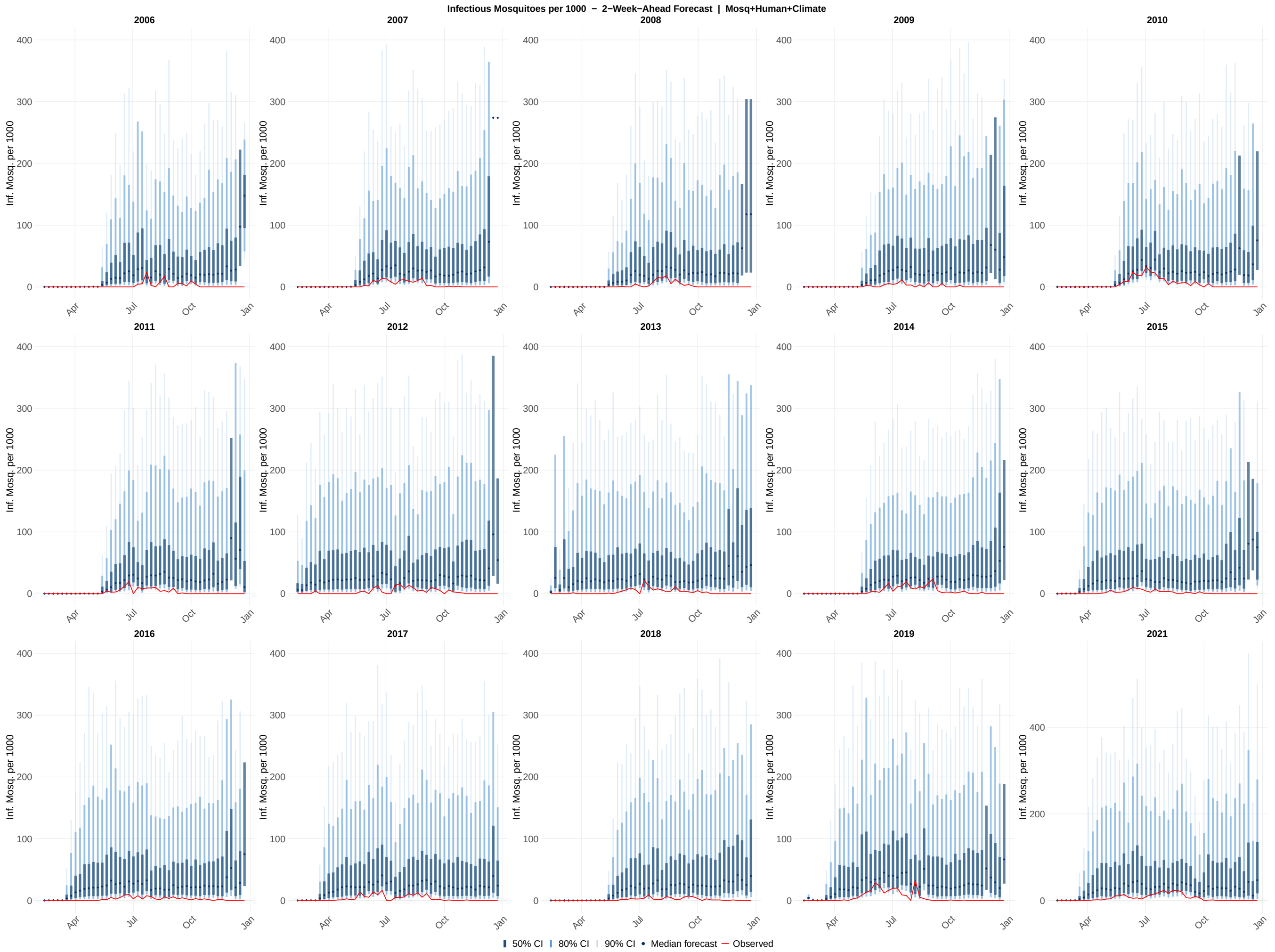

**Figure S35:** Probabilistic 2-week-ahead forecasts of infectious mosquito per 1000 across all 15 study years (2006–2019, 2021) for the mosquito + human with weather model configuration. Each panel shows the sequence of 2-week-ahead forecast distributions generated at each of the 46 assimilation steps within a single calendar year. Shaded ribbons indicate the 50% (darkest), 80% (medium), and 90% (lightest) prediction intervals; the solid line shows the forecast median; and the red line shows the observed infectious mosquito per 1000 values. The y-axis scale for 2021 is unconstrained; all other years share a common axis scale. Forecasts were generated by drawing 1,000 samples from the EnKF posterior at each assimilation step and propagating the time-varying parameters two steps forward via the Ornstein-Uhlenbeck process, with each successive step conditioned on the preceding OU-propagated parameter value.

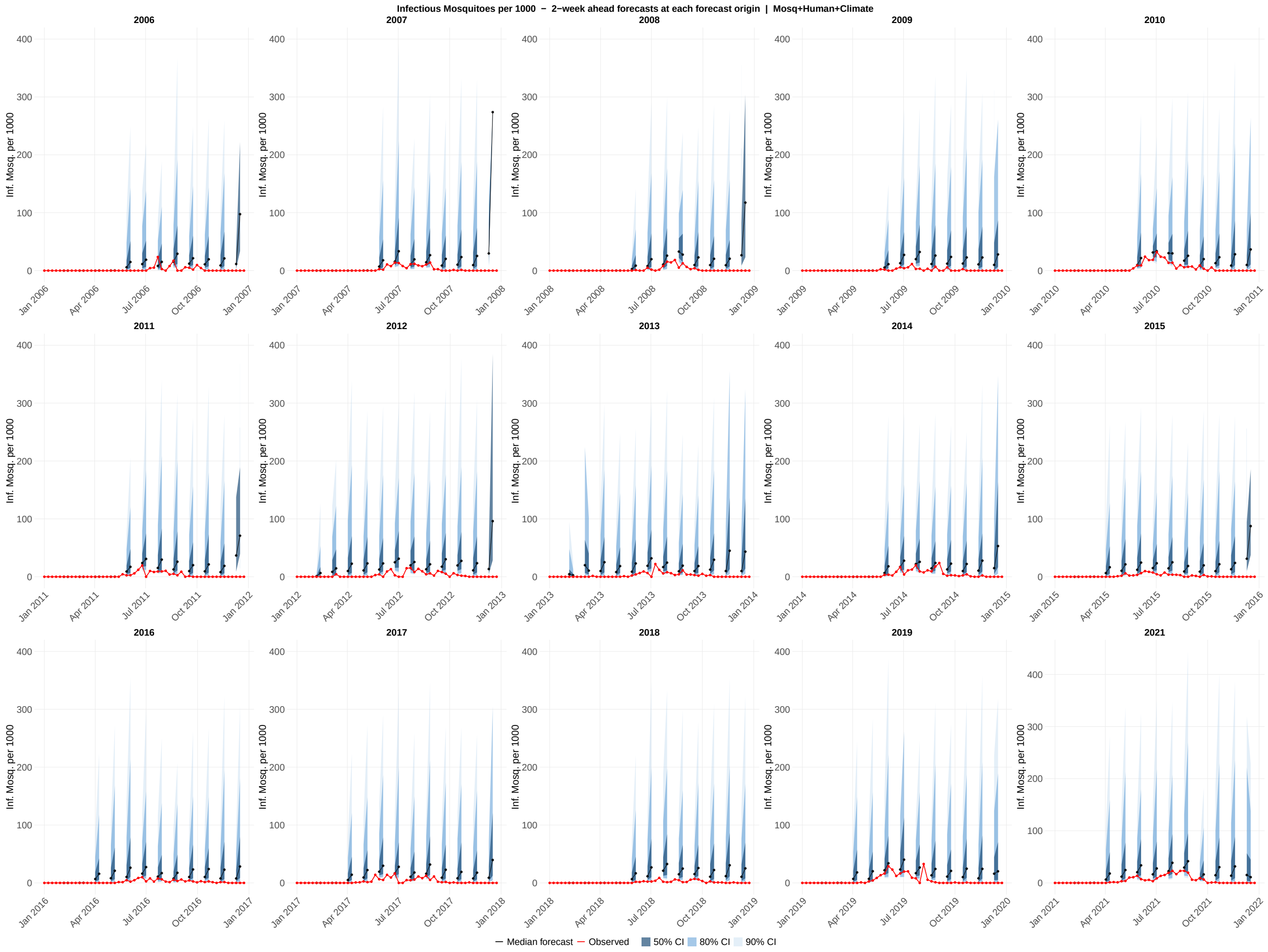

**Figure S36:** Probabilistic forecasts of infectious mosquitoes per 1000 all 15 study years (2006–2019, 2021) for the mosquito + human with weather model configuration. Each panel displays overlapping 2-week-ahead forecast generated at every fourth assimilation step within a single calendar year, providing a visual summary of how forecast uncertainty evolves across the transmission season. Each forecast horizon originates at a forecast origin date and extends through the 1-week-ahead and 2-week-ahead target dates; shaded ribbons show the 50% (darkest), 80% (medium), and 90% (lightest) prediction intervals at those two horizons; the colored line connecting the two horizon points shows the median forecast trajectory; the red line shows the observed infectious mosquito per 1000 values. The y-axis scale for 2021 is unconstrained; all other years share a common axis scale.

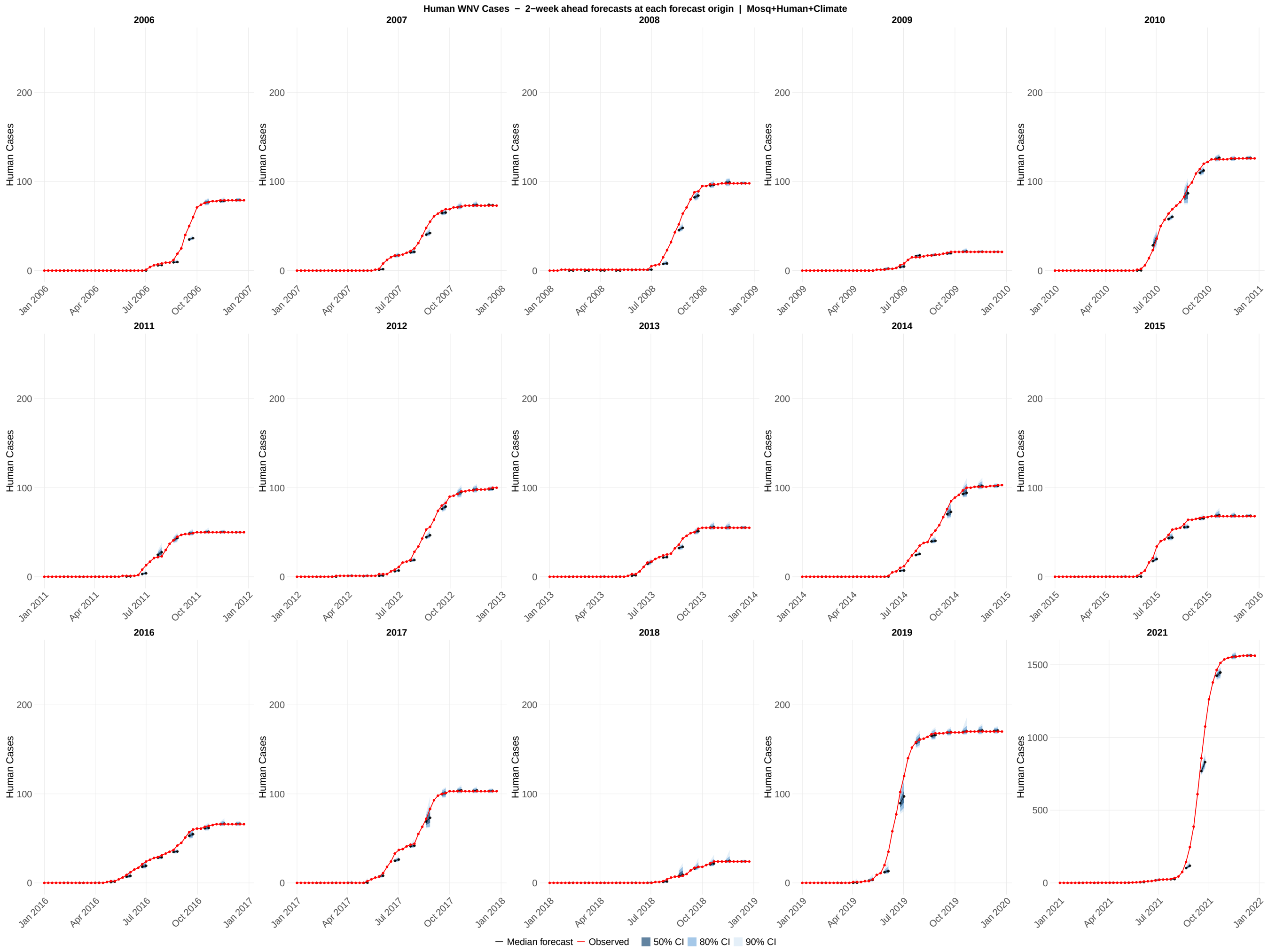

**Figure S37:** Probabilistic forecasts of human cases across all 15 study years (2006–2019, 2021) for the mosquito + human with weather model configuration. Each panel displays overlapping 2-week-ahead forecast generated at every fourth assimilation step within a single calendar year, providing a visual summary of how forecast uncertainty evolves across the transmission season. Each forecast horizon originates at a forecast origin date and extends through the 1-week-ahead and 2-week-ahead target dates; shaded ribbons show the 50% (darkest), 80% (medium), and 90% (lightest) prediction intervals at those two horizons; the colored line connecting the two horizon points shows the median forecast trajectory; the red line shows the observed human cases values. The y-axis scale for 2021 is unconstrained; all other years share a common axis scale.

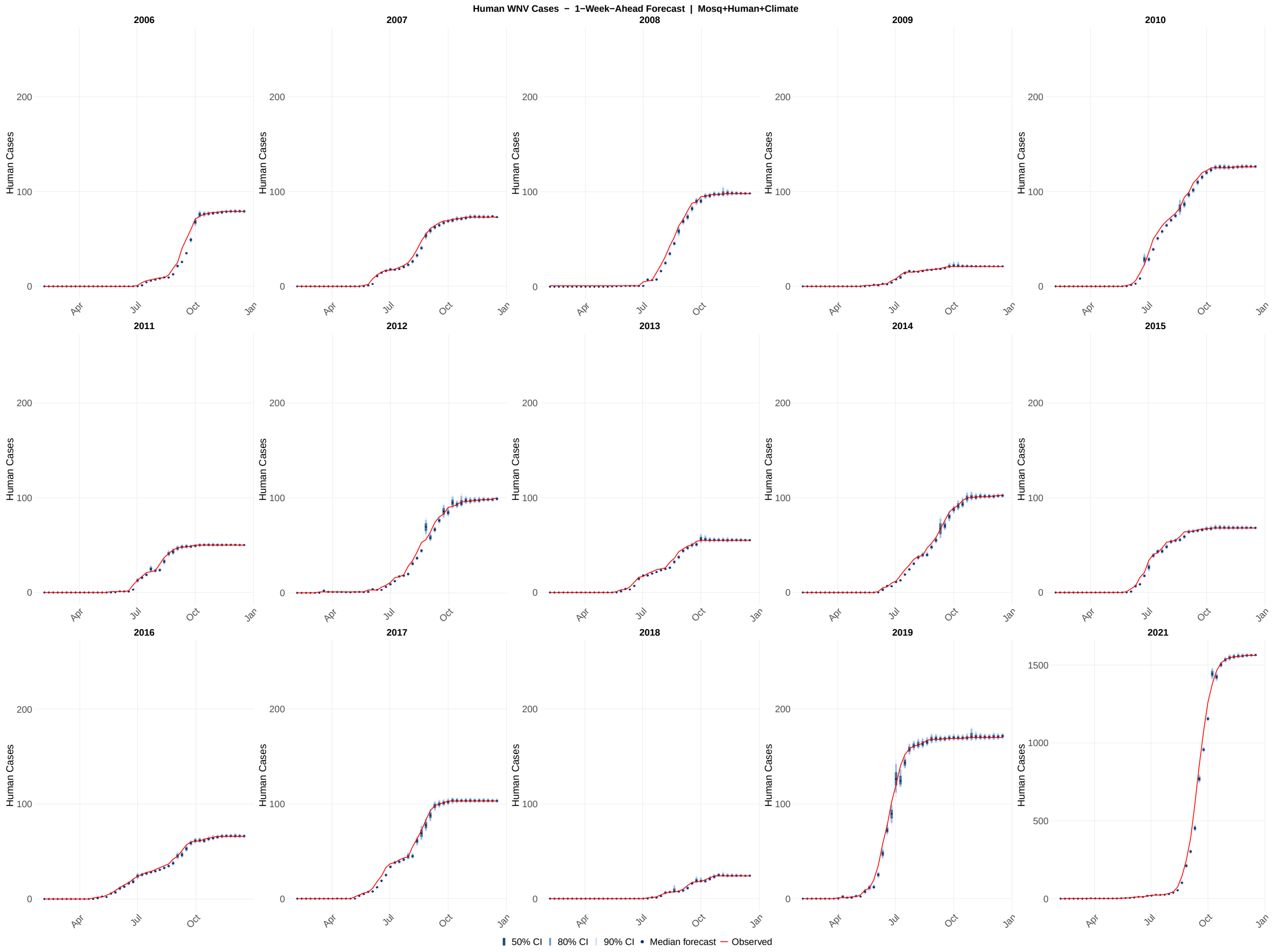

**Figure S38:** Probabilistic 1-week-ahead forecasts of human cases across all 15 study years (2006–2019, 2021) for the mosquito + human with weather model configuration. Each panel shows the sequence of 1-week-ahead forecast distributions generated at each of the 46 assimilation steps within a single calendar year. Shaded ribbons indicate the 50% (darkest), 80% (medium), and 90% (lightest) prediction intervals; the solid line shows the forecast median; the red line shows the observed human cases values. The y-axis scale for 2021 is unconstrained; all other years share a common axis scale. Forecasts were generated by drawing 1,000 samples from the EnKF posterior at each assimilation step, propagating the time-varying parameters one week forward via the Ornstein–Uhlenbeck process, and integrating the ODE model through the forecast horizon.

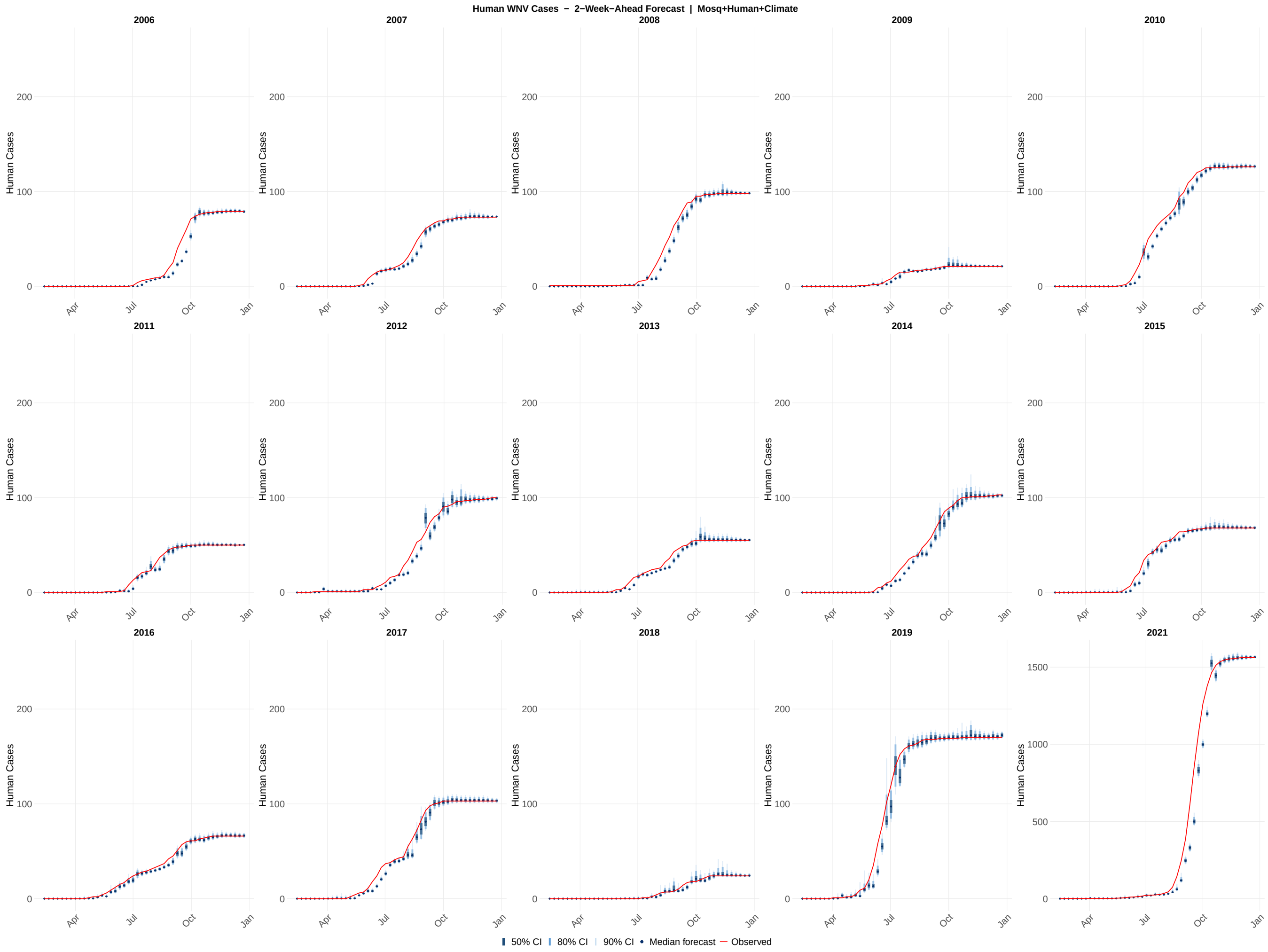

**Figure S39:** Probabilistic 2-week-ahead forecasts of human cases across all 15 study years (2006–2019, 2021) for the mosquito + human with weather model configuration. Each panel shows the sequence of 2-week-ahead forecast distributions generated at each of the 46 assimilation steps within a single calendar year. Shaded ribbons indicate the 50% (darkest), 80% (medium), and 90% (lightest) prediction intervals; the solid line shows the forecast median; the red line shows the observed human cases values. The y-axis scale for 2021 is unconstrained; all other years share a common axis scale. Forecasts were generated by drawing 1,000 samples from the EnKF posterior at each assimilation step and propagating the time-varying parameters two steps forward via the Ornstein–Uhlenbeck process, with each successive step conditioned on the preceding OU-propagated parameter value.

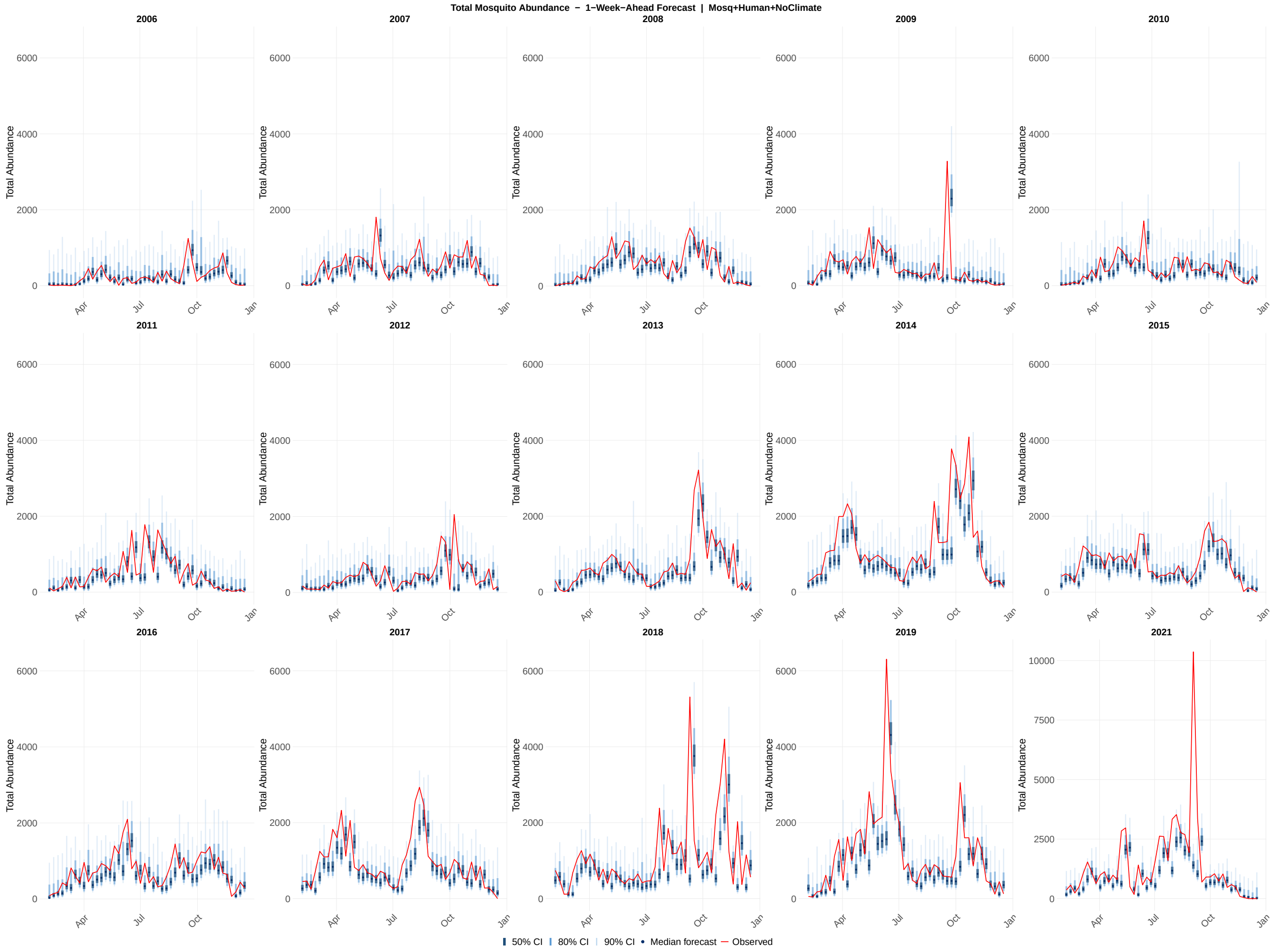

**Figure S40:** Probabilistic 1-week-ahead forecasts of total abundance across all 15 study years (2006–2019, 2021) for the mosquito + human without weather model configuration. Each panel shows the sequence of 1-week-ahead forecast distributions generated at each of the 46 assimilation steps within a single calendar year. Shaded ribbons indicate the 50% (darkest), 80% (medium), and 90% (lightest) prediction intervals; the solid line shows the forecast median; the red line shows the observed total abundance values. The y-axis scale for 2021 is unconstrained; all other years share a common axis scale. Forecasts were generated by drawing 1,000 samples from the EnKF posterior at each assimilation step, propagating the time-varying parameters one week forward via the Ornstein-Uhlenbeck process, and integrating the ODE model through the forecast horizon.

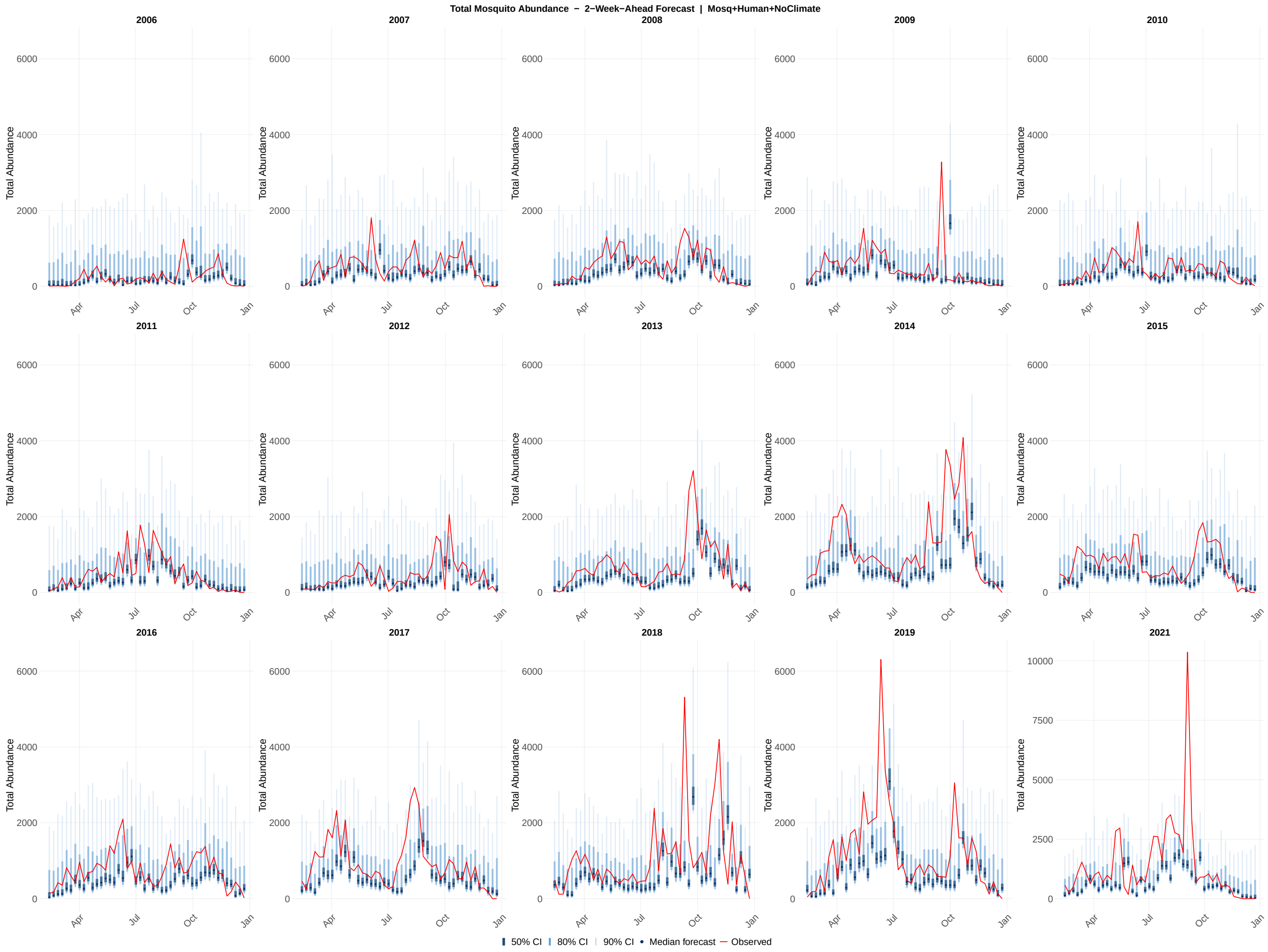

**Figure S41:** Probabilistic 2-week-ahead forecasts of total abundance across all 15 study years (2006–2019, 2021) for the mosquito + human without weather model configuration. Each panel shows the sequence of 2-week-ahead forecast distributions generated at each of the 46 assimilation steps within a single calendar year. Shaded ribbons indicate the 50% (darkest), 80% (medium), and 90% (lightest) prediction intervals; the solid line shows the forecast median; the red line shows the observed total abundance values. The y-axis scale for 2021 is unconstrained; all other years share a common axis scale. Forecasts were generated by drawing 1,000 samples from the EnKF posterior at each assimilation step and propagating the time-varying parameters two steps forward via the Ornstein-Uhlenbeck process, with each successive step conditioned on the preceding OU-propagated parameter value.

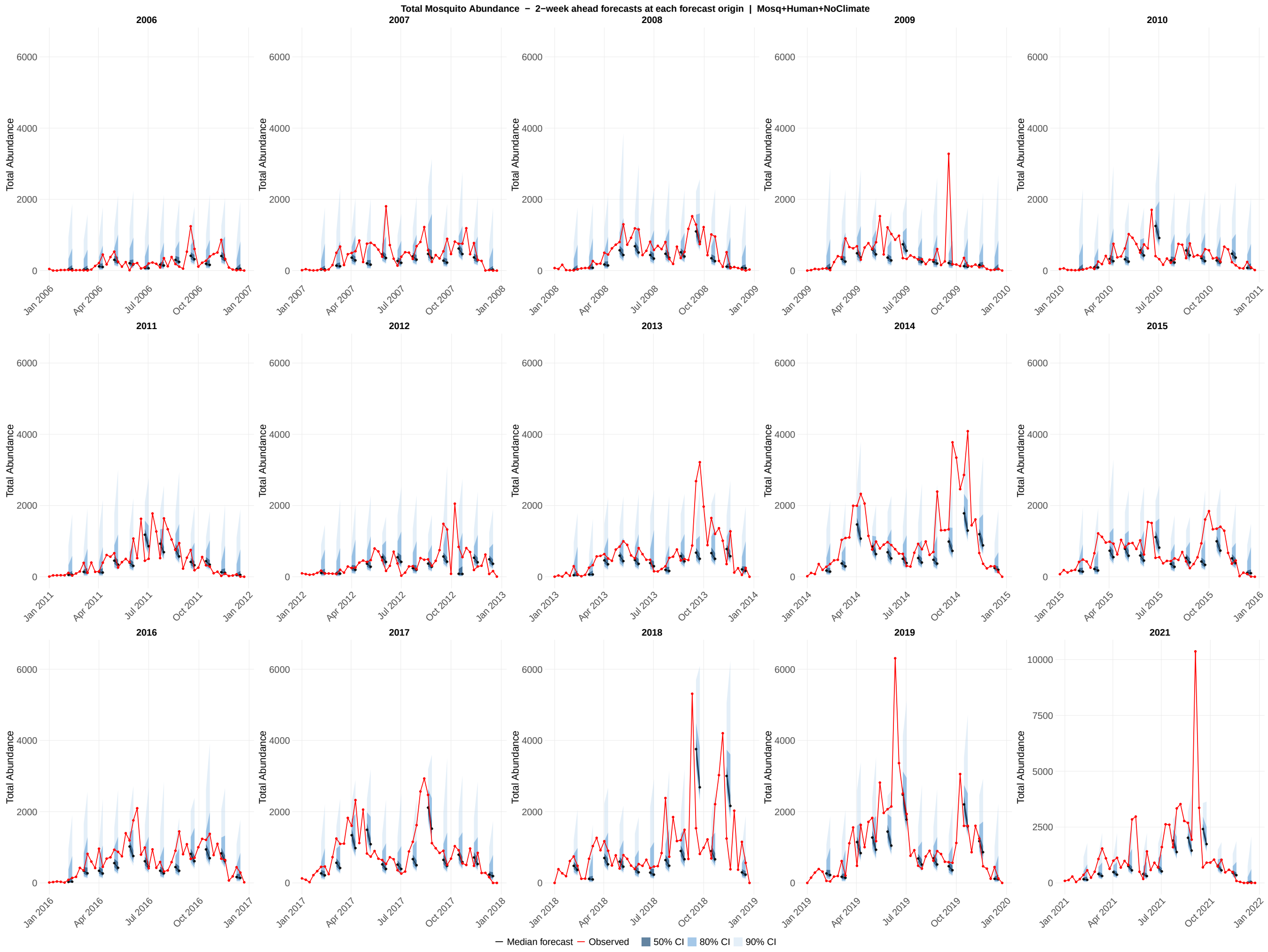

**Figure S42:** Probabilistic forecasts of total abundance across all 15 study years (2006–2019, 2021) for the mosquito + human without weather model configuration. Each panel displays an overlapping 2-week-ahead forecast generated at every fourth assimilation step within a single calendar year, providing a visual summary of how forecast uncertainty evolves across the transmission season. Each forecast horizon originates at a forecast origin date and extends through the 1-week-ahead and 2-week-ahead target dates; shaded ribbons show the 50% (darkest), 80% (medium), and 90% (lightest) prediction intervals at those two horizons; the colored line connecting the two horizon points shows the median forecast trajectory; the red line shows the observed total abundance values. The y-axis scale for 2021 is unconstrained; all other years share a common axis scale.

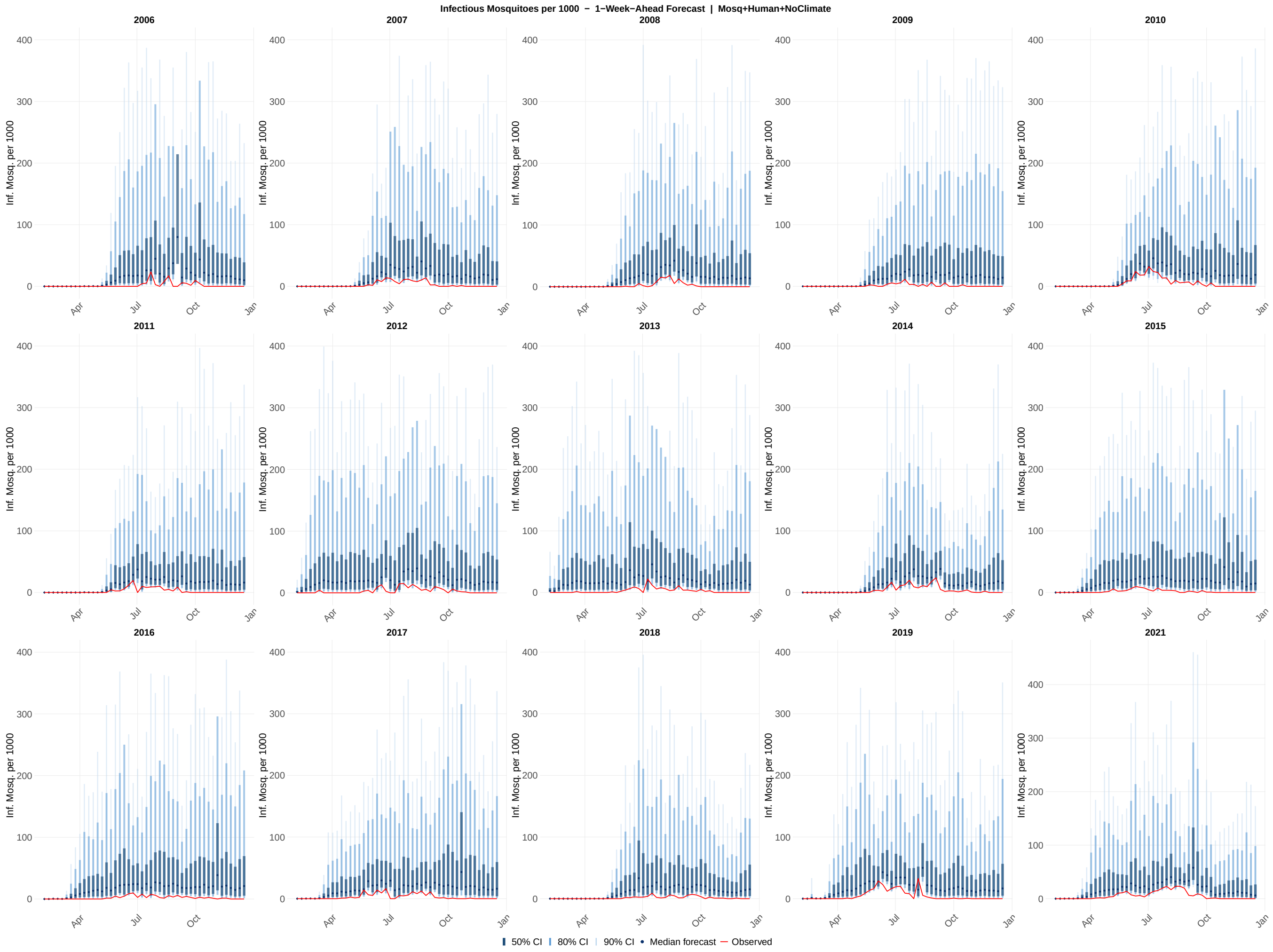

**Figure S43:** Probabilistic 1-week-ahead forecasts of infectious mosquitoes per 1000 across all 15 study years (2006–2019, 2021) for the mosquito + human without weather model configuration. Each panel shows the sequence of 1-week-ahead forecast distributions generated at each of the 46 assimilation steps within a single calendar year. Shaded ribbons indicate the 50% (darkest), 80% (medium), and 90% (lightest) prediction intervals; the solid line shows the forecast median; the red line shows the observed infectious mosquito per 1000 values. The y-axis scale for 2021 is unconstrained; all other years share a common axis scale. Forecasts were generated by drawing 1,000 samples from the EnKF posterior at each assimilation step, propagating the time-varying parameters one week forward via the Ornstein-Uhlenbeck process, and integrating the ODE model through the forecast horizon.

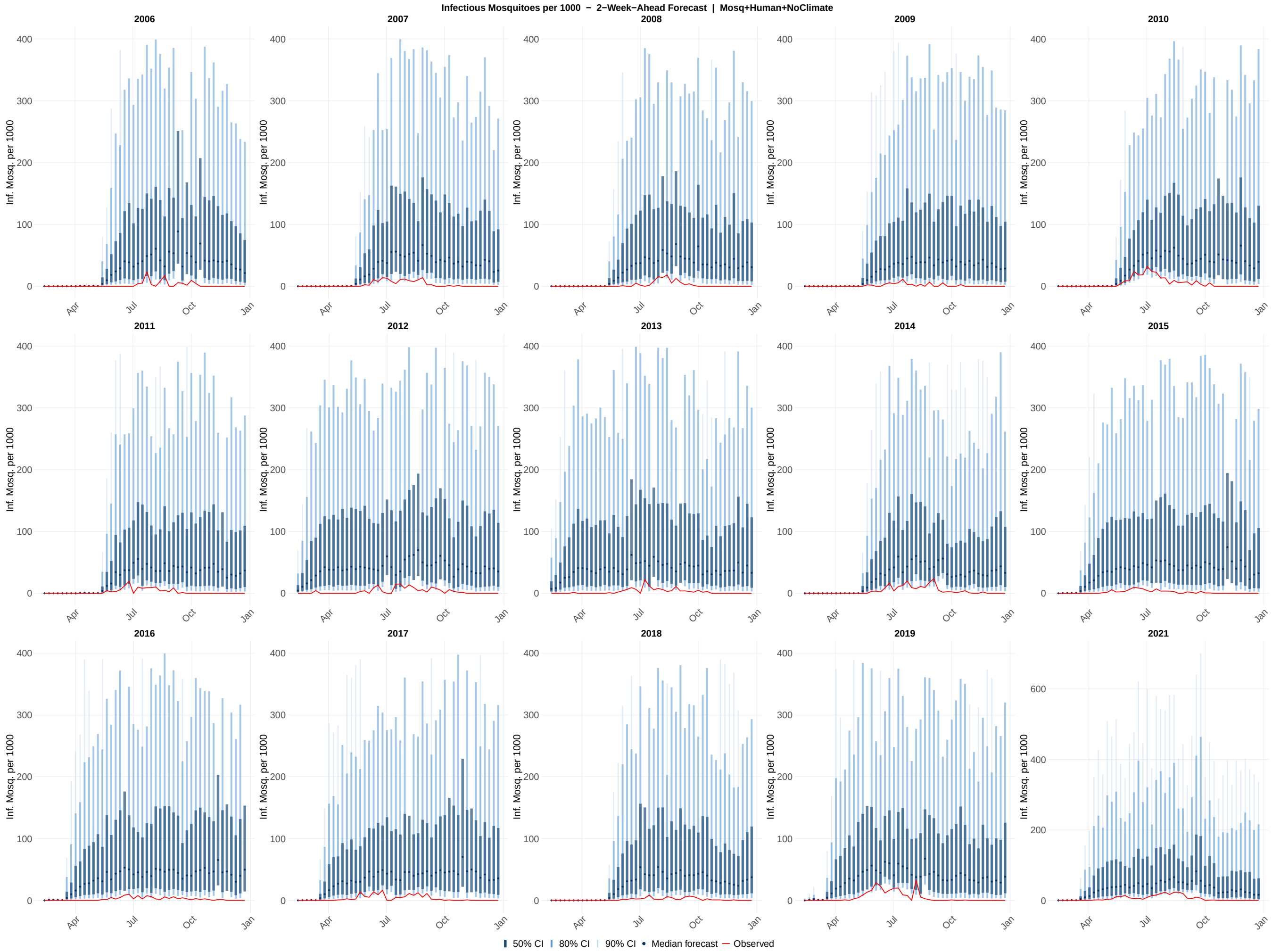

**Figure S44:** Probabilistic 2-week-ahead forecasts of infectious mosquito per 1000 across all 15 study years (2006–2019, 2021) for the mosquito + human without weather model configuration. Each panel shows the sequence of 2-week-ahead forecast distributions generated at each of the 46 assimilation steps within a single calendar year. Shaded ribbons indicate the 50% (darkest), 80% (medium), and 90% (lightest) prediction intervals; the solid line shows the forecast median; and the red line shows the observed infectious mosquito per 1000 values. The y-axis scale for 2021 is unconstrained; all other years share a common axis scale. Forecasts were generated by drawing 1,000 samples from the EnKF posterior at each assimilation step and propagating the time-varying parameters two steps forward via the Ornstein-Uhlenbeck process, with each successive step conditioned on the preceding OU-propagated parameter value.

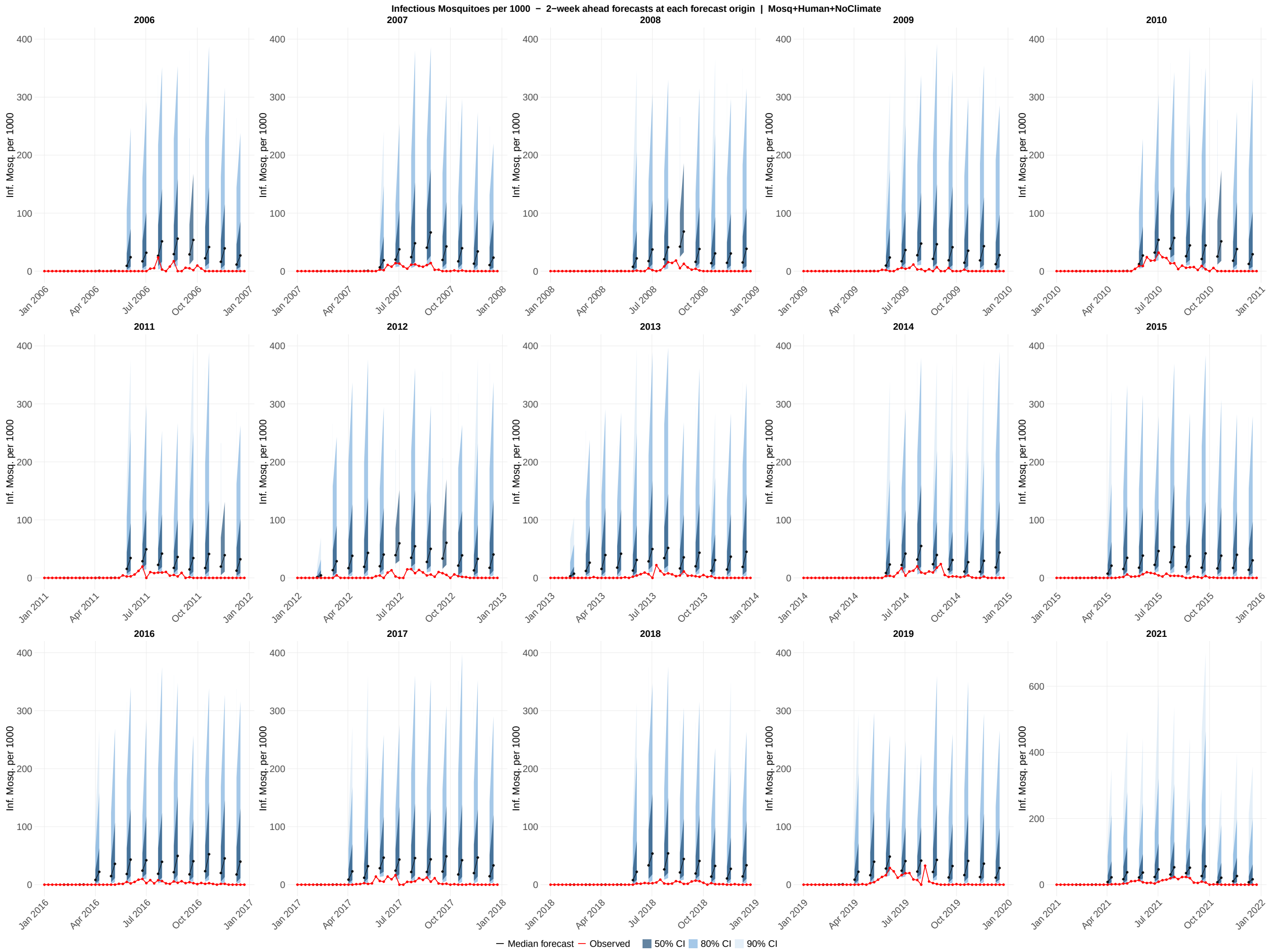

**Figure S45:** Probabilistic forecasts of infectious mosquitoes per 1000 all 15 study years (2006–2019, 2021) for the mosquito + human without weather model configuration. Each panel displays overlapping 2-week-ahead forecast generated at every fourth assimilation step within a single calendar year, providing a visual summary of how forecast uncertainty evolves across the transmission season. Each forecast horizon originates at a forecast origin date and extends through the 1-week-ahead and 2-week-ahead target dates; shaded ribbons show the 50% (darkest), 80% (medium), and 90% (lightest) prediction intervals at those two horizons; the colored line connecting the two horizon points shows the median forecast trajectory; the red line shows the observed infectious mosquito per 1000 values. The y-axis scale for 2021 is unconstrained; all other years share a common axis scale.

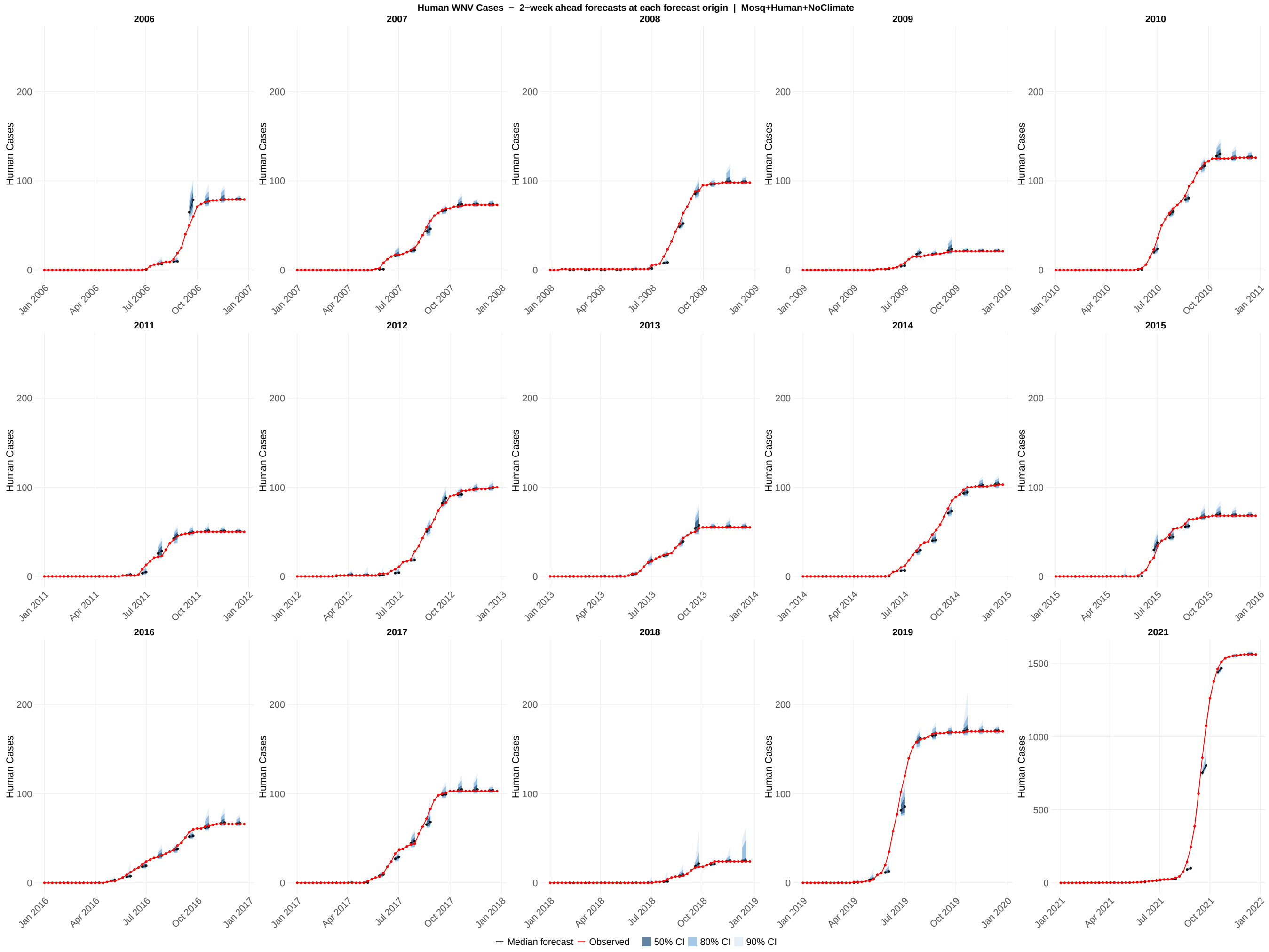

**Figure S46:** Probabilistic forecasts of human cases across all 15 study years (2006–2019, 2021) for the mosquito + human without weather model configuration. Each panel displays overlapping 2-week-ahead forecast generated at every fourth assimilation step within a single calendar year, providing a visual summary of how forecast uncertainty evolves across the transmission season. Each forecast horizon originates at a forecast origin date and extends through the 1-week-ahead and 2-week-ahead target dates; shaded ribbons show the 50% (darkest), 80% (medium), and 90% (lightest) prediction intervals at those two horizons; the colored line connecting the two horizon points shows the median forecast trajectory; the red line shows the observed human cases values. The y-axis scale for 2021 is unconstrained; all other years share a common axis scale.

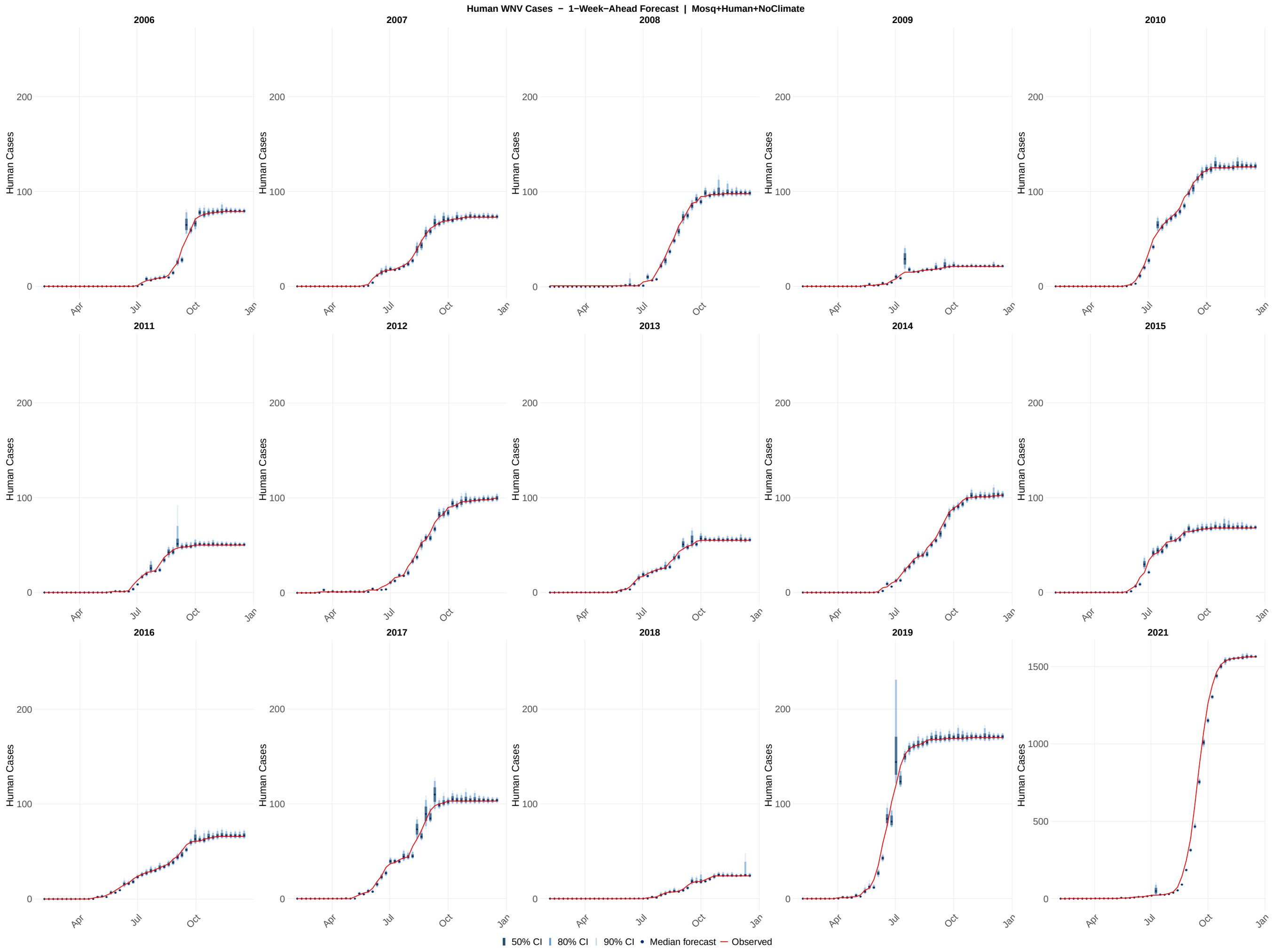

**Figure S47:** Probabilistic 1-week-ahead forecasts of human cases across all 15 study years (2006–2019, 2021) for the mosquito + human without weather model configuration. Each panel shows the sequence of 1-week-ahead forecast distributions generated at each of the 46 assimilation steps within a single calendar year. Shaded ribbons indicate the 50% (darkest), 80% (medium), and 90% (lightest) prediction intervals; the solid line shows the forecast median; the red line shows the observed human cases values. The y-axis scale for 2021 is unconstrained; all other years share a common axis scale. Forecasts were generated by drawing 1,000 samples from the EnKF posterior at each assimilation step, propagating the time-varying parameters one week forward via the Ornstein–Uhlenbeck process, and integrating the ODE model through the forecast horizon.

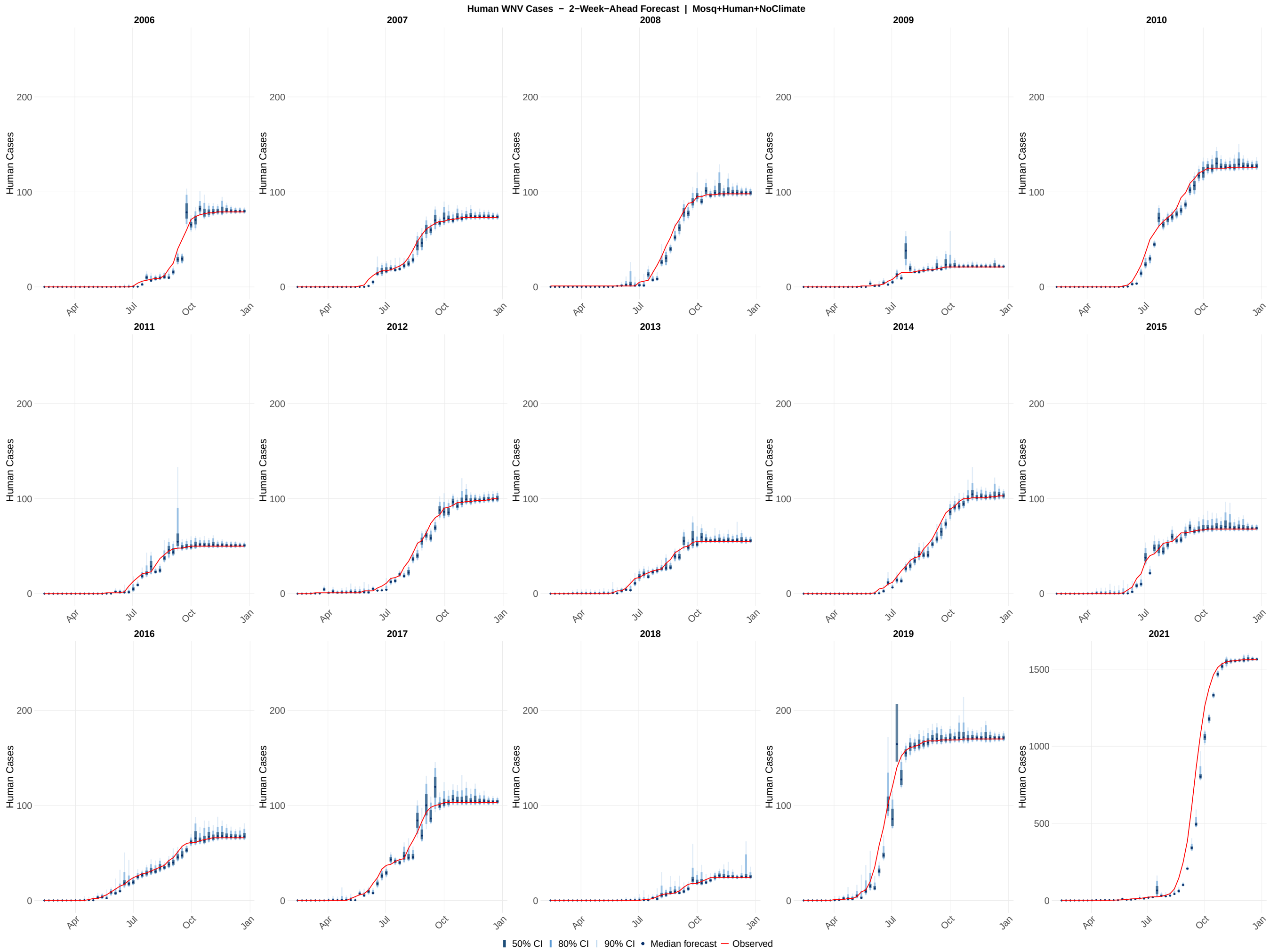

**Figure S48:** Probabilistic 2-week-ahead forecasts of human cases across all 15 study years (2006–2019, 2021) for the mosquito + human without weather model configuration. Each panel shows the sequence of 2-week-ahead forecast distributions generated at each of the 46 assimilation steps within a single calendar year. Shaded ribbons indicate the 50% (darkest), 80% (medium), and 90% (lightest) prediction intervals; the solid line shows the forecast median; the red line shows the observed human cases values. The y-axis scale for 2021 is unconstrained; all other years share a common axis scale. Forecasts were generated by drawing 1,000 samples from the EnKF posterior at each assimilation step and propagating the time-varying parameters two steps forward via the Ornstein–Uhlenbeck process, with each successive step conditioned on the preceding OU-propagated parameter value.

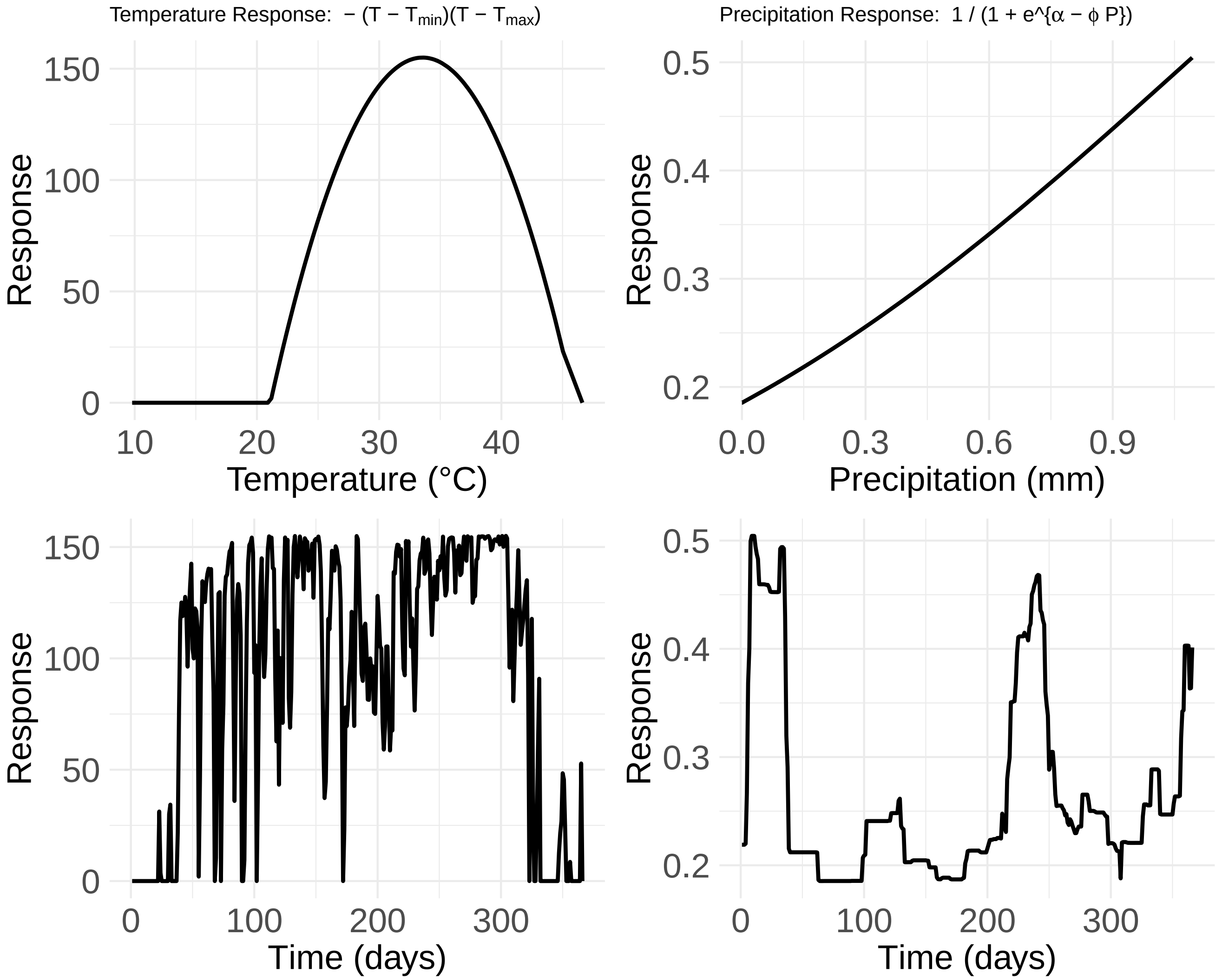

**Figure S49:** Temperature and precipitation response functions used in the model. We used 2016 temperature and precipitation data from PRISM. (**Top left**) Temperature response function defined as $-(T - T_{\text{min}})(T - T_{\text{max}})$, illustrating the parabolic relationship between temperature and suitability, with zero response below the minimum threshold $T_{\text{min}}$ and above the maximum threshold $T_{\text{max}}$, and maximum response at intermediate temperatures. (**Top right**) The precipitation response function is defined as $\frac{1}{1+\exp(\alpha-\phi P)}$, showing the sigmoidal increase in suitability with increasing daily precipitation, saturating at higher rainfall. (**Bottom left**) Time series of the temperature response across one year, showing variability in response driven by daily fluctuations in observed temperature. (**Bottom right**) Time series of the precipitation response across the same period, reflecting the nonlinear effects of rainfall pulses and dry periods on response values.

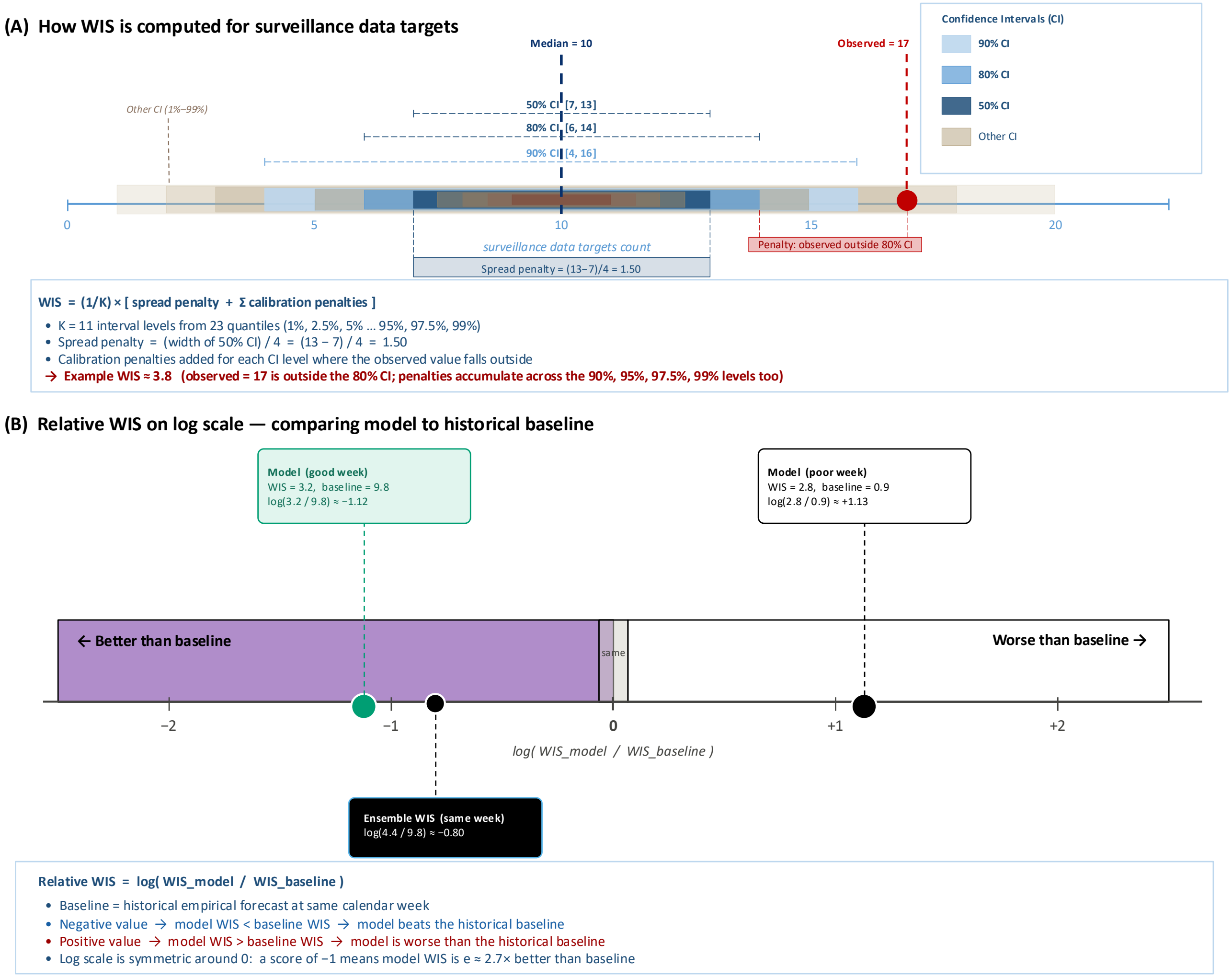

**Figure S50:** Schematic illustration of the Weighted Interval Score (WIS) and relative WIS used to evaluate probabilistic forecasts of cumulative human West Nile virus cases. (A) WIS computation for a representative forecast. The forecast is represented as 11 nested prediction intervals derived from 23 quantile levels (1%, 2.5%, 5%, . . . , 95%, 97.5%, 99%), shown as bars of decreasing width and increasing color intensity from the outermost (1%–99%, warm sand) to the innermost (50% CI, dark navy). The three highlighted intervals — 50% CI [7, 13], 80% CI [6, 14], and 90% CI [4, 16]—correspond to the standard reporting levels. The dashed navy vertical line indicates the median forecast (10 cases), and the red circle indicates the observed count (17 cases). WIS is computed as a weighted sum of a spread penalty (proportional to the width of the 50% CI) and calibration penalties incurred for each interval level where the observed value falls outside the forecast bounds; a lower WIS indicates a sharper and better-calibrated forecast. (B) Relative WIS on the natural log scale, used to benchmark each model against the historical baseline forecast. The baseline forecast at each calendar week was constructed from the empirical distribution of observed counts at the same week across all study years (2006–2019, 2021) using a leave-one-year-out approach. A relative WIS of zero indicates performance equivalent to the baseline; negative values (purple zone) indicate the model outperforms the baseline (model WIS $<$ baseline WIS); and positive values (white zone) indicate the model performs worse than the baseline (model WIS $>$ baseline WIS). Three example scores are shown: the Full Model during a peak transmission week (log ratio $\approx$ -1.12, green); the Full Model during an early-season week when the baseline is easier to beat (log ratio $\approx +1.13$, white); and the WIS-weighted ensemble (Ensemble model 4) during the same peak transmission week (log ratio $\approx$ -0.80, black). A log ratio of $-1$ corresponds to a model WIS approximately 2.7 times lower than the baseline.

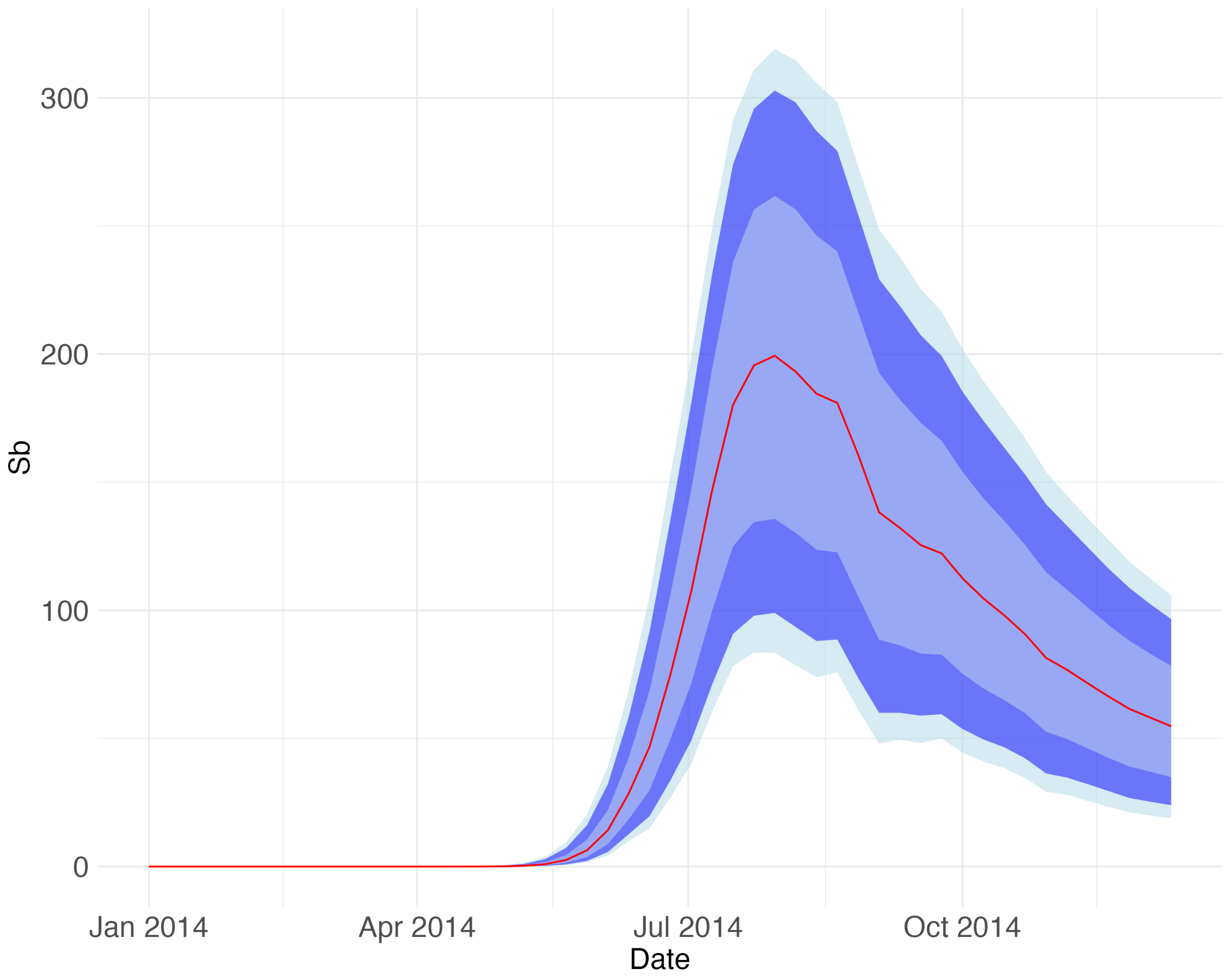

**Figure S51:** A representative figure for the shape of the simulated susceptible bird.

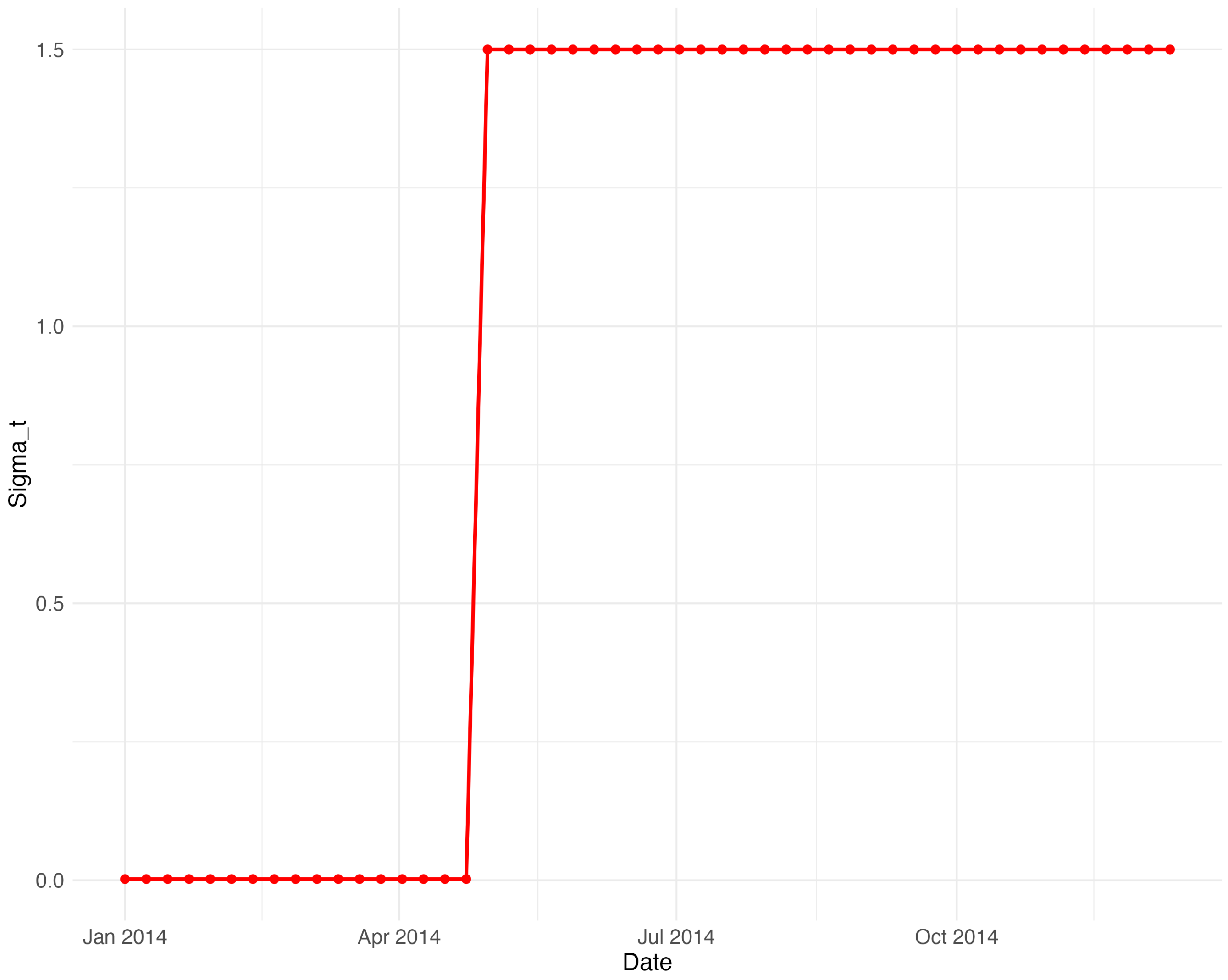

**Figure S52:** A representative figure for the shape of the diffusion parameter $\sigma$.

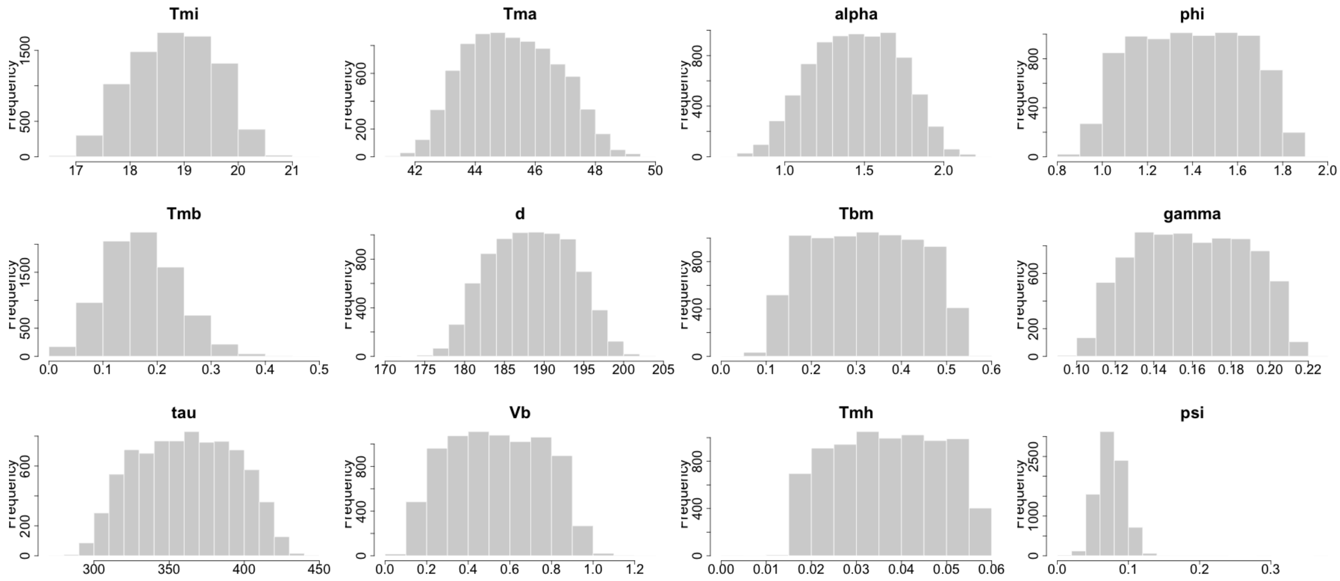

**Figure S53:** Histogram of the estimated static parameters for the full model for 2014. To access other model configurations and other years, visit the link: https://github.com/NAU-CCL/WNV-Forecasting and run the code for all models.

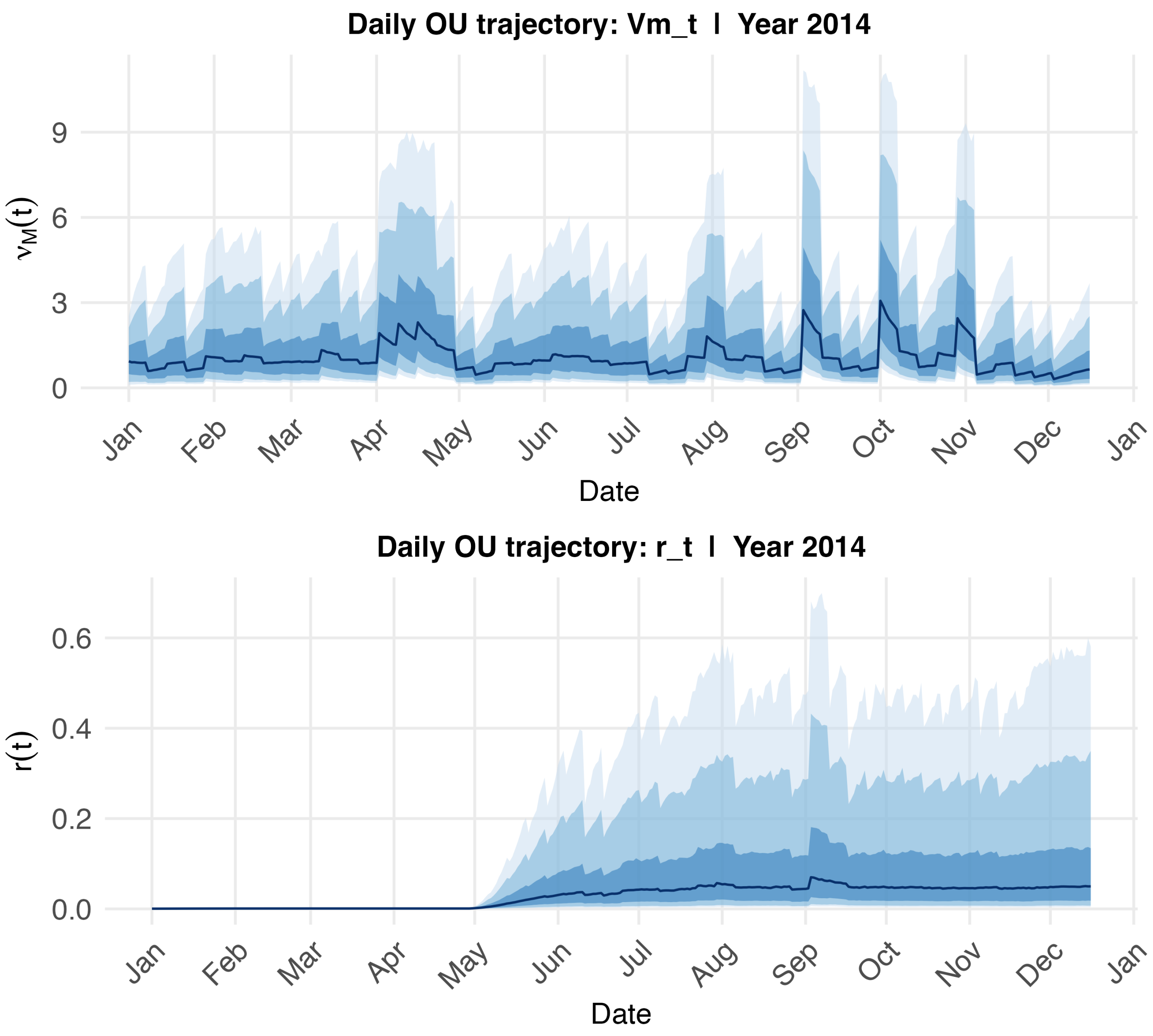

**Figure S54:** Daily ensemble trajectories of time-varying parameters estimated by the Ornstein-Uhlenbeck (OU) process during model fitting and forecasting, 2014. Upper panel: daily ensemble trajectories of the mosquito growth rate parameter ($\nu_M$), shown as the ensemble mean (solid line) with the interquartile range (shaded ribbon) across all N = 8,000 ensemble members. Lower panel: daily ensemble trajectories of the WNV transmission rate parameter ($r_t$). In both panels, the x-axis spans the full annual modeling period from January 1 to the end of the fitting window. Parameter values are propagated forward in daily steps via the OU process, seven successive daily steps per assimilation week. Wider ribbons indicate periods of greater ensemble spread, reflecting higher parameter uncertainty. Results are shown for the Full Model applied to Maricopa County, Arizona. To access other model configurations and other years, visit the link: https://github.com/kayodeoshinubi/WNV-Forecasting

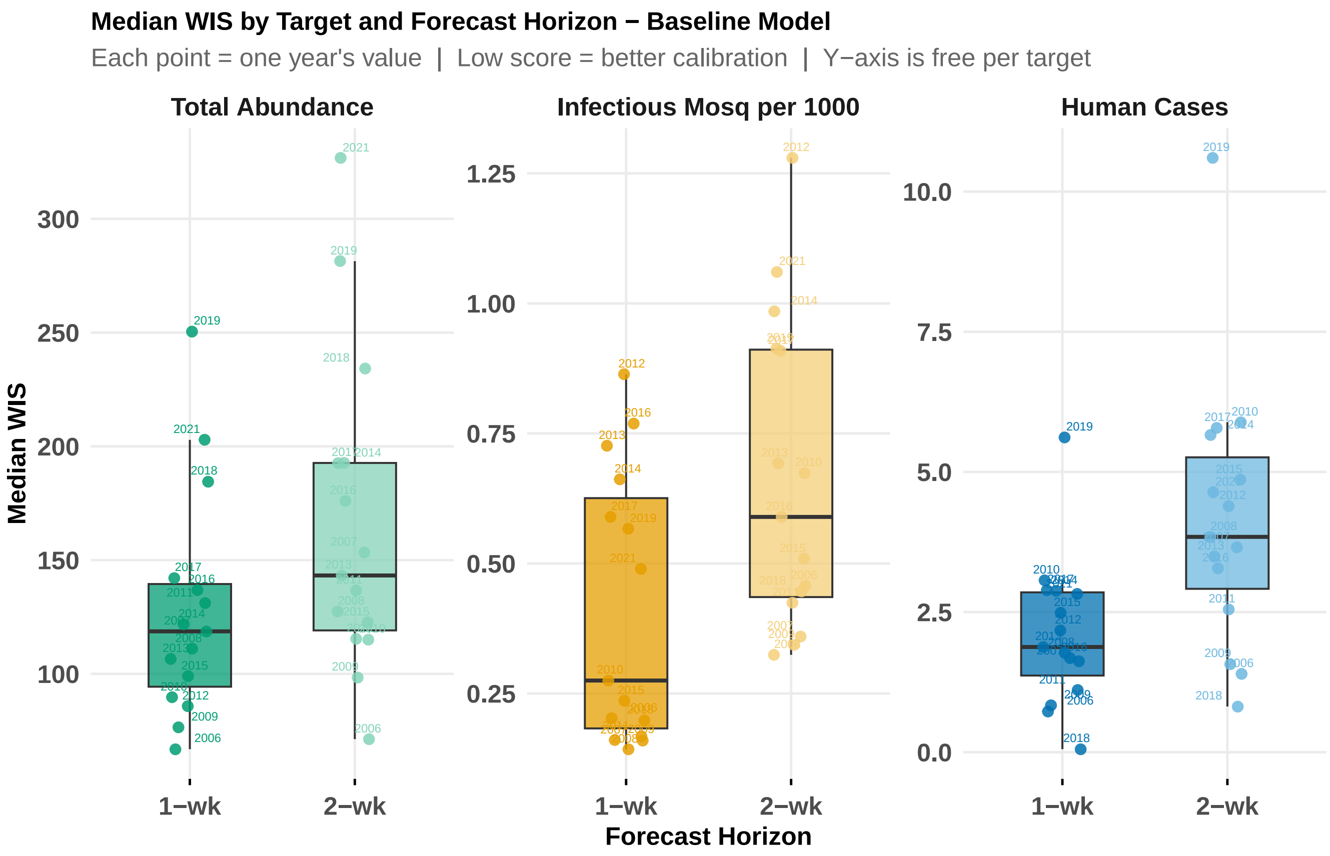

**Figure S55:** Median WIS for 1-week-ahead and 2-week-ahead probabilistic forecasts for the baseline model aggregated across all assimilation weeks per year, shown separately for human WNV cases (right), mosquito infection prevalence (IM1000) (center), and total mosquito abundance (left). Each point represents the median WIS for a single year; lower values indicate better fit accuracy.

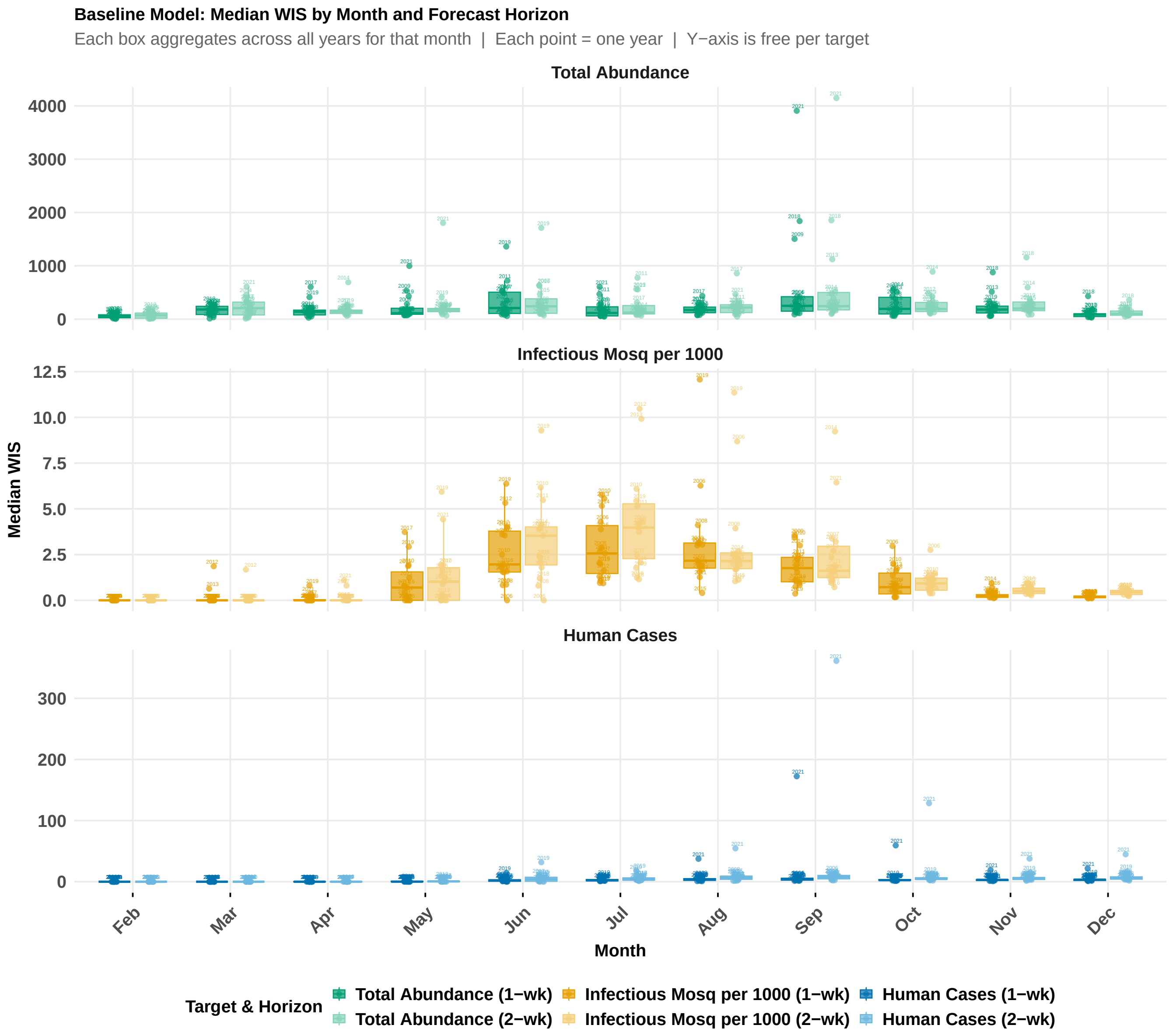

**Figure S56:** Seasonal patterns in 1-week-ahead and 2-week-ahead forecast accuracy for baseline model and surveillance targets, aggregated over the 15-year study period (2006–2019, 2021). Each boxplot summarizes the distribution of per-year median relative weighted interval score (WIS) for a given month and target, where each box aggregates across all available years for that calendar month. Results are shown for human WNV cases (bottom), mosquito infection prevalence (IM1000) (middle), and total mosquito abundance (top). January is excluded, as forecasts were not generated for that month.

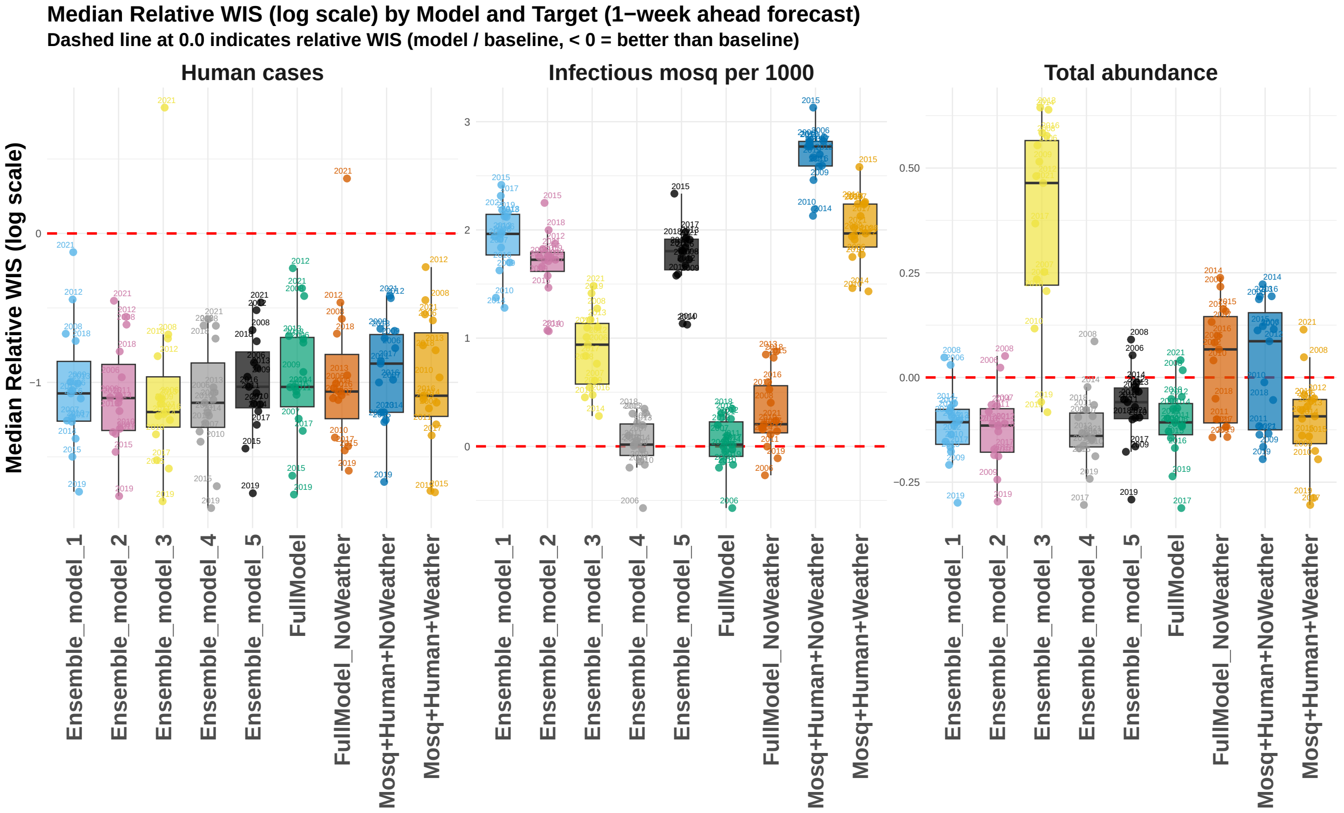

**Figure S57:** Median relative WIS on the log scale for 1-week-ahead probabilistic forecasts, where values below the red dashed line (0.0) indicate that the model outperforms the historical baseline. Each point represents the median relative WIS for a single year.

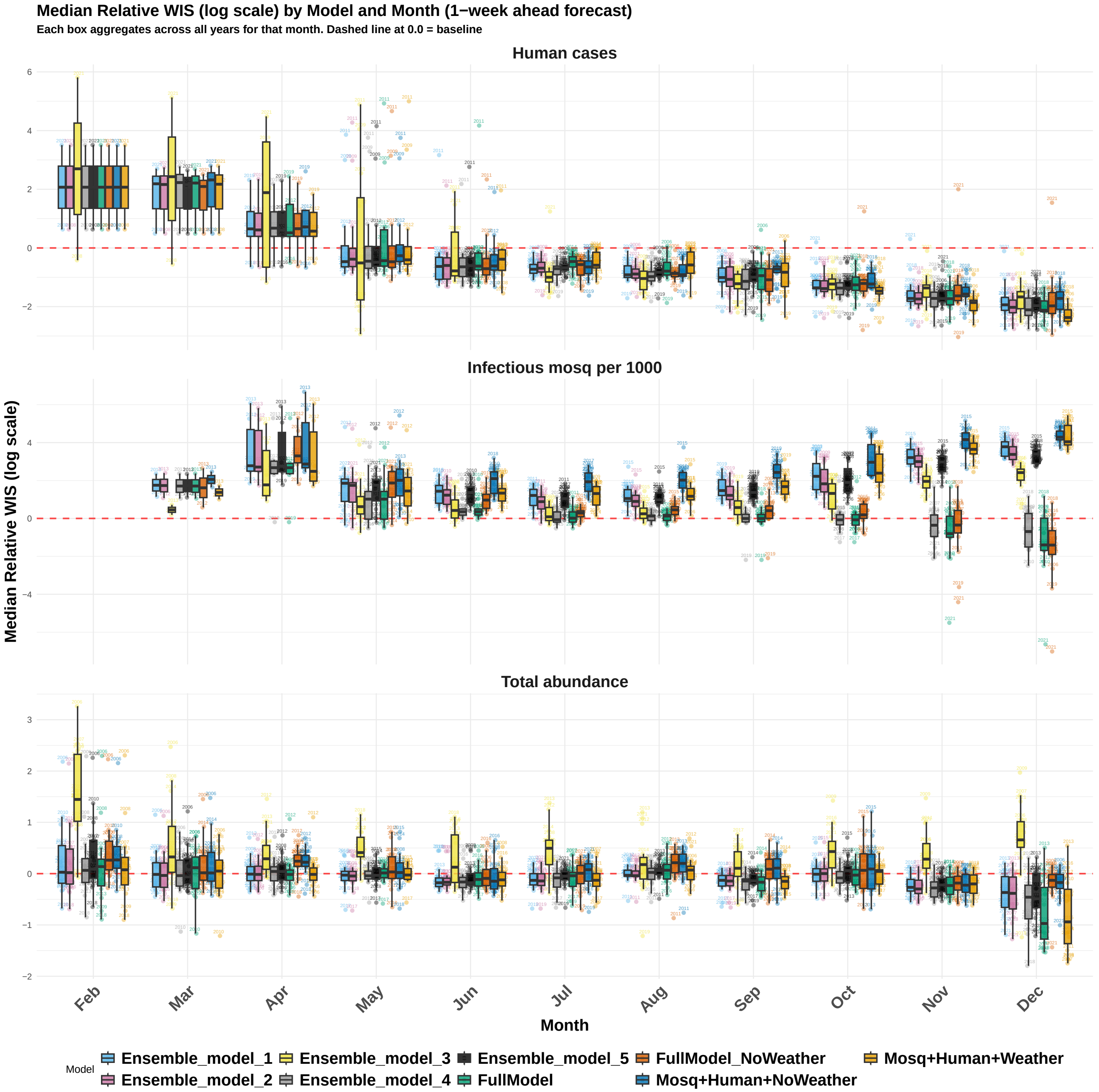

**Figure S58:** Seasonal patterns in 1-week-ahead forecast accuracy across model configurations and surveillance targets, aggregated over the 15-year study period (2006–2019, 2021). Each boxplot summarizes the distribution of per-year median relative weighted interval score (WIS) on the log scale for a given model, month, and target, where each box aggregates across all available years for that calendar month. The red dashed line at 0.0 marks the boundary between better-than-baseline (below) and worse-than-baseline (above) performance. Results are shown for human WNV cases (top), imosquito infection prevalence (IM1000) (middle), and total mosquito abundance (bottom). January is excluded, as forecasts were not generated for that month.

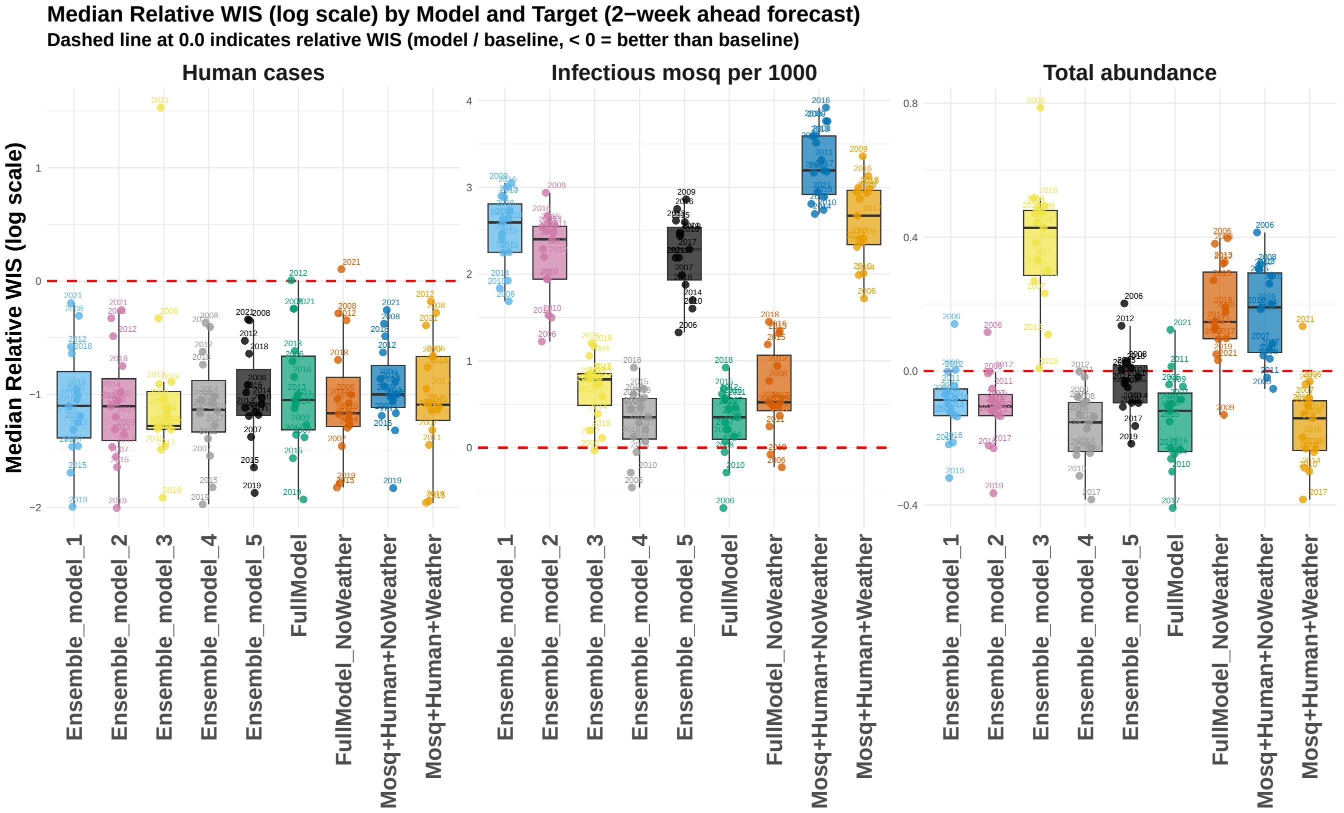

**Figure S59:** Median relative WIS on the log scale for 2-week-ahead probabilistic forecasts, where values below the red dashed line (0.0) indicate that the model outperforms the historical baseline. Each point represents the median relative WIS for a single year.

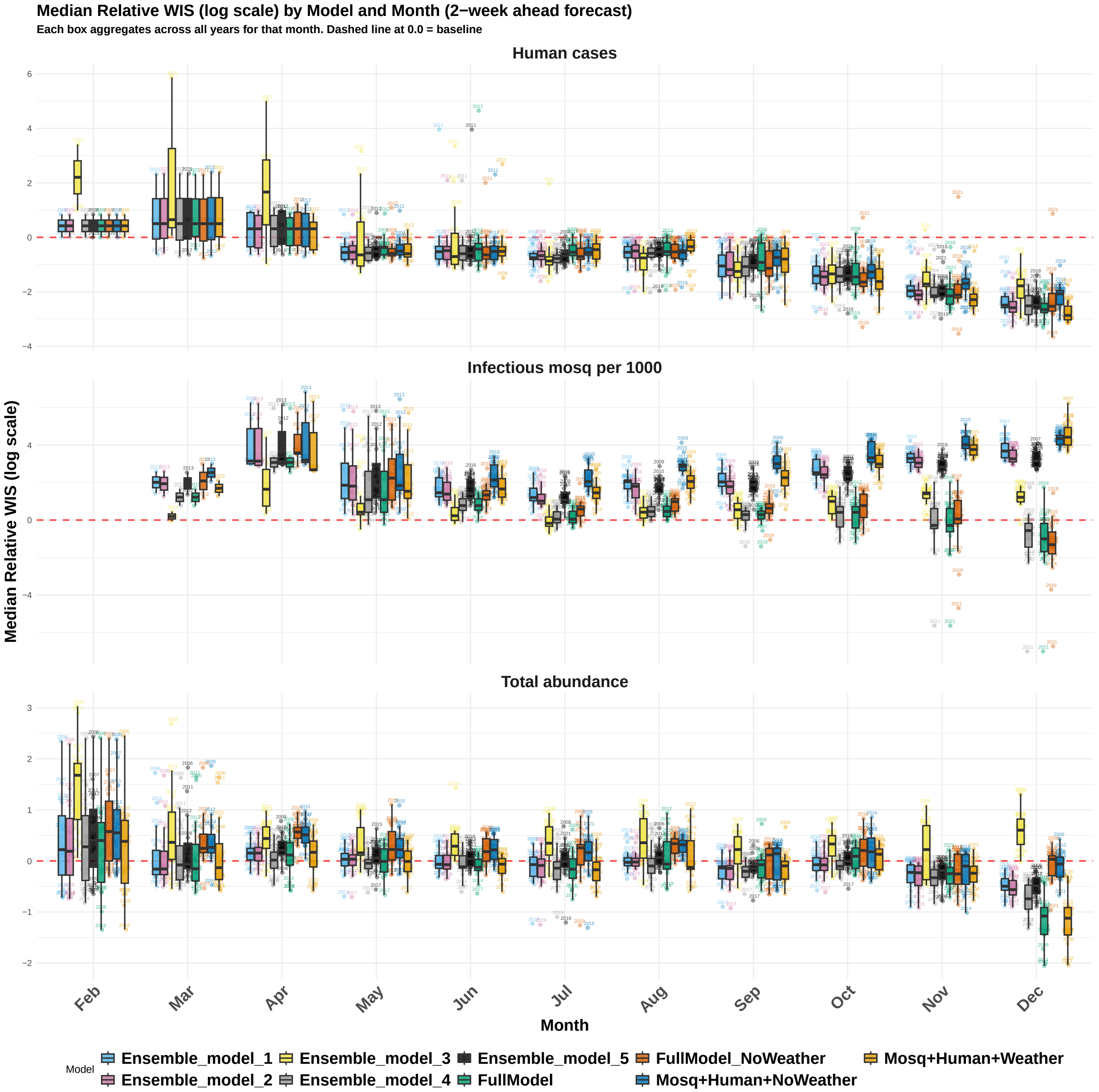

**Figure S60:** Seasonal patterns in 2-week-ahead forecast accuracy across model configurations and surveillance targets, aggregated over the 15-year study period (2006–2019, 2021). Each boxplot summarizes the distribution of per-year median relative weighted interval score (WIS) on the log scale for a given model, month, and target, where each box aggregates across all available years for that calendar month. The red dashed line at 0.0 marks the boundary between better-than-baseline (below) and worse-than-baseline (above) performance. Results are shown for human WNV cases (top), mosquito infection prevalence (IM1000) (middle), and total mosquito abundance (bottom). January is excluded as forecasts were not generated for that month.

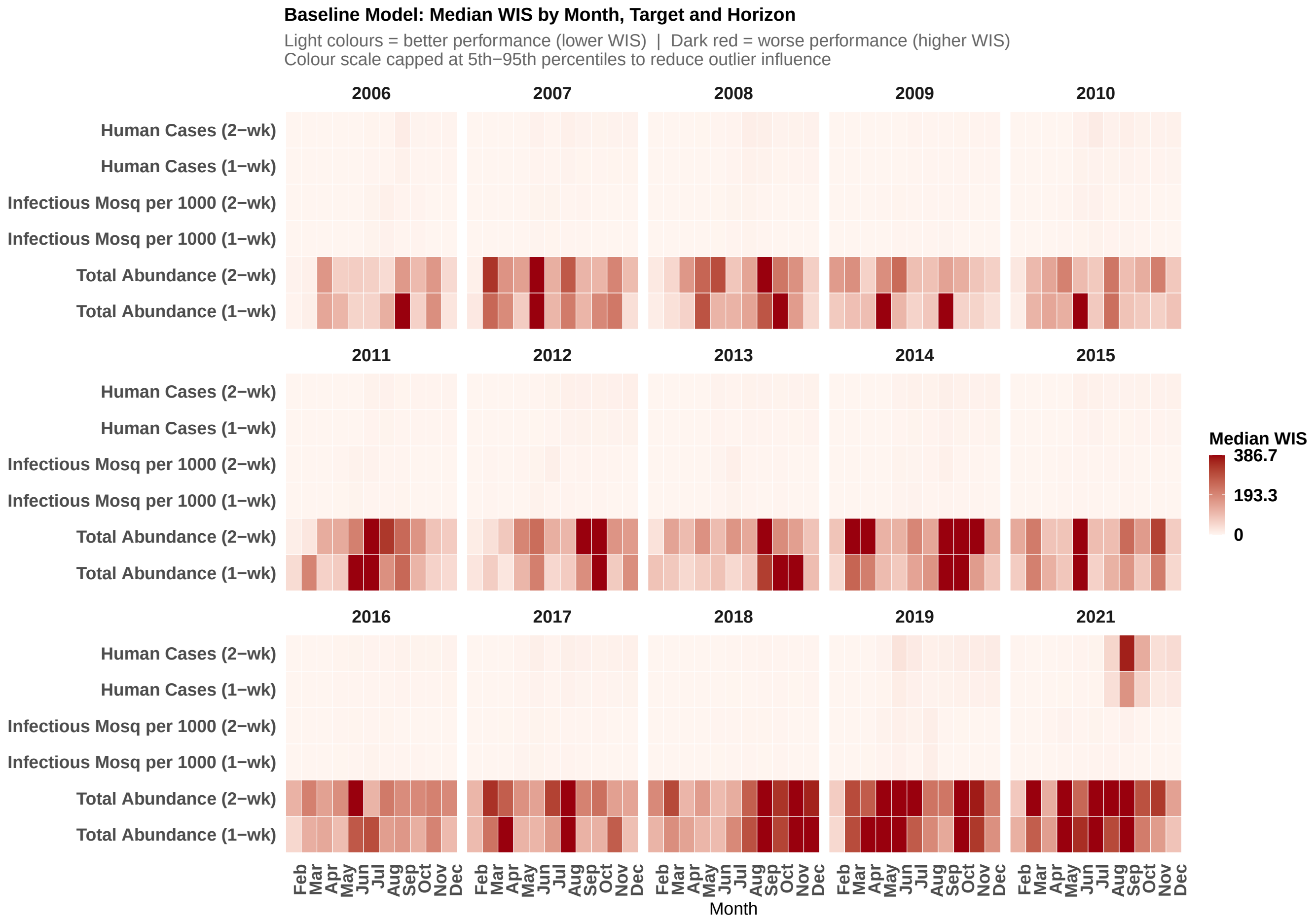

**Figure S61:** Median WIS for 1-week-ahead and 2-week-ahead probabilistic forecasts for the baseline model by calendar month and surveillance target. Each cell represents the median WIS aggregated across all forecast weeks falling within that calendar month. Darker red cells indicate higher (worse) WIS values; lighter cells indicate lower (better) WIS values. The color scale is calibrated independently using the 5th and 95th percentiles of the median WIS distribution across all model–month–year combinations for that target, rather than the global minimum and maximum, so that the color gradient reflects the typical range of forecast error while preventing extreme outlier values from compressing the scale and obscuring variation in the bulk of the distribution. Cells with values below the 5th percentile are rendered at the minimum color, and cells above the 95th percentile are rendered at the maximum color.

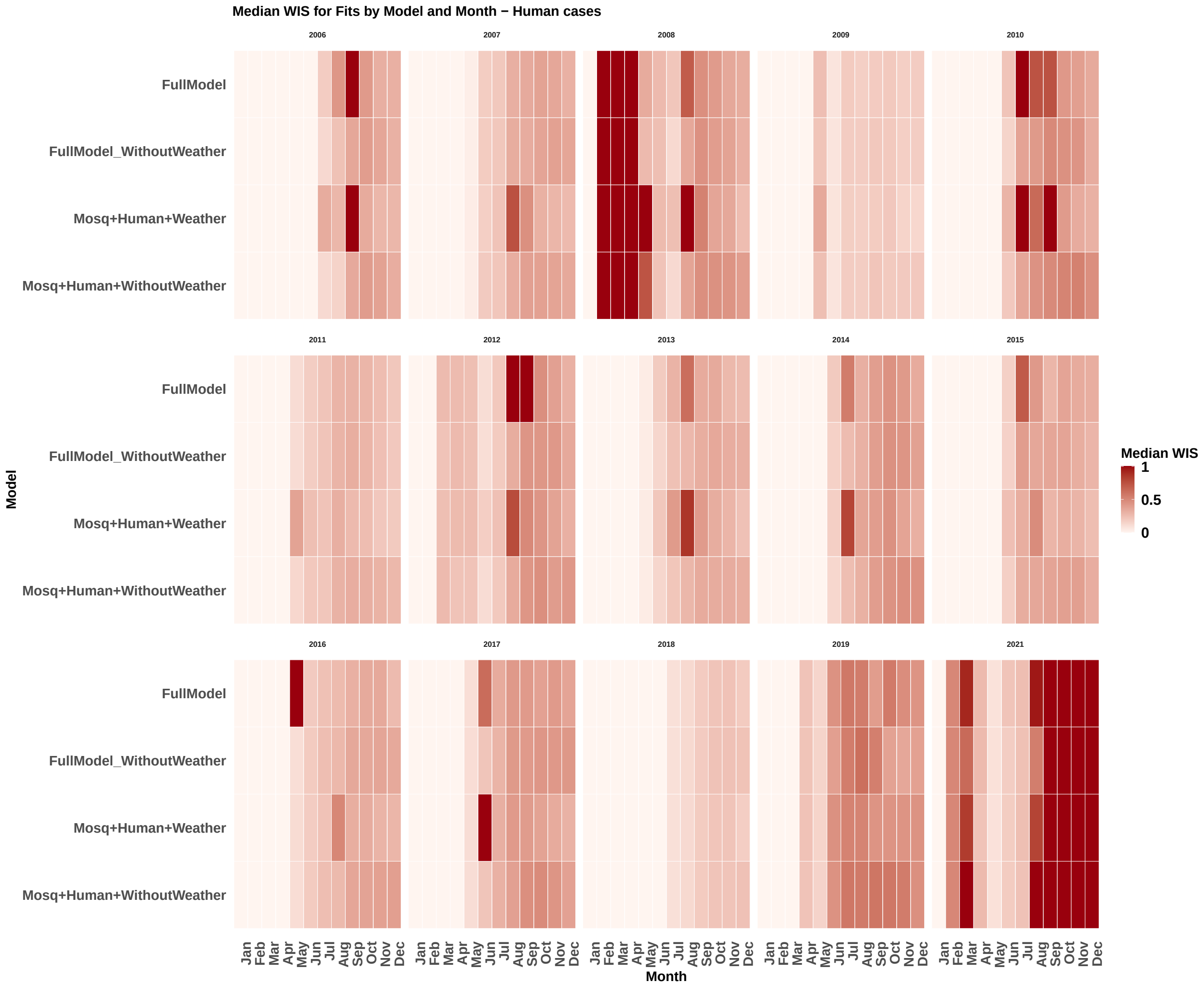

**Figure S62:** Median weighted interval score (WIS) for in-sample model fits by model, month, and year for human cases, Maricopa County, Arizona (2006–2019, 2021). Each cell represents the median WIS computed across all weekly forecasts within a given calendar month for a specific model and year. Darker red cells indicate higher (worse) WIS values; lighter cells indicate lower (better) WIS values. The color scale is calibrated independently using the 5th and 95th percentiles of the median WIS distribution across all model–month–year combinations for that target, rather than the global minimum and maximum, so that the color gradient reflects the typical range of forecast error while preventing extreme outlier values from compressing the scale and obscuring variation in the bulk of the distribution. Cells with values below the 5th percentile are rendered at the minimum color, and cells above the 95th percentile are rendered at the maximum color.

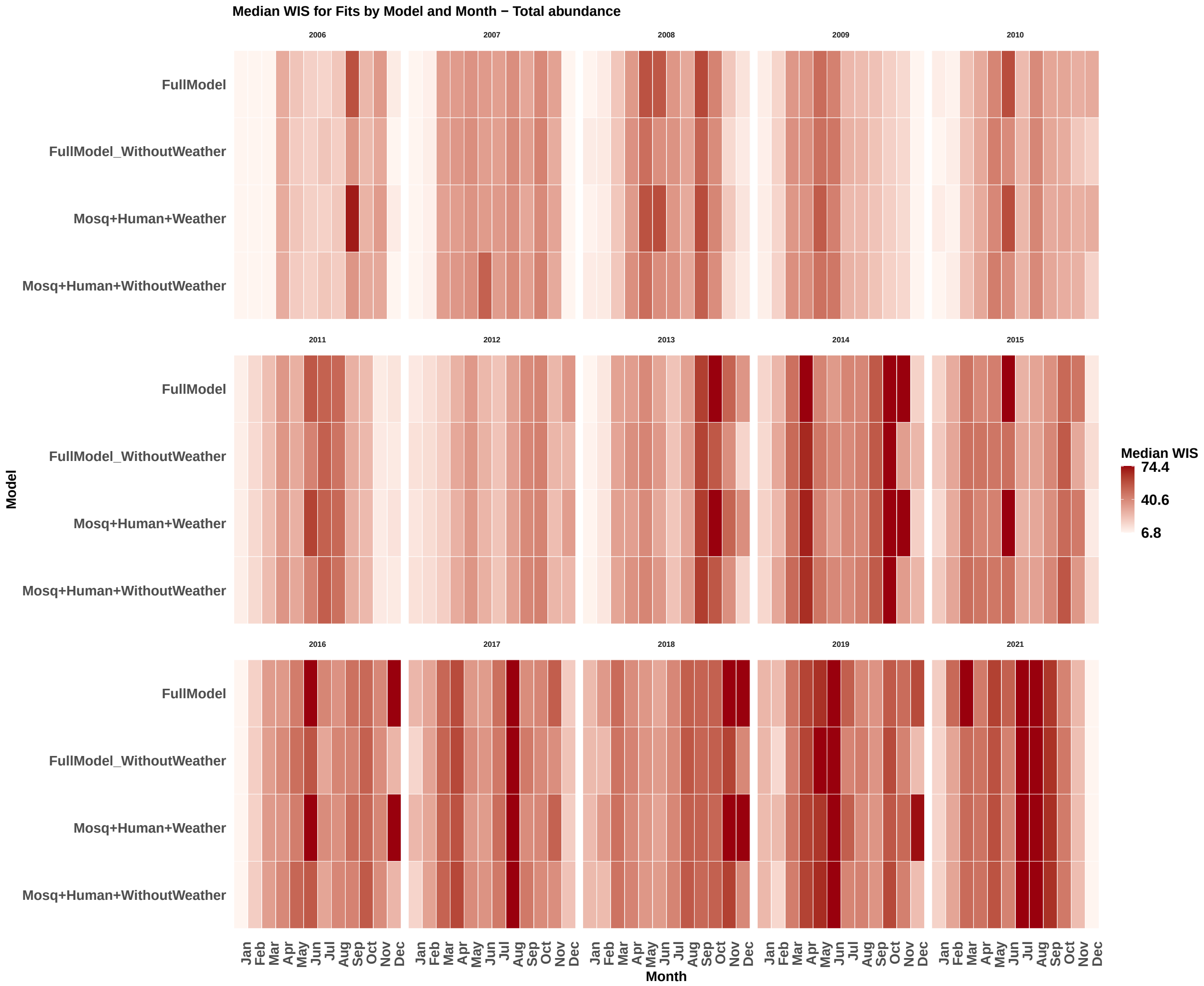

**Figure S63:** Median weighted interval score (WIS) for in-sample model fits by model, month, and year for total mosquito abundance, Maricopa County, Arizona (2006–2019, 2021). Each cell represents the median WIS computed across all weekly forecasts within a given calendar month for a specific model and year. Darker red cells indicate higher (worse) WIS values; lighter cells indicate lower (better) WIS values. The color scale is calibrated independently using the 5th and 95th percentiles of the median WIS distribution across all model–month–year combinations for that target, rather than the global minimum and maximum, so that the color gradient reflects the typical range of forecast error while preventing extreme outlier values from compressing the scale and obscuring variation in the bulk of the distribution. Cells with values below the 5th percentile are rendered at the minimum color, and cells above the 95th percentile are rendered at the maximum color.

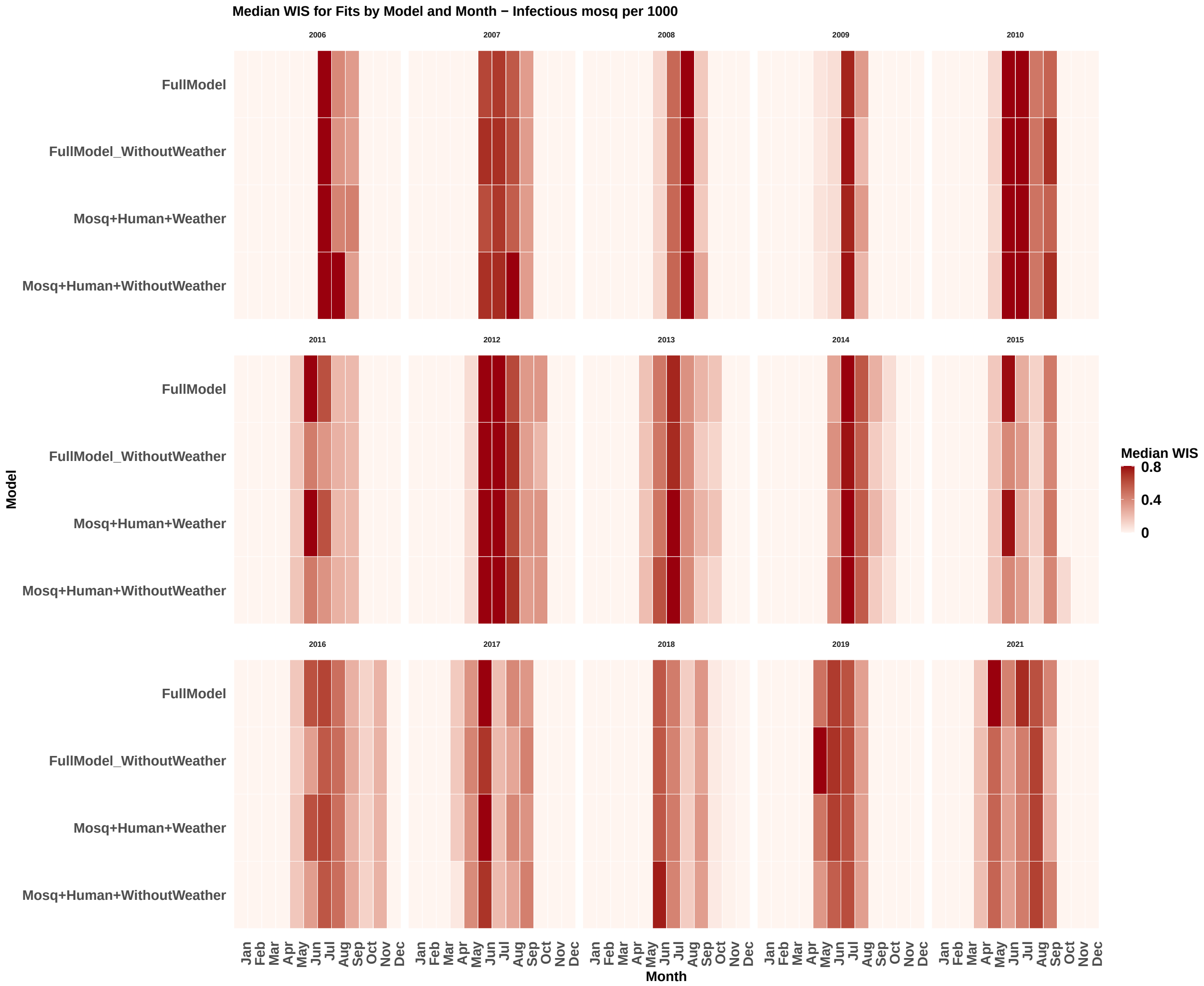

**Figure S64:** Median weighted interval score (WIS) for in-sample model fits by model, month, and year for mosquito infection prevalence (IM1000), Maricopa County, Arizona (2006–2019, 2021). Each cell represents the median WIS computed across all weekly forecasts within a given calendar month for a specific model and year. Darker red cells indicate higher (worse) WIS values; lighter cells indicate lower (better) WIS values. The color scale is calibrated independently using the 5th and 95th percentiles of the median WIS distribution across all model–month–year combinations for that target, rather than the global minimum and maximum, so that the color gradient reflects the typical range of forecast error while preventing extreme outlier values from compressing the scale and obscuring variation in the bulk of the distribution. Cells with values below the 5th percentile are rendered at the minimum color, and cells above the 95th percentile are rendered at the maximum color.

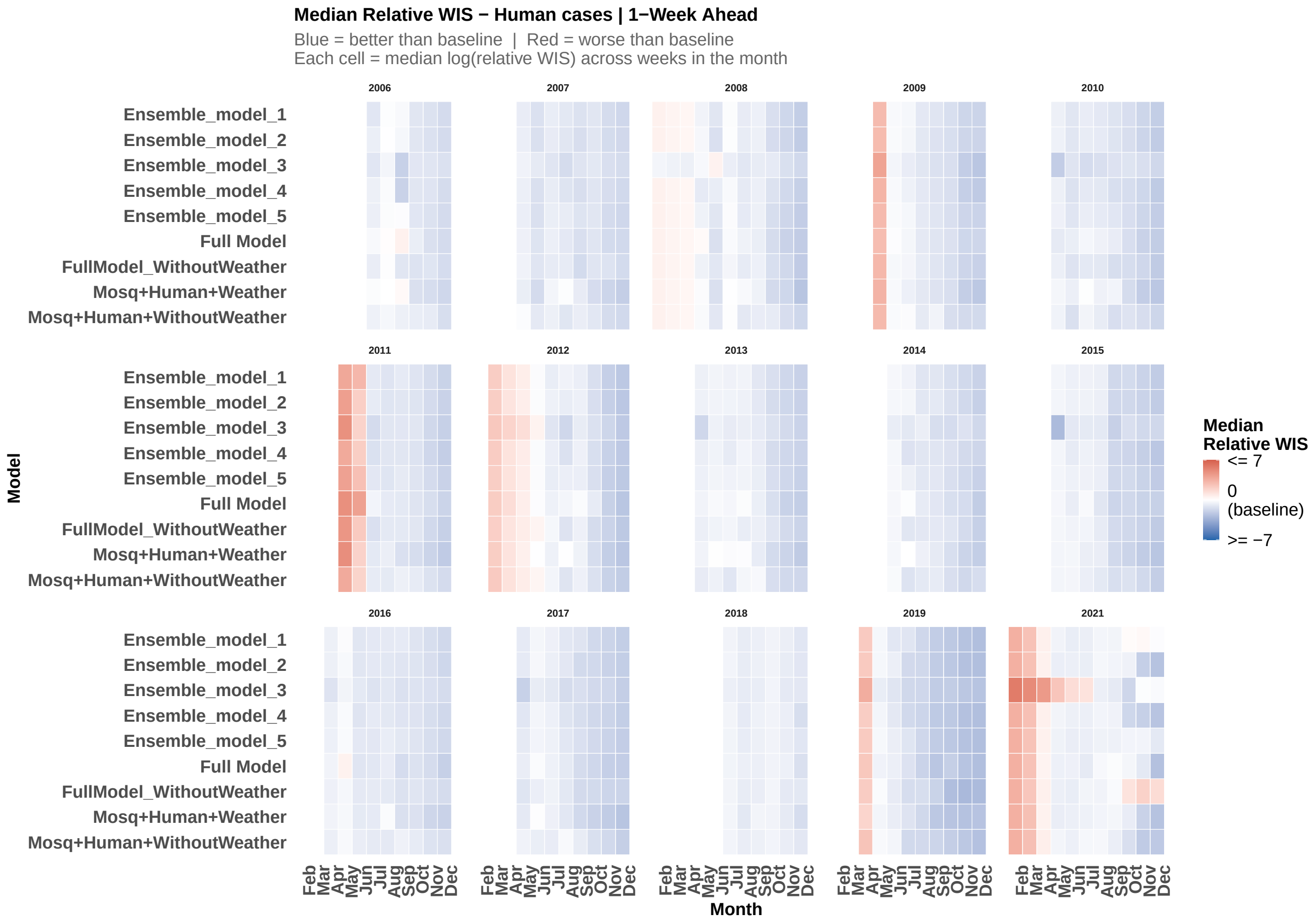

**Figure S65:** Median relative Weighted Interval Score (WIS) for human cases on the log scale for 1-week-ahead probabilistic forecasts by model configuration and calendar month. Each cell represents the median relative WIS aggregated across all forecast weeks falling within that calendar month, where relative WIS is defined as the log ratio of the model WIS to the historical baseline WIS. Values below zero (blue) indicate that the model outperformed the historical baseline; values above zero (red) indicate worse-than-baseline performance. Results are shown for all model configurations.

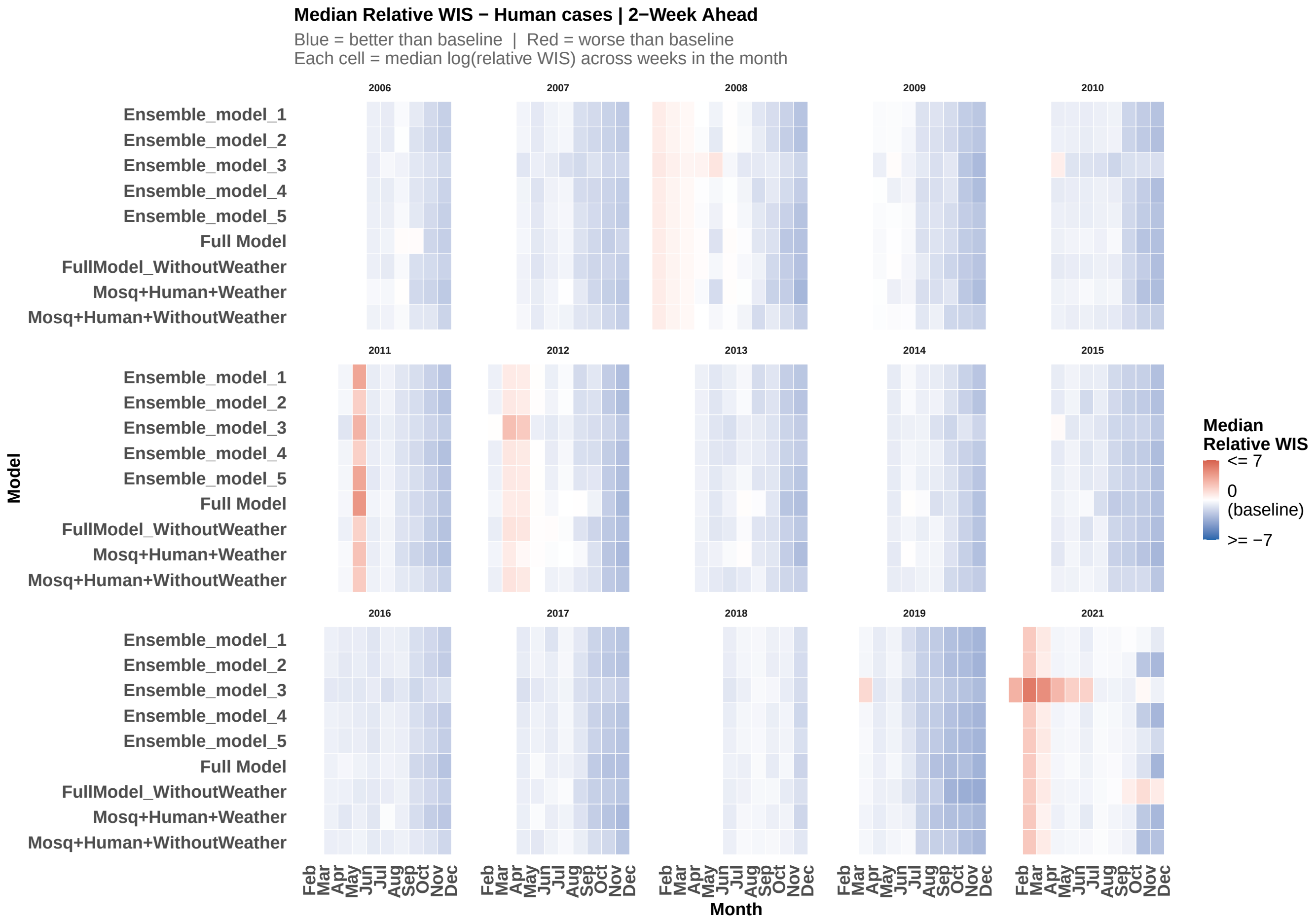

**Figure S66:** Median relative Weighted Interval Score (WIS) for human cases on the log scale for 2-week-ahead probabilistic forecasts by model configuration and calendar month. Each cell represents the median relative WIS aggregated across all forecast weeks falling within that calendar month, where relative WIS is defined as the log ratio of the model WIS to the historical baseline WIS. Values below zero (blue) indicate that the model outperformed the historical baseline; values above zero (red) indicate worse-than-baseline performance. Results are shown for all model configurations.

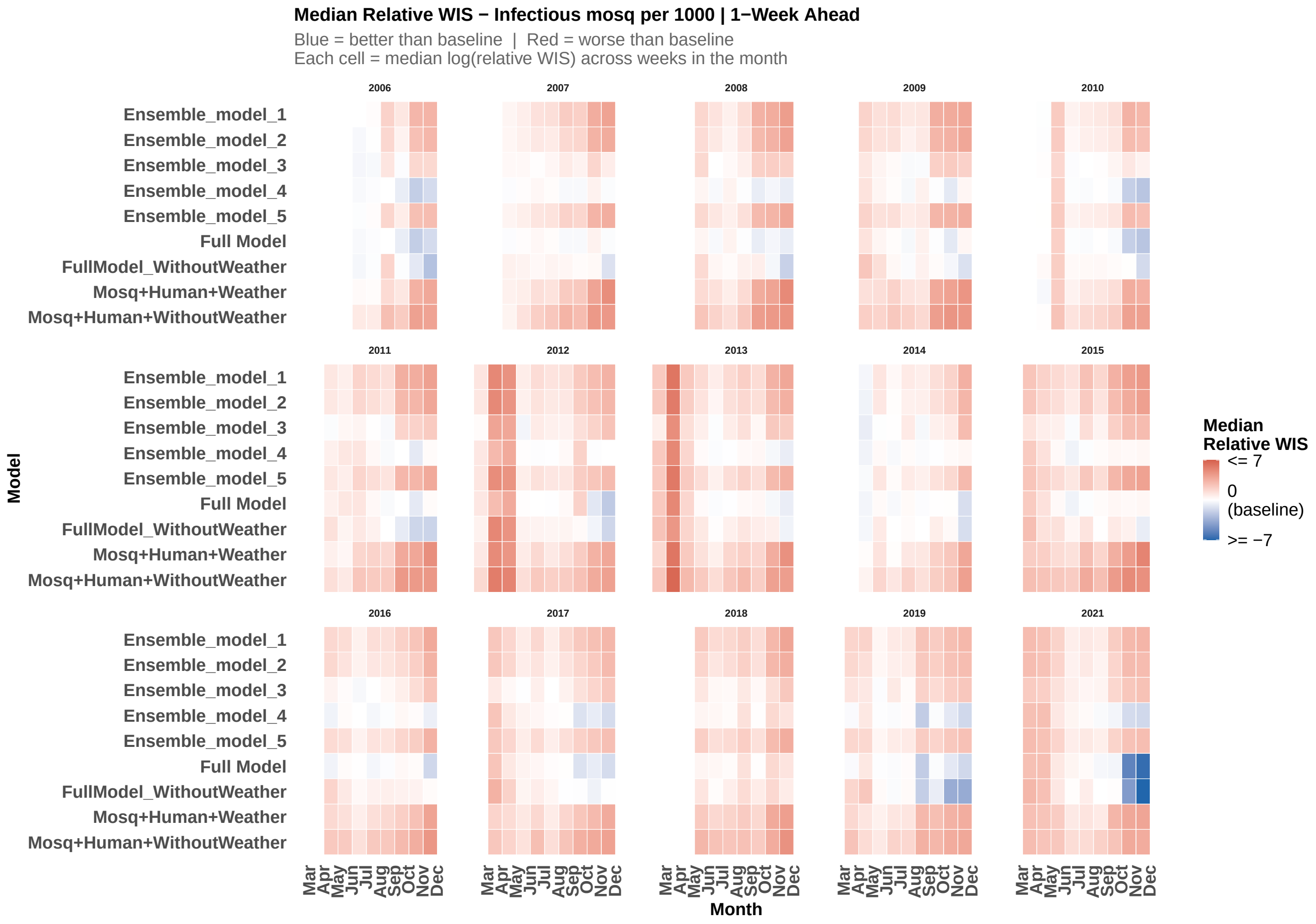

**Figure S67:** Median relative Weighted Interval Score (WIS) for mosquito infection prevalence (IM1000) on the log scale for 1-week-ahead probabilistic forecasts by model configuration and calendar month. Each cell represents the median relative WIS aggregated across all forecast weeks falling within that calendar month, where relative WIS is defined as the log ratio of the model WIS to the historical baseline WIS. Values below zero (blue) indicate that the model outperformed the historical baseline; values above zero (red) indicate worse-than-baseline performance. Results are shown for all model configurations.

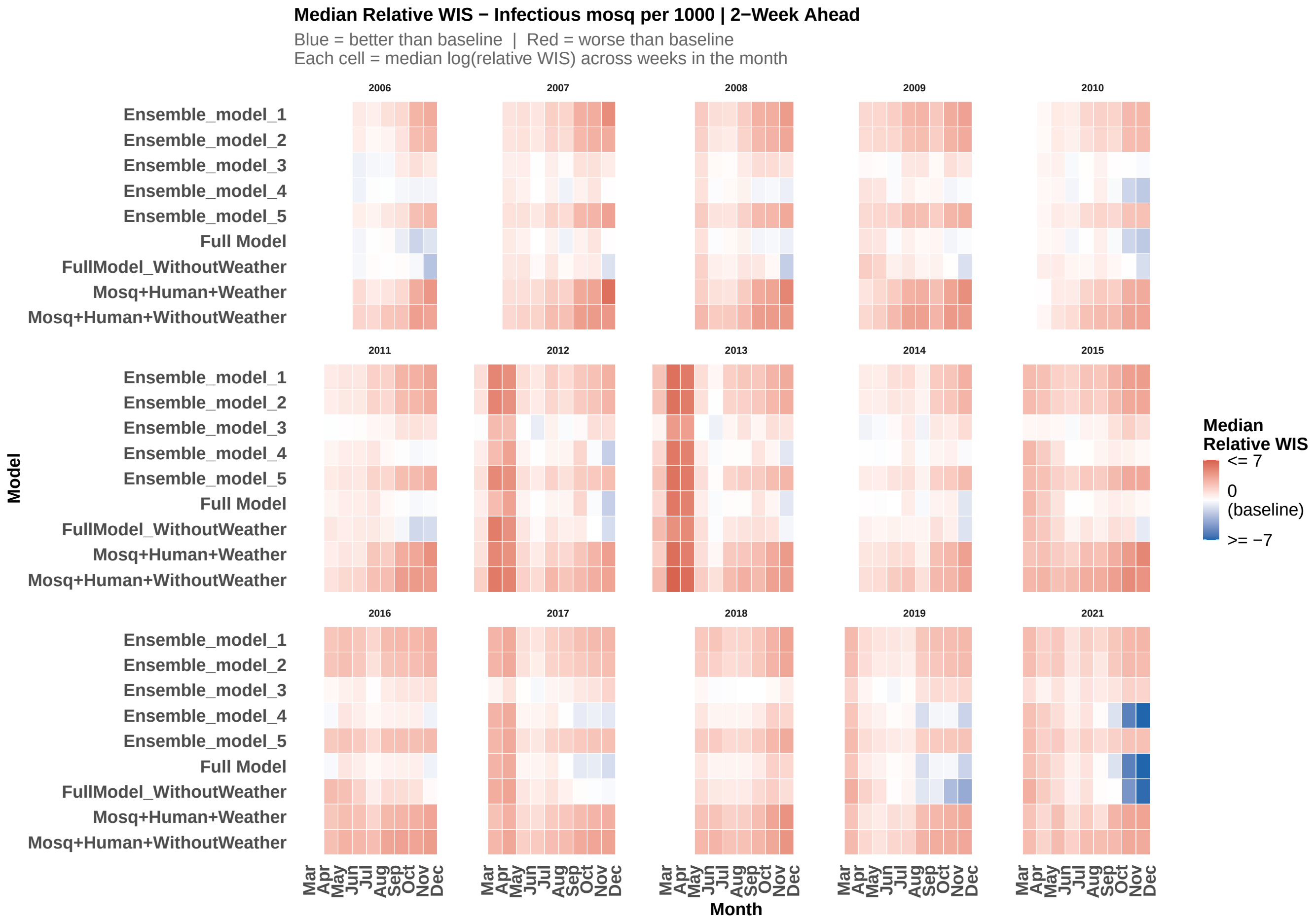

**Figure S68:** Median relative Weighted Interval Score (WIS) for mosquito infection prevalence (IM1000) on the log scale for 2-week-ahead probabilistic forecasts by model configuration and calendar month. Each cell represents the median relative WIS aggregated across all forecast weeks falling within that calendar month, where relative WIS is defined as the log ratio of the model WIS to the historical baseline WIS. Values below zero (blue) indicate that the model outperformed the historical baseline; values above zero (red) indicate worse-than-baseline performance. Results are shown for all model configurations.

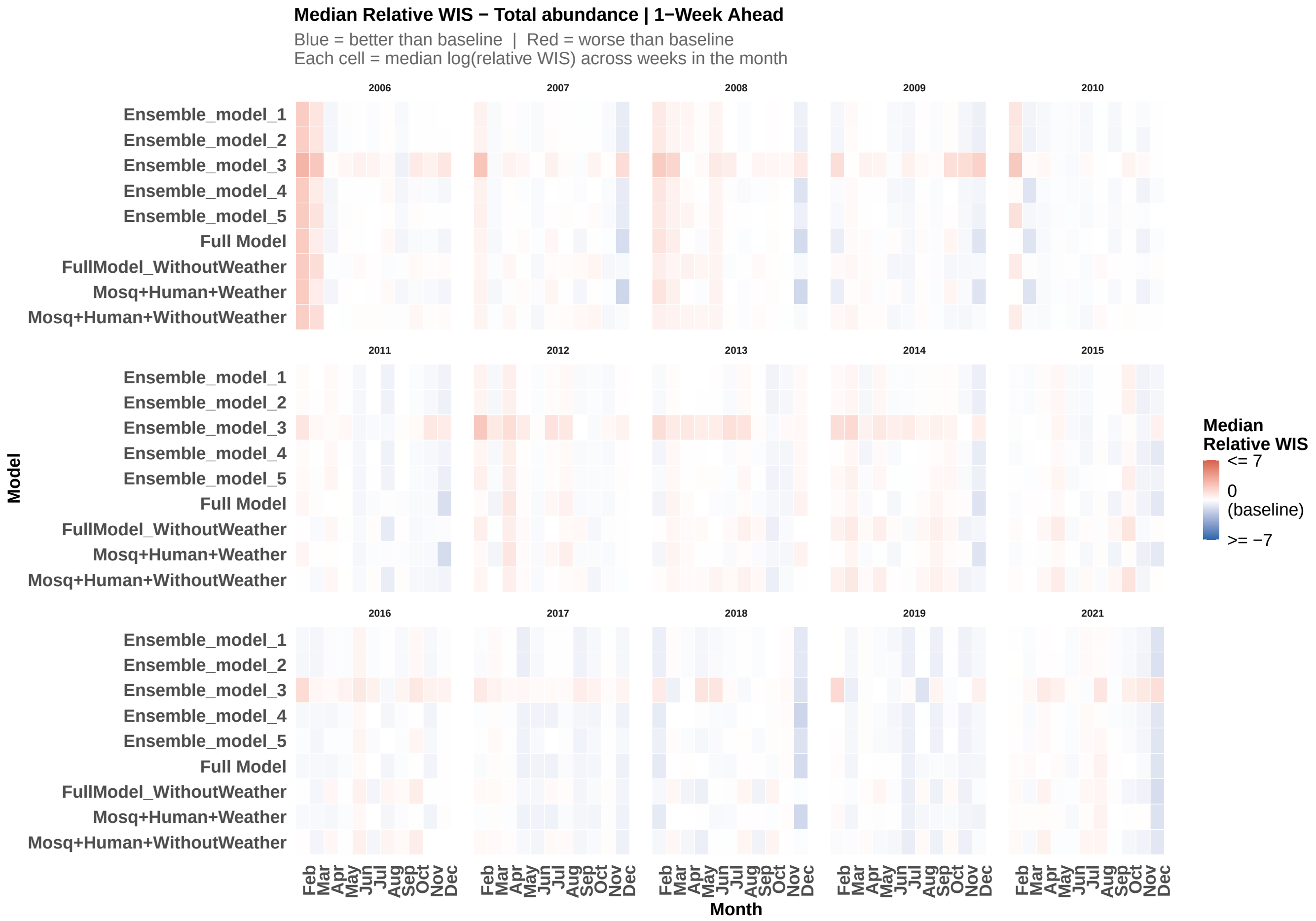

**Figure S69:** Median relative Weighted Interval Score (WIS) for total abundance on the log scale for 1-week-ahead probabilistic forecasts by model configuration and calendar month. Each cell represents the median relative WIS aggregated across all forecast weeks falling within that calendar month, where relative WIS is defined as the log ratio of the model WIS to the historical baseline WIS. Values below zero (blue) indicate that the model outperformed the historical baseline; values above zero (red) indicate worse-than-baseline performance. Results are shown for all model configurations.

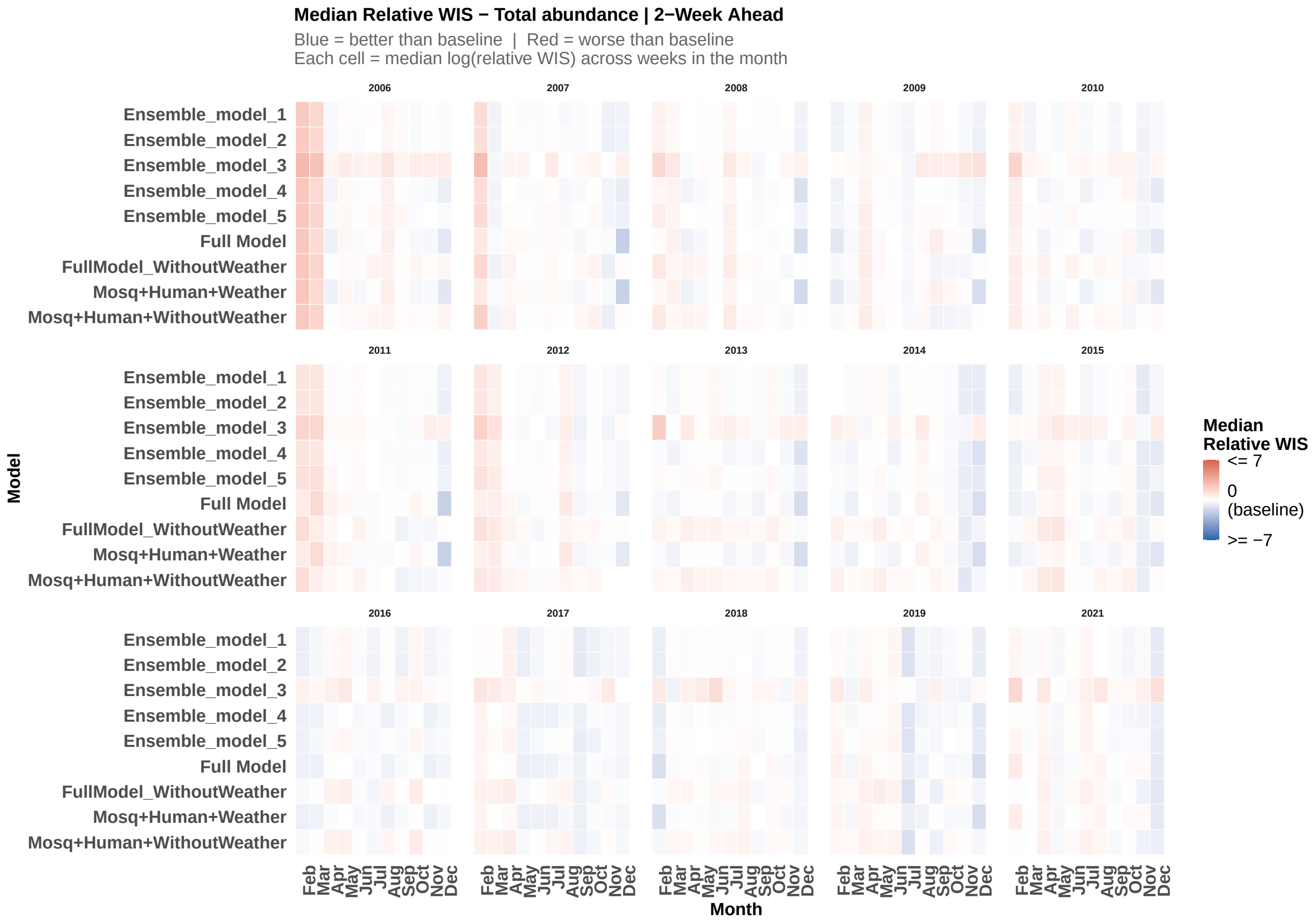

**Figure S70:** Median relative Weighted Interval Score (WIS) for total abundance on the log scale for 2-week-ahead probabilistic forecasts by model configuration and calendar month. Each cell represents the median relative WIS aggregated across all forecast weeks falling within that calendar month, where relative WIS is defined as the log ratio of the model WIS to the historical baseline WIS. Values below zero (blue) indicate that the model outperformed the historical baseline; values above zero (red) indicate worse-than-baseline performance. Results are shown for all model configurations.

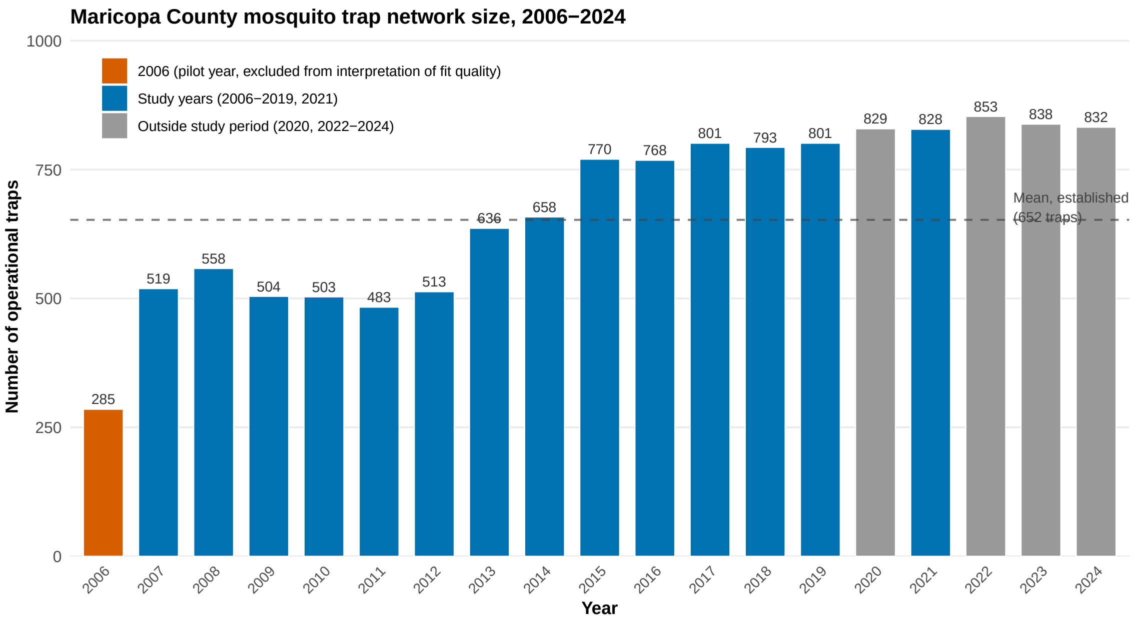

**Figure S71:** Number of operational mosquito traps in the Maricopa County surveillance network, 2006–2024. 2006, the pilot year of the trapping program, is shown in vermillion and had substantially fewer traps ($n = 285$) than all subsequent years. Study years (2006–2019, 2021) are shown in blue; years outside the study period (2020, 2022–2024, excluded due to COVID-19 surveillance disruptions or falling after the study window) are shown in grey for context. The dashed line marks the mean trap count across established study years (2007–2019, 2021; $n \approx 660$). Trap network size stabilized above 500 traps from 2009 onward and exceeded 750 traps from 2015 onward.

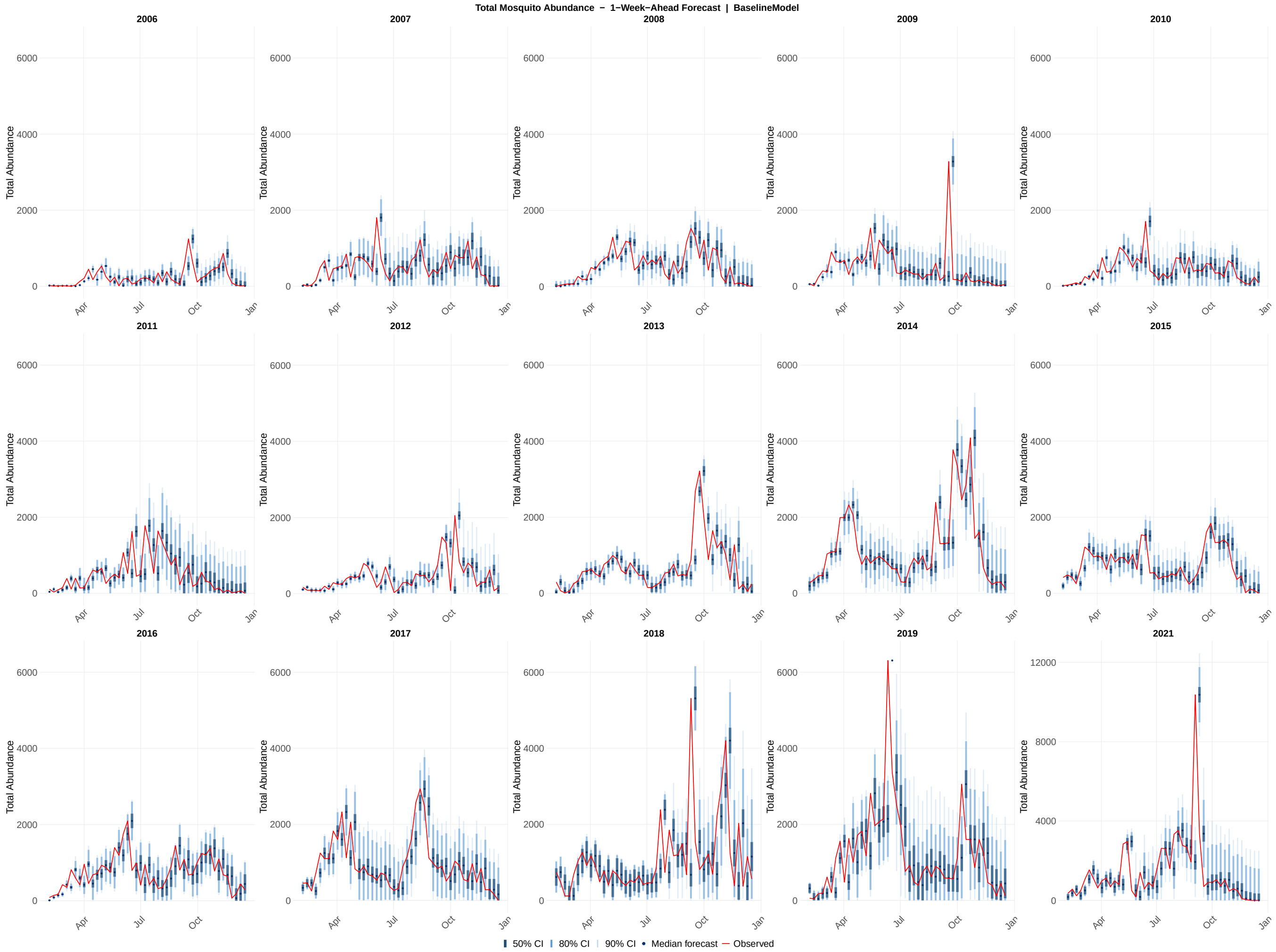

**Figure S72:** Probabilistic 1-week-ahead forecasts of total abundance across all 15 study years (2006–2019, 2021) for the baseline model. Each panel shows the sequence of 1-week-ahead forecast distributions generated at each of the 46 assimilation steps within a single calendar year. Shaded ribbons indicate the 50% (darkest), 80% (medium), and 90% (lightest) prediction intervals; the solid line shows the forecast median; the red line shows the observed total abundance values. The y-axis scale for 2021 is unconstrained; all other years share a common axis scale.

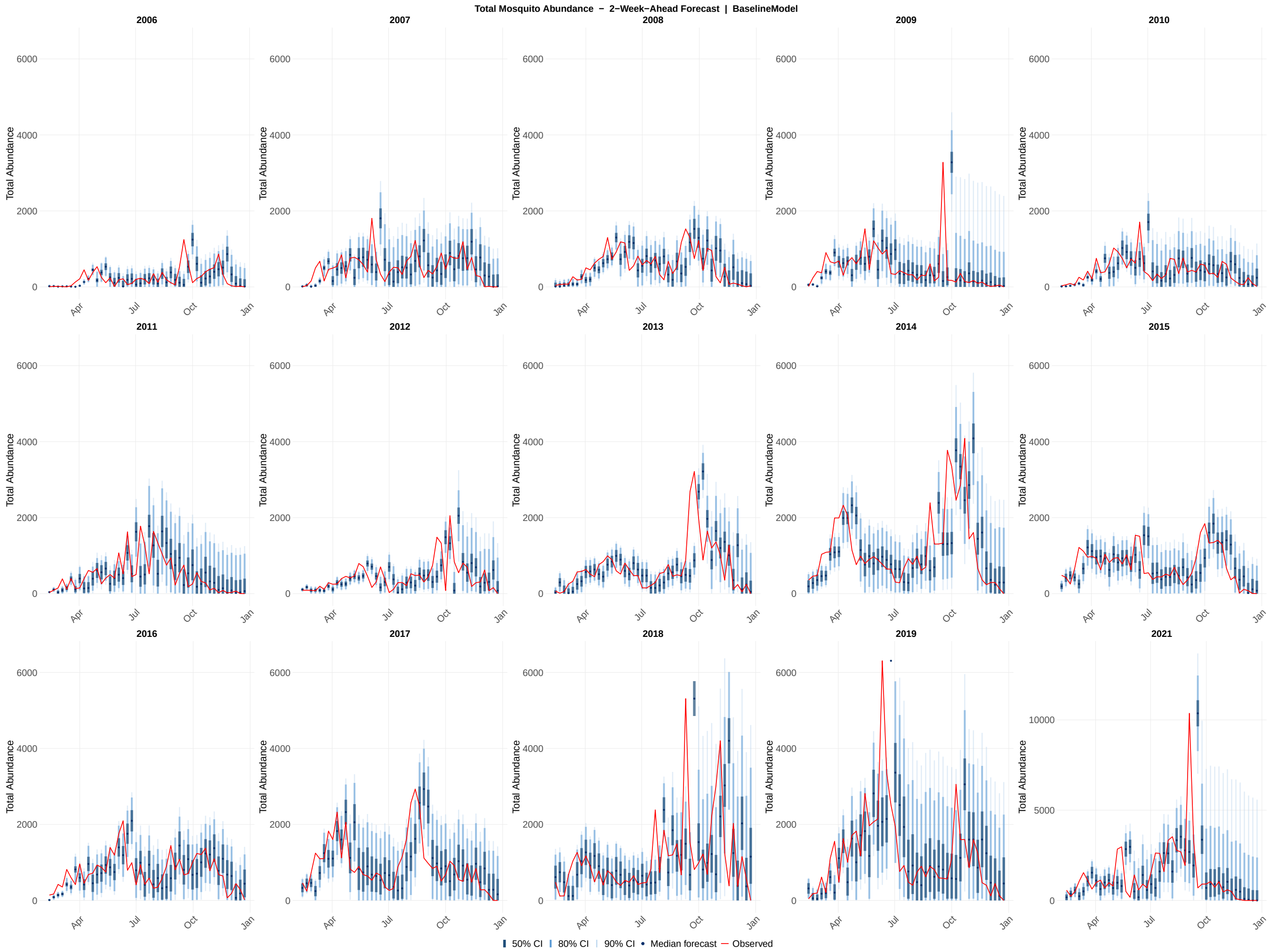

**Figure S73:** Probabilistic 2-week-ahead forecasts of total abundance across all 15 study years (2006–2019, 2021) for the baseline model. Each panel shows the sequence of 2-week-ahead forecast distributions generated at each of the 46 assimilation steps within a single calendar year. Shaded ribbons indicate the 50% (darkest), 80% (medium), and 90% (lightest) prediction intervals; the solid line shows the forecast median; the red line shows the observed total abundance values. The y-axis scale for 2021 is unconstrained; all other years share a common axis scale.

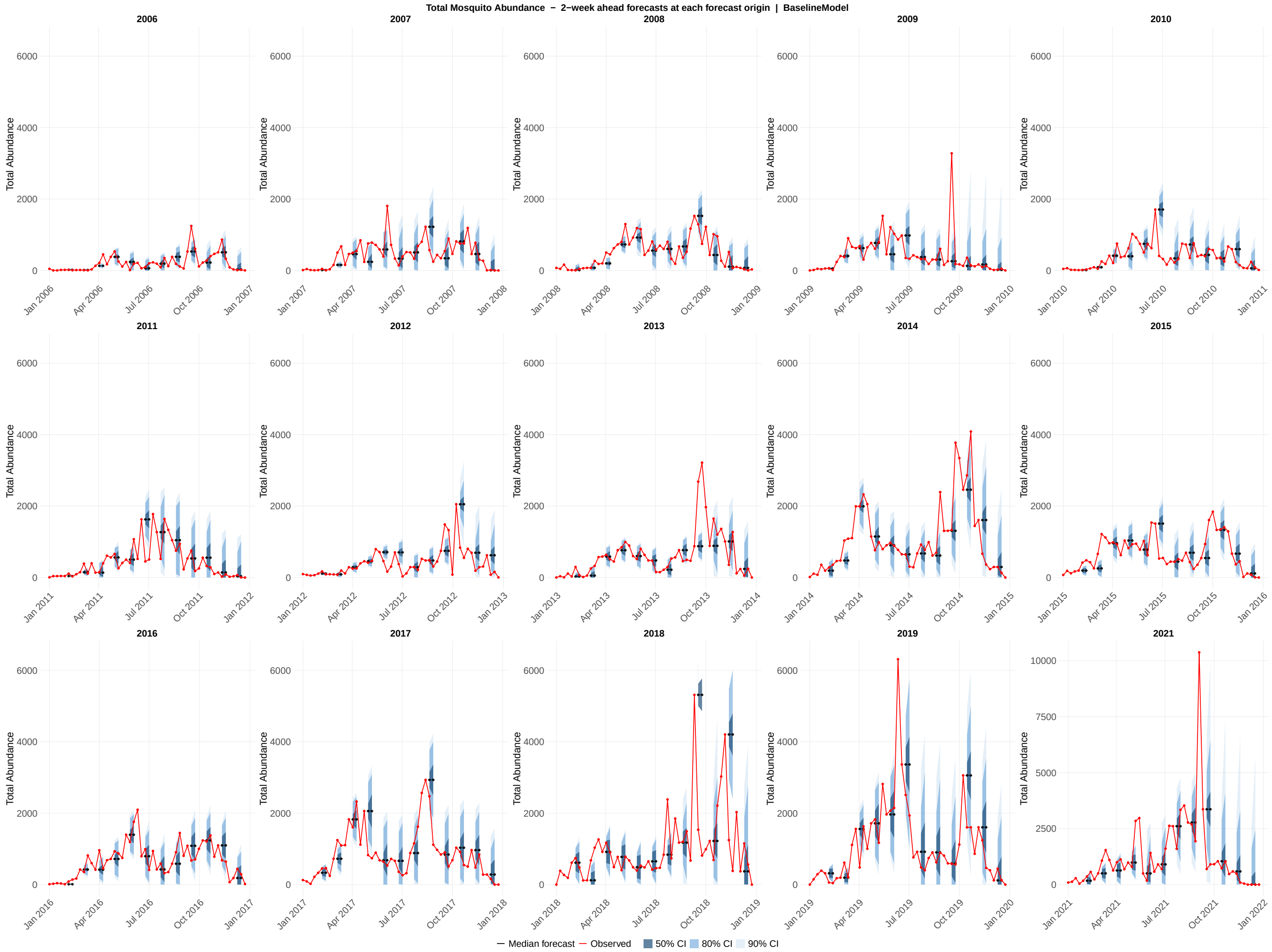

**Figure S74:** Probabilistic forecasts of total abundance across all 15 study years (2006–2019, 2021) for the baseline model. Each panel displays an overlapping 2-week-ahead forecast generated at every fourth assimilation step within a single calendar year, providing a visual summary of how forecast uncertainty evolves across the transmission season. Each forecast horizon originates at a forecast origin date and extends through the 1-week-ahead and 2-week-ahead target dates; shaded ribbons show the 50% (darkest), 80% (medium), and 90% (lightest) prediction intervals at those two horizons; the colored line connecting the two horizon points shows the median forecast trajectory; the red line shows the observed total abundance values. The y-axis scale for 2021 is unconstrained; all other years share a common axis scale.

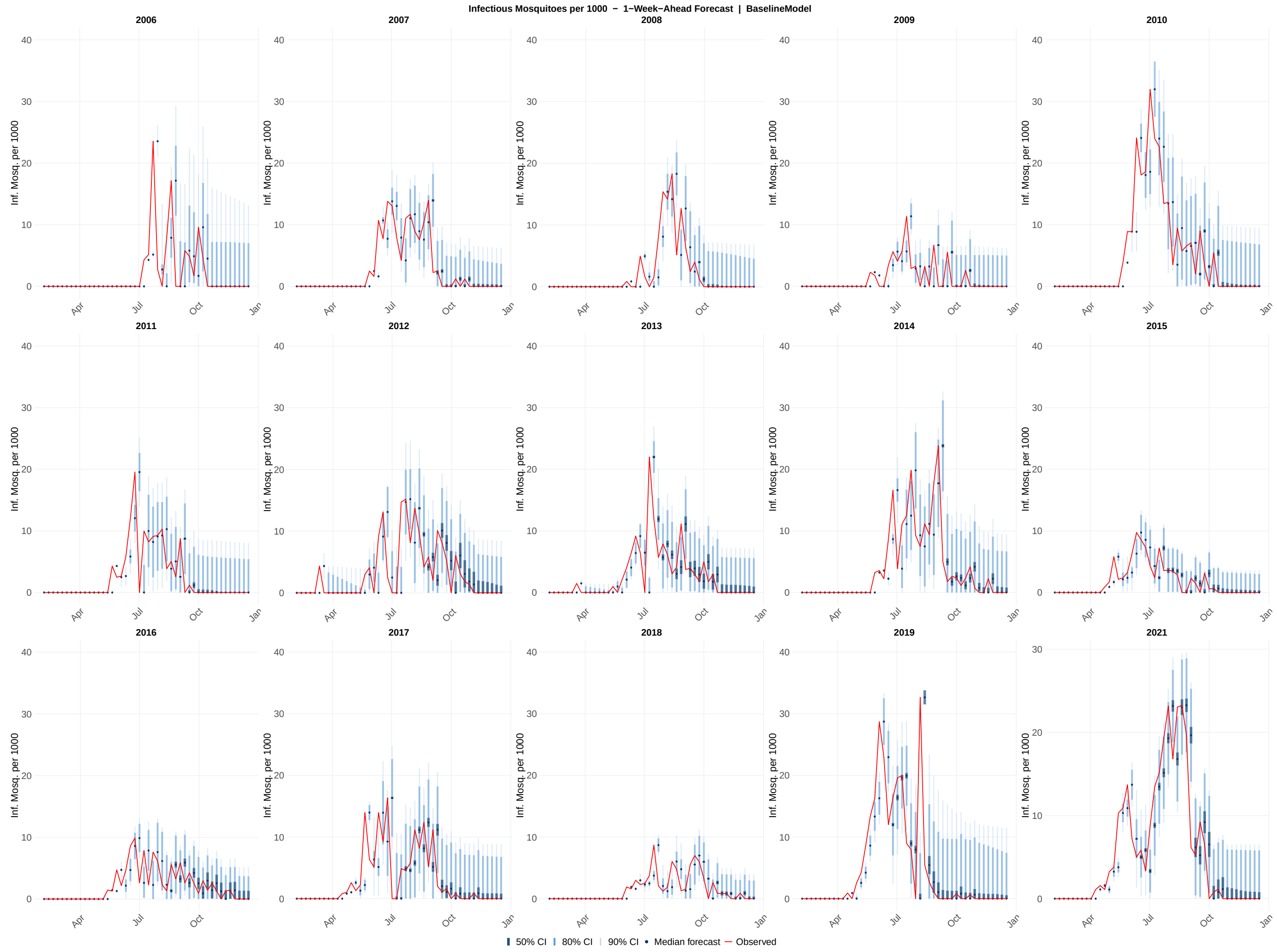

**Figure S75:** Probabilistic 1-week-ahead forecasts of infectious mosquitoes per 1000 across all 15 study years (2006–2019, 2021) for the baseline model. Each panel shows the sequence of 1-week-ahead forecast distributions generated at each of the 46 assimilation steps within a single calendar year. Shaded ribbons indicate the 50% (darkest), 80% (medium), and 90% (lightest) prediction intervals; the solid line shows the forecast median; the red line shows the observed infectious mosquito per 1000 values. The y-axis scale for 2021 is unconstrained; all other years share a common axis scale.

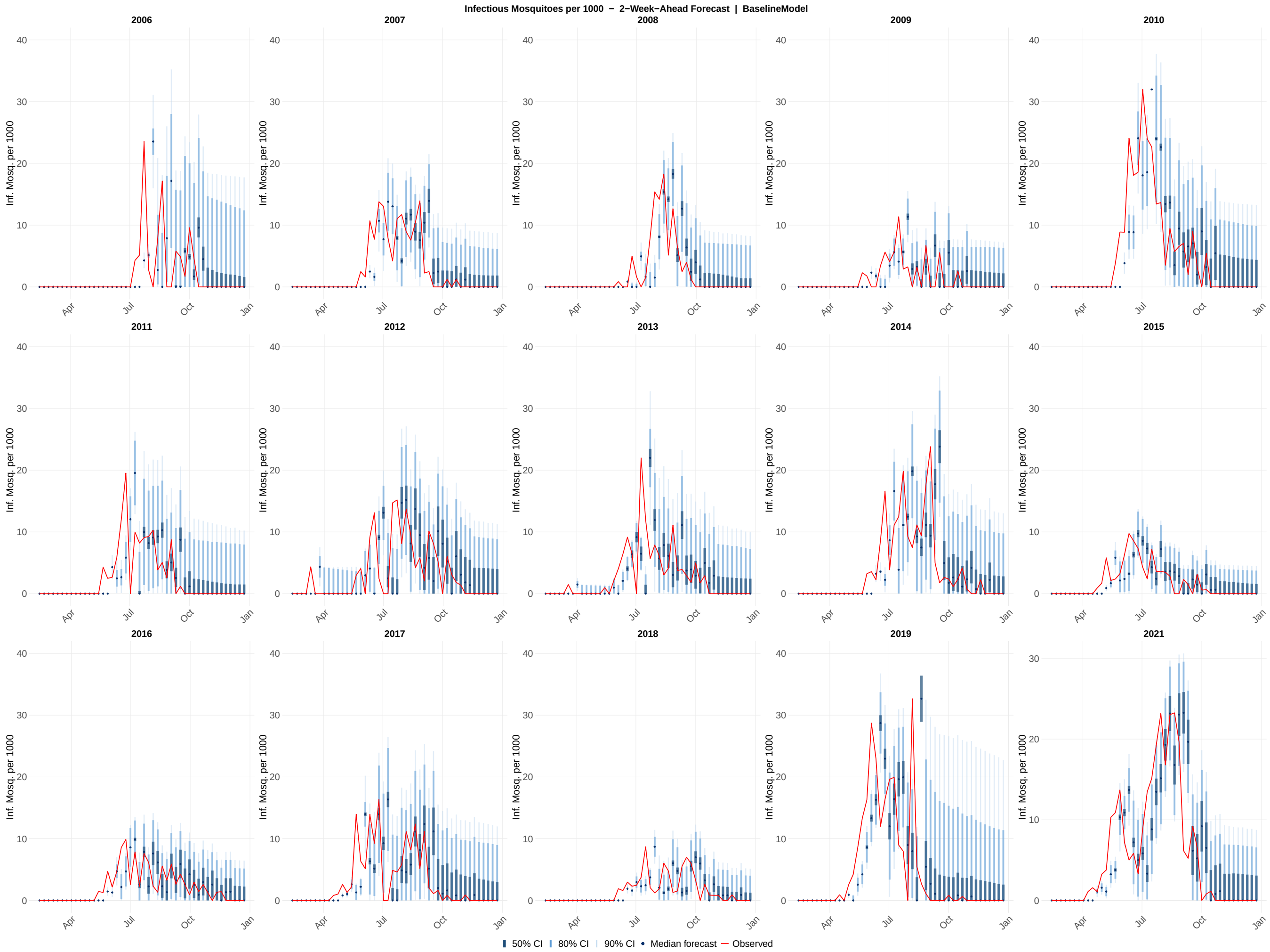

**Figure S76:** Probabilistic 2-week-ahead forecasts of infectious mosquito per 1000 across all 15 study years (2006–2019, 2021) for the baseline model. Each panel shows the sequence of 2-week-ahead forecast distributions generated at each of the 46 assimilation steps within a single calendar year. Shaded ribbons indicate the 50% (darkest), 80% (medium), and 90% (lightest) prediction intervals; the solid line shows the forecast median; and the red line shows the observed infectious mosquito per 1000 values. The y-axis scale for 2021 is unconstrained; all other years share a common axis scale.

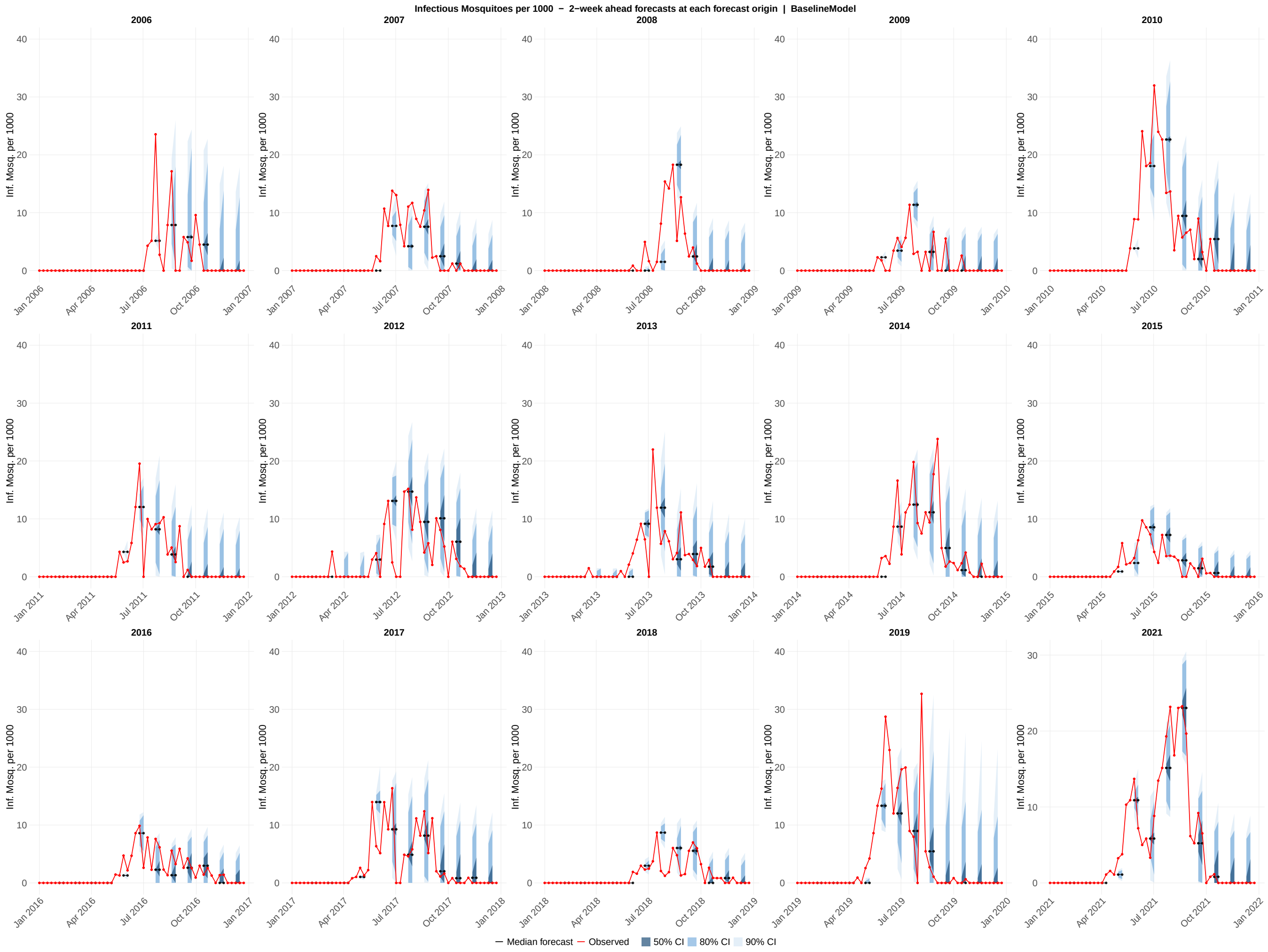

**Figure S77:** Probabilistic forecasts of infectious mosquitoes per 1000 all 15 study years (2006–2019, 2021) for the baseline model. Each panel displays overlapping 2-week-ahead forecast generated at every fourth assimilation step within a single calendar year, providing a visual summary of how forecast uncertainty evolves across the transmission season. Each forecast horizon originates at a forecast origin date and extends through the 1-week-ahead and 2-week-ahead target dates; shaded ribbons show the 50% (darkest), 80% (medium), and 90% (lightest) prediction intervals at those two horizons; the colored line connecting the two horizon points shows the median forecast trajectory; the red line shows the observed infectious mosquito per 1000 values. The y-axis scale for 2021 is unconstrained; all other years share a common axis scale.

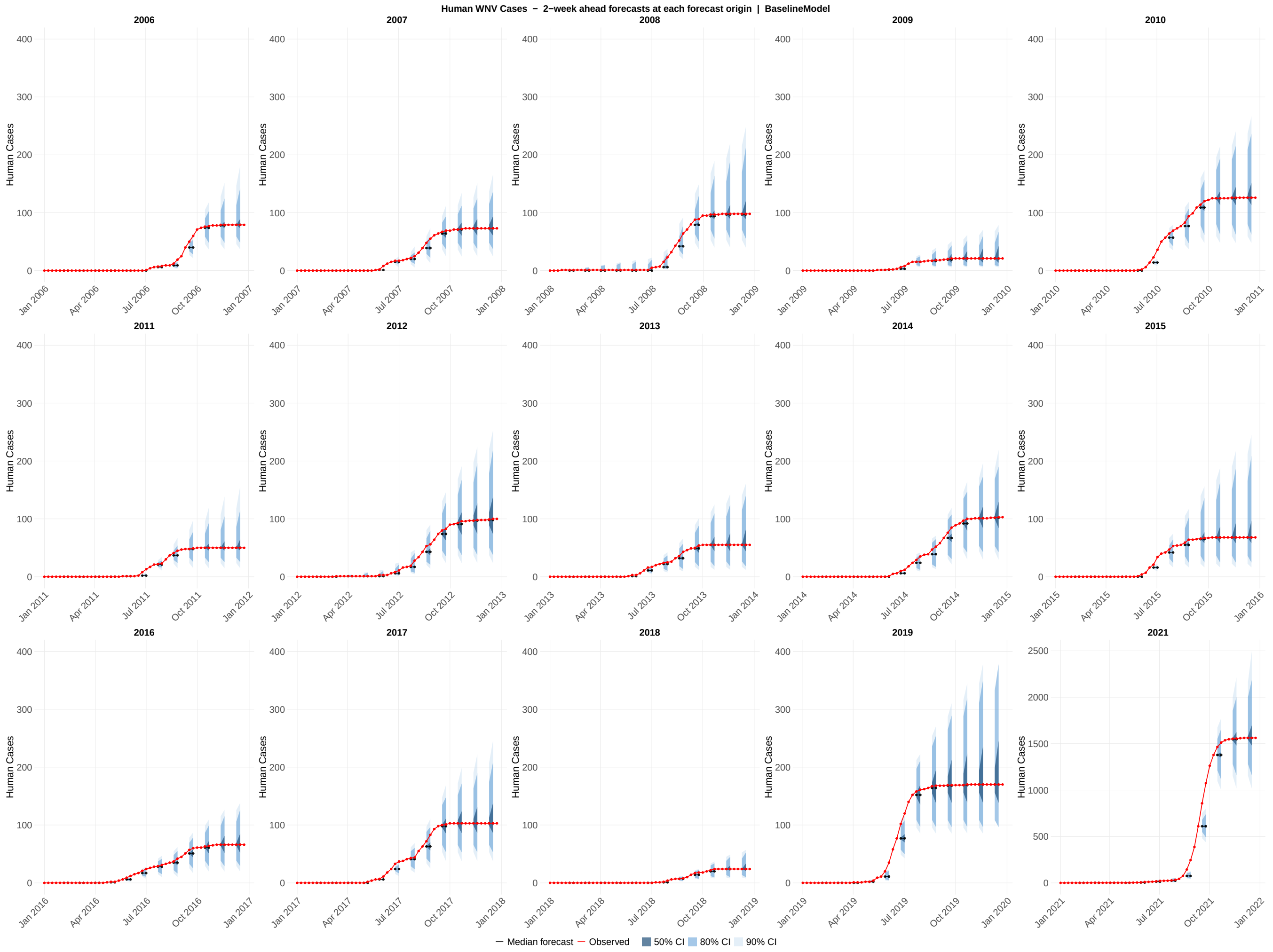

**Figure S78:** Probabilistic forecasts of human cases across all 15 study years (2006–2019, 2021) for the baseline model. Each panel displays overlapping 2-week-ahead forecast generated at every fourth assimilation step within a single calendar year, providing a visual summary of how forecast uncertainty evolves across the transmission season. Each forecast horizon originates at a forecast origin date and extends through the 1-week-ahead and 2-week-ahead target dates; shaded ribbons show the 50% (darkest), 80% (medium), and 90% (lightest) prediction intervals at those two horizons; the colored line connecting the two horizon points shows the median forecast trajectory; the red line shows the observed human cases values. The y-axis scale for 2021 is unconstrained; all other years share a common axis scale.

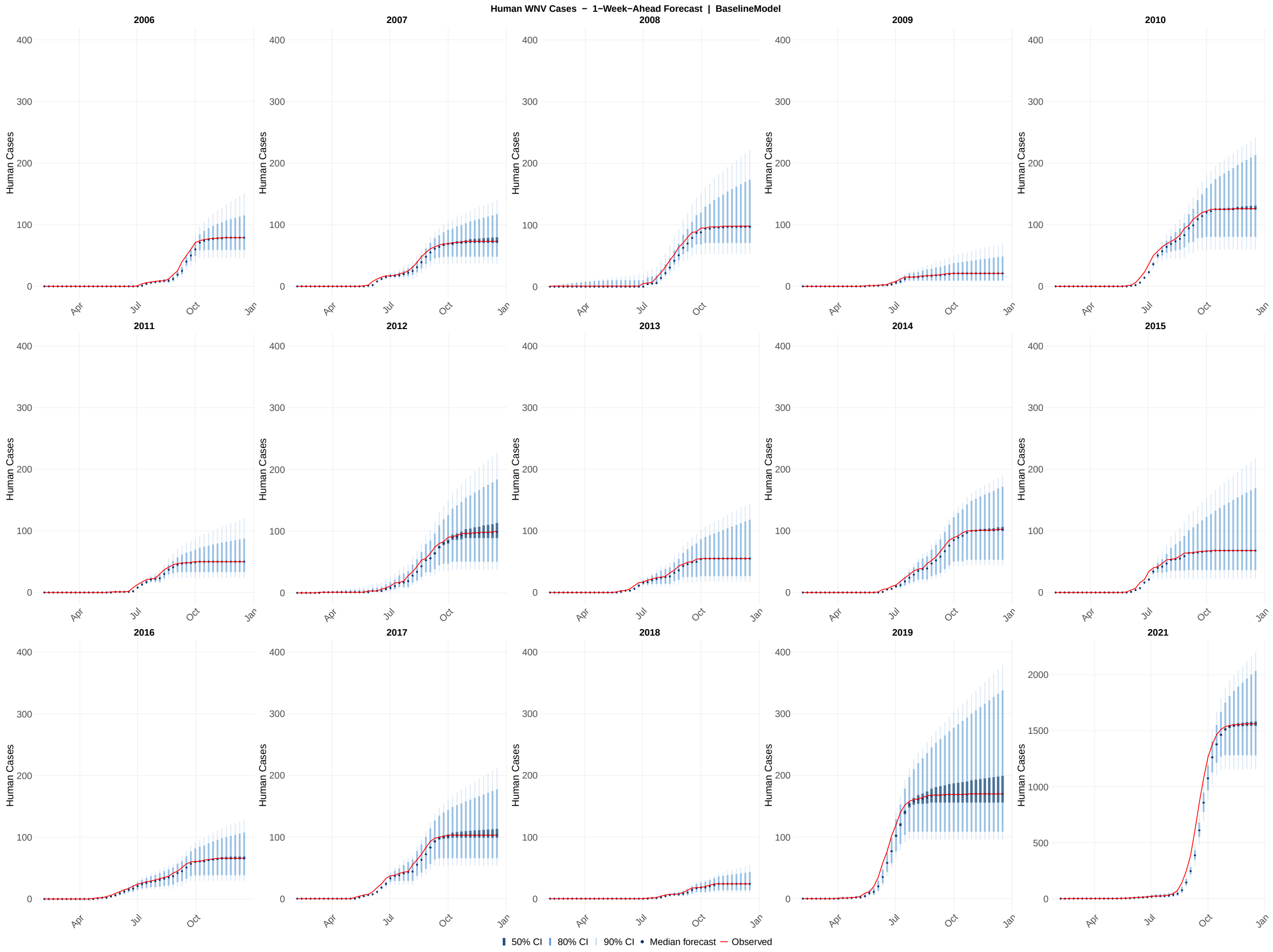

**Figure S79:** Probabilistic 1-week-ahead forecasts of human cases across all 15 study years (2006–2019, 2021) for the baseline model. Each panel shows the sequence of 1-week-ahead forecast distributions generated at each of the 46 assimilation steps within a single calendar year. Shaded ribbons indicate the 50% (darkest), 80% (medium), and 90% (lightest) prediction intervals; the solid line shows the forecast median; the red line shows the observed human cases values. The y-axis scale for 2021 is unconstrained; all other years share a common axis scale.

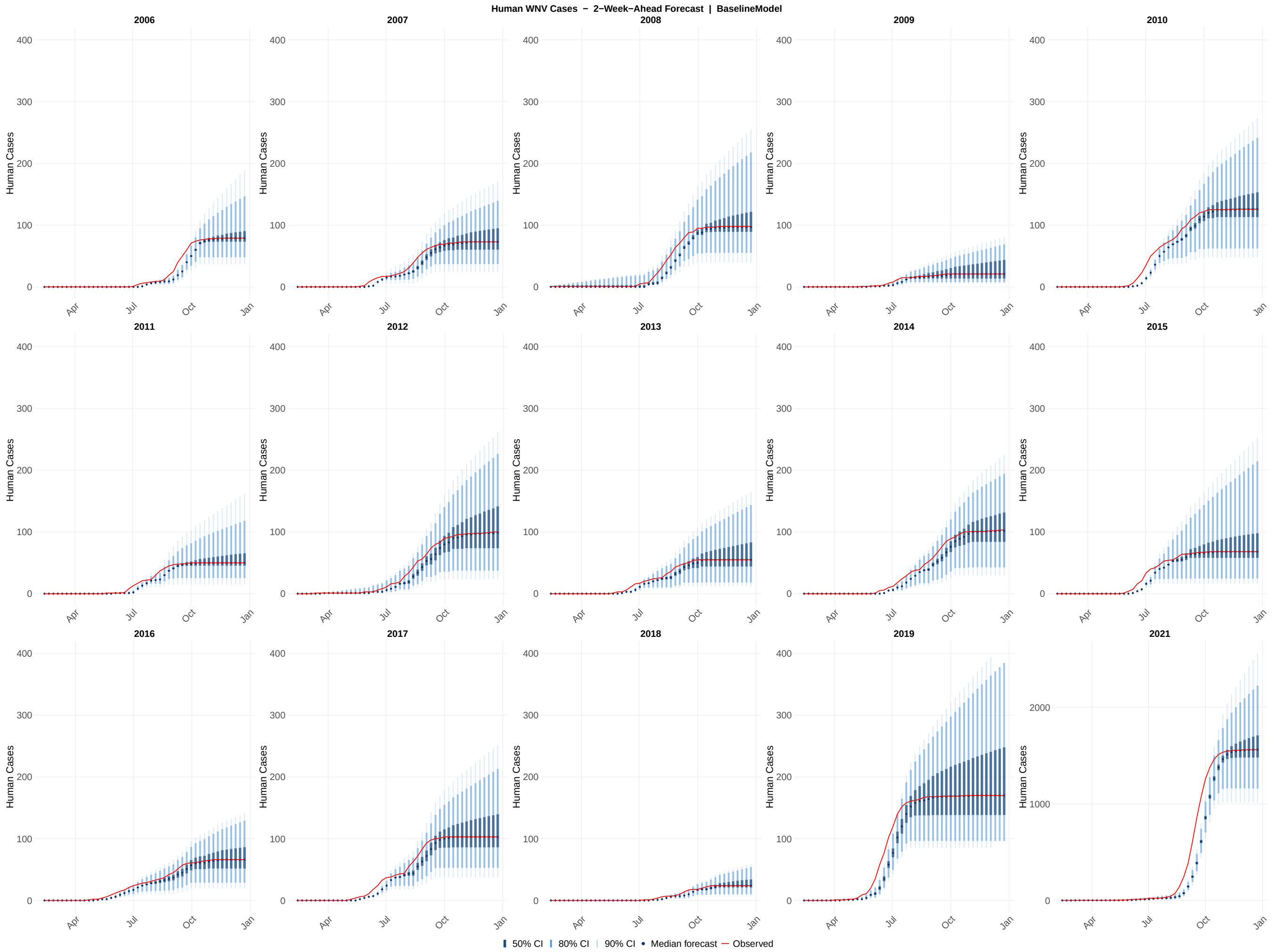

**Figure S80:** Probabilistic 2-week-ahead forecasts of human cases across all 15 study years (2006–2019, 2021) for the full model. Each panel shows the sequence of 2-week-ahead forecast distributions generated at each of the 46 assimilation steps within a single calendar year. Shaded ribbons indicate the 50% (darkest), 80% (medium), and 90% (lightest) prediction intervals; the solid line shows the forecast median; the red line shows the observed human cases values. The y-axis scale for 2021 is unconstrained; all other years share a common axis scale.
